# A novel proteomics-based plasma test for early detection of multiple cancers in the general population

**DOI:** 10.1101/2023.05.06.23289613

**Authors:** Bogdan Budnik, Hossein Amirkhani, Mohammad H. Forouzanfar, Ashkan Afshin

## Abstract

**Background:** Early detection of cancer is crucial for reducing the global burden of cancer and saving lives, but effective screening tests for many cancers do not exist. Genomics-based liquid biopsy tests for screening multiple cancers at once have been developed, but they have low sensitivity for early-stage cancers and are expensive. Recent advancements in measuring protein abundances in plasma offer new opportunities for developing multi-cancer screening tests.

**Methods:** We collected plasma samples from 440 individuals, healthy and diagnosed with 18 various types of early-stage solid tumours. Using Proximity Extension Assay, we measured more than 3000 high and low-abundance proteins in each sample. Then, using a multi-step statistical approach, we identified a limited set of proteins that could detect early-stage cancers and their tissue of origin with high diagnostic accuracy.

**Findings:** Our sex-specific cancer detection consisting of 10 proteins showed high accuracy for both males (AUC: 0.98) and females ((AUC: 0.983). At stage I and at the specificity of 99%, our detection panels were able to identify 89% of cancers among males and 75% of cancers among females.

Our sex-specific localization panels consisted of 150 proteins and were able to identify the tissue of origin of most cancers in more than 80% of cases. The analysis of the plasma concentrations of proteins selected showed that almost all the proteins were in the low-concentration part of the human plasma proteome.

**Interpretation:** The proteome-based screening test showed promising performance compared with other technologies and could be a starting point for developing a new generation of screening tests for the early detection of cancer and potentially other chronic diseases. This new approach may provide a more accessible and cost-effective alternative to existing methods for cancer detection and may help reduce cancer mortality rates globally.

## Introduction

Cancer is a leading cause of mortality globally, accounting for one in every six deaths.^1^ In the absence of established risk factors for many cancers, early detection and early treatment remain the cornerstone of clinical and public health strategies for reducing the global burden of cancer and saving lives. However, currently, no effective test exists for the early detection of many cancers. Nearly 60% of cancer-related deaths are due to cancers for which no screening test exists.^2^ Additionally, existing screening tests (i.e., colonoscopy, CT scan, mammography, pap test) have major limitations, including invasiveness, high cost, and low accuracy for early stages.

Liquid biopsy, the analysis of biomarkers in non-solid specimens, has emerged as a promising approach for developing novel biomarkers.^3^ In recent years, efforts have been made to develop a genomics-based liquid biopsy test for screening multiple cancers at once.^4 5^ For example, a blood test has been developed to identify the presence of over 50 cancers based on their methylation signatures in cell-free DNA.^6^ However, these genomics-based multi-cancer tests have shown low sensitivity for early-stage cancers (<50%).^7^ Additionally, they are too expensive (>500 USD) to be covered by most insurance companies and incorporated as routine screening tests in the health care system.

Protein biomarkers in the blood have the potential to be used for early detection and ongoing monitoring of diseases, but the current options have significant limitations due to a lack of diagnostic specificity.^8^ In this paper, we explore the potential use of plasma proteins as biomarkers for solid tumours (excluding melanoma) in specific organs and the need to search for biomarkers in the depths of the proteome that are currently undetectable. We discuss the lower sensitivity of current protein assays compared to nucleic acid detection methods and the need to be able to detect very small amounts of proteins to identify early stages of cancer growth through liquid biopsy.

## Methods

We collected plasma samples from a total of 440 individuals, healthy and those diagnosed with 18 various types of early-stage solid tumours. The 18 solid tumours (14 tumours in men and 16 in women) were chosen because of the invaluable opportunity of the cure at early stages before local invasion or spreading metastases happens and a potential diagnostic test at the early stage could reduce the mortality and cost of the cancer significantly. Deep proteomics profiling of plasma samples was conducted using Proximity Extension Assay technology^9-11^ and measured nearly 3,000 proteins in each sample. Then, we attempted to identify cancer protein signatures for the purpose of cancer detection. Our approach involved two main steps. In the first step, we searched for a limited number of proteins that could identify any cancer in its early stages. In the second step, we classified each type of cancer against the others to find a cancer-specific signature for localization (i.e., tissue of origin). Both steps were done for male and female samples separately and probability-based score was calculated at each step for a person’s protein array sample and the patient was classified by the calculated score. As with other biomarker studies, our experiment size was relatively small.^12^ Therefore, we used a multi-step approach to define a minimal set of proteins (also known as the “best set”) that could classify samples from a stable model and be generalizable. To select the features forming the minimum set, 100 bootstrap samples of the original dataset were analysed by logistic regressions with L1 penalty to identify the proteins with stronger association and best statistical significance for all cancers, pan cancer analysis, and differentiation of each cancer from other solid tumours (localization). The use of the L1 penalty ensures the sparsity of the selected biomarkers, preventing the simultaneous selection of correlated biomarkers. A leave-one-out validation was performed to evaluate the performance of the model with selected features. For cancer localization, the proteins were evaluated to identify the site of origin and/or subtype of the cancer for each cancer versus other cancers in the dataset. The top cancer specific proteins with the strongest associations with the target cancers were selected iteratively, which resulted in different size minimum sets for the cancers with the overall highest average performance. Then, for a given sample, the predicted probabilities of it belonging to different cancers were calculated using the selected proteins, and the cancer with the highest probability score was determined as the predicted cancer.

## Results

### The effect of protein correlation and sex

Out of the 3072 proteins that were analysed, 280 did not pass the quality measurements and were excluded from further analysis. The abundance of proteins showed varying correlation levels, with positive correlations potentially being a result of shared biological pathways. Many protein pairs displayed high positive correlation, as indicated by the correlation matrix (Fig 1). However, highly correlated proteins could make the analysis unstable, despite being related to cancer. Additionally, different biological pathways contribute to the initiation and progression of cancer, which highlights the need to capture diverse cancer types through different pathways and potentially less correlated proteins.^13^ Therefore, in our analysis, we only picked the most useful proteins out of internally correlated sets to be able to classify more patients.

**Figure 1.**
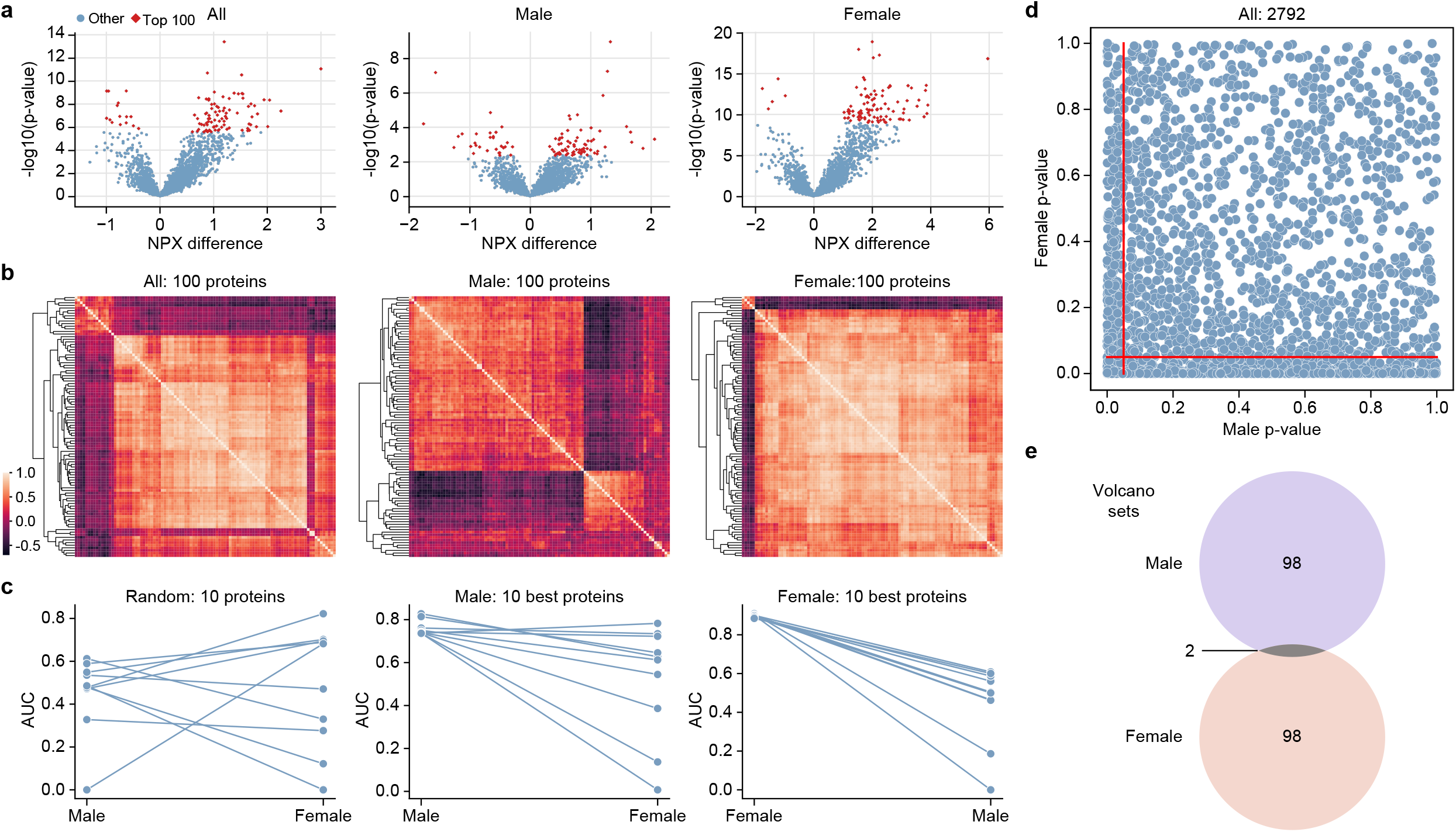
The difference between protein biomarkers in males and females. (a) The Volcano plots showing differential abundances of proteins among both sexes, males, and females. Top 100 proteins based on their p-value highlighted in red. (b) The correlation matrix of the top 100 proteins based on p-value among males, females, and both sexes. (c) Ranking of the top 10 proteins based on AUC in males and females as well as the ranking of 10 randomly selected proteins in males and females. (d) The scatter plot of p-values for each protein among males and females. (e) The Venn diagram of top 100 proteins based on volcano plots in males and females.

We found that the protein-cancer association varied significantly between male and females (Fig 1). A simple comparison between cancer and normal samples for men and women showed a poor correlation across protein profiles. Many proteins with a p-value of less than 0.001 in one sex showed no significant difference in the other one. An analysis of volcano plots showed minimal overlap between top 100 proteins selected based on their differential expression and their p-value. An analysis of the top 10 proteins using AUC (area under the curve) of the ROC curve (receiver operating characteristic curve) showed the order of importance of the proteins varied between males and females. The 10 proteins with the best AUC to differentiate cancer from normal in male samples performed poorly in females and vice versa. Thus, despite the sample size limitation, we decided to perform the remainder of the analyses on male and female samples separately to capture the sex specific patterns.

### Optimal number of proteins for cancer detection and localization

Finding the best sex-specific sets to identify a cancer was performed in two steps. At the first step, we detected the protein signature of any cancer (pan cancer/any cancer classifier) to classify any cancer from normal, followed by the second step, identifying the tissue of origin of cancer and cancer subtypes (i.e., small cell and non-small cell cancers of lung, and cervical and endometrial cancers of uterine).

For each step, we tested several numbers of proteins and expected to see an increasing performance of the model by AUC with more proteins to capture different cancer populations. The model performance increased quickly with adding a few more proteins but after 10 proteins in any cancer model, no more improvement in AUC was observed (Fig 2). A similar phenomenon was observed in the cancer localization step (Fig 4). Since the performance of different predictive models for specific cancers plateaued at different numbers, we employed an algorithm to pick the proteins-cancer pairs for additional proteins if improved overall AUC more. For the localization panels, the highest performance was observed at the level of 150 proteins. As expected, different numbers of proteins were picked as the best set by the algorithm. Moreover, some proteins were helpful in localizing more than one cancer.

**Figure 2.**
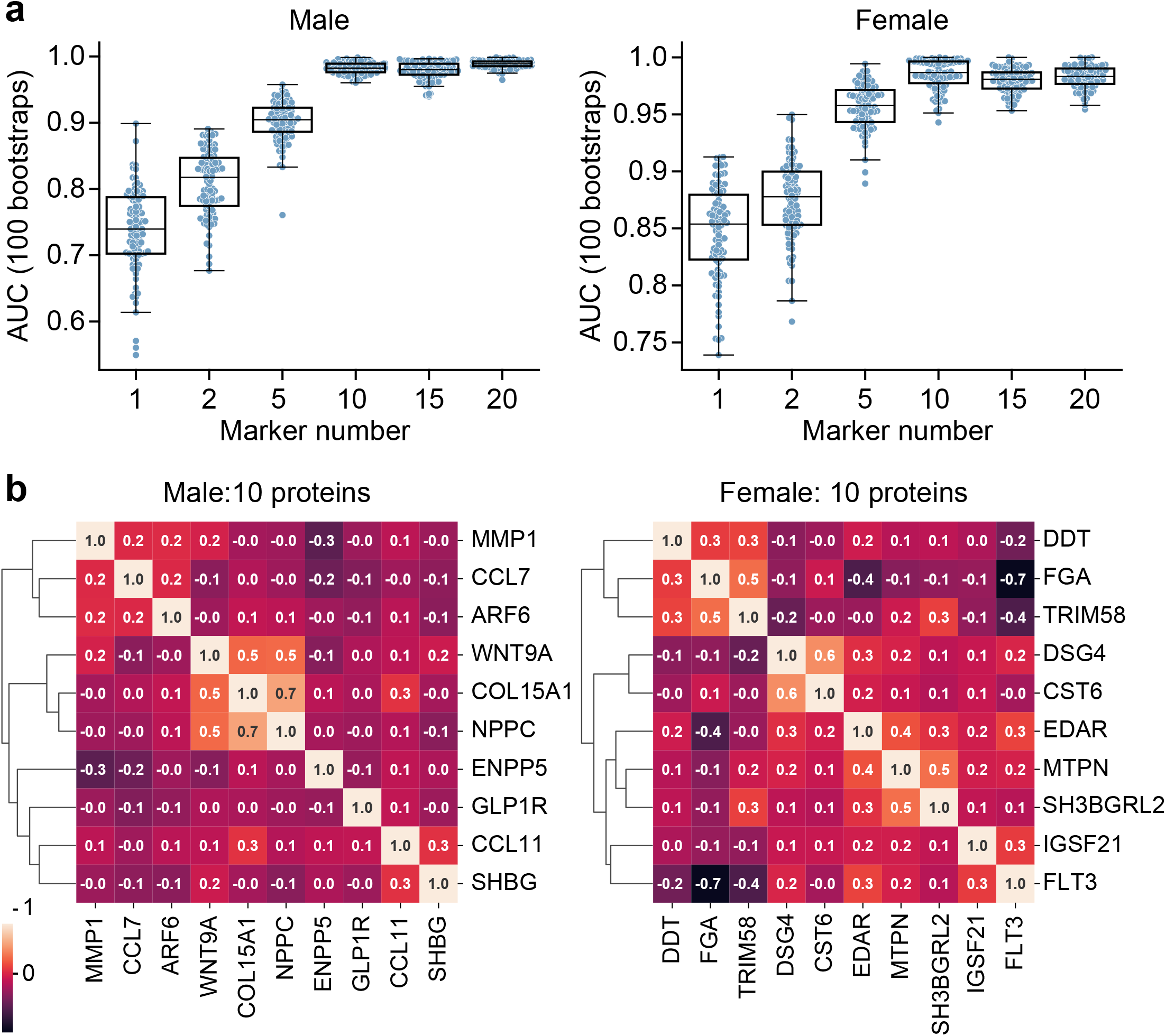
Selection of cancer detection panel. (a) The relationship between the number of proteins included in the protein panel and performance of the panel. (b) The correlation between proteins included the detection panel by sex.

### Sex-specific cancer detection panel

Our final detection panels for male and female each consisted of 10 proteins that were differentially expressed among normal and cancer plasma samples (Fig 2). Each protein of the panel alone had a low to medium detection accuracy but when assessed in combination with other proteins as a panel they achieved a very high accuracy in detection of early-stage cancers (Fig 3). The proteins in the panel showed low to medium correlation indicating each protein contributing new information and presenting a different pathway to the panel.

**Figure 3.**
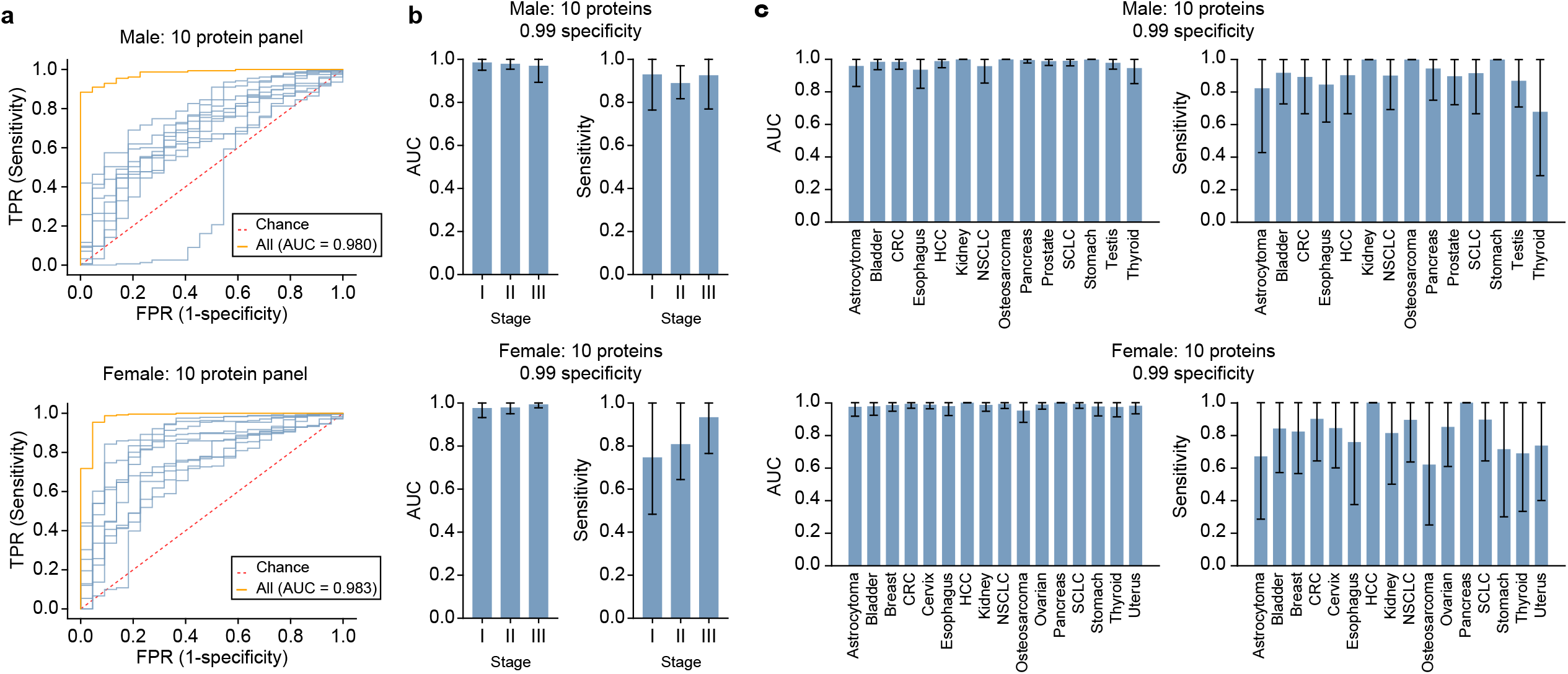
Performance of the detection panel. (a) ROC curve (receiver operating characteristic curve) of detection panel for males and females. (b) Sensitivity of the detection panel at the specificity of 99% for each stage of the cancer for males and females. (c) Sensitivity of detection panel at the specificity of 99% by stage for males and females.

**Figure 4.**
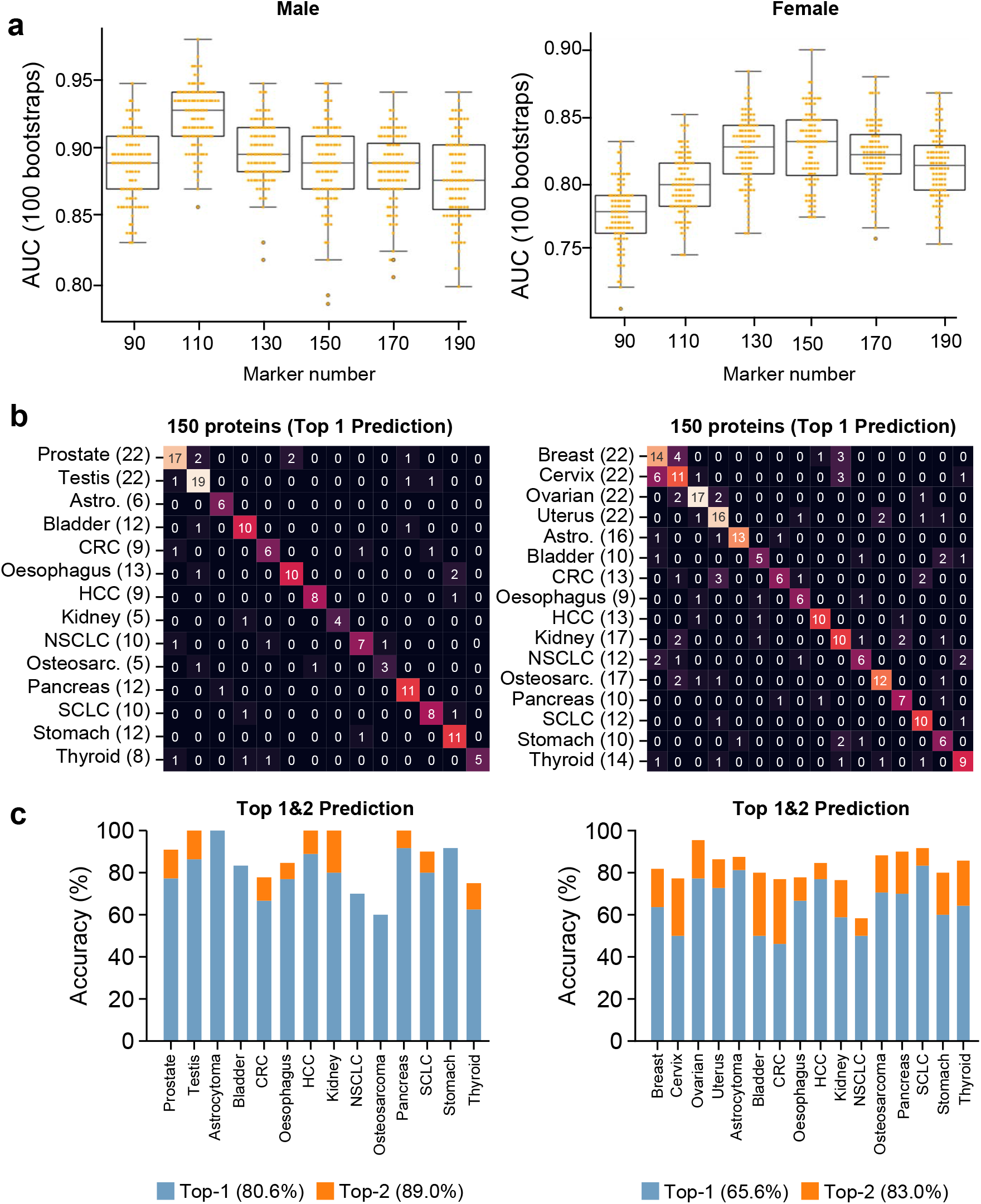
The selection and performance of the localization panels across cancers. (a) The relationship between the number of proteins included in the localization panel and the overall performance of the panel. (b) The confusion matrix showing the performance of the of the test in correctly identifying the source of the cancer in the first prediction. (c) The bar chart of the overall performance of the localization panel.

Overall, at the detection step, our protein panels showed high sensitivity and specificity among males and females (Fig 3). At the specificity of 99%, the overall sensitivity of our test was 88% among males and 74% among females. We also observed high accuracy across all stages of cancer among males and females. At stage I and at the specificity of 99%, our panel was able to identify 89% of cancers among males and 75% of cancers among females. The performance of the panels varied across cancers. Overall, some cancers were easier to detect (e.g., pancreas and kidney cancers). On the other hand, the detection of cancers like astrocytoma and thyroid was more challenging but achieved with relatively high accuracy after optimization.

To further evaluate the performance of the cancer set to detect any cancer, we excluded one cancer at a time and evaluated the fitted model on other cancers by the algorithm above on the excluded (unseen) cancer. Our analysis showed an acceptable performance with an AUC of, on average, above 0.9 for an unseen cancer (Fig S3). The cancer-out model has better performance on unseen cancers of pancreas and liver cancer but a lower performance to detect unseen thyroid cancer and astrocytoma. This shows that the protein set correlates very well with the cancer and serves as a cancer detection test. A plausible baseline cancer detection signature will enable efficient expansion to other cancers with a limited number of new samples added to the training set.

### Sex-specific cancer localization panel

Our localization panels consisted of 150 proteins. Each sample was fed into separate cancer versus other cancers and the prediction probability for that cancer was calculated. The top two highest probability was used to identify the tissue of origin. The number of proteins allocated to each cancer was selected in a way that optimized the overall performance of the test. In males, the highest number of proteins was allocated to bladder cancer and the lowest number of proteins were allocated to liver cancer. In females, the highest and the lowest number of proteins were allocated to ovarian cancer and bladder cancer, respectively.

We evaluated the overall performance of the test by its ability to correctly classify the samples at the detection and localization level. Overall, at the specificity of 99% our sex-specific test showed the detection sensitivity of 88% for male and 74% for female. The localization accuracy of the tests was 89% for males and 77% for females.

### The role of downregulated and low-concentration proteins

Through our analysis of 18 cancers and normal plasma samples, we found that only a few cancers can be uniquely identified by up-regulated proteins, which are typically preferred as biomarkers. We discovered that many cancers showed much higher specificity using downregulated proteins rather than just upregulated proteins. As the number of cancers included in a single pan-cancer test increases, it will be crucial to have both types of regulation biomarkers to achieve high cancer specificity among many different cancers.

Our evaluation of the plasma concentrations of proteins that were selected showed that almost all the proteins were in the low abundance group. The critical proteins such as CCL11 required no dilution, compared with high abundance proteins such as FGA which require 100,000-fold dilution to be detected reliably by proximity technology. This highlights the importance of low-concentration proteins to see precancerous states and early stages where the tumour has little systemic impact and generated footprints.

## Discussions

In this study, we showed that a measurement of limited set of plasma proteins could classify cancer samples from normal and differentiate different cancers. This finding is the foundation for a multi-cancer screening test for the early detection of 18 solid tumours that cover all major human organs of origin for such cancers at the earliest stage of their development with high accuracy. It is important to diagnose cancer at very early stages where curative treatments are achievable with surgery and available treatments. Additionally, for the first time to our knowledge, we found compelling evidence that the cancer protein signatures are most likely sex specific for all cancers. Our study also showed that biological signals for early-stage cancers are much more evident in the low-concentration part of the human plasma proteome. It was also promising to observe that a set of proteins could differentiate all cancers from normal and sensitive to detect unseen cancers

In our study, we analysed a range of proteins found in classical cancer pathways. However, we discovered that only a very small number of these proteins could be used as biomarkers for early-stage cancer. In contrast, many proteins that were effective biomarkers for early-stage cancer were found at low concentrations across the entire plasma proteome. This finding may be due to the fact that most of our knowledge about the role of proteins in cancer pathways comes from studies of transcriptome at the tissue level in advanced stages of cancer, and the expression of proteins at the mRNA and protein levels do not always correspond. In addition, the concentration of proteins in tissue and plasma may not be strongly correlated. Finally, our samples were mainly from early-stage cancers, where classic cancer pathways may not be highly active. This finding has major implication for developing the next generation of diagnostics highlighting the role low-abundance protein in early detection of disease.

The proteome-based diagnostic test showed promising performance compared with other technologies such as circulating tumour DNA tests (ctDNA)^14^ by significantly outperforming existing multi-cancer screening tests in detecting cancer across all stages (I, II, III) and among all types of cancers. At the specificity of 99% and in stage I of cancer, our test had a sensitivity that was much greater than Galleri^15^ and CancerSEEK^16^ tests. Additionally, our study demonstrated ability of our “best-test” to achieve much higher accuracy in identifying the tissue of origin of cancers in each sample in comparison to other tests. At the cancer-specific level, all our best-tests were more accurate than other available screening tests. Among the four screening tests that have received the highest recommendation (Level A) from the U.S. Preventive Services Task Force (colonoscopy for colon cancer, pap test for cervical cancer, mammography for breast cancer, and low-dose CT scan for lung cancer), only coloscopy and low-dose CT scan had an accuracy of above 90% for cancer detection. However, the sensitivity of our test for detecting early-stage cancer was still higher than the sensitivity of these tests.

Over the past decade, mRNA large-scale sequencing has provided a comprehensive view of gene expression in specific tissues, revealing the proteins that are present in different organs of the human body. The Human Protein Atlas is a useful resource for understanding mRNA and protein expression in multiple healthy tissues.^17^ However, it is important to note that tissues are typically composed of complex assemblies of distinct cells that may have different functions and developmental histories. Increasing amounts of information about RNA and protein expression in specific cell types is now becoming available for the individual cells that make up tissues and organs. A challenge in using protein detection for liquid biopsy is that cancer-specific protein biomarkers may be present at ultra-low levels in the blood.^18^ This is because proteins that are present at high concentrations in the blood of healthy individuals are unlikely to be significantly increased in patients with early stages of the disease or at early recurrence. The long history of plasma proteome analysis by mass spectrometry show that even proteome coverage was increased from several hundred proteins thirty years ago to more than five thousand proteins based on latest development in chromatography separation technique and DIA (Data Independent Acquisition) type of acquisition.^19^ Still the major problem of cheap and reproducible sample preparation protocols and reliably measuring proteins after first thousands of most abundant proteins prevent development of early stage multi cancer test by mass spectrometry at acceptable price per sample and general population scale. Thus, assays with greater sensitivity for biomarker proteins that are normally present at very low or undetectable levels in the blood may enable the detection of cancer at an earlier stage of the disease or even at premalignant stages. Our test is based on sensitive proximity assays that require the simultaneous binding of three separate antibodies. This ability to analyse plasma proteome profiles deeply and consistently allowed us to focus our attention on very low-abundant proteins, which we found to be the most precise and accurate biomarkers of early stages for all the cancers studied in our study. Advancing the PEA technology to measure ultra-low protein concentrations will provide better opportunities to detect and classify cancers at a very early stage and even at the precancerous stage.^20^

Our new generation protein-based plasma test has shown high sensitivity in detecting a variety of early-stage tumours in asymptomatic patients, making it a strong candidate for use as a population-wide screening tool that is not currently achievable with existing tests or techniques. Its high specificity can help alleviate concerns about causing harm to patients, and its low cost allows for widespread implementation. To be suitable for large-scale use, a screening test must have high sensitivity and the ability to reduce mortality and morbidity, as well as acceptable for healthcare system cost. In the case of cancer screening, it is also essential for the test to have high specificity to avoid causing undue harm to patients. Our test exhibits these desirable qualities, making it a promising option for cancer screening. We expect that the combination of lower cost and higher accuracy in our test will facilitate its integration into the healthcare system and eventual inclusion in routine annual check-ups. Early detection of cancer has the potential to greatly reduce the societal burden of both health and financial costs. In fact, implementing such interventions can not only be cost-effective but can also result in cost savings for society.

Our approach has major strengths, including the total number of proteins measured and accuracy of such measurements across all measured proteins down to very low abundant proteins, the focus on early-stage tumours, the number of studied cancers that represents all major organs of unmet needs included in the study.

Limitations should also be considered, including small cohort size and the existence of comorbidities. Therefore, our test needs to be further validated in a larger population cohort before being widely accepted into healthcare systems across various populations. We also foresee the expansion of the test to include all cancers with unmet test needs.

## Conclusions

In summary, the unique contributions of this study include: the analysis of the most extensive set of proteomics data available in a multi-cancer sample, the development of a cancer-specific protein signature for early-stage cancers that focuses on the baseline carcinogenic state rather than the end-stage tumour behaviours and human response, the identification of a sex-specific protein profiles and cancers signatures, the highlighting the importance of sex specific proteins set for early cancer detection and importance of downregulated proteins as sensitive biomarkers at the early stage, and the demonstration of the feasibility and potential performance of this approach for early-stage diagnosis of all major cancers at the population level.

## Data Availability

All data produced in the present study are available upon reasonable request to the authors

## References

1. Collaborators GBDCRF. The global burden of cancer attributable to risk factors, 2010-19: a systematic analysis for the Global Burden of Disease Study 2019. Lancet 2022;400(10352):563–91. doi:10.1016/S0140-6736(22)01438-6

2. National Cancer Institute. Crunching Numbers: What Cancer Screening Statistics Really Tell Us 2018 [cited 2023. Available from: https://www.cancer.gov/about-cancer/screening/research/what-screening-statistics-mean.

3. Geyer PE, Kulak NA, Pichler G, et al. Plasma Proteome Profiling to Assess Human Health and Disease. Cell Syst 2016;2(3):185–95. doi:10.1016/j.cels.2016.02.015 [published Online First: 20160323]

4. Marrugo-Ramirez J, Mir M, Samitier J. Blood-Based Cancer Biomarkers in Liquid Biopsy: A Promising Non-Invasive Alternative to Tissue Biopsy. Int J Mol Sci 2018;19(10) doi:10.3390/ijms19102877 [published Online First: 20180921]

5. Ding Z, Wang N, Ji N, et al. Proteomics technologies for cancer liquid biopsies. Mol Cancer 2022;21(1):53. doi:10.1186/s12943-022-01526-8 [published Online First: 20220215]

6. Hasenleithner SO, Speicher MR. How to detect cancer early using cell-free DNA. Cancer Cell 2022;40(12):1464–66. doi:10.1016/j.ccell.2022.11.009

7. Loomans-Kropp HA, Umar A, Minasian LM, et al. Multi-Cancer Early Detection Tests: Current Progress and Future Perspectives. Cancer Epidemiol Biomarkers Prev 2022;31(3):512–14. doi:10.1158/1055-9965.EPI-21-1387

8. Hackshaw A, Cohen SS, Reichert H, et al. Estimating the population health impact of a multicancer early detection genomic blood test to complement existing screening in the US and UK. Br J Cancer 2021;125(10):1432–42. doi:10.1038/s41416-021-01498-4 [published Online First: 20210823]

9. Fredriksson S, Gullberg M, Jarvius J, et al. Protein detection using proximity-dependent DNA ligation assays. Nat Biotechnol 2002;20(5):473–7. doi:10.1038/nbt0502-473

10. Lundberg M, Eriksson A, Tran B, et al. Homogeneous antibody-based proximity extension assays provide sensitive and specific detection of low-abundant proteins in human blood. Nucleic Acids Res 2011;39(15):e102. doi:10.1093/nar/gkr424 [published Online First: 20110606]

11. Darmanis S, Nong RY, Hammond M, et al. Sensitive plasma protein analysis by microparticle-based proximity ligation assays. Mol Cell Proteomics 2010;9(2):327–35. doi:10.1074/mcp.M900248-MCP200 [published Online First: 20091127]

12. Skates SJ, Gillette MA, LaBaer J, et al. Statistical design for biospecimen cohort size in proteomics-based biomarker discovery and verification studies. J Proteome Res 2013;12(12):5383–94. doi:10.1021/pr400132j [published Online First: 20131028]

13. Sever R, Brugge JS. Signal transduction in cancer. Cold Spring Harb Perspect Med 2015;5(4) doi:10.1101/cshperspect.a006098 [published Online First: 20150401]

14. Wan JCM, Massie C, Garcia-Corbacho J, et al. Liquid biopsies come of age: towards implementation of circulating tumour DNA. Nat Rev Cancer 2017;17(4):223–38. doi:10.1038/nrc.2017.7 [published Online First: 20170224]

15. Klein EA, Richards D, Cohn A, et al. Clinical validation of a targeted methylation-based multicancer early detection test using an independent validation set. Ann Oncol 2021;32(9):1167–77. doi:10.1016/j.annonc.2021.05.806 [published Online First: 20210624]

16. Cohen JD, Li L, Wang Y, et al. Detection and localization of surgically resectable cancers with a multi-analyte blood test. Science 2018;359(6378):926–30. doi:10.1126/science.aar3247 [published Online First: 20180118]

17. Rozenblatt-Rosen O, Stubbington MJT, Regev A, et al. The Human Cell Atlas: from vision to reality. Nature 2017;550(7677):451–53. doi:10.1038/550451a

18. Landegren U, Hammond M. Cancer diagnostics based on plasma protein biomarkers: hard times but great expectations. Mol Oncol 2021;15(6):1715–26. doi:10.1002/1878-0261.12809 [published Online First: 20201117]

19. Robbins JM, Peterson B, Schranner D, et al. Human plasma proteomic profiles indicative of cardiorespiratory fitness. Nat Metab 2021;3(6):786–97. doi:10.1038/s42255-021-00400-z [published Online First: 20210527]

20. Petrera A, von Toerne C, Behler J, et al. Multiplatform Approach for Plasma Proteomics: Complementarity of Olink Proximity Extension Assay Technology to Mass Spectrometry-Based Protein Profiling. J Proteome Res 2021;20(1):751–62. doi:10.1021/acs.jproteome.0c00641 [published Online First: 20201130]

