## Supplementary Materials for "A novel proteomics-based plasma test for early detection of multiple cancers in the general population"

### Materials and Methods

#### *1. Study Design*

We collected plasma samples from 440 patients diagnosed with 18 distinct types of cancer, as well as from healthy individuals (Fig S1). We focused on early-stage common solid tumours where early detection followed by medical/surgical treatment can significantly enhances the survival of the patients. Then, we used Olink's Proximity Extension Assay technology to measure proteins in the plasma samples. Using the machine learning approaches, we identified protein biomarkers for detection of cancer as well as identification of the site of origin of the cancer.

#### *2. Sample Collection*

The EDTA plasma samples used in this analysis were provided by the Ukraine Association of Biobank (UAB). (*21*) A summary of the number and type of samples is outlined in Table S1. The UAB was established in 2017 with the goal of growing and developing the biobank network across Ukraine. The network encompasses biobanks from the main Institutes of Medical Sciences in the country and operates under the guidance of the ESBB, ISBER, and NCI guidelines. The UAB has established policies and necessary documents such as the Patient Consent Policy, Patient Information Sheet, Biobank Consent Form, and Sample Application Form to ensure ethical and transparent operations in medical institutions and hospitals.
UAB places a strong emphasis on maintaining the highest ethical standards in its operations and has implemented strict bioethical policies and state-of-the-art procedures. The anonymity of donors is protected, and they are fully informed about the purpose of medical research and the potential benefits of scientific discoveries through legal consent documentation. All specimens are obtained with the fully informed and signed consent of donors.
Every specimen collected by the UAB is processed following a rigorous Standard Operating Procedure (SOP). Quality control is conducted on all collected tissue samples, and the specimens are handled with utmost care and precision from the time of excision to storage and shipment to ensure that the customers receive only the highest quality specimens.

#### *3. Protein Measurement*

The protein levels in plasma were determined by utilizing the Olink® Explore 3072 technology. A full description of Olink's Proximity Extension Assay (PEA) technology has been reported elsewhere. The list of the proteins measured, and their characteristics are provided in the Table S2.
Briefly, this technology employs antibody-based detection to assess the levels of 3,072 target proteins in plasma. The antibodies were each conjugated with two complementary probes and divided into four separate 384-plex panels. Each panel included three control assays for quality control purposes (interleukin-6 (IL6), interleukin-8 (CXCL8), and tumour necrosis factor (TNF)). The process began with an overnight incubation to allow the conjugated antibodies to bind to the target proteins in the samples. This was followed by an extension and pre-amplification step, where the hybridization and extension of the complementary probes took place. The extended DNA was then amplified through PCR and indexed to prepare the libraries, which were sequenced using the Illumina NovaSeq platform. The counts obtained from the sequencing were subject to a quality control and normalization procedure, which involved the

use of internal controls to reduce intra-assay variability. These included an incubation control consisting of a non-human antigen, an extension control consisting of a unique pair of probes, and an amplification control consisting of a double-stranded DNA sequence. Additionally, external controls such as a negative control (buffer sample) and plate controls (pool of plasma) were used to determine the limit of detection and adjust levels between plates, respectively. Finally, two known samples were used as sample controls to calculate the precision of the measurements. After quality control and normalization, the data was provided in a Normalized Protein eXpression (NPX) unit, which is on a log2 scale and indicates a high protein level with a high NPX value.

The analytical performance of Olink's panels has been carefully validated for sensitivity, dynamic range, specificity, precision, and scalability (Table S2). Analytical measuring range was defined by the lower limit of quantification (LLOQ) and upper limit of quantification (ULOQ) and reported in pg/mL. The high dose hook effect (a state of antigen excess relative to the reagent antibodies resulting in falsely lower values) has also been determined for each analyte. All assays have been thoroughly validated for precision (repeatability and reproducibility). Intra-assay variation (within-run) has been calculated as the mean CV for 6 individual samples, within each of 7 separate runs during the validation studies. Inter-assay variation (between-runs) was calculated as the mean CV, for the same 6 individual samples, among 7 separate runs during the validation studies.

***4. Biomarker Discovery***

We transformed each problem into a binary classification one. For the pan-cancer detection problem, the positive class encompassed all cancer samples, and the control class included normal samples. For the localization of each cancer, the samples of that specific cancer were considered positive cases, while other cancer samples were considered negative cases.

We employed Logistic Regression with an L1 penalty to identify a sparse set of informative biomarkers. Each protein was standardized to zero mean and unit standard deviation to ensure comparable coefficients. The strength of the L1 penalty was determined using stratified 5-fold cross-validation on a logarithmic scale ranging from 1e-4 to 1e4. The desired number of biomarkers was then selected based on the magnitude of their absolute coefficients. The pre-processing and modelling were carried out using the Scikit-learn Python library. (*22*)

After identifying the biomarkers, linear classifiers were trained on the training data utilizing the selected biomarkers. In the pan-cancer detection component, the predicted probability for each sample is compared to a threshold to determine whether it is normal or cancerous. The threshold is set to attain the desired level of specificity. The positive samples from the detection component then proceed to the localization component, which consists of one model for each type of cancer. The sample is assigned to the class with the highest predicted probability.

For data visualization, we used matplotlib (version 3.6.0), (*23*) seaborn (version 0.12.0), (*24*) and plotly (version 5.10.0). (*25*) Pre-processing and modelling were performed in Python (version 3.9.13), using scikit-learn (version 1.1.2), (*22*) pandas (version 1.4.4), (*26*) numpy (version 1.23.3), (*27*) and statsmodels (version 0.13.2). (*28*)

***5. Biomarker Validation***

After biomarkers selection and to maximize the use of data in our training models, we used the leave-one-out method as our evaluation method. In this approach, the number of folds equals the number of instances in the data set. We applied the learning algorithm once for each instance, using all other instances as a training set and using the selected instance as a single-item test set.

Fig S1. Biomarker discovery workflow for early-stage cancers.

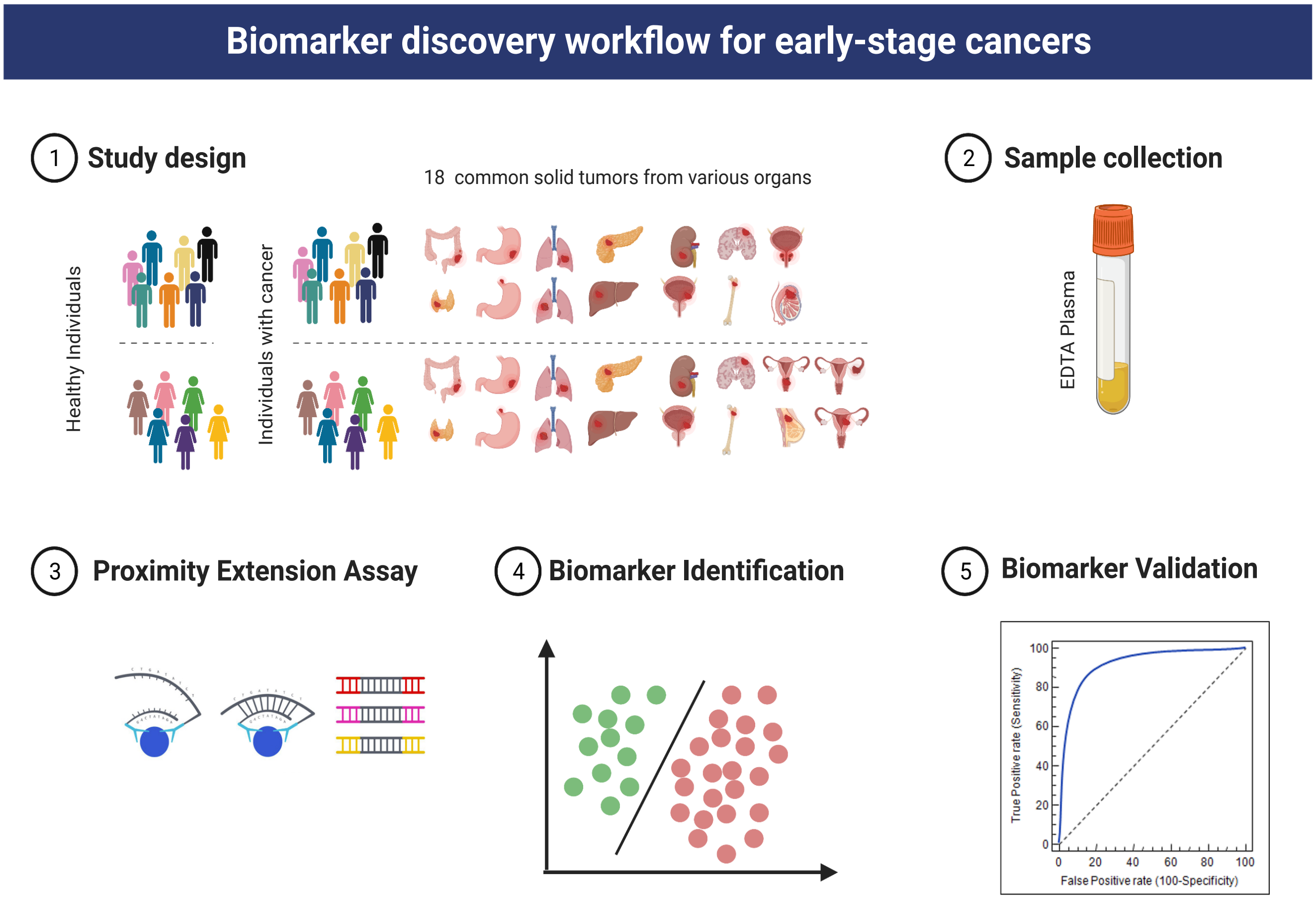

Fig S2. Distribution of proteins of the detection panel in tumour and normal plasma samples among males (a) and females (b).

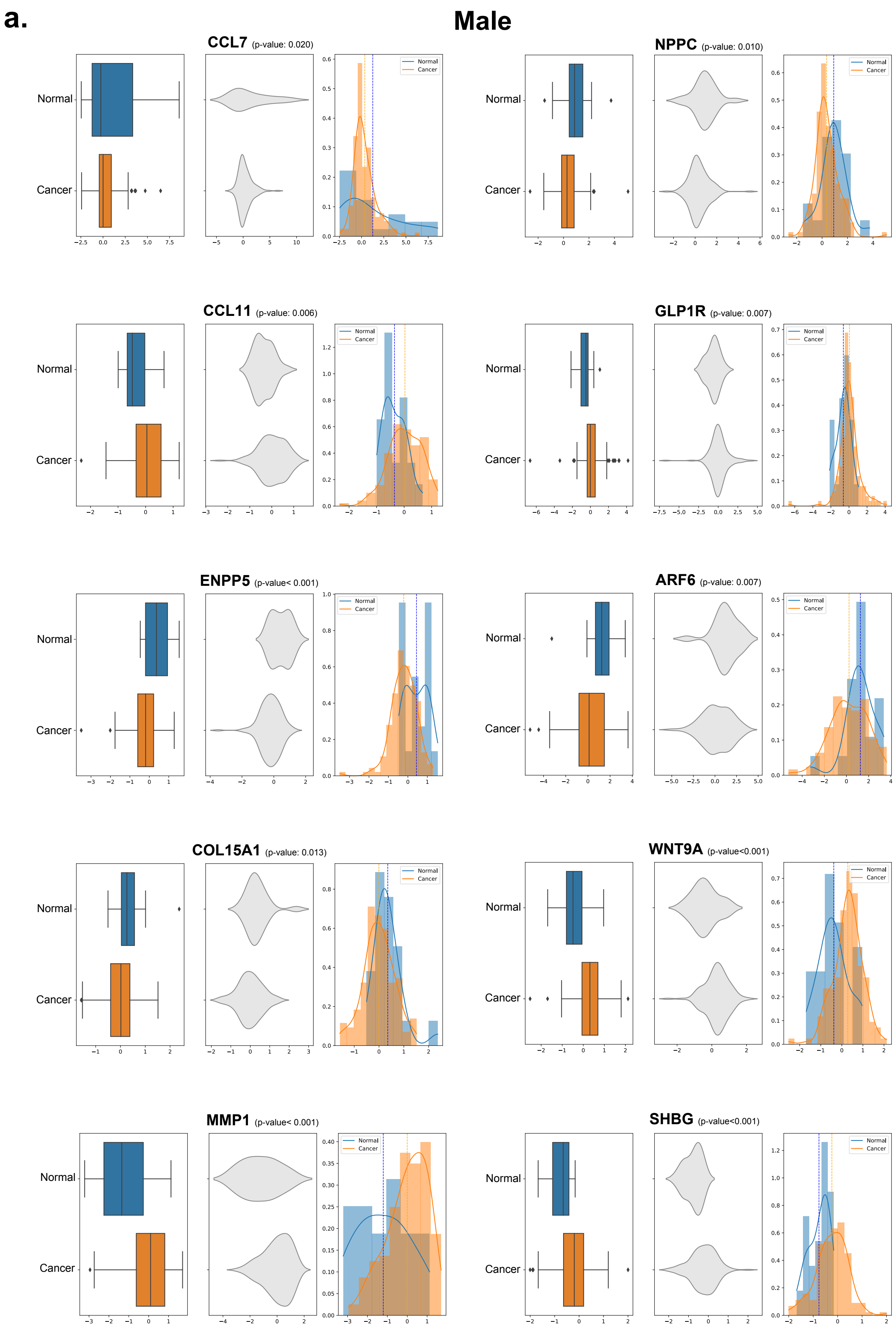

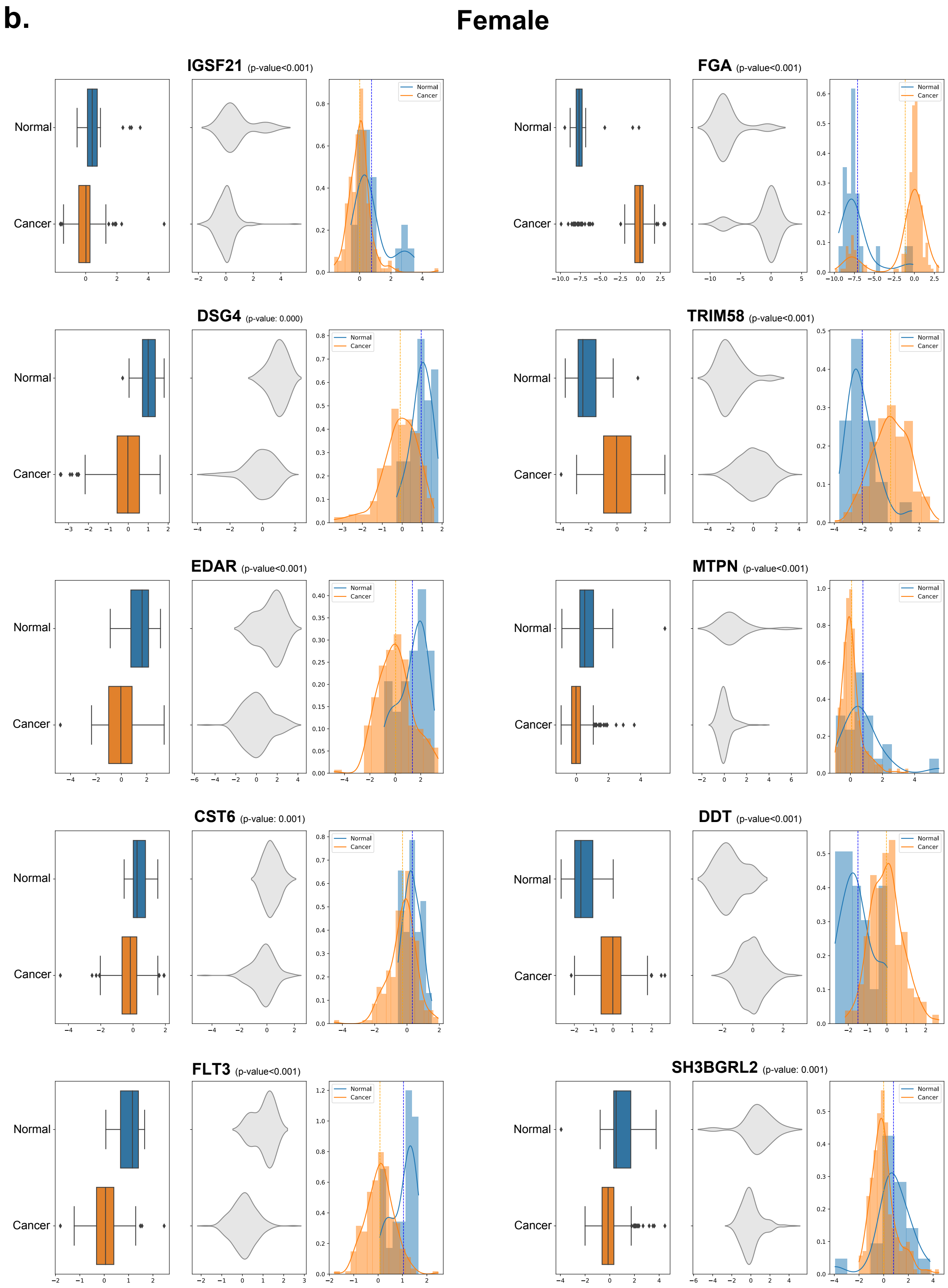
b.
Female
IGSF21 (p-value<0.001)
FGA (p-value<0.001)
DSG4 (p-value: 0.000)
TRIM58 (p-value<0.001)
EDAR (p-value<0.001)
MTPN (p-value<0.001)
CST6 (p-value: 0.001)
DDT (p-value<0.001)
FLT3 (p-value<0.001)
SH3BGRL2 (p-value: 0.001)
Normal
Cancer

Fig S3. The performance of the detection panel for unseen cancers among males (a) and females (b).

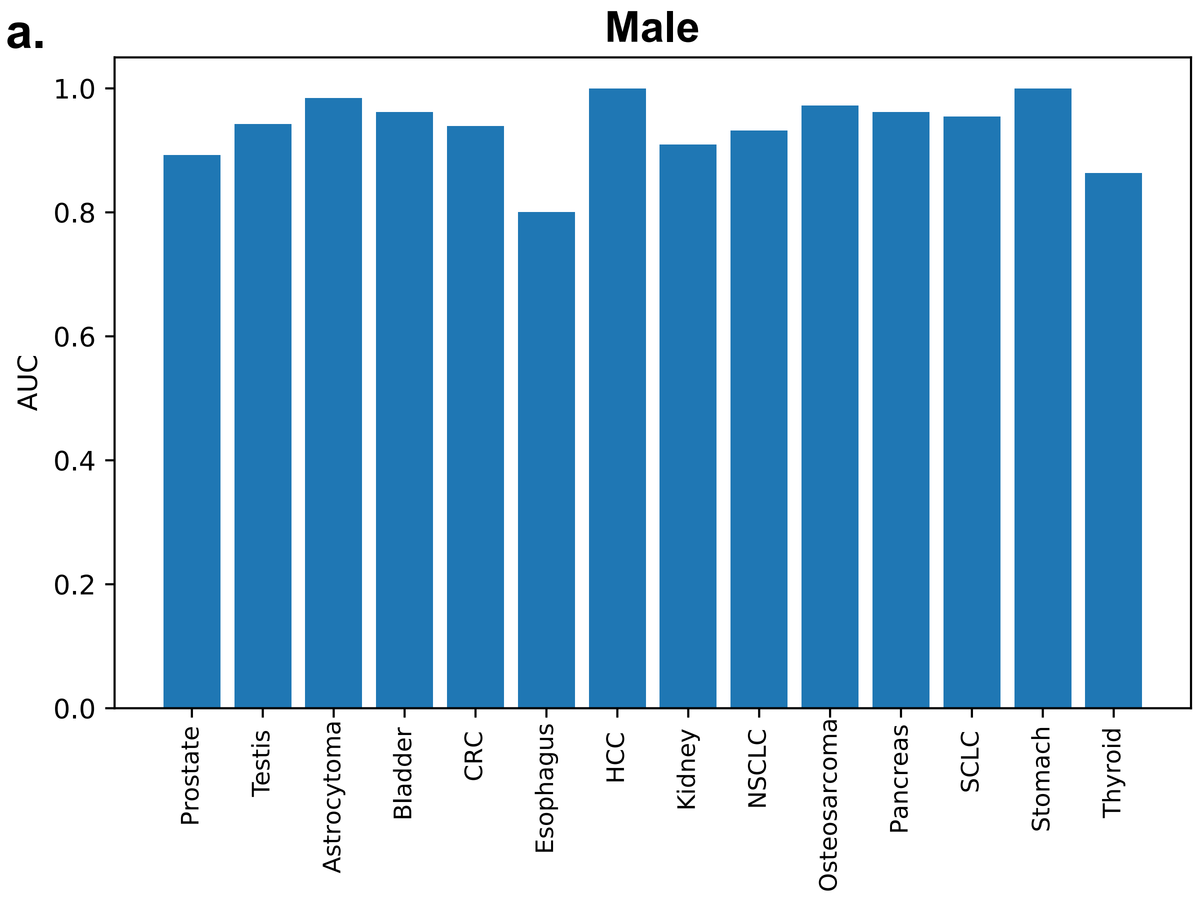


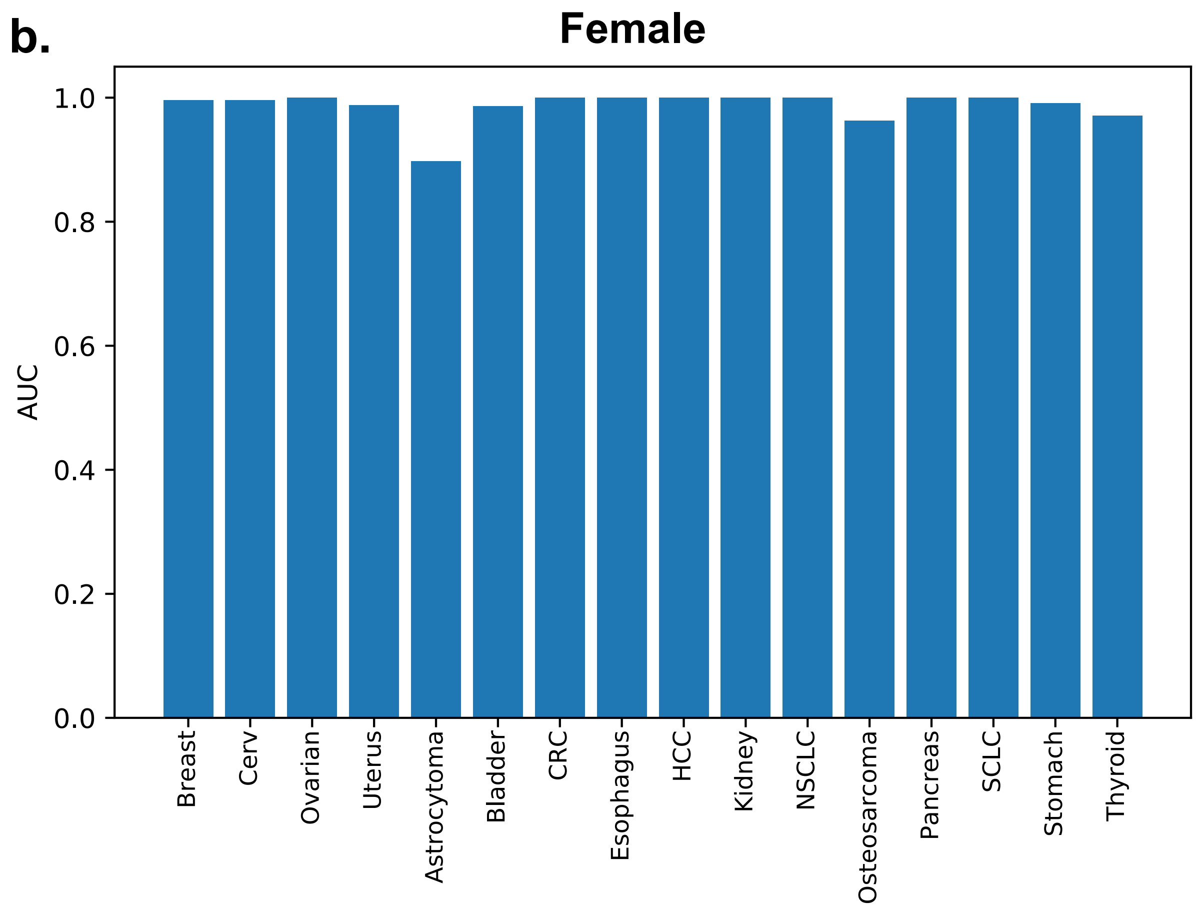

Fig S4. ROC curve (receiver operating characteristic curve) of localization panels by cancer for males (a) and females (b).

**a.**

### Male

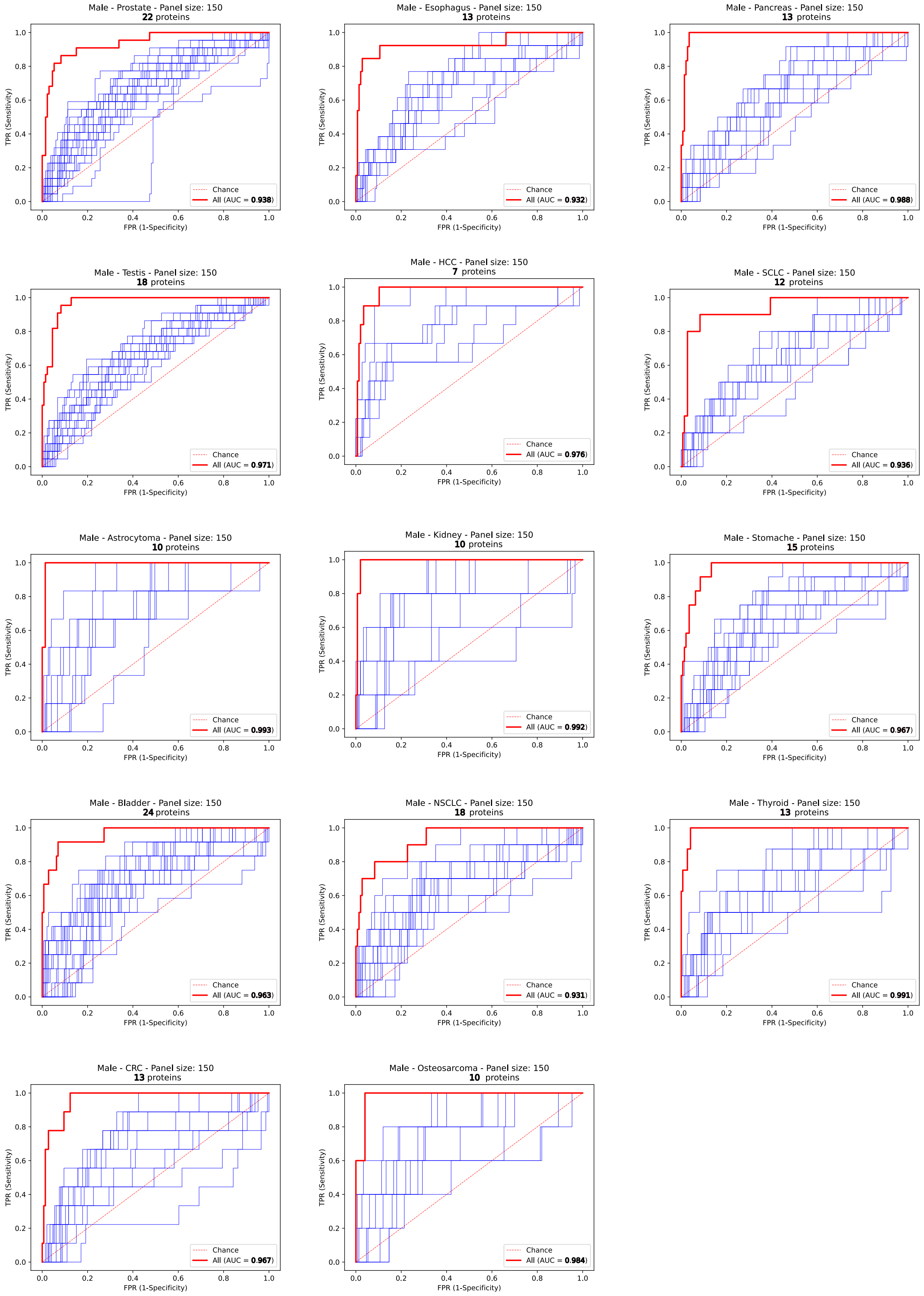

**b.**

### Female

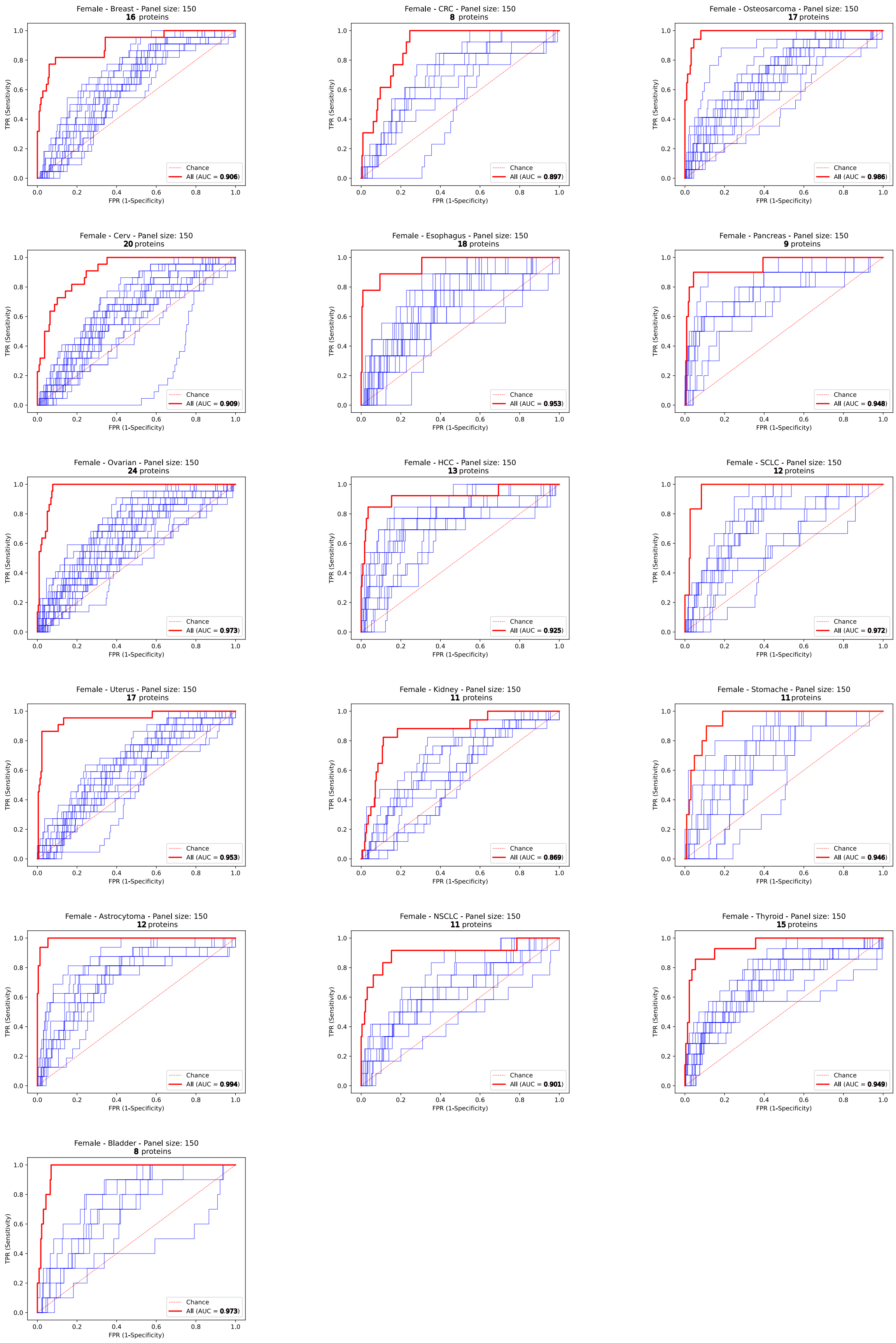

Female - Breast - Panel size: 150
16 proteins
All (AUC = 0.906)
Female - CRC - Panel size: 150
8 proteins
All (AUC = 0.897)
Female - Osteosarcoma - Panel size: 150
17 proteins
All (AUC = 0.986)
Female - Cerv - Panel size: 150
20 proteins
All (AUC = 0.909)
Female - Esophagus - Panel size: 150
18 proteins
All (AUC = 0.953)
Female - Pancreas - Panel size: 150
9 proteins
All (AUC = 0.948)
Female - Ovarian - Panel size: 150
24 proteins
All (AUC = 0.973)
Female - HCC - Panel size: 150
13 proteins
All (AUC = 0.925)
Female - SCLC - Panel size: 150
12 proteins
All (AUC = 0.972)
Female - Uterus - Panel size: 150
17 proteins
All (AUC = 0.953)
Female - Kidney - Panel size: 150
11 proteins
All (AUC = 0.869)
Female - Stomache - Panel size: 150
11 proteins
All (AUC = 0.946)
Female - Astrocytoma - Panel size: 150
12 proteins
All (AUC = 0.994)
Female - NSCLC - Panel size: 150
11 proteins
All (AUC = 0.901)
Female - Thyroid - Panel size: 150
15 proteins
All (AUC = 0.949)
Female - Bladder - Panel size: 150
8 proteins
All (AUC = 0.973)
Chance
TPR (Sensitivity)
FPR (1-Specificity)

**Table S1.** Characteristics of the samples included in the study.

| Cancer | Subtype | Number | Sex (% Male) | Age | Treatment history |
|---|---|---|---|---|---|
| Bladder | Urothelial carcinoma | 22 | 55% | 63.1 | Treatment Naive |
| Breast cancer | Invasive ductal carcinoma | 22 | 0% | 46.3 | Treatment Naive |
| Brain | Astrocytoma | 22 | 27% | 47.1 | Treatment Naive |
| Cervical | Cervix uteri carcinoma | 22 | 0% | 50.1 | Treatment Naive |
| Colorectal | Adenocarcinoma | 22 | 41% | 57.8 | Treatment Naive |
| Kidney | Kidney renal papillary cell carcinoma | 22 | 23% | 57.5 | Treatment Naive |
| Liver | Hepatocellular carcinoma | 22 | 41% | 56.7 | Treatment Naive |
| Lung | Non-Small Cell (NSCLC) | 22 | 45% | 58 | Treatment Naive |
| Lung | Small Cell (SCLC) | 22 | 45% | 57.2 | Treatment Naive |
| Oesophagus | squamous cell carcinoma | 22 | 59% | 61.4 | Treatment Naive |
| Osteosarcoma | osteoblastic osteosarcoma | 22 | 23% | 40.4 | Treatment Naive |
| Ovarian | Epithelial | 22 | 0% | 53.7 | Treatment Naive |
| Pancreas | Ductal adenocarcinoma | 22 | 55% | 62.7 | Treatment Naive |
| Prostate | Adenocarcinoma | 22 | 100% | 67.4 | Treatment Naive |
| Stomach | Adenocarcinoma | 22 | 55% | 69.2 | Treatment Naive |
| Testis | Seminoma | 22 | 100% | 52.2 | Treatment Naive |
| Thyroid | medullary thyroid cancer | 22 | 36% | 52.6 | Treatment Naive |
| Uterus | Endometrial Cancer | 22 | 0% | 52 | Treatment Naive |
| Normal | | 22 | 100% | 59.6 | |
| Normal | | 22 | 0% | 49.7 | |

**Table S2.** List of proteins included in the analysis.

| Protein | Dilution factor | LOD (pg/ml) | LLOQ (pg/ml) | ULOQ (pg/ml) | Hook (pg/mL) | Range (log10) | Intra-CV (%) | Inter-CV (%) |
|---|---|---|---|---|---|---|---|---|
| TIA1 | 1:1 | | | | | | 9 | 27 |
| COMT | 1:1 | 97.7 | 195.3 | 100000 | 800000 | 2.7 | 8 | 10 |
| DUOX2 | 1:1 | 97.7 | 195.3 | 100000 | 200000 | 2.7 | 7 | 31 |
| NPPB | 1:1 | | | | | | 10 | 18 |
| TSPAN1 | 1:1 | | | | | | 7 | 9 |
| CHEK2 | 1:1 | | | | | | 13 | 19 |
| RCOR1 | 1:1 | 1.5 | 3.1 | 3125 | 12500 | 3.0 | 7 | 12 |
| DNAJB8 | 1:1 | | | | | | 9 | 10 |
| MTPN | 1:1 | | | | | | 8 | 8 |
| EDIL3 | 1:1 | 24.4 | 48.8 | 200000 | 800000 | 3.6 | 8 | 13 |
| THPO | 1:1 | 97.7 | 781.3 | 200000 | 400000 | 2.4 | 9 | 14 |
| MPHOSPH8 | 1:1 | 48.8 | 97.7 | 12500 | 25000 | 2.1 | 8 | 19 |
| ADH4 | 1:1 | | | | | | 7 | 8 |
| S100P | 1:1 | | | | | | 12 | 15 |
| ADAMTS16 | 1:1 | 6.1 | 24.4 | 25000 | 200000 | 3.0 | 9 | 6 |
| GLO1 | 1:1 | | | | | | 7 | 9 |
| ACE2 | 1:1 | 48.8 | 97.7 | 100000 | 400000 | 3.0 | 9 | 10 |
| MSTN | 1:1 | 390.6 | 781.3 | 50000 | 400000 | 1.8 | 10 | 14 |
| PAG1 | 1:1 | 24.4 | 48.8 | 6250 | 200000 | 2.1 | 11 | 11 |
| ADAM15 | 1:1 | 24.4 | 48.8 | 25000 | 200000 | 2.7 | 7 | 10 |
| EPHX2 | 1:1 | 390.6 | 781.3 | 100000 | 200000 | 2.1 | 7 | 21 |
| CLTA | 1:1 | 3125.0 | 6250.0 | 6400000 | 12800000 | 3.0 | 7 | 18 |
| GRK5 | 1:1 | 1562.5 | 3125.0 | 400000 | 800000 | 2.1 | 8 | 14 |
| GP2 | 1:1 | 3.1 | 6.1 | 12500 | 200000 | 3.3 | 6 | 9 |
| TNF | 1:1 | 6.1 | 12.2 | 6250 | 200000 | 2.7 | 8 | 16 |
| HK2 | 1:1 | 97.7 | 195.3 | 50000 | 200000 | 2.4 | 9 | 39 |
| CTSH | 1:1 | | | | | | 11 | 10 |
| ENTPD6 | 1:1 | 195.3 | 781.3 | 50000 | 800000 | 1.8 | 8 | 8 |
| BOC | 1:1 | | | | | | 8 | 10 |
| BMP6 | 1:1 | 0.001 | 97.7 | 12500 | 200000 | 2.1 | 13 | 16 |
| IL6 | 1:1 | 0.4 | 0.8 | 3125 | 12500 | 3.6 | 6 | 12 |
| BAG6 | 1:1 | 390.6 | 781.3 | 200000 | 400000 | 2.4 | 8 | 10 |
| USP8 | 1:1 | 3125.0 | 6250.0 | 800000 | 800000 | 2.1 | 8 | 13 |
| ACTA2 | 1:1 | | | | | | 7 | 8 |
| FABP6 | 1:1 | | | | | | 10 | 24 |
| MMP7 | 1:1 | 97.7 | 97.7 | 6250 | 50000 | 1.8 | 6 | 8 |
| PON2 | 1:1 | | | | | | 8 | 8 |
| WASF1 | 1:1 | 6250.0 | 12500.0 | 800000 | 1600000 | 1.8 | 10 | 19 |
| CRX | 1:1 | | | | | | 8 | 13 |
| VIM | 1:1 | 12500.0 | 12500.0 | 1600000 | 3200000 | 2.1 | 6 | 60 |
| DCN | 1:1 | 195.3 | 390.6 | 100000 | 200000 | 2.4 | 7 | 11 |
| STK11 | 1:1 | 390.6 | 781.3 | 400000 | 800000 | 2.7 | 10 | 21 |
| TNNI3 | 1:1 | 97.7 | 195.3 | 25000 | 200000 | 2.1 | 15 | 14 |

| | | | | | | | | |
|---|---|---|---|---|---|---|---|---|
| ITGB1BP2 | 1:1 | 195.3 | 781.3 | 100000 | 200000 | 2.1 | 9 | 18 |
| TCL1B | 1:1 | 97.7 | 195.3 | 200000 | 800000 | 3.0 | 8 | 24 |
| SERPINB5 | 1:1 | 781.3 | 1562.5 | 100000 | 200000 | 1.8 | 11 | 21 |
| CTF1 | 1:1 | 195.3 | 781.3 | 100000 | 400000 | 2.1 | 11 | 22 |
| HNRNPK | 1:1 | 97.7 | 97.7 | 50000 | 200000 | 2.7 | 8 | 12 |
| CEBPB | 1:1 | | | | | | 11 | 41 |
| VSTM2L | 1:1 | | | | | | 8 | 11 |
| ZBTB17 | 1:1 | 6.1 | 12.2 | 6250 | 200000 | 2.7 | 8 | 8 |
| CEP43 | 1:1 | 781.3 | 1562.5 | 100000 | 3200000 | 1.8 | 10 | 24 |
| PTN | 1:1 | | | | | | 20 | 17 |
| CRHR1 | 1:1 | 24.4 | 48.8 | 6250 | 25000 | 2.1 | 7 | 24 |
| SUSD1 | 1:1 | 48.8 | 97.7 | 12500 | 200000 | 2.1 | 8 | 14 |
| TSLP | 1:1 | 6.1 | 48.8 | 12500 | 200000 | 2.4 | 7 | 14 |
| CA5A | 1:1 | 1.5 | 6.1 | 6250 | 25000 | 3.0 | 7 | 9 |
| MFAP3 | 1:1 | 24.4 | 48.8 | 100000 | 800000 | 3.3 | 9 | 26 |
| AKR1C4 | 1:1 | 390.6 | 781.3 | 100000 | 400000 | 2.1 | 12 | 17 |
| GPR37 | 1:1 | 781.3 | 1562.5 | 100000 | 400000 | 1.8 | 9 | 11 |
| IRAG2 | 1:1 | 48.8 | 97.7 | 12500 | 50000 | 2.1 | 8 | 12 |
| CEACAM8 | 1:1 | | | | | | 9 | 12 |
| HEBP1 | 1:1 | | | | | | 8 | 12 |
| CNPY2 | 1:1 | | | | | | 7 | 10 |
| VAMP5 | 1:1 | 390.6 | 781.3 | 200000 | 400000 | 2.4 | 8 | 30 |
| GYS1 | 1:1 | 12.2 | 48.8 | 25000 | 200000 | 2.7 | 8 | 16 |
| GZMH | 1:1 | | | | | | 8 | 12 |
| SLITRK6 | 1:1 | 24.4 | 48.8 | 100000 | 400000 | 3.3 | 7 | 14 |
| CLEC1A | 1:1 | 97.7 | 195.3 | 25000 | 400000 | 2.1 | 7 | 10 |
| ENTPD5 | 1:1 | 48.8 | 97.7 | 100000 | 800000 | 3.0 | 8 | 8 |
| TP53INP1 | 1:1 | 97.7 | 195.3 | 25000 | 200000 | 2.1 | 8 | 8 |
| TINAGL1 | 1:10 | 1.5 | 3.1 | 6250 | 50000 | 3.3 | 10 | 6 |
| CBLIF | 1:10 | 0.2 | 0.4 | 6250 | 50000 | 4.2 | 9 | 6 |
| FABP2 | 1:10 | 0.4 | 1.5 | 781 | 6250 | 2.7 | 9 | 8 |
| NPDC1 | 1:10 | 6.1 | 12.2 | 3125 | 50000 | 2.4 | 10 | 6 |
| RNASE3 | 1:10 | | | | | | 12 | 18 |
| GHRL | 1:10 | 24.4 | 48.8 | 50000 | 200000 | 3.0 | 8 | 13 |
| GLRX | 1:10 | | | | | | 9 | 11 |
| LEP | 1:10 | 12.2 | 48.8 | 12500 | 200000 | 2.4 | 7 | 11 |
| LGALS1 | 1:10 | 24.4 | 48.8 | 50000 | 200000 | 3.0 | 12 | 15 |
| IL6ST | 1:10 | 390.6 | 781.3 | 200000 | 800000 | 2.4 | 7 | 7 |
| NECTIN2 | 1:10 | 0.1 | 0.1 | 195 | 781 | 3.3 | 9 | 6 |
| ACOX1 | 1:10 | | | | | | 9 | 15 |
| NTRK2 | 1:10 | 48.8 | 48.8 | 6250 | 50000 | 2.1 | 10 | 10 |
| KITLG | 1:10 | 3.1 | 6.1 | 12500 | 50000 | 3.3 | 10 | 8 |
| SDC4 | 1:10 | 1.5 | 3.1 | 3125 | 200000 | 3.0 | 9 | 6 |
| QDPR | 1:10 | 12.2 | 24.4 | 50000 | 800000 | 3.3 | 7 | 7 |
| CANT1 | 1:10 | 6.1 | 12.2 | 6250 | 50000 | 2.7 | 8 | 5 |
| CLUL1 | 1:10 | 6.1 | 12.2 | 6250 | 50000 | 2.7 | 9 | 12 |
| PILRB | 1:10 | 0.8 | 1.5 | 3125 | 12500 | 3.3 | 7 | 6 |
| SNAP23 | 1:10 | 6.1 | 12.2 | 12500 | 50000 | 3.0 | 12 | 13 |
| CHRDL2 | 1:10 | 195.3 | 390.6 | 50000 | 200000 | 2.1 | 10 | 7 |
| NRCAM | 1:10 | 6.1 | 12.2 | 6250 | 12500 | 2.7 | 15 | 16 |
| AK1 | 1:10 | | | | | | 9 | 10 |
| STK4 | 1:10 | 195.3 | 390.6 | 6250 | 50000 | 1.2 | 6 | 28 |

| | | | | | | | | |
|---|---|---|---|---|---|---|---|---|
| ANXA4 | 1:10 | 97.7 | 195.3 | 25000 | 800000 | 2.1 | 7 | 14 |
| CLC | 1:10 | 390.6 | 781.3 | 50000 | 800000 | 1.8 | 10 | 8 |
| DPP7 | 1:10 | 97.7 | 195.3 | 50000 | 400000 | 2.4 | 11 | 15 |
| ACY1 | 1:10 | 390.6 | 781.3 | 400000 | 800000 | 2.7 | 10 | 11 |
| PRKAR1A | 1:10 | 24.4 | 48.8 | 100000 | 200000 | 3.3 | 12 | 12 |
| PLPBP | 1:10 | 12.2 | 24.4 | 12500 | 50000 | 2.7 | 9 | 12 |
| PDGFRA | 1:10 | 12.2 | 48.8 | 6250 | 50000 | 2.1 | 10 | 6 |
| DCTPP1 | 1:10 | 12.2 | 24.4 | 12500 | 50000 | 2.7 | 9 | 9 |
| PCDH17 | 1:10 | 24.4 | 48.8 | 50000 | 400000 | 3.0 | 13 | 10 |
| VSIR | 1:10 | 1.5 | 3.1 | 1563 | 6250 | 2.7 | 10 | 14 |
| NT-proBNP | 1:10 | 97.7 | 195.3 | 50000 | 200000 | 2.4 | 11 | 9 |
| S100A11 | 1:10 | 6.1 | 6.1 | 3125 | 12500 | 2.7 | 14 | 10 |
| CCL27 | 1:10 | | | | | | 17 | 14 |
| TSHB | 1:10 | 1.5 | 3.1 | 1563 | 3125 | 2.7 | 11 | 7 |
| TGM2 | 1:10 | 6.1 | 24.4 | 50000 | 200000 | 3.3 | 15 | 11 |
| AGXT | 1:10 | 390.6 | 781.3 | 1600000 | 12800000 | 3.3 | 9 | 10 |
| ICAM5 | 1:10 | 12.2 | 24.4 | 6250 | 50000 | 2.4 | 9 | 9 |
| PM20D1 | 1:10 | 48.8 | 97.7 | 100000 | 800000 | 3.0 | 11 | 13 |
| CLEC5A | 1:10 | 0.1 | 0.4 | 3125 | 6250 | 3.9 | 9 | 8 |
| CDHR5 | 1:10 | 12.2 | 48.8 | 25000 | 200000 | 2.7 | 10 | 7 |
| ACAN | 1:10 | 12.2 | 24.4 | 50000 | 200000 | 3.3 | 12 | 9 |
| CDH6 | 1:10 | 97.7 | 195.3 | 25000 | 200000 | 2.1 | 12 | 10 |
| PLIN3 | 1:10 | 195.3 | 390.6 | 50000 | 800000 | 2.1 | 12 | 13 |
| GSTA1 | 1:10 | 3.1 | 6.1 | 12500 | 400000 | 3.3 | 9 | 7 |
| THOP1 | 1:10 | 6.1 | 12.2 | 6250 | 200000 | 2.7 | 9 | 5 |
| IGSF8 | 1:10 | 12.2 | 12.2 | 25000 | 200000 | 3.3 | 9 | 9 |
| PDCD6 | 1:10 | 1562.5 | 1562.5 | 100000 | 800000 | 1.8 | 8 | 10 |
| PLXNB3 | 1:10 | 24.4 | 48.8 | 50000 | 200000 | 3.0 | 10 | 13 |
| MNDA | 1:10 | 390.6 | 781.3 | 800000 | 800000 | 3.0 | 9 | 10 |
| LEPR | 1:10 | | | | | | 8 | 12 |
| SIGLEC7 | 1:10 | 3.1 | 3.1 | 6250 | 50000 | 3.3 | 8 | 8 |
| APLP1 | 1:10 | 195.3 | 195.3 | 50000 | 400000 | 2.4 | 12 | 15 |
| NPTXR | 1:10 | 6.1 | 12.2 | 25000 | 50000 | 3.3 | 10 | 7 |
| GRAP2 | 1:10 | 1562.5 | 3125.0 | 400000 | 800000 | 2.1 | 8 | 10 |
| TYRO3 | 1:10 | 3.1 | 6.1 | 1563 | 6250 | 2.4 | 9 | 10 |
| CDH17 | 1:10 | 195.3 | 390.6 | 50000 | 400000 | 2.1 | 14 | 13 |
| SNX9 | 1:10 | 390.6 | 781.3 | 400000 | 800000 | 2.7 | 9 | 11 |
| EIF4EBP1 | 1:10 | 24.4 | 24.4 | 12500 | 50000 | 2.7 | 8 | 8 |
| IL19 | 1:10 | 3.1 | 12.2 | 100000 | 400000 | 3.9 | 9 | 7 |
| IGFBPL1 | 1:10 | 0.001 | 1.5 | 3125 | 6250 | 3.3 | 9 | 9 |
| SSC4D | 1:10 | 6.1 | 6.1 | 6250 | 200000 | 3.0 | 7 | 8 |
| OLR1 | 1:10 | 3.1 | 3.1 | 3125 | 12500 | 3.0 | 9 | 8 |
| CGREF1 | 1:10 | 24.4 | 48.8 | 6250 | 50000 | 2.1 | 8 | 7 |
| MCFD2 | 1:10 | 48.8 | 97.7 | 12500 | 400000 | 2.1 | 9 | 7 |
| AHCY | 1:10 | 195.3 | 390.6 | 100000 | 400000 | 2.4 | 8 | 13 |
| SERPINA12 | 1:10 | 24.4 | 97.7 | 6250 | 50000 | 1.8 | 9 | 14 |
| HMOX1 | 1:10 | 3.1 | 3.1 | 6250 | 25000 | 3.3 | 9 | 13 |
| PRSS27 | 1:10 | 12.2 | 24.4 | 25000 | 50000 | 3.0 | 8 | 6 |
| CNST | 1:10 | 12.2 | 48.8 | 6250 | 50000 | 2.1 | 10 | 21 |
| ADGRG2 | 1:10 | 12.2 | 48.8 | 12500 | 50000 | 2.4 | 10 | 8 |
| LPL | 1:10 | 195.3 | 195.3 | 400000 | 800000 | 3.3 | 8 | 11 |
| FADD | 1:10 | 781.3 | 3125.0 | 800000 | 800000 | 2.4 | 9 | 10 |

| | | | | | | | | |
|---|---|---|---|---|---|---|---|---|
| DIABLO | 1:10 | 3.1 | 6.1 | 12500 | 50000 | 3.3 | 12 | 28 |
| LRP11 | 1:10 | 12.2 | 48.8 | 6250 | 200000 | 2.1 | 11 | 7 |
| SFTPD | 1:10 | 6.1 | 12.2 | 6250 | 50000 | 2.7 | 8 | 5 |
| NADK | 1:10 | 62.5 | 125.0 | 256000 | 512000 | 3.3 | 10 | 6 |
| GPNMB | 1:10 | 24.4 | 48.8 | 12500 | 200000 | 2.4 | 12 | 10 |
| SDC1 | 1:10 | 6.1 | 12.2 | 6250 | 200000 | 2.7 | 14 | 12 |
| CXCL8 | 1:10 | 0.1 | 0.2 | 781 | 6250 | 3.6 | 10 | 15 |
| LACTB2 | 1:10 | 48.8 | 97.7 | 25000 | 200000 | 2.4 | 8 | 9 |
| SPON2 | 1:10 | 24.4 | 48.8 | 25000 | 400000 | 2.7 | 9 | 8 |
| MEP1B | 1:10 | 97.7 | 195.3 | 25000 | 50000 | 2.1 | 9 | 9 |
| CD2AP | 1:10 | 6.1 | 12.2 | 12500 | 50000 | 3.0 | 10 | 9 |
| PPP1R2 | 1:10 | 12.2 | 24.4 | 12500 | 50000 | 2.7 | 9 | 10 |
| SEMA3F | 1:10 | 148.4 | 296.9 | 38000 | 304000 | 2.1 | 9 | 8 |
| FCRL1 | 1:10 | 97.7 | 195.3 | 400000 | 800000 | 3.3 | 10 | 7 |
| CDH2 | 1:10 | 390.6 | 781.3 | 200000 | 800000 | 2.4 | 11 | 9 |
| REN | 1:10 | 6.1 | 12.2 | 12500 | 50000 | 3.0 | 9 | 9 |
| CA13 | 1:10 | 6.1 | 24.4 | 12500 | 50000 | 2.7 | 8 | 15 |
| CD69 | 1:10 | 0.8 | 3.1 | 12500 | 50000 | 3.6 | 12 | 14 |
| GDF2 | 1:10 | 1.5 | 6.1 | 12500 | 50000 | 3.3 | 10 | 8 |
| ANGPTL1 | 1:10 | 97.7 | 195.3 | 100000 | 400000 | 2.7 | 9 | 7 |
| MARCO | 1:10 | 24.4 | 24.4 | 50000 | 800000 | 3.3 | 7 | 6 |
| COL4A1 | 1:10 | 0.8 | 3.1 | 50000 | 200000 | 4.2 | 10 | 6 |
| THBD | 1:10 | 3.1 | 6.1 | 6250 | 50000 | 3.0 | 9 | 8 |
| GH1 | 1:10 | | | | | | 7 | 15 |
| LILRA5 | 1:10 | 0.8 | 1.5 | 3125 | 6250 | 3.3 | 8 | 7 |
| ROR1 | 1:10 | 24.4 | 48.8 | 3125 | 12500 | 1.8 | 10 | 7 |
| KYAT1 | 1:10 | 0.8 | 1.5 | 3125 | 12500 | 3.3 | 12 | 8 |
| FBP1 | 1:10 | 97.7 | 195.3 | 50000 | 200000 | 2.4 | 8 | 40 |
| CXCL5 | 1:10 | 3.1 | 6.1 | 3125 | 6250 | 2.7 | 11 | 10 |
| SOST | 1:10 | 6.1 | 6.1 | 25000 | 50000 | 3.6 | 8 | 11 |
| CCDC80 | 1:10 | 24.4 | 48.8 | 50000 | 800000 | 3.0 | 10 | 8 |
| TYMP | 1:10 | 390.6 | 781.3 | 200000 | 800000 | 2.4 | 9 | 17 |
| FAM3C | 1:10 | 3.1 | 6.1 | 6250 | 50000 | 3.0 | 8 | 7 |
| CTSL | 1:10 | 24.4 | 97.7 | 50000 | 200000 | 2.7 | 10 | 9 |
| HSPB1 | 1:10 | | | | | | 15 | 28 |
| SORT1 | 1:10 | 24.4 | 24.4 | 50000 | 400000 | 3.3 | 9 | 8 |
| DDC | 1:10 | 3.1 | 6.1 | 25000 | 50000 | 3.6 | 8 | 7 |
| DOK2 | 1:10 | 6.1 | 12.2 | 25000 | 200000 | 3.3 | 10 | 15 |
| UMOD | 1:100 | 1.5 | 3.1 | 12500 | 100000 | 3.6 | 4 | 3 |
| MFAP5 | 1:100 | 12.2 | 24.4 | 12500 | 100000 | 2.7 | 6 | 9 |
| PLAT | 1:100 | 0.8 | 3.1 | 25000 | 100000 | 3.9 | 8 | 8 |
| SEMA7A | 1:100 | 0.8 | 1.5 | 12500 | 50000 | 3.9 | 5 | 5 |
| CES1 | 1:100 | 24.4 | 48.8 | 400000 | 800000 | 3.9 | 5 | 7 |
| CA4 | 1:100 | 0.8 | 1.5 | 12500 | 100000 | 3.9 | 5 | 6 |
| PLTP | 1:100 | 6.1 | 12.2 | 25000 | 400000 | 3.3 | 5 | 11 |
| LDLR | 1:100 | 0.8 | 1.5 | 25000 | 100000 | 4.2 | 4 | 14 |
| ICAM2 | 1:100 | 97.7 | 195.3 | 200000 | 400000 | 3.0 | 6 | 9 |
| PCSK9 | 1:100 | 97.7 | 195.3 | 400000 | 800000 | 3.3 | 5 | 28 |
| FABP4 | 1:100 | 48.8 | 195.3 | 50000 | 200000 | 2.4 | 5 | 5 |
| ADAMTS13 | 1:100 | 6.1 | 12.2 | 6250 | 100000 | 2.7 | 6 | 9 |
| IGFBP1 | 1:100 | 1.5 | 3.1 | 12500 | 100000 | 3.6 | 3 | 3 |
| CDH5 | 1:100 | 195.3 | 390.6 | 200000 | 400000 | 2.7 | 5 | 8 |

| | | | | | | | | |
|---|---|---|---|---|---|---|---|---|
| IGFBP7 | 1:100 | 12.2 | 24.4 | 12500 | 200000 | 2.7 | 7 | 8 |
| EPHB4 | 1:100 | 1.5 | 3.1 | 12500 | 100000 | 3.6 | 4 | 5 |
| ESAM | 1:100 | 3.1 | 6.1 | 3125 | 12500 | 2.7 | 4 | 5 |
| ICAM3 | 1:100 | 1.5 | 1.5 | 3125 | 12500 | 3.3 | 3 | 4 |
| MSMB | 1:100 | 0.001 | 6.1 | 6250 | 12500 | 3.0 | 3 | 5 |
| REG3A | 1:100 | 0.4 | 0.8 | 1563 | 12500 | 3.3 | 4 | 5 |
| IL2RA | 1:100 | 0.02 | 0.0 | 391 | 3125 | 3.9 | 5 | 5 |
| CTSB | 1:100 | 12.2 | 24.4 | 12500 | 25000 | 2.7 | 4 | 4 |
| DKK3 | 1:100 | 6.1 | 12.2 | 25000 | 100000 | 3.3 | 5 | 5 |
| SERPINA11 | 1:100 | 3.1 | 6.1 | 12500 | 100000 | 3.3 | 4 | 14 |
| FUCA1 | 1:100 | 6.1 | 12.2 | 50000 | 200000 | 3.6 | 6 | 13 |
| LTBP2 | 1:100 | 6.1 | 12.2 | 25000 | 200000 | 3.3 | 4 | 5 |
| CRTAC1 | 1:100 | 6.1 | 24.4 | 100000 | 800000 | 3.6 | 6 | 8 |
| TNFRSF10C | 1:100 | 0.4 | 0.8 | 6250 | 12500 | 3.9 | 5 | 5 |
| PEAR1 | 1:100 | 12.2 | 24.4 | 12500 | 100000 | 2.7 | 5 | 4 |
| REG1B | 1:100 | 0.8 | 1.5 | 1563 | 12500 | 3.0 | 4 | 4 |
| CPA1 | 1:100 | 0.4 | 0.8 | 3125 | 12500 | 3.6 | 6 | 5 |
| DPT | 1:100 | 3.1 | 6.1 | 6250 | 400000 | 3.0 | 6 | 7 |
| LGALS3 | 1:100 | 12.2 | 48.8 | 12500 | 50000 | 2.4 | 5 | 5 |
| VWF | 1:100 | 1.5 | 3.1 | 12500 | 100000 | 3.6 | 9 | 32 |
| XG | 1:100 | 1.5 | 3.1 | 6250 | 12500 | 3.3 | 6 | 10 |
| TGFBR3 | 1:100 | 12.2 | 24.4 | 100000 | 200000 | 3.6 | 5 | 8 |
| PLA2G2A | 1:100 | | | | | | 6 | 9 |
| TNFSF13B | 1:100 | 0.2 | 0.8 | 12500 | 50000 | 4.2 | 4 | 4 |
| MET | 1:100 | 6.1 | 12.2 | 12500 | 50000 | 3.0 | 4 | 6 |
| CXCL16 | 1:100 | 0.8 | 1.5 | 12500 | 50000 | 3.9 | 5 | 6 |
| NOTCH3 | 1:100 | 1.5 | 3.1 | 6250 | 50000 | 3.3 | 6 | 8 |
| CA3 | 1:100 | 1562.5 | 12500.0 | 12800000 | 12800000 | 3.0 | 4 | 5 |
| PDGFRB | 1:100 | 3.1 | 6.1 | 3125 | 12500 | 2.7 | 4 | 4 |
| DLK1 | 1:100 | 0.8 | 3.1 | 6250 | 25000 | 3.3 | 4 | 5 |
| SCARF1 | 1:100 | 0.8 | 1.5 | 3125 | 12500 | 3.3 | 5 | 6 |
| GDF15 | 1:100 | 1.5 | 3.1 | 6250 | 25000 | 3.3 | 5 | 4 |
| CSTB | 1:100 | 3.1 | 6.1 | 3125 | 12500 | 2.7 | 4 | 3 |
| MB | 1:100 | 0.8 | 0.8 | 195 | 3125 | 2.4 | 4 | 5 |
| ENPP2 | 1:100 | 390.6 | 781.3 | 100000 | 800000 | 2.1 | 4 | 5 |
| GUSB | 1:100 | 3.1 | 6.1 | 6250 | 50000 | 3.0 | 4 | 6 |
| IL1RL1 | 1:100 | 0.8 | 3.1 | 3125 | 12500 | 3.0 | 4 | 4 |
| ALCAM | 1:100 | 3.1 | 6.1 | 3125 | 12500 | 2.7 | 4 | 5 |
| ADGRE5 | 1:100 | 3.1 | 6.1 | 25000 | 200000 | 3.6 | 5 | 7 |
| ENG | 1:100 | 1.5 | 3.1 | 3125 | 12500 | 3.0 | 4 | 4 |
| CORO1A | 1:100 | 6.1 | 24.4 | 100000 | 400000 | 3.6 | 6 | 7 |
| COL6A3 | 1:100 | 3.1 | 6.1 | 3125 | 100000 | 2.7 | 4 | 6 |
| SELE | 1:100 | 0.8 | 1.5 | 1563 | 3125 | 3.0 | 5 | 5 |
| CPB1 | 1:100 | 1.5 | 3.1 | 6250 | 25000 | 3.3 | 4 | 5 |
| CST6 | 1:100 | 12.2 | 24.4 | 3125 | 12500 | 2.1 | 5 | 5 |
| CD209 | 1:100 | 1.5 | 3.1 | 12500 | 100000 | 3.6 | 4 | 6 |
| PLA2G1B | 1:100 | 0.4 | 0.8 | 6250 | 12500 | 3.9 | 4 | 6 |
| MEGF9 | 1:100 | 0.8 | 1.5 | 6250 | 25000 | 3.6 | 5 | 6 |
| PAM | 1:100 | 97.7 | 195.3 | 200000 | 800000 | 3.0 | 4 | 6 |
| CCN3 | 1:100 | 0.001 | 1.5 | 6250 | 25000 | 3.6 | 3 | 5 |
| TCN2 | 1:100 | 0.8 | 1.5 | 6250 | 12500 | 3.6 | 5 | 9 |
| LILRB5 | 1:100 | 0.4 | 0.8 | 6250 | 25000 | 3.9 | 4 | 4 |

| | | | | | | | | |
|---|---|---|---|---|---|---|---|---|
| IGFBP2 | 1:100 | 12.2 | 24.4 | 50000 | 200000 | 3.3 | 5 | 7 |
| RNASET2 | 1:100 | 0.4 | 0.8 | 3125 | 12500 | 3.6 | 4 | 4 |
| AMY2B | 1:100 | 12.2 | 24.4 | 6250 | 25000 | 2.4 | 8 | 10 |
| CNTN1 | 1:100 | 3.1 | 6.1 | 12500 | 100000 | 3.3 | 5 | 6 |
| CHIT1 | 1:100 | 0.4 | 0.8 | 6250 | 25000 | 3.9 | 5 | 5 |
| COL18A1 | 1:100 | 3.1 | 6.1 | 3125 | 12500 | 2.7 | 4 | 8 |
| FAS | 1:100 | 0.1 | 0.2 | 6250 | 12500 | 4.5 | 5 | 6 |
| PPIB | 1:100 | 24.4 | 48.8 | 12500 | 200000 | 2.4 | 8 | 10 |
| CCL16 | 1:100 | 3.1 | 3.1 | 6250 | 12500 | 3.3 | 4 | 8 |
| BLMH | 1:100 | 24.4 | 48.8 | 12500 | 100000 | 2.4 | 4 | 5 |
| PTPRS | 1:100 | 3.1 | 6.1 | 6250 | 12500 | 3.0 | 5 | 5 |
| PI3 | 1:100 | 12.2 | 48.8 | 6250 | 12500 | 2.1 | 5 | 5 |
| TIE1 | 1:100 | | | | | | 4 | 6 |
| CELA3A | 1:100 | 1.5 | 6.1 | 6250 | 25000 | 3.0 | 4 | 7 |
| NOTCH1 | 1:100 | 0.2 | 0.4 | 6250 | 12500 | 4.2 | 3 | 4 |
| AMY2A | 1:100 | 24.4 | 24.4 | 6250 | 12500 | 2.4 | 8 | 8 |
| SIRPA | 1:100 | 0.4 | 0.8 | 6250 | 12500 | 3.9 | 5 | 7 |
| PDGFA | 1:100 | 0.4 | 0.8 | 3125 | 12500 | 3.6 | 4 | 6 |
| RETN | 1:100 | 0.2 | 0.4 | 3125 | 12500 | 3.9 | 6 | 6 |
| PLXNB2 | 1:100 | 0.8 | 1.5 | 6250 | 50000 | 3.6 | 4 | 5 |
| PRTN3 | 1:100 | 12.2 | 48.8 | 6250 | 100000 | 2.1 | 5 | 4 |
| ITGB2 | 1:100 | 1.5 | 3.1 | 6250 | 25000 | 3.3 | 6 | 8 |
| CD59 | 1:100 | 0.001 | 6.1 | 1563 | 3125 | 2.4 | 4 | 6 |
| PGLYRP1 | 1:100 | 0.8 | 1.5 | 3125 | 12500 | 3.3 | 5 | 6 |
| FAP | 1:100 | 12.2 | 24.4 | 12500 | 25000 | 2.7 | 5 | 6 |
| SERPINE1 | 1:100 | 0.8 | 1.5 | 6250 | 25000 | 3.6 | 6 | 5 |
| EGFR | 1:100 | 3.1 | 6.1 | 6250 | 12500 | 3.0 | 4 | 6 |
| ACP5 | 1:100 | 0.8 | 1.5 | 6250 | 12500 | 3.6 | 4 | 7 |
| COL1A1 | 1:100 | | | | | | 3 | 4 |
| AZU1 | 1:100 | 6.1 | 12.2 | 6250 | 12500 | 2.7 | 7 | 9 |
| CASP3 | 1:100 | 1.5 | 3.1 | 3125 | 12500 | 3.0 | 5 | 9 |
| SPP1 | 1:100 | 1.5 | 3.1 | 6250 | 12500 | 3.3 | 3 | 8 |
| QPCT | 1:100 | 24.4 | 48.8 | 6250 | 50000 | 2.1 | 5 | 7 |
| GAS6 | 1:100 | 48.8 | 97.7 | 400000 | 800000 | 3.6 | 5 | 7 |
| PRCP | 1:100 | 24.4 | 24.4 | 6250 | 25000 | 2.4 | 5 | 6 |
| BPIFB1 | 1:100 | 97.7 | 390.6 | 200000 | 800000 | 2.7 | 5 | 8 |
| CCL15 | 1:100 | 3.1 | 6.1 | 6250 | 12500 | 3.0 | 8 | 5 |
| LILRB2 | 1:100 | 0.8 | 1.5 | 6250 | 12500 | 3.6 | 4 | 6 |
| PTPRF | 1:100 | 3.1 | 6.1 | 25000 | 200000 | 3.6 | 4 | 7 |
| HYOU1 | 1:100 | 24.4 | 24.4 | 12500 | 400000 | 2.7 | 5 | 6 |
| ST6GAL1 | 1:100 | 24.4 | 48.8 | 400000 | 800000 | 3.9 | 6 | 9 |
| LILRB1 | 1:100 | 0.8 | 1.5 | 3125 | 12500 | 3.3 | 4 | 5 |
| MCAM | 1:1000 | 24.4 | 48.8 | 25000 | 800000 | 2.7 | 8 | 7 |
| F9 | 1:1000 | 48.8 | 97.7 | 50000 | 800000 | 2.7 | 7 | 9 |
| CDH1 | 1:1000 | 6.1 | 6.1 | 12500 | 50000 | 3.3 | 9 | 12 |
| CD46 | 1:1000 | 390.6 | 3125.0 | 6400000 | 12800000 | 3.3 | 11 | 13 |
| NCAM1 | 1:1000 | 6.1 | 12.2 | 25000 | 400000 | 3.3 | 7 | 9 |
| FCGR2A | 1:1000 | 0.2 | 0.4 | 1563 | 6250 | 3.6 | 10 | 10 |
| CTSZ | 1:1000 | 0.8 | 1.5 | 3125 | 6250 | 3.3 | 7 | 9 |
| LBP | 1:1000 | | | | | | 11 | 24 |
| HYAL1 | 1:1000 | 24.4 | 24.4 | 25000 | 100000 | 3.0 | 7 | 6 |
| PRSS2 | 1:1000 | 0.4 | 0.8 | 391 | 12500 | 2.7 | 7 | 6 |

| | | | | | | | | |
|---|---|---|---|---|---|---|---|---|
| GP1BA | 1:1000 | 1.5 | 3.1 | 12500 | 25000 | 3.6 | 7 | 11 |
| HSPG2 | 1:1000 | 0.4 | 1.5 | 6250 | 25000 | 3.6 | 5 | 9 |
| IL6R | 1:1000 | 0.1 | 0.2 | 3125 | 6250 | 4.2 | 7 | 10 |
| DEFA1_DEFA1B | 1:1000 | | | | | | 12 | 22 |
| TNC | 1:1000 | 0.4 | 1.5 | 3125 | 25000 | 3.3 | 9 | 10 |
| SELP | 1:1000 | 0.2 | 0.4 | 3125 | 12500 | 3.9 | 9 | 11 |
| SPARCL1 | 1:1000 | 6.1 | 12.2 | 12500 | 50000 | 3.0 | 9 | 9 |
| CD163 | 1:1000 | 6.1 | 12.2 | 25000 | 100000 | 3.3 | 8 | 7 |
| IL18BP | 1:1000 | 0.8 | 1.5 | 12500 | 25000 | 3.9 | 7 | 10 |
| CR2 | 1:1000 | 0.4 | 0.8 | 12500 | 25000 | 4.2 | 6 | 8 |
| ADA2 | 1:1000 | 0.4 | 0.8 | 6250 | 50000 | 3.9 | 6 | 7 |
| ART3 | 1:1000 | 1.5 | 3.1 | 3125 | 25000 | 3.0 | 7 | 7 |
| FCN2 | 1:1000 | 12.2 | 12.2 | 25000 | 800000 | 3.3 | 8 | 11 |
| LCN2 | 1:1000 | 0.8 | 1.5 | 1563 | 25000 | 3.0 | 8 | 9 |
| AXL | 1:1000 | 0.4 | 0.8 | 6250 | 25000 | 3.9 | 8 | 7 |
| OSMR | 1:1000 | 6.1 | 12.2 | 12500 | 25000 | 3.0 | 7 | 9 |
| PCOLCE | 1:1000 | 24.4 | 48.8 | 12500 | 25000 | 2.4 | 9 | 13 |
| NRP1 | 1:1000 | 6.1 | 12.2 | 12500 | 25000 | 3.0 | 8 | 8 |
| CNDP1 | 1:1000 | 390.6 | 781.3 | 400000 | 800000 | 2.7 | 8 | 15 |
| VASN | 1:1000 | 6.1 | 12.2 | 12500 | 50000 | 3.0 | 9 | 9 |
| VCAM1 | 1:1000 | 1.5 | 3.1 | 12500 | 50000 | 3.6 | 6 | 8 |
| SOD1 | 1:1000 | 24.4 | 97.7 | 6250 | 25000 | 1.8 | 5 | 19 |
| FCGR3B | 1:1000 | 3.1 | 6.1 | 6250 | 25000 | 3.0 | 7 | 7 |
| CTSD | 1:1000 | 48.8 | 195.3 | 25000 | 50000 | 2.1 | | |
| AOC3 | 1:1000 | 6.1 | 12.2 | 6250 | 25000 | 2.7 | 8 | 8 |
| ANPEP | 1:1000 | 24.4 | 48.8 | 100000 | 800000 | 3.3 | 7 | 8 |
| CD55 | 1:1000 | 0.2 | 0.4 | 3125 | 6250 | 3.9 | 8 | 9 |
| CHL1 | 1:1000 | 24.4 | 48.8 | 12500 | 400000 | 2.4 | 7 | 8 |
| TFRC | 1:1000 | 24.4 | 48.8 | 200000 | 800000 | 3.6 | 7 | 6 |
| ITGB1 | 1:1000 | 0.05 | 0.4 | 12500 | 25000 | 4.5 | 11 | 13 |
| TFPI | 1:1000 | 0.8 | 1.5 | 6250 | 50000 | 3.6 | 10 | 13 |
| C1QTNF1 | 1:1000 | 0.8 | 1.5 | 6250 | 50000 | 3.6 | 17 | 19 |
| CD93 | 1:1000 | 0.2 | 0.4 | 6250 | 25000 | 4.2 | 7 | 8 |
| KIT | 1:1000 | 0.4 | 0.8 | 1563 | 6250 | 3.3 | 8 | 10 |
| NID1 | 1:1000 | 3.1 | 6.1 | 50000 | 200000 | 3.9 | 7 | 8 |
| APOM | 1:1000 | 6.1 | 6.1 | 12500 | 50000 | 3.3 | 11 | 12 |
| TIMD4 | 1:1000 | 0.2 | 0.4 | 1563 | 6250 | 3.6 | 8 | 8 |
| CD14 | 1:1000 | 12.2 | 24.4 | 200000 | 400000 | 3.9 | 10 | 14 |
| RARRES2 | 1:1000 | 6.1 | 24.4 | 25000 | 800000 | 3.0 | 8 | 12 |
| SSC5D | 1:1000 | 48.8 | 97.7 | 50000 | 400000 | 2.7 | 9 | 10 |
| TFF3 | 1:1000 | 0.2 | 0.4 | 781 | 3125 | 3.3 | 10 | 10 |
| THBS4 | 1:1000 | 6.1 | 24.4 | 50000 | 200000 | 3.3 | 7 | 9 |
| CCL18 | 1:1000 | 0.4 | 0.8 | 3125 | 6250 | 3.6 | 10 | 23 |
| ANGPTL3 | 1:1000 | 1.5 | 3.1 | 12500 | 100000 | 3.6 | 10 | 9 |
| CCL14 | 1:1000 | 0.1 | 0.4 | 1563 | 6250 | 3.6 | 7 | 11 |
| F7 | 1:1000 | 0.8 | 3.1 | 6250 | 25000 | 3.3 | 6 | 10 |
| PTGDS | 1:1000 | 3.1 | 6.1 | 12500 | 50000 | 3.3 | 7 | 8 |
| C2 | 1:1000 | 48.8 | 97.7 | 200000 | 800000 | 3.3 | 7 | 11 |
| IGFBP6 | 1:1000 | 0.8 | 1.5 | 12500 | 25000 | 3.9 | 6 | 8 |
| TGFBI | 1:1000 | 97.7 | 195.3 | 100000 | 400000 | 2.7 | 8 | 12 |
| CHI3L1 | 1:1000 | 0.4 | 0.8 | 3125 | 6250 | 3.6 | 6 | 10 |
| ITIH3 | 1:1000 | 48.8 | 97.7 | 50000 | 100000 | 2.7 | 7 | 10 |

| | | | | | | | | |
|---|---|---|---|---|---|---|---|---|
| FETUB | 1:1000 | 12.2 | 48.8 | 50000 | 200000 | 3.0 | 7 | 7 |
| COMP | 1:1000 | 1.5 | 3.1 | 12500 | 50000 | 3.6 | 8 | 9 |
| GGH | 1:1000 | 195.3 | 390.6 | 50000 | 400000 | 2.1 | 6 | 7 |
| CA1 | 1:1000 | 0.8 | 1.5 | 25000 | 100000 | 4.2 | 6 | 7 |
| CCL5 | 1:1000 | 0.4 | 1.5 | 1563 | 12500 | 3.0 | 10 | 14 |
| REG1A | 1:1000 | 0.05 | 0.1 | 3125 | 6250 | 4.5 | 6 | 7 |
| EFEMP1 | 1:1000 | 24.4 | 48.8 | 50000 | 100000 | 3.0 | 10 | 14 |
| ANG | 1:1000 | 0.8 | 1.5 | 781 | 3125 | 2.7 | 7 | 11 |
| DPP4 | 1:1000 | 12.2 | 48.8 | 12500 | 100000 | 2.4 | 6 | 8 |
| PROC | 1:1000 | 97.7 | 195.3 | 100000 | 400000 | 2.7 | 7 | 8 |
| ICAM1 | 1:1000 | 0.4 | 0.8 | 6250 | 25000 | 3.9 | 8 | 8 |
| CST3 | 1:1000 | 12.2 | 48.8 | 6250 | 12500 | 2.1 | 6 | 13 |
| IGFBP3 | 1:1000 | 12.2 | 24.4 | 12500 | 25000 | 2.7 | 8 | 9 |
| TIMP1 | 1:1000 | 0.8 | 1.5 | 3125 | 12500 | 3.3 | 9 | 10 |
| IDO1 | 1:1 | 390.6 | 1562.5 | 200000 | 400000 | 2.1 | 10 | 13 |
| SCRIB | 1:1 | 195.3 | 390.6 | 50000 | 200000 | 2.1 | 8 | 22 |
| MYL4 | 1:1 | 3125.0 | 3125.0 | 100000 | 800000 | 1.5 | 11 | 20 |
| PECR | 1:1 | | | | | | 6 | 7 |
| EHD3 | 1:1 | 6250.0 | 12500.0 | 800000 | 800000 | 1.8 | 6 | 27 |
| ENTR1 | 1:1 | 781.3 | 781.3 | 400000 | 800000 | 2.7 | 9 | 15 |
| CEP170 | 1:1 | 48.8 | 48.8 | 3125 | 6250 | 1.8 | 6 | 11 |
| HSBP1 | 1:1 | 12.2 | 12.2 | 3125 | 25000 | 2.4 | 6 | 29 |
| SKIV2L | 1:1 | | | | | | 7 | 25 |
| CSF2 | 1:1 | 12.2 | 24.4 | 12500 | 50000 | 2.7 | 16 | 18 |
| RYR1 | 1:1 | 390.6 | 781.3 | 100000 | 200000 | 2.1 | 13 | 12 |
| MEGF11 | 1:1 | 97.7 | 97.7 | 25000 | 200000 | 2.4 | 10 | 10 |
| HIP1 | 1:1 | 1562.5 | 3125.0 | 200000 | 400000 | 1.8 | 8 | |
| RTKN2 | 1:1 | 3125.0 | 6250.0 | 400000 | 800000 | 1.8 | 5 | |
| UGDH | 1:1 | | | | | | 9 | 24 |
| CASQ2 | 1:1 | 6250.0 | 6250.0 | 400000 | 1600000 | 1.8 | 12 | 21 |
| HCG22 | 1:1 | 24.4 | 48.8 | 12500 | 50000 | 2.4 | 6 | 16 |
| NECAP2 | 1:1 | 12500.0 | 12500.0 | 400000 | 800000 | 1.5 | 15 | 25 |
| ATP2B4 | 1:1 | 390.6 | 390.6 | 25000 | 400000 | 1.8 | | |
| RBM19 | 1:1 | 390.6 | 390.6 | 100000 | 400000 | 2.4 | 14 | 18 |
| MYH4 | 1:1 | 390.6 | 390.6 | 25000 | 200000 | 1.8 | 14 | |
| ATP1B3 | 1:1 | 781.3 | 781.3 | 100000 | 400000 | 2.1 | | |
| CYP24A1 | 1:1 | 1562.5 | 3125.0 | 200000 | 800000 | 1.8 | 15 | |
| GCLM | 1:1 | 1562.5 | 3125.0 | 200000 | 800000 | 1.8 | 11 | 17 |
| TPM3 | 1:1 | 1562.5 | 1562.5 | 200000 | 400000 | 2.1 | 11 | 12 |
| KRT17 | 1:1 | 390.6 | 781.3 | 200000 | 800000 | 2.4 | | |
| LAMA1 | 1:1 | | | | | | | |
| BRD3 | 1:1 | | | | | | 12 | 32 |
| NOP56 | 1:1 | 3125.0 | 3125.0 | 200000 | 400000 | 1.8 | 13 | 37 |
| EIF2AK3 | 1:1 | 3125.0 | 6250.0 | 400000 | 800000 | 1.8 | 17 | 15 |
| NOS2 | 1:1 | 390.6 | 781.3 | 400000 | 800000 | 2.7 | 15 | 17 |
| MED21 | 1:1 | 781.3 | 1562.5 | 100000 | 400000 | 1.8 | 9 | 17 |
| ADRA2A | 1:1 | 3125.0 | 6250.0 | 200000 | 400000 | 1.5 | 9 | 15 |
| ACSL1 | 1:1 | 3125.0 | 6250.0 | 200000 | 400000 | 1.5 | | |
| ITPA | 1:1 | 781.3 | 1562.5 | 50000 | 400000 | 1.5 | 8 | 21 |
| ATP5F1D | 1:1 | 390.6 | 390.6 | 50000 | 200000 | 2.1 | 10 | 20 |
| FDX1 | 1:1 | | | | | | 9 | |
| SLC12A2 | 1:1 | | | | | | 8 | |

| | | | | | | | | |
|---|---|---|---|---|---|---|---|---|
| EXOSC10 | 1:1 | 3125.0 | 6250.0 | 400000 | 800000 | 1.8 | 12 | 3 |
| NXPE4 | 1:1 | 1562.5 | 1562.5 | 200000 | 400000 | 2.1 | 7 | 14 |
| REXO2 | 1:1 | 97.7 | 195.3 | 12500 | 400000 | 1.8 | 14 | |
| MAP1LC3B2 | 1:1 | 3125.0 | 3125.0 | 200000 | 800000 | 1.8 | 8 | 15 |
| PDE4D | 1:1 | 25000.0 | 50000.0 | 3200000 | 12800000 | 1.8 | 10 | 50 |
| RPS10 | 1:1 | | | | | | | |
| RAB39B | 1:1 | 3125.0 | 3125.0 | 200000 | 800000 | 1.8 | 13 | 16 |
| TMED10 | 1:1 | 390.6 | 781.3 | 100000 | 200000 | 2.1 | 7 | 12 |
| ACY3 | 1:1 | | | | | | 10 | 9 |
| ERVV-1 | 1:1 | 195.3 | 781.3 | 100000 | 200000 | 2.1 | 15 | 33 |
| TRAF3IP2 | 1:1 | 195.3 | 195.3 | 100000 | 200000 | 2.7 | 10 | 11 |
| CACNA1H | 1:1 | 390.6 | 781.3 | 100000 | 400000 | 2.1 | 12 | 20 |
| BHMT2 | 1:1 | 50000.0 | 100000.0 | 6400000 | 12800000 | 1.8 | | |
| CD2 | 1:1 | 781.3 | 1562.5 | 200000 | 400000 | 2.1 | 10 | 14 |
| SHD | 1:1 | 6250.0 | 6250.0 | 400000 | 800000 | 1.8 | 12 | 8 |
| SNU13 | 1:1 | | | | | | 8 | 12 |
| THRAP3 | 1:1 | 3125.0 | 3125.0 | 200000 | 800000 | 1.8 | 9 | 15 |
| TSPAN15 | 1:1 | 390.6 | 781.3 | 100000 | 200000 | 2.1 | 15 | 14 |
| UBQLN3 | 1:1 | 390.6 | 390.6 | 100000 | 200000 | 2.4 | | |
| DENND2B | 1:1 | 3125.0 | 3125.0 | 200000 | 400000 | 1.8 | 11 | 6 |
| GAMT | 1:1 | | | | | | 12 | |
| ELOB | 1:1 | 1562.5 | 1562.5 | 100000 | 400000 | 1.8 | 10 | 32 |
| NEB | 1:1 | 781.3 | 1562.5 | 200000 | 400000 | 2.1 | 13 | 8 |
| DDA1 | 1:1 | 1171.9 | 1171.9 | 300000 | 600000 | 2.4 | 11 | 11 |
| CPTP | 1:1 | 6250.0 | 12500.0 | 400000 | 800000 | 1.5 | 7 | 17 |
| PMM2 | 1:1 | | | | | | 11 | 39 |
| ATP1B1 | 1:1 | 195.3 | 195.3 | 25000 | 200000 | 2.1 | 11 | 11 |
| SLC51B | 1:1 | 781.3 | 781.3 | 100000 | 400000 | 2.1 | 10 | 22 |
| IGHMBP2 | 1:1 | 390.6 | 781.3 | 50000 | 200000 | 1.8 | | |
| RAB33A | 1:1 | 6250.0 | 12500.0 | 800000 | 800000 | 1.8 | 12 | 21 |
| ARL2BP | 1:1 | 1562.5 | 3125.0 | 200000 | 400000 | 1.8 | 9 | 17 |
| TTN | 1:1 | 195.3 | 390.6 | 12500 | 200000 | 1.5 | 10 | 12 |
| TPR | 1:1 | 97.7 | 97.7 | 12500 | 100000 | 2.1 | 14 | 16 |
| ECI2 | 1:1 | 6250.0 | 12500.0 | 400000 | 800000 | 1.5 | 13 | 35 |
| MYL1 | 1:1 | 6250.0 | 6250.0 | 200000 | 400000 | 1.5 | 6 | 14 |
| ATP1B2 | 1:1 | 781.3 | 1562.5 | 100000 | 400000 | 1.8 | | |
| GASK1A | 1:1 | | | | | | 12 | 15 |
| ABCA2 | 1:1 | 781.3 | 1562.5 | 200000 | 400000 | 2.1 | 19 | 26 |
| ZCCHC8 | 1:1 | 48.8 | 97.7 | 50000 | 100000 | 2.7 | 18 | 30 |
| CALY | 1:1 | 3125.0 | 3125.0 | 200000 | 800000 | 1.8 | 6 | 15 |
| NPR1 | 1:1 | 3125.0 | 6250.0 | 200000 | 800000 | 1.5 | 8 | 9 |
| MTR | 1:1 | 781.3 | 781.3 | 50000 | 400000 | 1.8 | 20 | 10 |
| CEP112 | 1:1 | 195.3 | 390.6 | 100000 | 200000 | 2.4 | 8 | 10 |
| HSD17B3 | 1:1 | 3125.0 | 3125.0 | 200000 | 800000 | 1.8 | 12 | 25 |
| EXTL1 | 1:1 | 390.6 | 390.6 | 100000 | 200000 | 2.4 | 10 | 14 |
| SAT1 | 1:1 | 195.3 | 195.3 | 12500 | 100000 | 1.8 | 2 | |
| SART1 | 1:1 | 390.6 | 1562.5 | 100000 | 400000 | 1.8 | 11 | 15 |
| USP47 | 1:1 | 390.6 | 781.3 | 100000 | 400000 | 2.1 | | |
| SCGB2A2 | 1:1 | 1562.5 | 3125.0 | 200000 | 400000 | 1.8 | 23 | |
| FDX2 | 1:1 | 195.3 | 195.3 | 12500 | 200000 | 1.8 | 13 | 18 |
| IFNW1 | 1:1 | 195.3 | 390.6 | 200000 | 400000 | 2.7 | 4 | 1 |
| RIPK4 | 1:1 | 1562.5 | 1562.5 | 200000 | 800000 | 2.1 | 15 | 31 |

| | | | | | | | | |
|---|---|---|---|---|---|---|---|---|
| NUP50 | 1:1 | | | | | | 11 | |
| DHODH | 1:1 | 12500.0 | 12500.0 | 800000 | 800000 | 1.8 | 10 | |
| GBP6 | 1:1 | | | | | | | |
| EDEM2 | 1:1 | 1562.5 | 3125.0 | 200000 | 800000 | 1.8 | 13 | 6 |
| ENPEP | 1:1 | 390.6 | 781.3 | 100000 | 400000 | 2.1 | 13 | 21 |
| NFYA | 1:1 | | | | | | 11 | 10 |
| PGLYRP4 | 1:1 | 390.6 | 390.6 | 50000 | 100000 | 2.1 | 8 | 25 |
| TRPV3 | 1:1 | 24.4 | 24.4 | 12500 | 50000 | 2.7 | 14 | 23 |
| NUDT15 | 1:1 | 390.6 | 390.6 | 25000 | 200000 | 1.8 | 14 | 19 |
| GUK1 | 1:1 | | | | | | 13 | 3 |
| HADH | 1:1 | | | | | | | |
| MYOM2 | 1:1 | 97.7 | 195.3 | 12500 | 100000 | 1.8 | 18 | 17 |
| PRKD2 | 1:1 | 6250.0 | 12500.0 | 800000 | 3200000 | 1.8 | 9 | 10 |
| TEF | 1:1 | 195.3 | 781.3 | 200000 | 800000 | 2.4 | 14 | 11 |
| MANSC4 | 1:1 | 390.6 | 390.6 | 25000 | 200000 | 1.8 | 10 | 33 |
| ABRAXAS2 | 1:1 | | | | | | 9 | 18 |
| IGF2BP3 | 1:1 | 12500.0 | 12500.0 | 200000 | 800000 | 1.2 | | |
| PROCR | 1:1 | | | | | | 8 | 26 |
| KIF22 | 1:1 | | | | | | 15 | 37 |
| COPB2 | 1:1 | 195.3 | 195.3 | 6250 | 400000 | 1.5 | 16 | 32 |
| PMCH | 1:1 | 3125.0 | 3125.0 | 100000 | 800000 | 1.5 | 8 | 12 |
| UHRF2 | 1:1 | 781.3 | 1562.5 | 100000 | 200000 | 1.8 | | |
| ASS1 | 1:1 | 781.3 | 3125.0 | 200000 | 400000 | 1.8 | 9 | 18 |
| KRT6C | 1:1 | 390.6 | 390.6 | 25000 | 200000 | 1.8 | | |
| ARNTL | 1:1 | 3125.0 | 6250.0 | 400000 | 800000 | 1.8 | | |
| ATP6V1G2 | 1:1 | | | | | | 22 | 17 |
| CSRP3 | 1:1 | 1562.5 | 3125.0 | 50000 | 200000 | 1.2 | 24 | 22 |
| ARL13B | 1:1 | 195.3 | 195.3 | 25000 | 400000 | 2.1 | 13 | 24 |
| GSTM4 | 1:1 | 3125.0 | 3125.0 | 200000 | 800000 | 1.8 | 12 | 12 |
| KIF1C | 1:1 | 97.7 | 97.7 | 25000 | 50000 | 2.4 | 9 | 10 |
| LILRA4 | 1:1 | | | | | | 12 | |
| ANK2 | 1:1 | 195.3 | 390.6 | 50000 | 100000 | 2.1 | 11 | 11 |
| NFKB1 | 1:1 | | | | | | 13 | 19 |
| MMUT | 1:1 | | | | | | 12 | 34 |
| NFX1 | 1:1 | 1562.5 | 3125.0 | 100000 | 400000 | 1.5 | 14 | 22 |
| HMGCS1 | 1:1 | 6250.0 | 6250.0 | 200000 | 400000 | 1.5 | 8 | 6 |
| B3GAT3 | 1:1 | | | | | | | |
| CBX2 | 1:1 | 3125.0 | 3125.0 | 400000 | 800000 | 2.1 | 10 | |
| MRPL24 | 1:1 | 3125.0 | 6250.0 | 400000 | 800000 | 1.8 | 11 | 16 |
| BRDT | 1:1 | 48.8 | 97.7 | 12500 | 50000 | 2.1 | 13 | 20 |
| NUDT10 | 1:1 | 781.3 | 781.3 | 50000 | 200000 | 1.8 | | |
| GNPDA1 | 1:1 | 195.3 | 195.3 | 12500 | 100000 | 1.8 | 9 | 12 |
| SEL1L | 1:1 | 390.6 | 781.3 | 100000 | 400000 | 2.1 | 7 | 8 |
| TOR1AIP1 | 1:1 | 6250.0 | 12500.0 | 800000 | 3200000 | 1.8 | 10 | 26 |
| PNMA1 | 1:1 | 6250.0 | 12500.0 | 800000 | 800000 | 1.8 | 9 | 24 |
| VIPR1 | 1:1 | 195.3 | 390.6 | 100000 | 800000 | 2.4 | | |
| ACTN2 | 1:1 | 781.3 | 1562.5 | 100000 | 400000 | 1.8 | 18 | 18 |
| LRCH4 | 1:1 | | | | | | 17 | 17 |
| LPP | 1:1 | | | | | | 14 | 15 |
| VSIG10L | 1:1 | 6250.0 | 6250.0 | 200000 | 800000 | 1.5 | 10 | |
| NAA10 | 1:1 | | | | | | 20 | 35 |
| FHIP2A | 1:1 | | | | | | 8 | 7 |

| | | | | | | | | |
|---|---|---|---|---|---|---|---|---|
| PACS2 | 1:1 | 97.7 | 195.3 | 25000 | 200000 | 2.1 | 13 | 25 |
| TSNAX | 1:1 | | | | | | 11 | 20 |
| PKD2 | 1:1 | 781.3 | 1562.5 | 100000 | 200000 | 1.8 | 12 | 17 |
| CSDE1 | 1:1 | | | | | | 16 | 20 |
| PALM3 | 1:1 | | | | | | 12 | 19 |
| YOD1 | 1:1 | | | | | | 14 | 21 |
| C1GALT1C1 | 1:1 | | | | | | 16 | 26 |
| EIF5 | 1:1 | 781.3 | 1562.5 | 100000 | 400000 | 1.8 | 16 | 27 |
| FRMD4B | 1:1 | | | | | | 11 | 19 |
| GP1BB | 1:1 | | | | | | 15 | 28 |
| SPINK8 | 1:1 | 781.3 | 1562.5 | 200000 | 400000 | 2.1 | 10 | |
| PDZD2 | 1:1 | 390.6 | 390.6 | 200000 | 400000 | 2.7 | 14 | 20 |
| PYY | 1:1 | | | | | | 14 | 18 |
| TMED4 | 1:1 | 1562.5 | 1562.5 | 100000 | 400000 | 1.8 | 11 | |
| ENOX2 | 1:1 | 1562.5 | 3125.0 | 800000 | 3200000 | 2.4 | 8 | 27 |
| LMOD1 | 1:1 | 195.3 | 390.6 | 200000 | 800000 | 2.7 | 7 | 24 |
| MYBPC2 | 1:1 | 781.3 | 1562.5 | 100000 | 800000 | 1.8 | 7 | 21 |
| IGSF21 | 1:1 | 1562.5 | 1562.5 | 100000 | 400000 | 1.8 | 7 | 15 |
| ENO3 | 1:1 | | | | | | | |
| M6PR | 1:1 | 97.7 | 97.7 | 6250 | 25000 | 1.8 | 9 | 12 |
| HRC | 1:1 | 781.3 | 1562.5 | 200000 | 3200000 | 2.1 | 8 | 16 |
| SIL1 | 1:1 | 390.6 | 781.3 | 50000 | 200000 | 1.8 | 6 | 9 |
| PAGR1 | 1:1 | 195.3 | 195.3 | 50000 | 400000 | 2.4 | 15 | 17 |
| FSHB | 1:1 | 97.7 | 195.3 | 12500 | 100000 | 1.8 | 6 | 9 |
| SNED1 | 1:1 | 1562.5 | 3125.0 | 400000 | 800000 | 2.1 | 12 | 21 |
| APOBR | 1:1 | | | | | | 7 | 12 |
| RECK | 1:1 | 1562.5 | 1562.5 | 50000 | 200000 | 1.5 | 7 | 5 |
| DMP1 | 1:1 | 3125.0 | 6250.0 | 800000 | 6400000 | 2.1 | 8 | 35 |
| KHK | 1:1 | 195.3 | 195.3 | 12500 | 100000 | 1.8 | 6 | 20 |
| BECN1 | 1:1 | 12500.0 | 25000.0 | 800000 | 800000 | 1.5 | 10 | 14 |
| RNF5 | 1:1 | | | | | | 12 | 21 |
| CCER2 | 1:1 | 1562.5 | 1562.5 | 100000 | 400000 | 1.8 | 9 | 12 |
| COQ7 | 1:1 | 390.6 | 781.3 | 100000 | 400000 | 2.1 | 12 | 18 |
| HS1BP3 | 1:1 | 3125.0 | 3125.0 | 100000 | 800000 | 1.5 | 8 | 25 |
| COMMD1 | 1:1 | 24.4 | 48.8 | 12500 | 50000 | 2.4 | 8 | 16 |
| OXCT1 | 1:1 | 781.3 | 3125.0 | 200000 | 800000 | 1.8 | 10 | 21 |
| ATP6V1G1 | 1:1 | 3125.0 | 3125.0 | 200000 | 800000 | 1.8 | 12 | 35 |
| DMD | 1:1 | 390.6 | 781.3 | 200000 | 800000 | 2.4 | 9 | 17 |
| SNX5 | 1:1 | | | | | | 5 | 9 |
| EDN1 | 1:1 | | | | | | 6 | 15 |
| ROBO4 | 1:1 | | | | | | 6 | 9 |
| AHNAK2 | 1:1 | 97.7 | 195.3 | 12500 | 50000 | 1.8 | 6 | 9 |
| HMGCL | 1:1 | 781.3 | 1562.5 | 100000 | 400000 | 1.8 | 8 | 25 |
| ITPR1 | 1:1 | 781.3 | 1562.5 | 100000 | 400000 | 1.8 | 13 | 14 |
| AIF1L | 1:1 | 390.6 | 781.3 | 200000 | 800000 | 2.4 | 7 | 19 |
| LONP1 | 1:1 | 781.3 | 781.3 | 50000 | 200000 | 1.8 | 10 | 28 |
| AAMDC | 1:1 | | | | | | 8 | 13 |
| OPLAH | 1:1 | | | | | | 7 | 24 |
| PYDC1 | 1:1 | 6.1 | 12.2 | 1563 | 12500 | 2.1 | 7 | 11 |
| DNAJC6 | 1:1 | | | | | | 6 | 30 |
| RNF149 | 1:1 | 97.7 | 97.7 | 6250 | 50000 | 1.8 | 9 | 11 |
| GADD45GIP1 | 1:1 | | | | | | 10 | 25 |

| | | | | | | | | |
|---|---|---|---|---|---|---|---|---|
| RNASE10 | 1:1 | 195.3 | 195.3 | 12500 | 100000 | 1.8 | 7 | 13 |
| CBS | 1:1 | 3125.0 | 3125.0 | 800000 | 800000 | 2.4 | 9 | 25 |
| ECHS1 | 1:1 | | | | | | 8 | 27 |
| FAM172A | 1:1 | 390.6 | 781.3 | 100000 | 400000 | 2.1 | 8 | 20 |
| ANP32C | 1:1 | 97.7 | 195.3 | 50000 | 400000 | 2.4 | 1 | |
| CALCOCO2 | 1:1 | 195.3 | 195.3 | 50000 | 200000 | 2.4 | 8 | 15 |
| SEC31A | 1:1 | | | | | | 7 | 31 |
| NFE2 | 1:1 | | | | | | 8 | 12 |
| BOLA2 | 1:1 | 195.3 | 195.3 | 25000 | 200000 | 2.1 | 7 | 11 |
| LIPF | 1:1 | 195.3 | 390.6 | 50000 | 200000 | 2.1 | 7 | 22 |
| ZP3 | 1:1 | 48.8 | 97.7 | 25000 | 50000 | 2.4 | | |
| GRP | 1:1 | | | | | | 6 | 22 |
| EIF2S2 | 1:1 | | | | | | 11 | 25 |
| MAN1A2 | 1:1 | 781.3 | 781.3 | 100000 | 800000 | 2.1 | 5 | 8 |
| SIGLEC8 | 1:1 | 12.2 | 12.2 | 6250 | 25000 | 2.7 | 6 | 11 |
| AHNAK | 1:1 | 97.7 | 195.3 | 50000 | 200000 | 2.4 | 7 | 14 |
| INPP5D | 1:1 | 3125.0 | 3125.0 | 200000 | 800000 | 1.8 | 8 | 12 |
| RANBP1 | 1:1 | | | | | | 7 | 14 |
| RBPMS2 | 1:1 | 48.8 | 97.7 | 6250 | 12500 | 1.8 | 6 | 30 |
| ANXA2 | 1:1 | 6250.0 | 12500.0 | 800000 | 800000 | 1.8 | 9 | 14 |
| HSDL2 | 1:1 | | | | | | 8 | 12 |
| NAGK | 1:1 | 195.3 | 390.6 | 12500 | 100000 | 1.5 | 7 | 22 |
| CRYZL1 | 1:1 | 48.8 | 195.3 | 25000 | 50000 | 2.1 | 7 | 28 |
| TMEM132A | 1:1 | | | | | | | |
| COL2A1 | 1:1 | 97.7 | 97.7 | 12500 | 50000 | 2.1 | 6 | 15 |
| CNP | 1:1 | 781.3 | 781.3 | 100000 | 800000 | 2.1 | 6 | 14 |
| RPL14 | 1:1 | 50000.0 | 100000.0 | 6400000 | 12800000 | 1.8 | 14 | 18 |
| ASRGL1 | 1:1 | | | | | | 16 | 31 |
| POMC | 1:1 | | | | | | | |
| B3GNT7 | 1:1 | 781.3 | 781.3 | 200000 | 400000 | 2.4 | 7 | 11 |
| BPIFA2 | 1:1 | 1562.5 | 3125.0 | 100000 | 400000 | 1.5 | 7 | 32 |
| SCPEP1 | 1:1 | 195.3 | 390.6 | 100000 | 400000 | 2.4 | 7 | 9 |
| SCN4B | 1:1 | | | | | | 9 | 26 |
| PRRT3 | 1:1 | 781.3 | 1562.5 | 200000 | 800000 | 2.1 | 6 | 11 |
| ADAMTSL4 | 1:1 | | | | | | 8 | 13 |
| MECR | 1:1 | 6250.0 | 6250.0 | 400000 | 800000 | 1.8 | | |
| ENPP6 | 1:1 | 12.2 | 24.4 | 6250 | 25000 | 2.4 | 8 | 22 |
| LAMB1 | 1:1 | 195.3 | 390.6 | 50000 | 400000 | 2.1 | 6 | 12 |
| DLL4 | 1:1 | | | | | | 17 | 5 |
| GIP | 1:1 | | | | | | 13 | 12 |
| EPPK1 | 1:1 | 390.6 | 781.3 | 100000 | 400000 | 2.1 | 7 | 18 |
| FSTL1 | 1:1 | | | | | | 5 | 9 |
| GATD3 | 1:1 | | | | | | | |
| MYL3 | 1:1 | 390.6 | 781.3 | 50000 | 800000 | 1.8 | 7 | 21 |
| NIT1 | 1:1 | 6250.0 | 6250.0 | 400000 | 800000 | 1.8 | 6 | 13 |
| ECHDC3 | 1:10 | 24.4 | 48.8 | 25000 | 100000 | 2.7 | 4 | 14 |
| NT5C | 1:10 | 12500.0 | 12500.0 | 800000 | 800000 | 1.8 | 5 | 21 |
| GALNT5 | 1:10 | 6250.0 | 6250.0 | 400000 | 800000 | 1.8 | 10 | 10 |
| MYBPC1 | 1:10 | 781.3 | 1562.5 | 200000 | 800000 | 2.1 | 7 | 24 |
| PTGR1 | 1:10 | 390.6 | 390.6 | 50000 | 200000 | 2.1 | 5 | 17 |
| GRHPR | 1:10 | 1562.5 | 1562.5 | 200000 | 800000 | 2.1 | 9 | 18 |
| GGACT | 1:10 | 48.8 | 48.8 | 1563 | 12500 | 1.5 | 12 | 28 |

| | | | | | | | | |
|---|---|---|---|---|---|---|---|---|
| BNIP2 | 1:10 | 3125.0 | 3125.0 | 100000 | 800000 | 1.5 | 11 | 13 |
| CD80 | 1:10 | | | | | | 17 | 18 |
| SPESP1 | 1:10 | 1562.5 | 3125.0 | 200000 | 800000 | 1.8 | 8 | 24 |
| MMP15 | 1:10 | | | | | | 5 | 2 |
| SARG | 1:10 | 97.7 | 195.3 | 12500 | 100000 | 1.8 | | |
| UBE2L6 | 1:10 | 1562.5 | 1562.5 | 50000 | 200000 | 1.5 | 7 | 11 |
| FCAMR | 1:10 | 6250.0 | 6250.0 | 400000 | 800000 | 1.8 | 6 | 22 |
| DTYMK | 1:10 | 3125.0 | 6250.0 | 400000 | 800000 | 1.8 | 5 | 21 |
| SCRG1 | 1:10 | 3125.0 | 3125.0 | 100000 | 400000 | 1.5 | 10 | 12 |
| VIT | 1:10 | 195.3 | 390.6 | 25000 | 100000 | 1.8 | 6 | 16 |
| EEF1D | 1:10 | 3125.0 | 3125.0 | 400000 | 800000 | 2.1 | 4 | 12 |
| HBZ | 1:10 | | | | | | 7 | 31 |
| NIT2 | 1:10 | | | | | | 10 | 32 |
| HEPH | 1:10 | 97.7 | 97.7 | 12500 | 50000 | 2.1 | 5 | 7 |
| MAN2B2 | 1:10 | 390.6 | 781.3 | 50000 | 800000 | 1.8 | 4 | 12 |
| TPK1 | 1:10 | 24.4 | 48.8 | 12500 | 100000 | 2.4 | 5 | 8 |
| COCH | 1:10 | 195.3 | 195.3 | 25000 | 200000 | 2.1 | 5 | 9 |
| SYTL4 | 1:10 | | | | | | 13 | 26 |
| GMFG | 1:10 | 781.3 | 781.3 | 50000 | 200000 | 1.8 | 6 | 23 |
| MAMDC2 | 1:10 | 195.3 | 195.3 | 50000 | 200000 | 2.4 | 5 | 10 |
| SRPX | 1:10 | 781.3 | 781.3 | 50000 | 200000 | 1.8 | 6 | 16 |
| IGSF9 | 1:10 | 97.7 | 97.7 | 25000 | 100000 | 2.4 | 8 | 19 |
| BPIFB2 | 1:10 | 1562.5 | 3125.0 | 100000 | 200000 | 1.5 | | |
| CRYBB1 | 1:10 | 12.2 | 12.2 | 6250 | 25000 | 2.7 | 3 | 16 |
| TIMP2 | 1:10 | | | | | | 11 | 10 |
| CPXM2 | 1:10 | 12500.0 | 12500.0 | 800000 | 800000 | 1.8 | 8 | 18 |
| PDAP1 | 1:10 | 3125.0 | 6250.0 | 200000 | 800000 | 1.5 | | |
| COX6B1 | 1:10 | | | | | | 10 | 20 |
| PTPRB | 1:10 | 781.3 | 3125.0 | 100000 | 200000 | 1.5 | 6 | 10 |
| TCOF1 | 1:10 | 390.6 | 781.3 | 50000 | 200000 | 1.8 | 7 | 9 |
| GGCT | 1:10 | 195.3 | 390.6 | 25000 | 200000 | 1.8 | 9 | 29 |
| RAB11FIP3 | 1:10 | 48.8 | 97.7 | 25000 | 200000 | 2.4 | 6 | 27 |
| TCTN3 | 1:10 | 1562.5 | 1562.5 | 100000 | 800000 | 1.8 | 6 | 12 |
| L3HYPDH | 1:10 | 781.3 | 781.3 | 50000 | 200000 | 1.8 | 9 | 30 |
| CHMP6 | 1:10 | 781.3 | 781.3 | 100000 | 800000 | 2.1 | 8 | 22 |
| MYDGF | 1:10 | 97.7 | 195.3 | 12500 | 200000 | 1.8 | 5 | 29 |
| PKD1 | 1:10 | 24.4 | 97.7 | 6250 | 25000 | 1.8 | 6 | 10 |
| NPL | 1:10 | 781.3 | 1562.5 | 100000 | 400000 | 1.8 | 5 | 18 |
| BCAT1 | 1:10 | 1562.5 | 1562.5 | 100000 | 800000 | 1.8 | 8 | 10 |
| ASPN | 1:10 | | | | | | 8 | 26 |
| SGSH | 1:10 | 97.7 | 97.7 | 12500 | 50000 | 2.1 | 4 | 6 |
| LECT2 | 1:10 | 6250.0 | 6250.0 | 800000 | 800000 | 2.1 | 9 | 35 |
| PTPRZ1 | 1:10 | 781.3 | 781.3 | 100000 | 200000 | 2.1 | 7 | 10 |
| CDA | 1:10 | 1562.5 | 3125.0 | 100000 | 800000 | 1.5 | 9 | 13 |
| NGFR | 1:10 | 390.6 | 390.6 | 25000 | 100000 | 1.8 | 14 | 22 |
| LILRA6 | 1:10 | 97.7 | 195.3 | 25000 | 200000 | 2.1 | 6 | 21 |
| EHBP1 | 1:10 | 195.3 | 390.6 | 25000 | 100000 | 1.8 | 8 | 20 |
| MELTF | 1:10 | 97.7 | 195.3 | 50000 | 200000 | 2.4 | 5 | 8 |
| PGD | 1:10 | 6250.0 | 6250.0 | 200000 | 800000 | 1.5 | 8 | 19 |
| GET3 | 1:10 | 1562.5 | 3125.0 | 400000 | 800000 | 2.1 | 8 | 26 |
| ELN | 1:10 | | | | | | 11 | 16 |
| RILPL2 | 1:10 | 195.3 | 390.6 | 50000 | 200000 | 2.1 | 6 | 35 |

| | | | | | | | | |
|---|---|---|---|---|---|---|---|---|
| FAM20A | 1:10 | 390.6 | 390.6 | 100000 | 800000 | 2.4 | 5 | 14 |
| ADGRF5 | 1:10 | 1562.5 | 3125.0 | 200000 | 800000 | 1.8 | 6 | 8 |
| LRRC59 | 1:10 | | | | | | 12 | 29 |
| CMC1 | 1:10 | | | | | | 6 | 29 |
| SERPINE2 | 1:10 | | | | | | 5 | 17 |
| TIMM10 | 1:10 | 781.3 | 781.3 | 50000 | 200000 | 1.8 | 10 | 10 |
| RAB10 | 1:10 | | | | | | 13 | 16 |
| ACADSB | 1:10 | 3125.0 | 3125.0 | 400000 | 800000 | 2.1 | 12 | 24 |
| ADAMTSL5 | 1:10 | 390.6 | 390.6 | 50000 | 100000 | 2.1 | 11 | 19 |
| FABP3 | 1:10 | | | | | | 8 | 16 |
| TOMM20 | 1:10 | 97.7 | 195.3 | 25000 | 100000 | 2.1 | 12 | 29 |
| UPB1 | 1:10 | 781.3 | 781.3 | 50000 | 200000 | 1.8 | 14 | 13 |
| PPM1F | 1:10 | 6250.0 | 6250.0 | 800000 | 800000 | 2.1 | 10 | 26 |
| RAB27B | 1:10 | 195.3 | 390.6 | 100000 | 400000 | 2.4 | | |
| PDIA4 | 1:100 | 390.6 | 390.6 | 50000 | 100000 | 2.1 | 8 | 27 |
| BDNF | 1:100 | 48.8 | 195.3 | 50000 | 100000 | 2.4 | 9 | 28 |
| DDT | 1:100 | | | | | | 10 | 19 |
| ADAMTSL2 | 1:100 | 3125.0 | 3125.0 | 400000 | 800000 | 2.1 | 9 | 12 |
| SLC4A1 | 1:100 | | | | | | | |
| LCP1 | 1:100 | 1562.5 | 1562.5 | 400000 | 800000 | 2.4 | 10 | 19 |
| AMDHD2 | 1:100 | 3125.0 | 25000.0 | 800000 | 800000 | 1.5 | 8 | 19 |
| ISM2 | 1:100 | 195.3 | 195.3 | 25000 | 200000 | 2.1 | 11 | 22 |
| KLK3 | 1:100 | 195.3 | 195.3 | 25000 | 200000 | 2.1 | 5 | 5 |
| CTLA4 | 1:100 | 97.7 | 97.7 | 25000 | 100000 | 2.4 | 1 | |
| CEACAM6 | 1:100 | 3125.0 | 3125.0 | 400000 | 800000 | 2.1 | 10 | 15 |
| LILRA3 | 1:100 | | | | | | 5 | 13 |
| MUC2 | 1:100 | 6.1 | 12.2 | 6250 | 25000 | 2.7 | 7 | 30 |
| FBLN2 | 1:100 | 195.3 | 195.3 | 25000 | 100000 | 2.1 | 4 | 9 |
| COL3A1 | 1:100 | 97.7 | 195.3 | 25000 | 100000 | 2.1 | 6 | 10 |
| SBSN | 1:100 | | | | | | 7 | 19 |
| CSPG4 | 1:100 | 97.7 | 97.7 | 25000 | 100000 | 2.4 | 5 | 6 |
| COL15A1 | 1:100 | 195.3 | 195.3 | 12500 | 50000 | 1.8 | 5 | 10 |
| FGFBP2 | 1:100 | 195.3 | 195.3 | 25000 | 50000 | 2.1 | 4 | 15 |
| CD248 | 1:100 | 97.7 | 195.3 | 25000 | 100000 | 2.1 | 5 | 10 |
| CTHRC1 | 1:100 | 781.3 | 1562.5 | 50000 | 200000 | 1.5 | 8 | 13 |
| PRAP1 | 1:100 | | | | | | 4 | 12 |
| NPTX2 | 1:100 | 48.8 | 97.7 | 25000 | 100000 | 2.4 | 7 | 14 |
| BGLAP | 1:100 | 781.3 | 1562.5 | 100000 | 400000 | 1.8 | 6 | 16 |
| SHISA5 | 1:100 | 390.6 | 390.6 | 25000 | 50000 | 1.8 | 6 | 9 |
| TWF2 | 1:100 | 781.3 | 781.3 | 200000 | 800000 | 2.4 | | |
| PPIF | 1:100 | 6250.0 | 6250.0 | 200000 | 800000 | 1.5 | 10 | 28 |
| TNFSF8 | 1:100 | 12.2 | 24.4 | 12500 | 50000 | 2.7 | 11 | 11 |
| PRG3 | 1:100 | 97.7 | 195.3 | 25000 | 200000 | 2.1 | 5 | 14 |
| MYH9 | 1:100 | | | | | | | |
| PRG2 | 1:100 | 97.7 | 97.7 | 25000 | 50000 | 2.4 | 5 | 15 |
| PTPRC | 1:100 | 48.8 | 48.8 | 12500 | 100000 | 2.4 | 7 | 7 |
| TREH | 1:100 | 48.8 | 97.7 | 25000 | 800000 | 2.4 | 5 | 20 |
| CYTL1 | 1:100 | 390.6 | 390.6 | 25000 | 200000 | 1.8 | 6 | 12 |
| CILP | 1:100 | | | | | | 9 | 26 |
| CLEC4M | 1:100 | 24.4 | 48.8 | 6250 | 25000 | 2.1 | 6 | 14 |
| CPQ | 1:100 | 195.3 | 195.3 | 50000 | 200000 | 2.4 | 4 | 11 |
| CHGA | 1:100 | 1562.5 | 3125.0 | 200000 | 800000 | 1.8 | 9 | 22 |

| | | | | | | | | |
|---|---|---|---|---|---|---|---|---|
| NOTCH2 | 1:100 | 48.8 | 97.7 | 6250 | 50000 | 1.8 | 5 | 6 |
| ITGBL1 | 1:100 | 97.7 | 195.3 | 25000 | 100000 | 2.1 | 6 | 13 |
| HPSE | 1:100 | | | | | | 6 | 33 |
| IGDCC4 | 1:100 | 48.8 | 48.8 | 6250 | 50000 | 2.1 | 6 | 8 |
| POSTN | 1:100 | | | | | | 9 | 15 |
| CELA2A | 1:100 | 48.8 | 48.8 | 25000 | 100000 | 2.7 | 5 | 9 |
| TNN | 1:100 | 50000.0 | 100000.0 | 6400000 | 12800000 | 1.8 | 7 | 8 |
| GM2A | 1:100 | 781.3 | 1562.5 | 50000 | 200000 | 1.5 | 7 | 17 |
| TALDO1 | 1:100 | 97.7 | 195.3 | 25000 | 200000 | 2.1 | 4 | 8 |
| STAB2 | 1:100 | 390.6 | 1562.5 | 100000 | 800000 | 1.8 | 5 | 9 |
| SLURP1 | 1:100 | 1562.5 | 1562.5 | 100000 | 800000 | 1.8 | 7 | 9 |
| CKB | 1:100 | | | | | | 9 | 15 |
| ASAH1 | 1:100 | 195.3 | 390.6 | 25000 | 200000 | 1.8 | 8 | 18 |
| LAMP1 | 1:100 | 97.7 | 390.6 | 50000 | 100000 | 2.1 | 7 | 6 |
| PSAP | 1:100 | 1562.5 | 1562.5 | 50000 | 200000 | 1.5 | 6 | 7 |
| GOT1 | 1:100 | 390.6 | 390.6 | 25000 | 50000 | 1.8 | 8 | 12 |
| NPC2 | 1:100 | 390.6 | 390.6 | 12500 | 50000 | 1.5 | 5 | 8 |
| NCF2 | 1:1 | 0.8 | 3.1 | 12500 | 25000 | 3.6 | 8 | 6 |
| IL15 | 1:1 | 1.5 | 3.1 | 50000 | 400000 | 4.2 | 9 | 4 |
| BCR | 1:1 | 781.3 | 1562.5 | 200000 | 800000 | 2.1 | 7 | 4 |
| MAP2K6 | 1:1 | 24.4 | 48.8 | 25000 | 200000 | 2.7 | 9 | 10 |
| DAPP1 | 1:1 | 3.1 | 12.2 | 25000 | 200000 | 3.3 | 10 | 9 |
| IL4R | 1:1 | 3.1 | 6.1 | 25000 | 100000 | 3.6 | 10 | 12 |
| PIK3AP1 | 1:1 | 781.3 | 1562.5 | 400000 | 800000 | 2.4 | 14 | 13 |
| CD4 | 1:1 | 97.7 | 97.7 | 400000 | 800000 | 3.6 | 8 | 9 |
| KLRD1 | 1:1 | 1.5 | 3.1 | 6250 | 25000 | 3.3 | 3 | 4 |
| BSG | 1:1 | 0.2 | 0.4 | 12500 | 50000 | 4.5 | 3 | 3 |
| SERPINB8 | 1:1 | 12.2 | 12.2 | 12500 | 50000 | 3.0 | 6 | 6 |
| VEGFD | 1:1 | 12.2 | 24.4 | 25000 | 100000 | 3.0 | 4 | 4 |
| AGRP | 1:1 | 48.8 | 97.7 | 50000 | 400000 | 2.7 | 6 | 14 |
| CCL3 | 1:1 | 0.2 | 0.4 | 781 | 1563 | 3.3 | 9 | 5 |
| TBC1D5 | 1:1 | | | | | | 7 | 7 |
| EGLN1 | 1:1 | 6.1 | 12.2 | 12500 | 25000 | 3.0 | 9 | 5 |
| NUDC | 1:1 | 24.4 | 97.7 | 100000 | 800000 | 3.0 | 4 | 5 |
| SELPLG | 1:1 | 3.1 | 6.1 | 6250 | 25000 | 3.0 | 9 | 5 |
| KLRB1 | 1:1 | 0.8 | 1.5 | 3125 | 12500 | 3.3 | 3 | 2 |
| CEACAM21 | 1:1 | 0.4 | 0.8 | 1563 | 12500 | 3.3 | 8 | 6 |
| IL17RB | 1:1 | 24.4 | 24.4 | 25000 | 400000 | 3.0 | 8 | 8 |
| IL5RA | 1:1 | 6.1 | 12.2 | 25000 | 400000 | 3.3 | 9 | 8 |
| C1QA | 1:1 | 29.2 | 58.3 | 119500 | 239000 | 3.3 | 4 | 5 |
| LY9 | 1:1 | 3.1 | 6.1 | 12500 | 50000 | 3.3 | 3 | 4 |
| HEXIM1 | 1:1 | 1.5 | 3.1 | 781 | 1563 | 2.4 | 10 | 7 |
| PCDH1 | 1:1 | 195.3 | 390.6 | 200000 | 800000 | 2.7 | 4 | 4 |
| LY75 | 1:1 | 24.4 | 48.8 | 50000 | 400000 | 3.0 | 10 | 7 |
| TLR3 | 1:1 | 1.5 | 3.1 | 12500 | 100000 | 3.6 | 4 | 3 |
| IDS | 1:1 | 97.7 | 195.3 | 400000 | 800000 | 3.3 | 3 | 4 |
| CTSO | 1:1 | 48.8 | 97.7 | 50000 | 400000 | 2.7 | 4 | 4 |
| HGF | 1:1 | 3.1 | 12.2 | 100000 | 200000 | 3.9 | 4 | 3 |
| MERTK | 1:1 | 97.7 | 195.3 | 100000 | 800000 | 2.7 | 5 | 6 |
| CCL11 | 1:1 | 1.5 | 3.1 | 25000 | 200000 | 3.9 | 3 | 4 |
| IL12RB1 | 1:1 | 97.7 | 195.3 | 100000 | 800000 | 2.7 | 8 | 17 |
| PARP1 | 1:1 | 48.8 | 97.7 | 50000 | 200000 | 2.7 | 7 | 14 |

| | | | | | | | | |
|---|---|---|---|---|---|---|---|---|
| RABGAP1L | 1:1 | 48.8 | 48.8 | 25000 | 200000 | 2.7 | 10 | 19 |
| MYO9B | 1:1 | | | | | | 10 | 19 |
| IL33 | 1:1 | 6.1 | 24.4 | 6250 | 25000 | 2.4 | 8 | 58 |
| IL2RB | 1:1 | 97.7 | 390.6 | 200000 | 800000 | 2.7 | 10 | 37 |
| LILRB4 | 1:1 | 97.7 | 195.3 | 100000 | 800000 | 2.7 | 8 | 15 |
| IL1B | 1:1 | 0.4 | 1.5 | 25000 | 100000 | 4.2 | 10 | 20 |
| IL2 | 1:1 | 0.4 | 0.8 | 6250 | 25000 | 3.9 | 7 | 20 |
| SCGN | 1:1 | 97.7 | 390.6 | 200000 | 800000 | 2.7 | 9 | 11 |
| CXCL14 | 1:1 | | | | | | 8 | 15 |
| IL22RA1 | 1:1 | 0.4 | 1.5 | 3125 | 12500 | 3.3 | 8 | 22 |
| IL10 | 1:1 | 6.1 | 24.4 | 200000 | 800000 | 3.9 | 10 | 18 |
| CEP164 | 1:1 | 12.2 | 48.8 | 12500 | 25000 | 2.4 | 7 | 18 |
| LAP3 | 1:1 | | | | | | 12 | 20 |
| NBN | 1:1 | | | | | | 11 | 9 |
| RGS8 | 1:1 | | | | | | 18 | 14 |
| MVK | 1:1 | 390.6 | 781.3 | 400000 | 800000 | 2.7 | 11 | 12 |
| WNT9A | 1:1 | 3.1 | 6.1 | 25000 | 100000 | 3.6 | 9 | 11 |
| PRKAB1 | 1:1 | 48.8 | 97.7 | 25000 | 100000 | 2.4 | 7 | 21 |
| IL11 | 1:1 | 6.1 | 12.2 | 6250 | 100000 | 2.7 | 9 | 9 |
| JUN | 1:1 | 390.6 | 781.3 | 100000 | 3200000 | 2.1 | 11 | 14 |
| ACTN4 | 1:1 | 390.6 | 781.3 | 100000 | 400000 | 2.1 | 7 | 14 |
| IL17F | 1:1 | 1.5 | 6.1 | 6250 | 25000 | 3.0 | 8 | 18 |
| IL4 | 1:1 | 0.8 | 1.5 | 6250 | 25000 | 3.6 | 7 | 33 |
| IL13 | 1:1 | 1.5 | 6.1 | 100000 | 800000 | 4.2 | 9 | 22 |
| RAB37 | 1:1 | 12.2 | 24.4 | 12500 | 100000 | 2.7 | 9 | 19 |
| IL20 | 1:1 | 24.4 | 97.7 | 25000 | 800000 | 2.4 | 7 | 16 |
| FCRL3 | 1:1 | 48.8 | 97.7 | 50000 | 800000 | 2.7 | 8 | 18 |
| AMBN | 1:1 | 12.2 | 24.4 | 25000 | 400000 | 3.0 | 9 | 15 |
| IL24 | 1:1 | 24.4 | 48.8 | 12500 | 25000 | 2.4 | 14 | 16 |
| PREB | 1:1 | 390.6 | 781.3 | 200000 | 800000 | 2.4 | 9 | 12 |
| LTO1 | 1:1 | 12.2 | 24.4 | 50000 | 100000 | 3.3 | 8 | 17 |
| PADI2 | 1:1 | 48.8 | 97.7 | 400000 | 800000 | 3.6 | 8 | 15 |
| SIT1 | 1:1 | 0.04 | 4.5 | 18500 | 37000 | 3.6 | 8 | 16 |
| IFNLR1 | 1:1 | 0.8 | 1.5 | 25000 | 100000 | 4.2 | 11 | 5 |
| ALDH3A1 | 1:1 | 0.8 | 3.1 | 3125 | 25000 | 3.0 | 10 | 10 |
| BACH1 | 1:1 | 12.2 | 24.4 | 25000 | 200000 | 3.0 | 8 | 7 |
| AOC1 | 1:1 | 97.7 | 195.3 | 100000 | 800000 | 2.7 | 10 | 21 |
| IL17A | 1:1 | 6.1 | 12.2 | 50000 | 100000 | 3.6 | 7 | 7 |
| YTHDF3 | 1:1 | 12.2 | 24.4 | 12500 | 100000 | 2.7 | 9 | 32 |
| SPINK4 | 1:1 | | | | | | 10 | 6 |
| IL3RA | 1:1 | | | | | | 7 | 28 |
| IL17D | 1:1 | 3.1 | 6.1 | 6250 | 25000 | 3.0 | 8 | 14 |
| IL17C | 1:1 | 12.2 | 24.4 | 50000 | 100000 | 3.3 | 10 | 14 |
| MILR1 | 1:1 | | | | | | 7 | 21 |
| PNPT1 | 1:1 | | | | | | 9 | 11 |
| ARTN | 1:1 | 0.8 | 1.5 | 6250 | 25000 | 3.6 | 9 | 9 |
| NRTN | 1:1 | 0.8 | 1.5 | 3125 | 25000 | 3.3 | 15 | 31 |
| CXCL12 | 1:1 | | | | | | 8 | 17 |
| PRKCQ | 1:1 | | | | | | 13 | 18 |
| TRAF2 | 1:1 | | | | | | 9 | 4 |
| WAS | 1:1 | | | | | | 8 | 31 |
| TNFAIP8 | 1:1 | | | | | | 11 | 16 |

| | | | | | | | | |
|---|---|---|---|---|---|---|---|---|
| PAPPA | 1:1 | 195.3 | 390.6 | 50000 | 200000 | 2.1 | 12 | 15 |
| DGKZ | 1:1 | 3.1 | 6.1 | 3125 | 25000 | 2.7 | 8 | 13 |
| EIF5A | 1:1 | | | | | | 11 | 10 |
| ITM2A | 1:1 | | | | | | 7 | 9 |
| PSPN | 1:1 | 3.1 | 6.1 | 12500 | 100000 | 3.3 | 11 | 5 |
| TPT1 | 1:1 | 1562.5 | 6250.0 | 400000 | 800000 | 1.8 | 16 | 19 |
| FGF2 | 1:1 | 12.2 | 12.2 | 12500 | 400000 | 3.0 | 4 | 6 |
| NUB1 | 1:1 | 3.1 | 6.1 | 12500 | 25000 | 3.3 | 8 | 12 |
| LRRN1 | 1:1 | 12.2 | 24.4 | 25000 | 400000 | 3.0 | 6 | 17 |
| TNF | 1:1 | 1.5 | 3.1 | 6250 | 100000 | 3.3 | 9 | 15 |
| IL10RA | 1:1 | 1.5 | 6.1 | 400000 | 800000 | 4.8 | 7 | 17 |
| TNFRSF13C | 1:1 | 12.2 | 24.4 | 6250 | 100000 | 2.4 | 9 | 13 |
| ARNT | 1:1 | | | | | | 8 | 14 |
| NCLN | 1:1 | | | | | | 6 | 17 |
| IL20RA | 1:1 | 0.8 | 1.5 | 12500 | 400000 | 3.9 | 8 | 13 |
| SULT2A1 | 1:1 | 195.3 | 390.6 | 200000 | 400000 | 2.7 | 10 | 8 |
| METAP1D | 1:1 | 0.4 | 0.8 | 12500 | 25000 | 4.2 | 8 | 9 |
| FABP9 | 1:1 | 3.1 | 12.2 | 12500 | 25000 | 3.0 | 10 | 7 |
| SH2D1A | 1:1 | 6.1 | 12.2 | 6250 | 25000 | 2.7 | 8 | 5 |
| NFATC3 | 1:1 | 6.1 | 24.4 | 3125 | 12500 | 2.1 | 8 | 9 |
| SRPK2 | 1:1 | 3.1 | 6.1 | 1563 | 12500 | 2.4 | 5 | 11 |
| HLA-DRA | 1:1 | 390.6 | 781.3 | 400000 | 800000 | 2.7 | 10 | 9 |
| BTN3A2 | 1:1 | 48.8 | 97.7 | 200000 | 800000 | 3.3 | 10 | 7 |
| BCL2L11 | 1:1 | | | | | | 9 | 8 |
| IL1A | 1:1 | 1.5 | 3.1 | 3148 | 12594 | 3.0 | 8 | 47 |
| IFNG | 1:1 | 1.5 | 6.1 | 12500 | 50000 | 3.3 | 15 | 22 |
| ICA1 | 1:1 | 48.8 | 195.3 | 100000 | 400000 | 2.7 | 8 | 16 |
| VASH1 | 1:1 | 195.3 | 390.6 | 100000 | 400000 | 2.4 | 6 | 9 |
| IL5 | 1:1 | 6.1 | 12.2 | 100000 | 400000 | 3.9 | 10 | 21 |
| SPRY2 | 1:1 | 24.4 | 97.7 | 50000 | 400000 | 2.7 | 9 | 15 |
| IL15RA | 1:1 | 6.1 | 12.2 | 3125 | 25000 | 2.4 | 7 | 23 |
| FGF5 | 1:1 | 3.1 | 6.1 | 50000 | 100000 | 3.9 | 11 | 19 |
| TANK | 1:1 | 48.8 | 195.3 | 100000 | 400000 | 2.7 | 8 | 15 |
| NFATC1 | 1:1 | 390.6 | 781.3 | 400000 | 800000 | 2.7 | 10 | 9 |
| CSF3 | 1:1 | 97.7 | 390.6 | 400000 | 800000 | 3.0 | 9 | 14 |
| AMN | 1:1 | | | | | | 8 | 19 |
| SLAMF1 | 1:1 | 781.3 | 1562.5 | 400000 | 800000 | 2.4 | 14 | 17 |
| IRAK1 | 1:1 | 781.3 | 3125.0 | 400000 | 800000 | 2.1 | 11 | 14 |
| FOXO1 | 1:1 | | | | | | 8 | 8 |
| GALNT3 | 1:1 | 781.3 | 1562.5 | 400000 | 800000 | 2.4 | 11 | 14 |
| PRDX3 | 1:1 | | | | | | 8 | 12 |
| GBP2 | 1:1 | 390.6 | 781.3 | 400000 | 800000 | 2.7 | 12 | 20 |
| JCHAIN | 1:1 | | | | | | 13 | 19 |
| BID | 1:1 | 781.3 | 3125.0 | 400000 | 3200000 | 2.1 | 11 | 11 |
| SCRN1 | 1:1 | 48.8 | 97.7 | 50000 | 400000 | 2.7 | 9 | 12 |
| SLC39A5 | 1:1 | | | | | | 10 | 9 |
| ENAH | 1:1 | 48.8 | 48.8 | 6250 | 25000 | 2.1 | 10 | 20 |
| IKBKG | 1:1 | 3.1 | 12.2 | 12500 | 25000 | 3.0 | 13 | 14 |
| ITGB6 | 1:1 | 1.5 | 3.1 | 6250 | 25000 | 3.3 | 8 | 14 |
| PROK1 | 1:1 | 3.1 | 12.2 | 12500 | 25000 | 3.0 | 9 | 5 |
| IL1RL2 | 1:1 | 0.8 | 1.5 | 6250 | 100000 | 3.6 | 8 | 16 |
| IL6 | 1:1 | 0.2 | 0.4 | 3125 | 25000 | 3.9 | 10 | 14 |

| | | | | | | | | |
|---|---|---|---|---|---|---|---|---|
| MLN | 1:1 | 390.6 | 781.3 | 200000 | 400000 | 2.4 | 12 | 10 |
| FXYD5 | 1:1 | 6.1 | 12.2 | 3125 | 100000 | 2.4 | 8 | 13 |
| SIGLEC10 | 1:1 | 12.2 | 48.8 | 200000 | 800000 | 3.6 | 8 | 4 |
| PSIP1 | 1:1 | | | | | | 9 | 8 |
| TRIM21 | 1:1 | 24.4 | 97.7 | 400000 | 800000 | 3.6 | 12 | 9 |
| ISM1 | 1:1 | 97.7 | 195.3 | 200000 | 400000 | 3.0 | 10 | 10 |
| IL7 | 1:1 | 1.5 | 3.1 | 6250 | 12500 | 3.3 | 12 | 9 |
| HLA-E | 1:1 | 12500.0 | 25000.0 | 1600000 | 3200000 | 1.8 | 6 | 4 |
| STX8 | 1:1 | 12.2 | 48.8 | 12500 | 25000 | 2.4 | 4 | 13 |
| LSP1 | 1:1 | 97.7 | 97.7 | 25000 | 400000 | 2.4 | 10 | 8 |
| CLEC4C | 1:1 | 24.4 | 48.8 | 25000 | 400000 | 2.7 | 8 | 4 |
| OSM | 1:1 | 0.2 | 0.4 | 781 | 3125 | 3.3 | 12 | 6 |
| SAMD9L | 1:1 | 195.3 | 390.6 | 200000 | 800000 | 2.7 | 7 | 8 |
| CD200R1 | 1:1 | 6.1 | 12.2 | 25000 | 100000 | 3.3 | 7 | 7 |
| CNTNAP2 | 1:1 | 12.2 | 24.4 | 12500 | 400000 | 2.7 | 10 | 10 |
| ANXA11 | 1:1 | 1562.5 | 3125.0 | 800000 | 800000 | 2.4 | 7 | 9 |
| FCRL6 | 1:1 | 97.7 | 195.3 | 200000 | 800000 | 3.0 | 10 | 5 |
| NPPC | 1:1 | 12.2 | 24.4 | 12500 | 25000 | 2.7 | 11 | 8 |
| BANK1 | 1:1 | 195.3 | 390.6 | 200000 | 400000 | 2.7 | 9 | 16 |
| CD83 | 1:1 | 1.5 | 3.1 | 12500 | 25000 | 3.6 | 9 | 6 |
| MAPK9 | 1:1 | | | | | | 12 | 13 |
| CCL28 | 1:1 | 97.7 | 195.3 | 100000 | 400000 | 2.7 | 11 | 12 |
| ARHGEF12 | 1:1 | 781.3 | 3125.0 | 400000 | 800000 | 2.1 | 8 | 9 |
| GOPC | 1:1 | 12.2 | 24.4 | 12500 | 25000 | 2.7 | 7 | 16 |
| PTPRM | 1:1 | 97.7 | 195.3 | 200000 | 800000 | 3.0 | 11 | 11 |
| CD40LG | 1:1 | 0.8 | 1.5 | 6250 | 12500 | 3.6 | 4 | 4 |
| MGMT | 1:1 | 195.3 | 390.6 | 100000 | 400000 | 2.4 | 12 | 13 |
| PSMG3 | 1:1 | 390.6 | 781.3 | 200000 | 400000 | 2.4 | 9 | 10 |
| DPP10 | 1:1 | 6.1 | 12.2 | 25000 | 200000 | 3.3 | 9 | 9 |
| HSD11B1 | 1:1 | | | | | | 9 | 7 |
| EDAR | 1:1 | 0.4 | 0.8 | 781 | 12500 | 3.0 | 9 | 5 |
| COL9A1 | 1:1 | 6.1 | 12.2 | 12500 | 25000 | 3.0 | 11 | 6 |
| PLXNA4 | 1:1 | 24.4 | 97.7 | 50000 | 400000 | 2.7 | 12 | 6 |
| EPO | 1:1 | 3.9 | 7.8 | 1000 | 4000 | 2.1 | 11 | 11 |
| ITGA6 | 1:1 | | | | | | 9 | 7 |
| CCL7 | 1:1 | 0.4 | 0.8 | 781 | 25000 | 3.0 | 10 | 8 |
| NCR1 | 1:1 | 3.1 | 6.1 | 12500 | 25000 | 3.3 | 9 | 6 |
| LTA | 1:1 | 0.8 | 1.5 | 3125 | 12500 | 3.3 | 8 | 4 |
| CASP2 | 1:1 | 195.3 | 390.6 | 200000 | 800000 | 2.7 | 7 | 5 |
| FCAR | 1:1 | 0.8 | 3.1 | 12500 | 25000 | 3.6 | 9 | 9 |
| IRAK4 | 1:1 | 390.6 | 1562.5 | 400000 | 800000 | 2.4 | 9 | 14 |
| CLEC4G | 1:1 | 3.1 | 6.1 | 50000 | 200000 | 3.9 | 10 | 6 |
| HPCAL1 | 1:1 | 48.8 | 97.7 | 50000 | 200000 | 2.7 | 4 | 5 |
| CCL26 | 1:1 | 97.7 | 781.3 | 400000 | 800000 | 2.7 | 15 | 17 |
| ITGA11 | 1:1 | 97.7 | 390.6 | 200000 | 800000 | 2.7 | 10 | 15 |
| NT5C3A | 1:1 | 97.7 | 195.3 | 200000 | 800000 | 3.0 | 9 | 7 |
| KRT19 | 1:1 | 48.8 | 97.7 | 100000 | 400000 | 3.0 | 8 | 9 |
| RAB6A | 1:1 | 48.8 | 97.7 | 200000 | 800000 | 3.3 | 8 | 8 |
| CD84 | 1:1 | 24.4 | 48.8 | 12500 | 200000 | 2.4 | 9 | 7 |
| CXADR | 1:1 | 3.1 | 3.1 | 1563 | 12500 | 2.7 | 12 | 7 |
| CD70 | 1:1 | 12.2 | 48.8 | 100000 | 200000 | 3.3 | 10 | 6 |
| MICA_MICB | 1:1 | 6.1 | 12.2 | 12500 | 25000 | 3.0 | 10 | 8 |

| | | | | | | | | |
|---|---|---|---|---|---|---|---|---|
| TNFSF11 | 1:1 | 12.2 | 24.4 | 50000 | 200000 | 3.3 | 11 | 7 |
| CLIP2 | 1:1 | 195.3 | 195.3 | 50000 | 800000 | 2.4 | 13 | 20 |
| TRIM5 | 1:1 | | | | | | 10 | 17 |
| PTX3 | 1:1 | | | | | | 9 | 7 |
| CKAP4 | 1:1 | | | | | | 3 | 6 |
| CXCL6 | 1:1 | | | | | | 6 | 13 |
| NTF3 | 1:1 | 0.4 | 0.8 | 3125 | 12500 | 3.6 | 15 | 7 |
| ICAM4 | 1:1 | 781.3 | 1562.5 | 400000 | 800000 | 2.4 | 9 | 5 |
| DECR1 | 1:1 | | | | | | 11 | 11 |
| TNFSF10 | 1:1 | 24.4 | 97.7 | 25000 | 50000 | 2.4 | 4 | 4 |
| CLEC4D | 1:1 | 3.1 | 6.1 | 12500 | 25000 | 3.3 | 10 | 8 |
| NFASC | 1:1 | | | | | | 3 | 2 |
| FLT3LG | 1:1 | 0.4 | 0.8 | 1563 | 12500 | 3.3 | 6 | 4 |
| IL16 | 1:1 | 1.5 | 3.1 | 3125 | 12500 | 3.0 | 3 | 6 |
| CDSN | 1:1 | 390.6 | 781.3 | 400000 | 800000 | 2.7 | 4 | 4 |
| AXIN1 | 1:1 | 781.3 | 1562.5 | 400000 | 800000 | 2.4 | 6 | 9 |
| SLAMF7 | 1:1 | 195.3 | 390.6 | 200000 | 800000 | 2.7 | 10 | 6 |
| CLEC4A | 1:1 | 0.8 | 3.1 | 6250 | 25000 | 3.3 | 9 | 7 |
| LAT | 1:1 | 48.8 | 97.7 | 25000 | 100000 | 2.4 | 7 | 8 |
| CXCL8 | 1:1 | 0.1 | 0.2 | 1563 | 6250 | 3.9 | 3 | 4 |
| PPP1R9B | 1:1 | 12.2 | 24.4 | 25000 | 800000 | 3.0 | 7 | 12 |
| CD200 | 1:1 | 6.1 | 12.2 | 3125 | 25000 | 2.4 | 10 | 6 |
| HCLS1 | 1:1 | 195.3 | 195.3 | 25000 | 400000 | 2.1 | 8 | 8 |
| CD244 | 1:1 | 1.5 | 3.1 | 6250 | 25000 | 3.3 | 4 | 5 |
| KYNU | 1:1 | 48.8 | 48.8 | 400000 | 800000 | 3.9 | 3 | 11 |
| DFFA | 1:1 | 0.8 | 3.1 | 6250 | 50000 | 3.3 | 5 | 6 |
| IL18R1 | 1:1 | 0.2 | 0.8 | 6250 | 25000 | 3.9 | 3 | 3 |
| ADAM23 | 1:1 | 12.2 | 24.4 | 50000 | 200000 | 3.3 | 5 | 3 |
| LAMP3 | 1:1 | 12.2 | 24.4 | 25000 | 200000 | 3.0 | 5 | 4 |
| IL32 | 1:1 | 0.8 | 3.1 | 12500 | 25000 | 3.6 | 9 | 7 |
| GMPR | 1:1 | | | | | | 4 | 4 |
| CD6 | 1:1 | 1.5 | 1.5 | 6250 | 25000 | 3.6 | 4 | 4 |
| CD22 | 1:1 | 3.1 | 6.1 | 50000 | 200000 | 3.9 | 3 | 3 |
| CXCL17 | 1:1 | 3.1 | 6.1 | 12500 | 50000 | 3.3 | 4 | 4 |
| FKBP1B | 1:1 | 390.6 | 781.3 | 200000 | 400000 | 2.4 | 5 | 26 |
| GZMA | 1:1 | 390.6 | 390.6 | 800000 | 800000 | 3.3 | 5 | 5 |
| CD160 | 1:1 | 1.5 | 3.1 | 12500 | 25000 | 3.6 | 3 | 7 |
| TNFRSF4 | 1:1 | 0.8 | 1.5 | 6250 | 12500 | 3.6 | 5 | 4 |
| TGFB1 | 1:1 | | | | | | 4 | 4 |
| TPSAB1 | 1:1 | 6.1 | 12.2 | 100000 | 200000 | 3.9 | 4 | 3 |
| EIF4G1 | 1:1 | 12.2 | 24.4 | 25000 | 200000 | 3.0 | 6 | 10 |
| FASLG | 1:1 | 0.2 | 0.2 | 6250 | 12500 | 4.5 | 4 | 4 |
| CD79B | 1:1 | 0.8 | 1.5 | 6250 | 25000 | 3.6 | 4 | 4 |
| PTH1R | 1:1 | 3.1 | 3.1 | 6250 | 25000 | 3.3 | 7 | 8 |
| TNFRSF11A | 1:1 | 0.8 | 1.5 | 6250 | 12500 | 3.6 | 4 | 4 |
| FCRL2 | 1:1 | 24.4 | 48.8 | 50000 | 200000 | 3.0 | 5 | 3 |
| CLEC7A | 1:1 | 3.1 | 6.1 | 3125 | 12500 | 2.7 | 3 | 5 |
| CLSTN2 | 1:1 | 6.1 | 6.1 | 25000 | 200000 | 3.6 | 5 | 3 |
| IL12B | 1:1 | 0.4 | 0.8 | 3125 | 12500 | 3.6 | 5 | 4 |
| LIFR | 1:1 | 6.1 | 48.8 | 12500 | 400000 | 2.4 | 12 | 7 |
| CCL13 | 1:1 | 0.8 | 6.1 | 3125 | 6250 | 2.7 | 5 | 5 |
| ADA | 1:1 | 781.3 | 1562.5 | 3200000 | 12800000 | 3.3 | 4 | 3 |

| | | | | | | | | |
|---|---|---|---|---|---|---|---|---|
| PRDX5 | 1:1 | 1562.5 | 3125.0 | 800000 | 800000 | 2.4 | 4 | 25 |
| DNAJA2 | 1:1 | 97.7 | 390.6 | 400000 | 800000 | 3.0 | 6 | 16 |
| VEGFA | 1:1 | 0.8 | 1.5 | 6250 | 12500 | 3.6 | 7 | 7 |
| TNFSF12 | 1:1 | 12.2 | 24.4 | 100000 | 400000 | 3.6 | 5 | 4 |
| CCL25 | 1:1 | 3.1 | 24.4 | 25000 | 100000 | 3.0 | 5 | 6 |
| GZMB | 1:1 | | | | | | 12 | 7 |
| TGFA | 1:1 | 0.2 | 0.4 | 1563 | 25000 | 3.6 | 12 | 7 |
| CCL20 | 1:1 | 3.1 | 6.1 | 6250 | 25000 | 3.0 | 4 | 5 |
| MMP1 | 1:1 | 0.8 | 1.5 | 3125 | 12500 | 3.3 | 4 | 5 |
| PGF | 1:1 | 0.4 | 0.8 | 6250 | 12500 | 3.9 | 4 | 3 |
| ANGPTL4 | 1:10 | 12.2 | 24.4 | 25000 | 50000 | 3.0 | 7 | 4 |
| FGF19 | 1:10 | 3.1 | 6.1 | 12500 | 50000 | 3.3 | 6 | 4 |
| TNFRSF13B | 1:10 | 6.1 | 24.4 | 50000 | 200000 | 3.3 | 8 | 4 |
| CRKL | 1:10 | 6.1 | 12.2 | 25000 | 100000 | 3.3 | 6 | 8 |
| PLAUR | 1:10 | 0.2 | 0.4 | 1563 | 6250 | 3.6 | 6 | 4 |
| DNPH1 | 1:10 | 6.1 | 12.2 | 6250 | 12500 | 2.7 | 7 | 8 |
| SPON1 | 1:10 | 48.8 | 97.7 | 100000 | 200000 | 3.0 | 5 | 4 |
| ESM1 | 1:10 | 3.1 | 6.1 | 6250 | 25000 | 3.0 | 8 | 5 |
| PRSS8 | 1:10 | 0.4 | 0.8 | 6250 | 12500 | 3.9 | 8 | 3 |
| AGRN | 1:10 | 6.1 | 12.2 | 6250 | 12500 | 2.7 | 7 | 5 |
| PTPN6 | 1:10 | 781.3 | 1562.5 | 100000 | 200000 | 1.8 | 6 | 7 |
| CXCL9 | 1:10 | 0.05 | 0.1 | 6250 | 12500 | 4.8 | 7 | 5 |
| ATP5IF1 | 1:10 | 195.3 | 390.6 | 50000 | 200000 | 2.1 | 8 | 7 |
| LHPP | 1:10 | 0.8 | 1.5 | 12500 | 25000 | 3.9 | 8 | 5 |
| CXCL3 | 1:10 | 12.2 | 48.8 | 50000 | 200000 | 3.0 | 10 | 9 |
| ENPP7 | 1:10 | 0.8 | 1.5 | 12500 | 200000 | 3.9 | 6 | 4 |
| CXCL1 | 1:10 | 0.4 | 0.8 | 6250 | 12500 | 3.9 | 8 | 8 |
| SMOC2 | 1:10 | 24.4 | 48.8 | 50000 | 200000 | 3.0 | 7 | 3 |
| CCL17 | 1:10 | 0.4 | 0.8 | 781 | 3125 | 3.0 | 7 | 14 |
| LGMN | 1:10 | 0.8 | 3.1 | 6250 | 50000 | 3.3 | 9 | 5 |
| LGALS9 | 1:10 | 48.8 | 97.7 | 50000 | 200000 | 2.7 | 7 | 4 |
| TFF2 | 1:10 | 0.8 | 1.5 | 3125 | 12500 | 3.3 | 8 | 6 |
| CTSC | 1:10 | 24.4 | 48.8 | 50000 | 200000 | 3.0 | 8 | 12 |
| CD276 | 1:10 | 97.7 | 195.3 | 400000 | 800000 | 3.3 | 7 | 4 |
| CCL23 | 1:10 | 6.1 | 12.2 | 6250 | 25000 | 2.7 | 6 | 8 |
| F2R | 1:10 | 48.8 | 195.3 | 200000 | 400000 | 3.0 | 8 | 5 |
| TREM2 | 1:10 | | | | | | 6 | 10 |
| SHMT1 | 1:10 | 12.2 | 24.4 | 100000 | 400000 | 3.6 | 8 | 5 |
| ANGPT1 | 1:10 | 12.2 | 24.4 | 100000 | 200000 | 3.6 | 6 | 3 |
| CCL22 | 1:10 | 6.1 | 6.1 | 3125 | 12500 | 2.7 | 14 | 20 |
| TNFSF13 | 1:10 | 3.1 | 6.1 | 25000 | 100000 | 3.6 | 5 | 5 |
| PLA2G4A | 1:10 | 781.3 | 1562.5 | 400000 | 800000 | 2.4 | 6 | 7 |
| PON3 | 1:10 | 390.6 | 390.6 | 200000 | 800000 | 2.7 | 5 | 9 |
| DAG1 | 1:10 | 3.1 | 6.1 | 6250 | 12500 | 3.0 | 7 | 4 |
| SIGLEC1 | 1:10 | 12.2 | 24.4 | 100000 | 200000 | 3.6 | 7 | 4 |
| DNER | 1:10 | 1.5 | 3.1 | 25000 | 100000 | 3.9 | 5 | 4 |
| MGLL | 1:10 | 1562.5 | 12500.0 | 3200000 | 12800000 | 2.4 | 9 | 9 |
| CCL21 | 1:10 | 24.4 | 48.8 | 200000 | 800000 | 3.6 | 8 | 4 |
| CD58 | 1:10 | 0.4 | 0.8 | 3125 | 12500 | 3.6 | 6 | 4 |
| CD48 | 1:10 | 6.1 | 12.2 | 100000 | 200000 | 3.9 | 6 | 4 |
| COLEC12 | 1:10 | 1.5 | 3.1 | 6250 | 25000 | 3.3 | 7 | 6 |
| LAIR1 | 1:10 | 0.1 | 0.2 | 391 | 3125 | 3.3 | 7 | 4 |

| | | | | | | | | |
|---|---|---|---|---|---|---|---|---|
| CSF1 | 1:10 | 0.05 | 0.2 | 3125 | 6250 | 4.2 | 4 | 3 |
| PRELP | 1:10 | 6.1 | 12.2 | 12500 | 200000 | 3.0 | 6 | 4 |
| EPCAM | 1:10 | 0.4 | 0.4 | 6250 | 25000 | 4.2 | 7 | 7 |
| EGF | 1:10 | 0.1 | 0.2 | 781 | 1563 | 3.6 | 13 | 9 |
| CXCL10 | 1:10 | 0.2 | 6.1 | 6250 | 25000 | 3.0 | 7 | 9 |
| SMPDL3A | 1:10 | 6.1 | 12.2 | 100000 | 200000 | 3.9 | 6 | 4 |
| BTN2A1 | 1:10 | 3.1 | 6.1 | 6250 | 25000 | 3.0 | 7 | 4 |
| SPINT2 | 1:10 | 6.1 | 12.2 | 25000 | 50000 | 3.3 | 8 | 6 |
| ROBO1 | 1:10 | 3.1 | 6.1 | 100000 | 200000 | 4.2 | 6 | 4 |
| MZB1 | 1:10 | 0.4 | 0.8 | 6250 | 50000 | 3.9 | 7 | 2 |
| TIMP3 | 1:10 | 390.6 | 781.3 | 200000 | 400000 | 2.4 | 13 | 13 |
| NCK2 | 1:10 | | | | | | 5 | 10 |
| CST7 | 1:10 | 0.4 | 0.8 | 6250 | 12500 | 3.9 | 7 | 5 |
| MATN2 | 1:10 | 24.4 | 48.8 | 25000 | 50000 | 2.7 | 8 | 3 |
| CRLF1 | 1:10 | 390.6 | 781.3 | 100000 | 800000 | 2.1 | 6 | 7 |
| DBNL | 1:10 | 24.4 | 97.7 | 50000 | 200000 | 2.7 | 10 | 10 |
| AGER | 1:10 | 0.8 | 3.1 | 6250 | 25000 | 3.3 | 7 | 5 |
| IL10RB | 1:10 | 1.5 | 6.1 | 3125 | 12500 | 2.7 | 8 | 3 |
| GLOD4 | 1:10 | 0.8 | 3.1 | 6250 | 25000 | 3.3 | 7 | 4 |
| EPHA1 | 1:10 | 3.1 | 6.1 | 50000 | 200000 | 3.9 | 7 | 3 |
| IL1R2 | 1:10 | 0.8 | 1.5 | 6250 | 12500 | 3.6 | 8 | 3 |
| ENPP5 | 1:10 | 6.1 | 6.1 | 3125 | 12500 | 2.7 | 4 | 3 |
| CRIM1 | 1:10 | 1.5 | 3.1 | 6250 | 50000 | 3.3 | 8 | 4 |
| IFNGR1 | 1:10 | 1.5 | 3.1 | 3125 | 12500 | 3.0 | 7 | 4 |
| ERBB3 | 1:10 | 6.1 | 12.2 | 12500 | 50000 | 3.0 | 7 | 5 |
| MMP10 | 1:10 | 0.2 | 0.4 | 6250 | 12500 | 4.2 | 7 | 7 |
| PDLIM7 | 1:10 | 24.4 | 48.8 | 25000 | 50000 | 2.7 | 7 | 11 |
| SCGB1A1 | 1:10 | 390.6 | 781.3 | 400000 | 800000 | 2.7 | 6 | 4 |
| IL1RN | 1:10 | 0.2 | 0.4 | 1563 | 12500 | 3.6 | 6 | 3 |
| LY6D | 1:10 | 6.1 | 12.2 | 12500 | 200000 | 3.0 | 9 | 5 |
| IL18 | 1:10 | 0.1 | 0.2 | 6250 | 12500 | 4.5 | 5 | 4 |
| CCL4 | 1:10 | 0.2 | 0.4 | 1563 | 200000 | 3.6 | 7 | 5 |
| NELL2 | 1:10 | 12.2 | 24.4 | 50000 | 100000 | 3.3 | 6 | 2 |
| LTBR | 1:10 | 0.4 | 0.8 | 6250 | 12500 | 3.9 | 7 | 5 |
| PKLR | 1:10 | 3.1 | 6.1 | 50000 | 200000 | 3.9 | 6 | 7 |
| MANF | 1:10 | 3.1 | 6.1 | 12500 | 25000 | 3.3 | 7 | 14 |
| SIRPB1 | 1:10 | 1.5 | 1.5 | 6250 | 25000 | 3.6 | 4 | 4 |
| CCN2 | 1:10 | 24.4 | 48.8 | 25000 | 200000 | 2.7 | 4 | 5 |
| HSPA1A | 1:10 | 48.8 | 97.7 | 50000 | 200000 | 2.7 | 6 | 4 |
| TNFRSF11B | 1:10 | 0.1 | 0.2 | 6250 | 25000 | 4.5 | 7 | 3 |
| SCG3 | 1:10 | 12.2 | 24.4 | 25000 | 200000 | 3.0 | 7 | 4 |
| TPP1 | 1:10 | 1.5 | 3.1 | 12500 | 100000 | 3.6 | 6 | 3 |
| MEGF10 | 1:10 | 87.9 | 175.8 | 90000 | 360000 | 2.7 | 5 | 5 |
| CDON | 1:10 | 12.2 | 24.4 | 50000 | 200000 | 3.3 | 6 | 4 |
| MPIG6B | 1:10 | 0.8 | 1.5 | 1563 | 3125 | 3.0 | 12 | 14 |
| LGALS4 | 1:10 | 1.5 | 6.1 | 6250 | 25000 | 3.0 | 5 | 4 |
| SKAP2 | 1:10 | 24.4 | 48.8 | 50000 | 200000 | 3.0 | 9 | 11 |
| PDGFB | 1:10 | 6.1 | 6.1 | 6250 | 12500 | 3.0 | 10 | 11 |
| SCGB3A2 | 1:10 | 3.1 | 12.2 | 50000 | 200000 | 3.6 | 8 | 11 |
| FSTL3 | 1:10 | 12.2 | 12.2 | 6250 | 25000 | 2.7 | 6 | 5 |
| ANGPTL2 | 1:10 | 781.3 | 1562.5 | 6400000 | 12800000 | 3.6 | 9 | 8 |
| REG4 | 1:10 | 48.8 | 97.7 | 200000 | 400000 | 3.3 | 7 | 9 |

| | | | | | | | | |
|---|---|---|---|---|---|---|---|---|
| CRHBP | 1:10 | 24.4 | 97.7 | 100000 | 800000 | 3.0 | 7 | 5 |
| OMD | 1:10 | 195.3 | 390.6 | 50000 | 200000 | 2.1 | 7 | 3 |
| NME3 | 1:10 | 6.1 | 12.2 | 6250 | 25000 | 2.7 | 8 | 5 |
| FIS1 | 1:10 | 195.3 | 781.3 | 800000 | 800000 | 3.0 | 9 | 12 |
| FST | 1:10 | 12.2 | 24.4 | 25000 | 50000 | 3.0 | 5 | 6 |
| B4GALT1 | 1:10 | 24.4 | 48.8 | 100000 | 200000 | 3.3 | 10 | 6 |
| CKMT1A_CKMT1 | 1:10 | 48.8 | 97.7 | 400000 | 800000 | 3.6 | 7 | 15 |
| ADGRE2 | 1:10 | 0.8 | 1.5 | 6250 | 25000 | 3.6 | 7 | 5 |
| MEPE | 1:10 | 24.4 | 48.8 | 12500 | 25000 | 2.4 | 8 | 3 |
| CRELD2 | 1:10 | 3.1 | 6.1 | 12500 | 200000 | 3.3 | 8 | 3 |
| CTRC | 1:10 | 3.1 | 6.1 | 12500 | 50000 | 3.3 | 7 | 9 |
| FABP1 | 1:10 | 3.1 | 6.1 | 100000 | 200000 | 4.2 | 8 | 5 |
| LAMA4 | 1:10 | 12.2 | 24.4 | 100000 | 200000 | 3.6 | 7 | 5 |
| TNFRSF14 | 1:10 | 0.8 | 1.5 | 6250 | 25000 | 3.6 | 8 | 4 |
| GAL | 1:10 | 24.4 | 48.8 | 100000 | 200000 | 3.3 | 8 | 7 |
| WFIKKN2 | 1:10 | 3.1 | 6.1 | 6250 | 25000 | 3.0 | 5 | 4 |
| OSCAR | 1:10 | 3.1 | 6.1 | 6250 | 200000 | 3.0 | 7 | 5 |
| CCL24 | 1:10 | 0.8 | 0.8 | 3125 | 6250 | 3.6 | 8 | 10 |
| CD40 | 1:10 | 0.1 | 0.2 | 3125 | 12500 | 4.2 | 9 | 8 |
| PNLIPRP2 | 1:10 | 3.1 | 6.1 | 25000 | 200000 | 3.6 | 6 | 4 |
| CHRDL1 | 1:10 | 390.6 | 390.6 | 100000 | 200000 | 2.4 | 8 | 6 |
| DAND5 | 1:1 | | | | | | 10 | |
| FOLH1 | 1:1 | 6250.0 | 6250.0 | 400000 | 800000 | 1.8 | 12 | 14 |
| RAP1A | 1:1 | | | | | | 11 | 16 |
| GLI2 | 1:1 | 195.3 | 195.3 | 12500 | 25000 | 1.8 | | |
| FGF12 | 1:1 | 24.4 | 48.8 | 3125 | 25000 | 1.8 | | |
| TLR4 | 1:1 | | | | | | | |
| MKI67 | 1:1 | 48.8 | 97.7 | 12500 | 100000 | 2.1 | 13 | 24 |
| CCNE1 | 1:1 | 390.6 | 781.3 | 50000 | 400000 | 1.8 | | |
| UBE2Z | 1:1 | 390.6 | 781.3 | 100000 | 800000 | 2.1 | 13 | 21 |
| PRR5 | 1:1 | 3125.0 | 6250.0 | 400000 | 800000 | 1.8 | 13 | 0.3 |
| CEBPA | 1:1 | | | | | | | |
| KDM3A | 1:1 | 1562.5 | 3125.0 | 200000 | 800000 | 1.8 | | |
| FGF20 | 1:1 | 3125.0 | 6250.0 | 800000 | 800000 | 2.1 | 6 | 18 |
| PGR | 1:1 | 781.3 | 781.3 | 200000 | 800000 | 2.4 | 10 | 9 |
| IL20RB | 1:1 | 390.6 | 781.3 | 200000 | 800000 | 2.4 | | |
| NPHS2 | 1:1 | 390.6 | 781.3 | 50000 | 800000 | 1.8 | 6 | 20 |
| NME1 | 1:1 | 3125.0 | 3125.0 | 200000 | 400000 | 1.8 | | |
| LRIG3 | 1:1 | | | | | | | |
| GAD2 | 1:1 | 1562.5 | 6250.0 | 400000 | 800000 | 1.8 | | |
| CD3G | 1:1 | 390.6 | 781.3 | 50000 | 400000 | 1.8 | 10 | 13 |
| POLR2A | 1:1 | | | | | | 6 | |
| EP300 | 1:1 | 390.6 | 390.6 | 12500 | 25000 | 1.5 | 3 | |
| IL21R | 1:1 | 781.3 | 781.3 | 50000 | 400000 | 1.8 | | |
| PTPN9 | 1:1 | 1562.5 | 6250.0 | 400000 | 800000 | 1.8 | | |
| ST8SIA1 | 1:1 | 390.6 | 1562.5 | 200000 | 800000 | 2.1 | 14 | 20 |
| HDAC8 | 1:1 | | | | | | | |
| FGF6 | 1:1 | 1562.5 | 3125.0 | 400000 | 800000 | 2.1 | 13 | 16 |
| RELB | 1:1 | | | | | | 12 | 7 |
| TNFSF9 | 1:1 | | | | | | 12 | 17 |
| ESR1 | 1:1 | | | | | | 8 | 29 |
| FOXJ3 | 1:1 | 195.3 | 390.6 | 25000 | 800000 | 1.8 | 12 | 12 |

| | | | | | | | | |
|---|---|---|---|---|---|---|---|---|
| YY1 | 1:1 | | | | | | | |
| RICTOR | 1:1 | | | | | | 4 | 17 |
| STX5 | 1:1 | | | | | | 7 | 10 |
| UNC5D | 1:1 | 48.8 | 97.7 | 12500 | 100000 | 2.1 | 15 | 14 |
| DDX4 | 1:1 | 195.3 | 195.3 | 12500 | 100000 | 1.8 | | |
| PALLD | 1:1 | 781.3 | 1562.5 | 100000 | 400000 | 1.8 | 10 | 27 |
| HIF1A | 1:1 | 12.2 | 24.4 | 1563 | 12500 | 1.8 | 8 | 20 |
| RPA2 | 1:1 | | | | | | | |
| IL12RB2 | 1:1 | 3125.0 | 3125.0 | 400000 | 800000 | 2.1 | 2 | 28 |
| PTP4A3 | 1:1 | | | | | | 8 | 10 |
| EDNRB | 1:1 | 48.8 | 97.7 | 6250 | 25000 | 1.8 | 8 | 20 |
| SSBP1 | 1:1 | | | | | | 12 | 16 |
| WASL | 1:1 | | | | | | | |
| KLRF1 | 1:1 | 6.1 | 12.2 | 3125 | 25000 | 2.4 | 9 | 13 |
| BABAM1 | 1:1 | | | | | | 4 | |
| SPRED2 | 1:1 | | | | | | 14 | 21 |
| PIKFYVE | 1:1 | | | | | | 8 | 12 |
| TPSG1 | 1:1 | 390.6 | 390.6 | 25000 | 800000 | 1.8 | 19 | |
| SIRT1 | 1:1 | 195.3 | 390.6 | 25000 | 200000 | 1.8 | 9 | 19 |
| TP53BP1 | 1:1 | 390.6 | 1562.5 | 400000 | 800000 | 2.4 | 11 | 18 |
| SMPD3 | 1:1 | 3125.0 | 6250.0 | 400000 | 800000 | 1.8 | 10 | |
| POF1B | 1:1 | 390.6 | 390.6 | 100000 | 200000 | 2.4 | 17 | 24 |
| TRAF3 | 1:1 | | | | | | 10 | 13 |
| IL31 | 1:1 | 781.3 | 1562.5 | 200000 | 800000 | 2.1 | 8 | 9 |
| NEDD9 | 1:1 | 195.3 | 195.3 | 12500 | 50000 | 1.8 | 11 | |
| AKR7L | 1:1 | | | | | | 15 | 23 |
| BAG4 | 1:1 | | | | | | 13 | 14 |
| TGFBR1 | 1:1 | 390.6 | 390.6 | 50000 | 200000 | 2.1 | 1 | |
| MCEMP1 | 1:1 | 24.4 | 48.8 | 6250 | 25000 | 2.1 | 21 | 28 |
| IGFL4 | 1:1 | 24.4 | 48.8 | 12500 | 50000 | 2.4 | 13 | 25 |
| MRPS16 | 1:1 | 3125.0 | 6250.0 | 400000 | 800000 | 1.8 | | |
| RNF168 | 1:1 | | | | | | | |
| HS6ST2 | 1:1 | | | | | | 9 | 24 |
| CEMIP2 | 1:1 | 48.8 | 195.3 | 25000 | 200000 | 2.1 | 12 | 14 |
| BCL2L15 | 1:1 | 48.8 | 97.7 | 12500 | 50000 | 2.1 | 13 | 15 |
| ALPI | 1:1 | 390.6 | 781.3 | 25000 | 100000 | 1.5 | 10 | 36 |
| REPS1 | 1:1 | | | | | | 14 | 16 |
| TSC1 | 1:1 | 3125.0 | 6250.0 | 400000 | 800000 | 1.8 | 14 | 21 |
| XIAP | 1:1 | 3125.0 | 3125.0 | 400000 | 800000 | 2.1 | 8 | 15 |
| CPOX | 1:1 | 6250.0 | 6250.0 | 800000 | 800000 | 2.1 | 9 | 17 |
| DDX39A | 1:1 | 97.7 | 390.6 | 25000 | 200000 | 1.8 | 5 | 16 |
| DENR | 1:1 | 390.6 | 781.3 | 25000 | 100000 | 1.5 | 11 | 21 |
| CSNK1D | 1:1 | | | | | | 12 | 36 |
| THSD1 | 1:1 | 1562.5 | 3125.0 | 200000 | 800000 | 1.8 | 7 | 35 |
| IL31RA | 1:1 | 1562.5 | 3125.0 | 200000 | 800000 | 1.8 | 11 | 16 |
| RBPMS | 1:1 | | | | | | 10 | 16 |
| DAPK2 | 1:1 | 1562.5 | 1562.5 | 100000 | 800000 | 1.8 | 18 | 22 |
| SUMF1 | 1:1 | 3125.0 | 6250.0 | 400000 | 800000 | 1.8 | 8 | 17 |
| CA8 | 1:1 | 3125.0 | 6250.0 | 800000 | 6400000 | 2.1 | | |
| EVI5 | 1:1 | 195.3 | 390.6 | 100000 | 200000 | 2.4 | 9 | 15 |
| ANKMY2 | 1:1 | 781.3 | 1562.5 | 100000 | 400000 | 1.8 | 11 | 23 |
| TERF1 | 1:1 | 781.3 | 1562.5 | 50000 | 400000 | 1.5 | 12 | |

| | | | | | | | | |
|---|---|---|---|---|---|---|---|---|
| PRSS22 | 1:1 | | | | | | 12 | 12 |
| ADAMTS4 | 1:1 | 781.3 | 781.3 | 100000 | 800000 | 2.1 | 13 | 25 |
| TYRP1 | 1:1 | 6250.0 | 12500.0 | 800000 | 800000 | 1.8 | 10 | 16 |
| SHH | 1:1 | | | | | | | |
| C1QL2 | 1:1 | 1562.5 | 1562.5 | 100000 | 400000 | 1.8 | 10 | 16 |
| RNF31 | 1:1 | | | | | | 11 | 15 |
| PLCB1 | 1:1 | 6250.0 | 6250.0 | 400000 | 800000 | 1.8 | | |
| PSMG4 | 1:1 | | | | | | 12 | 12 |
| ADAM12 | 1:1 | 390.6 | 781.3 | 50000 | 800000 | 1.8 | 14 | 14 |
| IL36A | 1:1 | 195.3 | 195.3 | 25000 | 200000 | 2.1 | 21 | |
| ZNF174 | 1:1 | 390.6 | 390.6 | 25000 | 200000 | 1.8 | 9 | 18 |
| NAGA | 1:1 | 3125.0 | 6250.0 | 400000 | 800000 | 1.8 | 13 | 17 |
| TSPYL1 | 1:1 | 390.6 | 1562.5 | 100000 | 800000 | 1.8 | 9 | 17 |
| PER3 | 1:1 | | | | | | 16 | 37 |
| CD36 | 1:1 | 781.3 | 781.3 | 50000 | 400000 | 1.8 | 9 | 14 |
| ACRV1 | 1:1 | 12.2 | 48.8 | 6250 | 50000 | 2.1 | 9 | 24 |
| SLITRK1 | 1:1 | 195.3 | 195.3 | 25000 | 200000 | 2.1 | 11 | 19 |
| LMOD1 | 1:1 | | | | | | 17 | 23 |
| IDO1 | 1:1 | 195.3 | 390.6 | 100000 | 400000 | 2.4 | 13 | 17 |
| SCRIB | 1:1 | | | | | | 16 | 26 |
| NDUFA5 | 1:1 | 781.3 | 1562.5 | 200000 | 800000 | 2.1 | 16 | 17 |
| NECTIN1 | 1:1 | | | | | | 9 | 13 |
| FGF3 | 1:1 | 1562.5 | 1562.5 | 100000 | 800000 | 1.8 | | |
| GIPR | 1:1 | 24.4 | 48.8 | 12500 | 50000 | 2.4 | | |
| BLNK | 1:1 | 12500.0 | 12500.0 | 800000 | 800000 | 1.8 | 16 | 18 |
| NPHS1 | 1:1 | 781.3 | 1562.5 | 100000 | 400000 | 1.8 | 12 | 17 |
| ERMAP | 1:1 | 12.2 | 24.4 | 1563 | 6250 | 1.8 | 13 | 15 |
| MTDH | 1:1 | 781.3 | 781.3 | 50000 | 100000 | 1.8 | | |
| ITGAL | 1:1 | | | | | | 13 | 30 |
| STAT2 | 1:1 | 781.3 | 1562.5 | 100000 | 800000 | 1.8 | 11 | 24 |
| FGF16 | 1:1 | 6250.0 | 6250.0 | 400000 | 800000 | 1.8 | 17 | 14 |
| DEFB103A | 1:1 | | | | | | 6 | 26 |
| GIT1 | 1:1 | | | | | | 15 | 17 |
| TLR1 | 1:1 | 12.2 | 24.4 | 12500 | 100000 | 2.7 | 14 | 17 |
| SEMA6C | 1:1 | 781.3 | 1562.5 | 200000 | 800000 | 2.1 | 10 | 14 |
| CASP9 | 1:1 | | | | | | | |
| S100A13 | 1:1 | 3.1 | 12.2 | 12500 | 400000 | 3.0 | 10 | 31 |
| ADAMTS1 | 1:1 | 390.6 | 781.3 | 50000 | 100000 | 1.8 | 14 | 13 |
| DGKA | 1:1 | 6250.0 | 12500.0 | 800000 | 800000 | 1.8 | 8 | 18 |
| CSF3R | 1:1 | 97.7 | 195.3 | 12500 | 100000 | 1.8 | 10 | 11 |
| PPM1B | 1:1 | 1562.5 | 3125.0 | 200000 | 800000 | 1.8 | 9 | 17 |
| PDZK1 | 1:1 | 3125.0 | 6250.0 | 400000 | 800000 | 1.8 | 15 | 18 |
| NUMB | 1:1 | 390.6 | 1562.5 | 100000 | 800000 | 1.8 | 8 | 30 |
| RALB | 1:1 | | | | | | 21 | |
| INSR | 1:1 | 1562.5 | 1562.5 | 50000 | 400000 | 1.5 | 6 | 6 |
| NXPH3 | 1:1 | 390.6 | 390.6 | 25000 | 200000 | 1.8 | 9 | 12 |
| PPL | 1:1 | 48.8 | 97.7 | 6250 | 50000 | 1.8 | 9 | 14 |
| PTGES2 | 1:1 | | | | | | 17 | 27 |
| STX7 | 1:1 | | | | | | 10 | 19 |
| TOP2B | 1:1 | 781.3 | 1562.5 | 50000 | 400000 | 1.5 | 11 | 29 |
| EIF4E | 1:1 | 195.3 | 195.3 | 25000 | 100000 | 2.1 | 11 | 37 |
| CD226 | 1:1 | | | | | | 15 | 21 |

| | | | | | | | | |
|---|---|---|---|---|---|---|---|---|
| VAMP8 | 1:1 | 1562.5 | 3125.0 | 400000 | 3200000 | 2.1 | 12 | 30 |
| VEGFB | 1:1 | | | | | | 9 | 12 |
| CSF2RB | 1:1 | 97.7 | 97.7 | 12500 | 100000 | 2.1 | 9 | 22 |
| CLEC12A | 1:1 | | | | | | 8 | 9 |
| IL36G | 1:1 | | | | | | 16 | 16 |
| DBN1 | 1:1 | 24.4 | 48.8 | 3125 | 800000 | 1.8 | 9 | 17 |
| ZBP1 | 1:1 | 6250.0 | 12500.0 | 800000 | 800000 | 1.8 | 9 | 10 |
| CD7 | 1:1 | 781.3 | 781.3 | 200000 | 800000 | 2.4 | 15 | 11 |
| BNIP3L | 1:1 | 195.3 | 390.6 | 12500 | 100000 | 1.5 | 15 | 18 |
| ACP1 | 1:1 | 6250.0 | 6250.0 | 400000 | 800000 | 1.8 | | |
| NEDD4L | 1:1 | | | | | | 8 | 21 |
| NFAT5 | 1:1 | | | | | | 12 | 18 |
| LATS1 | 1:1 | | | | | | 14 | 38 |
| AFAP1 | 1:10 | | | | | | 6 | 23 |
| ITGA2 | 1:10 | | | | | | 8 | 14 |
| APPL2 | 1:10 | 195.3 | 390.6 | 50000 | 800000 | 2.1 | | |
| TNFRSF17 | 1:10 | | | | | | 5 | 12 |
| NHLRC3 | 1:10 | | | | | | 5 | 7 |
| ADGRD1 | 1:10 | 48.8 | 97.7 | 12500 | 50000 | 2.1 | 5 | 10 |
| BMPER | 1:10 | 24.4 | 48.8 | 25000 | 100000 | 2.7 | 5 | 11 |
| COL5A1 | 1:10 | 12.2 | 12.2 | 6250 | 50000 | 2.7 | 6 | 11 |
| JAM3 | 1:10 | 48.8 | 97.7 | 12500 | 100000 | 2.1 | 14 | 33 |
| ACHE | 1:10 | 24.4 | 48.8 | 12500 | 100000 | 2.4 | 6 | 16 |
| PAXX | 1:10 | 97.7 | 195.3 | 25000 | 100000 | 2.1 | 5 | 9 |
| PDE5A | 1:10 | 1562.5 | 3125.0 | 200000 | 800000 | 1.8 | 5 | 27 |
| MXRA8 | 1:10 | 195.3 | 390.6 | 25000 | 200000 | 1.8 | 5 | 14 |
| DIPK2B | 1:10 | | | | | | 6 | 10 |
| CST1 | 1:10 | 390.6 | 781.3 | 50000 | 400000 | 1.8 | 6 | 32 |
| ANXA1 | 1:10 | | | | | | 7 | 16 |
| DTD1 | 1:10 | 781.3 | 781.3 | 50000 | 800000 | 1.8 | | |
| HEG1 | 1:10 | 390.6 | 781.3 | 50000 | 400000 | 1.8 | 5 | 9 |
| ERP29 | 1:10 | 97.7 | 97.7 | 6250 | 12500 | 1.8 | | |
| SCGB3A1 | 1:10 | | | | | | 6 | 13 |
| TBCA | 1:10 | 24.4 | 48.8 | 6250 | 50000 | 2.1 | 5 | 27 |
| CD300A | 1:10 | 48.8 | 97.7 | 6250 | 50000 | 1.8 | 5 | 9 |
| HMCN2 | 1:10 | 390.6 | 781.3 | 100000 | 400000 | 2.1 | 5 | 21 |
| GLA | 1:10 | | | | | | 5 | 11 |
| LEG1 | 1:10 | 12.2 | 48.8 | 6250 | 25000 | 2.1 | 5 | 18 |
| PRR4 | 1:10 | | | | | | 7 | 18 |
| PCBD1 | 1:10 | | | | | | 5 | 12 |
| CPA4 | 1:10 | | | | | | 6 | 13 |
| YWHAQ | 1:10 | 390.6 | 390.6 | 25000 | 200000 | 1.8 | 6 | 29 |
| SERPINI1 | 1:10 | 195.3 | 195.3 | 6250 | 50000 | 1.5 | 8 | 12 |
| UROD | 1:10 | 390.6 | 390.6 | 25000 | 200000 | 1.8 | 6 | 16 |
| DDI2 | 1:10 | 195.3 | 390.6 | 50000 | 100000 | 2.1 | 6 | 17 |
| CELSR2 | 1:10 | | | | | | 5 | 8 |
| GP5 | 1:10 | 390.6 | 781.3 | 50000 | 200000 | 1.8 | 5 | 16 |
| GAPDH | 1:10 | | | | | | 6 | 21 |
| SEMA3G | 1:10 | | | | | | 6 | 13 |
| ADH1B | 1:10 | 3125.0 | 6250.0 | 400000 | 800000 | 1.8 | 9 | 19 |
| MFAP4 | 1:10 | 390.6 | 781.3 | 50000 | 200000 | 1.8 | 5 | 6 |
| GNPDA2 | 1:10 | 390.6 | 390.6 | 12500 | 50000 | 1.5 | 6 | 15 |

| | | | | | | | | |
|---|---|---|---|---|---|---|---|---|
| PSTPIP2 | 1:10 | 1562.5 | 3125.0 | 200000 | 800000 | 1.8 | 10 | 37 |
| MENT | 1:10 | | | | | | 5 | 7 |
| DNAJB6 | 1:10 | 781.3 | 3125.0 | 200000 | 800000 | 1.8 | 6 | 27 |
| RLN2 | 1:10 | 781.3 | 3125.0 | 800000 | 800000 | 2.4 | 20 | 24 |
| BMP10 | 1:10 | 781.3 | 781.3 | 50000 | 100000 | 1.8 | 7 | 11 |
| TPD52L2 | 1:10 | 781.3 | 781.3 | 50000 | 800000 | 1.8 | 6 | 37 |
| CRELD1 | 1:10 | 195.3 | 195.3 | 25000 | 200000 | 2.1 | 5 | 11 |
| AKAP12 | 1:10 | 781.3 | 1562.5 | 100000 | 1600000 | 1.8 | 8 | 10 |
| AMOT | 1:10 | 12.2 | 24.4 | 3125 | 12500 | 2.1 | 7 | 20 |
| GHR | 1:10 | | | | | | 5 | 10 |
| C1QTNF5 | 1:10 | 1562.5 | 3125.0 | 200000 | 1600000 | 1.8 | 5 | 9 |
| PDIA3 | 1:10 | | | | | | 11 | 18 |
| C1QTNF9 | 1:10 | 195.3 | 195.3 | 12500 | 400000 | 1.8 | 4 | 18 |
| GCHFR | 1:10 | | | | | | 5 | 8 |
| RABEP1 | 1:10 | 48.8 | 195.3 | 25000 | 100000 | 2.1 | 7 | 14 |
| SLC9A3R1 | 1:10 | | | | | | 14 | 33 |
| SFRP4 | 1:10 | | | | | | 5 | 10 |
| SDK2 | 1:10 | 195.3 | 390.6 | 25000 | 100000 | 1.8 | 10 | 15 |
| CACYBP | 1:10 | 390.6 | 781.3 | 50000 | 200000 | 1.8 | 6 | 35 |
| CTSE | 1:10 | 97.7 | 97.7 | 50000 | 200000 | 2.7 | 6 | 13 |
| SUSD5 | 1:10 | 195.3 | 390.6 | 25000 | 100000 | 1.8 | 4 | 16 |
| SPINK2 | 1:10 | 48.8 | 48.8 | 6250 | 25000 | 2.1 | 5 | 10 |
| PINLYP | 1:10 | 390.6 | 781.3 | 50000 | 200000 | 1.8 | 11 | 16 |
| FUOM | 1:10 | 48.8 | 195.3 | 12500 | 100000 | 1.8 | 5 | 22 |
| KLK7 | 1:10 | | | | | | 6 | 14 |
| INHBB | 1:10 | 390.6 | 390.6 | 25000 | 100000 | 1.8 | 7 | 18 |
| DNAJB2 | 1:10 | 195.3 | 390.6 | 25000 | 100000 | 1.8 | 6 | 11 |
| GLRX5 | 1:10 | 48.8 | 97.7 | 6250 | 50000 | 1.8 | | |
| FGFR4 | 1:10 | 390.6 | 781.3 | 50000 | 100000 | 1.8 | 5 | 13 |
| TP53I3 | 1:10 | 6.1 | 24.4 | 6250 | 50000 | 2.4 | 5 | 10 |
| MOCS2 | 1:10 | 1562.5 | 3125.0 | 200000 | 800000 | 1.8 | 7 | 11 |
| GIMAP7 | 1:10 | 12.2 | 48.8 | 12500 | 100000 | 2.4 | 7 | 11 |
| LZTFL1 | 1:10 | 24.4 | 97.7 | 50000 | 200000 | 2.7 | 9 | 24 |
| VTI1A | 1:10 | 195.3 | 195.3 | 12500 | 25000 | 1.8 | 9 | 13 |
| OLFM4 | 1:10 | 781.3 | 1562.5 | 400000 | 800000 | 2.4 | 6 | 34 |
| SYAP1 | 1:10 | | | | | | 9 | 24 |
| PENK | 1:10 | 3125.0 | 3125.0 | 200000 | 400000 | 1.8 | 6 | 8 |
| ARHGAP45 | 1:10 | 1562.5 | 1562.5 | 400000 | 800000 | 2.4 | 9 | 16 |
| CD72 | 1:10 | 97.7 | 195.3 | 12500 | 100000 | 1.8 | 6 | 14 |
| RIDA | 1:10 | 48.8 | 97.7 | 3125 | 12500 | 1.5 | 5 | 10 |
| DAAM1 | 1:10 | 6250.0 | 6250.0 | 400000 | 800000 | 1.8 | 9 | 27 |
| NAGPA | 1:10 | 12.2 | 48.8 | 6250 | 50000 | 2.1 | 5 | 9 |
| PHYKPL | 1:10 | 3125.0 | 3125.0 | 400000 | 800000 | 2.1 | 8 | 14 |
| SNX15 | 1:10 | 195.3 | 195.3 | 12500 | 100000 | 1.8 | 11 | 16 |
| MYOM3 | 1:10 | 12.2 | 24.4 | 6250 | 50000 | 2.4 | 5 | 36 |
| SNCA | 1:10 | | | | | | | |
| ACYP1 | 1:10 | 12.2 | 48.8 | 3125 | 12500 | 1.8 | 5 | 15 |
| UBXN1 | 1:10 | | | | | | 10 | 21 |
| TNFAIP8L2 | 1:10 | | | | | | 8 | 22 |
| DCTD | 1:10 | 195.3 | 390.6 | 25000 | 100000 | 1.8 | 6 | 24 |
| MARS1 | 1:10 | 1562.5 | 3125.0 | 400000 | 800000 | 2.1 | 6 | 16 |
| CHAD | 1:10 | 195.3 | 390.6 | 25000 | 100000 | 1.8 | 5 | 18 |

| | | | | | | | | |
|---|---|---|---|---|---|---|---|---|
| GMPR2 | 1:10 | 97.7 | 195.3 | 25000 | 100000 | 2.1 | 6 | 15 |
| ADD1 | 1:10 | | | | | | 10 | 21 |
| EPHA4 | 1:10 | | | | | | 6 | 9 |
| PRKG1 | 1:10 | | | | | | 5 | 31 |
| SPART | 1:10 | 97.7 | 390.6 | 50000 | 400000 | 2.1 | 6 | 16 |
| PTRHD1 | 1:10 | 390.6 | 390.6 | 50000 | 800000 | 2.1 | 7 | 26 |
| ST13 | 1:10 | 390.6 | 390.6 | 50000 | 800000 | 2.1 | 5 | 9 |
| PRDX2 | 1:1000 | | | | | | 8 | 12 |
| CSH1 | 1:1000 | 97.7 | 97.7 | 12500 | 50000 | 2.1 | | |
| MDH1 | 1:1000 | 1562.5 | 6250.0 | 400000 | 800000 | 1.8 | 4 | 8 |
| TXN | 1:1000 | 48.8 | 97.7 | 6250 | 12500 | 1.8 | 24 | 23 |
| NRGN | 1:1000 | 48.8 | 48.8 | 12500 | 800000 | 2.4 | 7 | 28 |
| FGL1 | 1:1000 | 390.6 | 390.6 | 100000 | 200000 | 2.4 | 4 | 27 |
| APOF | 1:1000 | | | | | | 6 | 20 |
| CSF1R | 1:1000 | 97.7 | 195.3 | 12500 | 50000 | 1.8 | 5 | 9 |
| GSR | 1:1000 | 195.3 | 390.6 | 25000 | 100000 | 1.8 | 5 | 6 |
| PGA4 | 1:1000 | 12.2 | 24.4 | 25000 | 100000 | 3.0 | 4 | 9 |
| C9 | 1:1000 | | | | | | 19 | 28 |
| CR1 | 1:1000 | 6.1 | 6.1 | 6250 | 50000 | 3.0 | 6 | 9 |
| ACE | 1:1000 | 390.6 | 1562.5 | 200000 | 800000 | 2.1 | 7 | 9 |
| PI16 | 1:1000 | | | | | | 6 | 6 |
| MRC1 | 1:1000 | 24.4 | 48.8 | 12500 | 200000 | 2.4 | 5 | 9 |
| PNLIP | 1:1000 | 6.1 | 12.2 | 12500 | 50000 | 3.0 | 5 | 13 |
| VASP | 1:1000 | 6250.0 | 12500.0 | 800000 | 800000 | 1.8 | 9 | 36 |
| LCAT | 1:1000 | 390.6 | 781.3 | 50000 | 400000 | 1.8 | 5 | 9 |
| TGOLN2 | 1:1000 | 97.7 | 195.3 | 12500 | 800000 | 1.8 | 7 | 10 |
| FCN1 | 1:1000 | | | | | | 11 | 22 |
| RNASE6 | 1:1000 | 6.1 | 24.4 | 3125 | 12500 | 2.1 | 6 | 7 |
| PEPD | 1:1000 | 195.3 | 390.6 | 25000 | 200000 | 1.8 | 6 | 9 |
| NEXN | 1:1000 | 48.8 | 97.7 | 25000 | 200000 | 2.4 | | |
| TCN1 | 1:1000 | 12.2 | 24.4 | 6250 | 50000 | 2.4 | 5 | 10 |
| APOA2 | 1:1000 | | | | | | | |
| SHBG | 1:1000 | 1562.5 | 3125.0 | 400000 | 800000 | 2.1 | 3 | 10 |
| F10 | 1:1000 | 97.7 | 195.3 | 12500 | 50000 | 1.8 | 8 | 9 |
| ASGR2 | 1:1000 | 195.3 | 390.6 | 25000 | 200000 | 1.8 | 8 | 14 |
| SOD3 | 1:1000 | 3.1 | 12.2 | 1563 | 12500 | 2.1 | 4 | 11 |
| TREML1 | 1:1000 | 97.7 | 390.6 | 25000 | 200000 | 1.8 | 8 | 29 |
| DBH | 1:1000 | 97.7 | 195.3 | 100000 | 400000 | 2.7 | 4 | 32 |
| PON1 | 1:1000 | | | | | | 7 | 11 |
| ABO | 1:1000 | 12.2 | 48.8 | 50000 | 200000 | 3.0 | 8 | 9 |
| VNN1 | 1:1000 | 24.4 | 48.8 | 6250 | 50000 | 2.1 | 4 | 20 |
| HGFAC | 1:1000 | 48.8 | 195.3 | 12500 | 50000 | 1.8 | 4 | 7 |
| BCHE | 1:1000 | | | | | | 4 | 12 |
| CFHR5 | 1:1000 | | | | | | 6 | 15 |
| CRISP3 | 1:1000 | 390.6 | 390.6 | 25000 | 50000 | 1.8 | 3 | 7 |
| GSN | 1:1000 | 1562.5 | 3125.0 | 200000 | 800000 | 1.8 | 4 | 8 |
| CGB3 | 1:1000 | | | | | | 13 | |
| MST1 | 1:1000 | 97.7 | 195.3 | 25000 | 200000 | 2.1 | 4 | 14 |
| CAT | 1:1000 | | | | | | 7 | 11 |
| RNASE4 | 1:1000 | 781.3 | 781.3 | 50000 | 200000 | 1.8 | 6 | 9 |
| RNASE1 | 1:1000 | | | | | | 10 | 11 |
| CTBS | 1:1000 | 195.3 | 195.3 | 12500 | 50000 | 1.8 | 4 | 5 |

| | | | | | | | | |
|---|---|---|---|---|---|---|---|---|
| GPI | 1:1000 | 195.3 | 390.6 | 50000 | 200000 | 2.1 | 7 | 16 |
| APOB | 1:1000 | | | | | | 9 | 29 |
| AGT | 1:1000 | 1562.5 | 3125.0 | 200000 | 800000 | 1.8 | 6 | 31 |
| AMY1A | 1:1000 | 195.3 | 195.3 | 12500 | 50000 | 1.8 | 6 | 12 |
| AHSG | 1:1000 | | | | | | 5 | 14 |
| APOL1 | 1:1000 | 97.7 | 97.7 | 25000 | 100000 | 2.4 | 4 | 33 |
| C8B | 1:100000 | | | | | | 12 | 14 |
| CFHR2 | 1:100000 | 195.3 | 390.6 | 25000 | 100000 | 1.8 | 8 | 16 |
| SERPINA6 | 1:100000 | 390.6 | 781.3 | 200000 | 800000 | 2.4 | 5 | 8 |
| SERPINA7 | 1:100000 | 195.3 | 390.6 | 25000 | 200000 | 1.8 | 7 | 8 |
| SERPINF1 | 1:100000 | 195.3 | 390.6 | 50000 | 800000 | 2.1 | 3 | 8 |
| PGLYRP2 | 1:100000 | 1562.5 | 3125.0 | 400000 | 800000 | 2.1 | 4 | 6 |
| LGALS3BP | 1:100000 | 97.7 | 195.3 | 12500 | 50000 | 1.8 | 7 | 11 |
| CLU | 1:100000 | 781.3 | 1562.5 | 100000 | 800000 | 1.8 | 5 | 8 |
| B2M | 1:100000 | 48.8 | 97.7 | 6250 | 25000 | 1.8 | 6 | 5 |
| AFM | 1:100000 | 97.7 | 195.3 | 50000 | 200000 | 2.4 | 6 | 7 |
| CFHR4 | 1:100000 | 48.8 | 97.7 | 12500 | 50000 | 2.1 | 4 | 12 |
| SAA4 | 1:100000 | | | | | | 5 | 9 |
| IGLC2 | 1:100000 | 6.1 | 12.2 | 781 | 50000 | 1.8 | 4 | 5 |
| F12 | 1:100000 | 97.7 | 195.3 | 25000 | 200000 | 2.1 | 9 | 23 |
| MBL2 | 1:100000 | 1.5 | 12.2 | 3125 | 25000 | 2.4 | 5 | 27 |
| F2 | 1:100000 | | | | | | 6 | 7 |
| APOE | 1:100000 | | | | | | 10 | 17 |
| LYVE1 | 1:100000 | 3.1 | 6.1 | 1563 | 12500 | 2.4 | 6 | 11 |
| CFD | 1:100000 | 12.2 | 24.4 | 12500 | 50000 | 2.7 | 7 | 6 |
| PROS1 | 1:100000 | 195.3 | 390.6 | 50000 | 200000 | 2.1 | 5 | 7 |
| KLKB1 | 1:100000 | 195.3 | 390.6 | 25000 | 200000 | 1.8 | 5 | 7 |
| ECM1 | 1:100000 | 6250.0 | 6250.0 | 400000 | 800000 | 1.8 | 7 | 15 |
| ATRN | 1:100000 | 195.3 | 390.6 | 50000 | 200000 | 2.1 | 5 | 6 |
| SERPINA5 | 1:100000 | | | | | | 5 | 14 |
| QSOX1 | 1:100000 | 390.6 | 390.6 | 12500 | 50000 | 1.5 | 6 | 7 |
| PPBP | 1:100000 | 24.4 | 48.8 | 12500 | 100000 | 2.4 | 5 | 28 |
| CPB2 | 1:100000 | 195.3 | 390.6 | 25000 | 200000 | 1.8 | 6 | 9 |
| PZP | 1:100000 | 390.6 | 390.6 | 100000 | 800000 | 2.4 | 4 | 6 |
| APOA4 | 1:100000 | 781.3 | 1562.5 | 100000 | 800000 | 1.8 | 8 | 16 |
| C1S | 1:100000 | 195.3 | 390.6 | 50000 | 200000 | 2.1 | 6 | 7 |
| PF4 | 1:100000 | | | | | | 11 | 28 |
| CLEC3B | 1:100000 | 97.7 | 195.3 | 25000 | 200000 | 2.1 | 5 | 6 |
| ITIH4 | 1:100000 | 390.6 | 781.3 | 50000 | 200000 | 1.8 | 4 | 14 |
| SERPIND1 | 1:100000 | 3125.0 | 3125.0 | 400000 | 800000 | 2.1 | 5 | 11 |
| APOA1 | 1:100000 | | | | | | 8 | 14 |
| ADIPOQ | 1:100000 | 781.3 | 781.3 | 25000 | 50000 | 1.5 | 7 | 18 |
| C7 | 1:100000 | 24.4 | 48.8 | 12500 | 100000 | 2.4 | 9 | 6 |
| LPA | 1:100000 | 24.4 | 48.8 | 3125 | 50000 | 1.8 | | |
| C1R | 1:100000 | 390.6 | 1562.5 | 100000 | 200000 | 1.8 | 5 | 6 |
| APOD | 1:100000 | 781.3 | 1562.5 | 50000 | 200000 | 1.5 | 7 | 17 |
| SELL | 1:100000 | 3.1 | 6.1 | 1563 | 6250 | 2.4 | 4 | 6 |
| BTD | 1:100000 | 12.2 | 24.4 | 3125 | 25000 | 2.1 | 5 | 7 |
| C1RL | 1:100000 | 12.2 | 24.4 | 6250 | 50000 | 2.4 | 5 | 7 |
| ORM1 | 1:100000 | 24.4 | 48.8 | 6250 | 25000 | 2.1 | 6 | 5 |
| ITIH1 | 1:100000 | 195.3 | 195.3 | 25000 | 200000 | 2.1 | 6 | 8 |
| APOC1 | 1:100000 | 195.3 | 390.6 | 100000 | 800000 | 2.4 | 9 | 16 |

| | | | | | | | | |
|---|---|---|---|---|---|---|---|---|
| CFB | 1:100000 | 781.3 | 1562.5 | 100000 | 800000 | 1.8 | 6 | 12 |
| SERPINA1 | 1:100000 | 3125.0 | 6250.0 | 400000 | 800000 | 1.8 | 3 | 4 |
| SELENOP | 1:100000 | 97.7 | 195.3 | 25000 | 100000 | 2.1 | 15 | 19 |
| C5 | 1:100000 | 24.4 | 48.8 | 3125 | 12500 | 1.8 | 7 | 7 |
| LRG1 | 1:100000 | 97.7 | 97.7 | 6250 | 25000 | 1.8 | 5 | 6 |
| HRG | 1:100000 | 97.7 | 195.3 | 25000 | 200000 | 2.1 | 6 | 9 |
| PLG | 1:100000 | 97.7 | 195.3 | 50000 | 200000 | 2.4 | 4 | 11 |
| CFP | 1:100000 | 195.3 | 390.6 | 25000 | 200000 | 1.8 | 4 | 7 |
| TF | 1:100000 | | | | | | 5 | 5 |
| FN1 | 1:100000 | | | | | | 4 | 23 |
| TTR | 1:100000 | 195.3 | 195.3 | 12500 | 50000 | 1.8 | 5 | 5 |
| F13B | 1:100000 | 390.6 | 781.3 | 25000 | 100000 | 1.5 | 6 | 10 |
| GC | 1:100000 | 97.7 | 390.6 | 25000 | 200000 | 1.8 | 4 | 7 |
| SERPINF2 | 1:100000 | 390.6 | 781.3 | 100000 | 400000 | 2.1 | 4 | 6 |
| FGA | 1:100000 | 781.3 | 1562.5 | 200000 | 800000 | 2.1 | 6 | 19 |
| SERPING1 | 1:100000 | 390.6 | 390.6 | 25000 | 200000 | 1.8 | 4 | 7 |
| SERPINC1 | 1:100000 | | | | | | 4 | 4 |
| C3 | 1:100000 | | | | | | 7 | 40 |
| CD5L | 1:100000 | 24.4 | 24.4 | 12500 | 50000 | 2.7 | 4 | 13 |
| SERPINA4 | 1:100000 | | | | | | 5 | 6 |
| APCS | 1:100000 | 195.3 | 390.6 | 25000 | 100000 | 1.8 | 4 | 10 |
| SERPINA3 | 1:100000 | | | | | | 3 | 6 |
| A1BG | 1:100000 | 48.8 | 195.3 | 12500 | 200000 | 1.8 | 4 | 6 |
| CFH | 1:100000 | | | | | | 5 | 8 |
| F11 | 1:100000 | 12.2 | 48.8 | 12500 | 50000 | 2.4 | 6 | 9 |
| CFI | 1:100000 | 195.3 | 390.6 | 50000 | 200000 | 2.1 | 5 | 9 |
| ODAM | 1:1 | 195.3 | 781.3 | 100000 | 200000 | 2.1 | 8 | 14 |
| NPM1 | 1:1 | 24.4 | 48.8 | 12500 | 200000 | 2.4 | 8 | 10 |
| PRTFDC1 | 1:1 | 3125.0 | 6250.0 | 400000 | 800000 | 1.8 | 10 | 11 |
| NXPH1 | 1:1 | 48.8 | 195.3 | 100000 | 200000 | 2.7 | 8 | 13 |
| MUC13 | 1:1 | 6.1 | 12.2 | 6250 | 25000 | 2.7 | 9 | 19 |
| VSTM1 | 1:1 | 48.8 | 97.7 | 6250 | 25000 | 1.8 | 9 | 9 |
| SLC39A14 | 1:1 | 97.7 | 390.6 | 12500 | 50000 | 1.5 | 9 | 19 |
| TBC1D17 | 1:1 | 195.3 | 390.6 | 50000 | 200000 | 2.1 | 8 | 12 |
| ADGRB3 | 1:1 | 12.2 | 24.4 | 100000 | 800000 | 3.6 | 7 | 6 |
| PSME1 | 1:1 | 97.7 | 195.3 | 50000 | 200000 | 2.4 | 8 | 8 |
| FRZB | 1:1 | 97.7 | 195.3 | 25000 | 50000 | 2.1 | 7 | 5 |
| GLB1 | 1:1 | 195.3 | 390.6 | 100000 | 800000 | 2.4 | 8 | 10 |
| GLT8D2 | 1:1 | 3125.0 | 6250.0 | 1600000 | 3200000 | 2.4 | 9 | 9 |
| ASAH2 | 1:1 | 97.7 | 195.3 | 25000 | 200000 | 2.1 | 11 | 6 |
| AGR2 | 1:1 | 1562.5 | 1562.5 | 100000 | 800000 | 1.8 | 21 | 13 |
| GFRA3 | 1:1 | 24.4 | 48.8 | 12500 | 50000 | 2.4 | 9 | 8 |
| CHMP1A | 1:1 | | | | | | 5 | 10 |
| TST | 1:1 | 6250.0 | 6250.0 | 800000 | 1600000 | 2.1 | 11 | 13 |
| MAX | 1:1 | 97.7 | 390.6 | 25000 | 100000 | 1.8 | 7 | 19 |
| GSTP1 | 1:1 | 12500.0 | 12500.0 | 400000 | 1600000 | 1.5 | 8 | 17 |
| ISLR2 | 1:1 | 24.4 | 48.8 | 25000 | 200000 | 2.7 | 6 | 15 |
| CST5 | 1:1 | 24.4 | 24.4 | 6250 | 25000 | 2.4 | 6 | 6 |
| CX3CL1 | 1:1 | 24.4 | 48.8 | 6250 | 12500 | 2.1 | 8 | 8 |
| CASP1 | 1:1 | 97.7 | 195.3 | 50000 | 100000 | 2.4 | 8 | 21 |
| NEFL | 1:1 | 390.6 | 781.3 | 200000 | 800000 | 2.4 | 12 | 12 |
| TDRKH | 1:1 | 6.1 | 12.2 | 6250 | 25000 | 2.7 | 6 | 13 |

| | | | | | | | | |
|---|---|---|---|---|---|---|---|---|
| CLSTN1 | 1:1 | 390.6 | 1562.5 | 200000 | 800000 | 2.1 | 10 | 11 |
| KIRREL2 | 1:1 | 97.7 | 195.3 | 50000 | 200000 | 2.4 | 8 | 14 |
| PFDN2 | 1:1 | 97.7 | 195.3 | 100000 | 800000 | 2.7 | 7 | 23 |
| HNMT | 1:1 | 12.2 | 48.8 | 25000 | 50000 | 2.7 | 7 | 6 |
| TMPRSS5 | 1:1 | 3.1 | 6.1 | 6250 | 50000 | 3.0 | 8 | 5 |
| TNFRSF10B | 1:1 | 0.8 | 1.5 | 3125 | 12500 | 3.3 | 10 | 7 |
| PLA2G10 | 1:1 | 1.5 | 3.1 | 3125 | 12500 | 3.0 | 9 | 6 |
| LY96 | 1:1 | 6250.0 | 12500.0 | 1600000 | 3200000 | 2.1 | 9 | 6 |
| EBAG9 | 1:1 | 97.7 | 390.6 | 50000 | 200000 | 2.1 | 8 | 6 |
| PSG1 | 1:1 | 6.1 | 12.2 | 3125 | 12500 | 2.4 | 8 | 7 |
| CSF2RA | 1:1 | 781.3 | 1562.5 | 400000 | 800000 | 2.4 | 9 | 8 |
| ECE1 | 1:1 | | | | | | 13 | 17 |
| WFIKKN1 | 1:1 | 12.2 | 24.4 | 50000 | 200000 | 3.3 | 10 | 5 |
| GPC5 | 1:1 | 12.2 | 48.8 | 25000 | 50000 | 2.7 | 10 | 6 |
| CRADD | 1:1 | | | | | | 10 | 6 |
| CCS | 1:1 | 781.3 | 1562.5 | 400000 | 800000 | 2.4 | 9 | 10 |
| ASGR1 | 1:1 | 195.3 | 390.6 | 100000 | 800000 | 2.4 | 7 | 6 |
| CCL2 | 1:1 | 6.1 | 12.2 | 1563 | 6250 | 2.1 | 9 | 8 |
| KRT14 | 1:1 | 97.7 | 390.6 | 200000 | 800000 | 2.7 | 6 | 18 |
| RAB6B | 1:1 | 195.3 | 781.3 | 50000 | 200000 | 1.8 | 8 | 11 |
| SIGLEC15 | 1:1 | 97.7 | 195.3 | 100000 | 200000 | 2.7 | 6 | 15 |
| METAP1 | 1:1 | 781.3 | 1562.5 | 200000 | 800000 | 2.1 | 7 | 23 |
| IL34 | 1:1 | 48.8 | 195.3 | 200000 | 800000 | 3.0 | 8 | 14 |
| IFNL1 | 1:1 | 6.1 | 12.2 | 25000 | 100000 | 3.3 | 9 | 16 |
| LYPD1 | 1:1 | 195.3 | 390.6 | 100000 | 400000 | 2.4 | 6 | 12 |
| RASA1 | 1:1 | 6250.0 | 6250.0 | 3200000 | 6400000 | 2.7 | 9 | 15 |
| ATP5PO | 1:1 | 48.8 | 48.8 | 100000 | 800000 | 3.3 | 6 | 20 |
| IDI2 | 1:1 | 195.3 | 781.3 | 200000 | 800000 | 2.4 | 12 | 20 |
| ARID4B | 1:1 | 12.2 | 24.4 | 12500 | 200000 | 2.7 | 6 | 15 |
| SSB | 1:1 | 390.6 | 1562.5 | 200000 | 800000 | 2.1 | 6 | 10 |
| SLC27A4 | 1:1 | 24.4 | 24.4 | 100000 | 200000 | 3.6 | 8 | 18 |
| SIRT5 | 1:1 | 1562.5 | 1562.5 | 200000 | 400000 | 2.1 | 12 | 24 |
| KCNIP4 | 1:1 | 48.8 | 48.8 | 12500 | 200000 | 2.4 | 8 | 8 |
| SLC16A1 | 1:1 | 781.3 | 1562.5 | 400000 | 800000 | 2.4 | 13 | 9 |
| SESTD1 | 1:1 | 3125.0 | 3125.0 | 400000 | 800000 | 2.1 | 14 | 6 |
| PXN | 1:1 | | | | | | 7 | 14 |
| SMARCA2 | 1:1 | 97.7 | 195.3 | 50000 | 200000 | 2.4 | 7 | 12 |
| ANXA5 | 1:1 | | | | | | 10 | 17 |
| LIF | 1:1 | 3.1 | 6.1 | 3125 | 100000 | 2.7 | 6 | 9 |
| TDGF1 | 1:1 | 390.6 | 781.3 | 100000 | 200000 | 2.1 | 8 | 34 |
| ATF2 | 1:1 | 1406.3 | 2812.5 | 360000 | 720000 | 2.1 | 8 | 16 |
| PAK4 | 1:1 | 390.6 | 1562.5 | 200000 | 800000 | 2.1 | 12 | 15 |
| CDHR1 | 1:1 | 48.8 | 97.7 | 50000 | 200000 | 2.7 | 8 | 10 |
| MMP13 | 1:1 | 24.4 | 48.8 | 100000 | 400000 | 3.3 | 8 | 13 |
| MAPT | 1:1 | 1562.5 | 1562.5 | 50000 | 200000 | 1.5 | 11 | 22 |
| COPE | 1:1 | 781.3 | 1562.5 | 400000 | 800000 | 2.4 | 8 | 12 |
| FGR | 1:1 | 390.6 | 390.6 | 100000 | 400000 | 2.4 | 8 | 19 |
| FOSB | 1:1 | 195.3 | 390.6 | 200000 | 800000 | 2.7 | 6 | 22 |
| CLSPN | 1:1 | | | | | | 17 | 32 |
| FMNL1 | 1:1 | 3125.0 | 3125.0 | 200000 | 800000 | 1.8 | 11 | 19 |
| S100A16 | 1:1 | 97.7 | 390.6 | 25000 | 400000 | 1.8 | 13 | 13 |
| DCTN6 | 1:1 | 781.3 | 3125.0 | 200000 | 800000 | 1.8 | 5 | 14 |

| | | | | | | | | |
|---|---|---|---|---|---|---|---|---|
| SCARB1 | 1:1 | 1562.5 | 3125.0 | 400000 | 800000 | 2.1 | 7 | 14 |
| CALB2 | 1:1 | 195.3 | 781.3 | 200000 | 400000 | 2.4 | 9 | 19 |
| ATXN10 | 1:1 | 195.3 | 390.6 | 50000 | 200000 | 2.1 | 9 | 13 |
| MRPL46 | 1:1 | | | | | | 11 | 21 |
| BAX | 1:1 | 14.2 | 28.3 | 29000 | 116000 | 3.0 | 9 | 21 |
| ATP6V1F | 1:1 | | | | | | 7 | 16 |
| TPPP3 | 1:1 | 195.3 | 390.6 | 25000 | 100000 | 1.8 | 8 | 10 |
| KEL | 1:1 | 1562.5 | 3125.0 | 200000 | 800000 | 1.8 | 6 | 8 |
| NDRG1 | 1:1 | 48.8 | 195.3 | 100000 | 400000 | 2.7 | 9 | 15 |
| IVD | 1:1 | 3125.0 | 3125.0 | 400000 | 800000 | 2.1 | 10 | 12 |
| PADI4 | 1:1 | | | | | | 9 | 14 |
| GGA1 | 1:1 | | | | | | 11 | 11 |
| HSP90B1 | 1:1 | | | | | | 7 | 11 |
| EPHA10 | 1:1 | 97.7 | 195.3 | 100000 | 800000 | 2.7 | 8 | 15 |
| BRK1 | 1:1 | 24.4 | 48.8 | 100000 | 200000 | 3.3 | 9 | 13 |
| PMVK | 1:1 | 3125.0 | 3125.0 | 800000 | 6400000 | 2.4 | 17 | 21 |
| HMOX2 | 1:1 | 24.4 | 48.8 | 100000 | 200000 | 3.3 | 8 | 10 |
| KLB | 1:1 | 1562.5 | 1562.5 | 200000 | 800000 | 2.1 | 12 | 22 |
| ILKAP | 1:1 | 781.3 | 1562.5 | 400000 | 800000 | 2.4 | 10 | 17 |
| C2CD2L | 1:1 | 25000.0 | 50000.0 | 6400000 | 12800000 | 2.1 | 5 | |
| SLIT2 | 1:1 | 97.7 | 390.6 | 100000 | 200000 | 2.4 | 7 | 11 |
| KRT5 | 1:1 | | | | | | 11 | 17 |
| GKN1 | 1:1 | 1562.5 | 3125.0 | 800000 | 3200000 | 2.4 | 6 | 26 |
| PTK7 | 1:1 | 390.6 | 390.6 | 100000 | 200000 | 2.4 | 7 | 11 |
| AMFR | 1:1 | 97.7 | 195.3 | 100000 | 400000 | 2.7 | 7 | 21 |
| C19orf12 | 1:1 | 48.8 | 195.3 | 100000 | 200000 | 2.7 | 7 | 18 |
| NOS3 | 1:1 | 24.4 | 48.8 | 100000 | 200000 | 3.3 | 10 | 14 |
| PTEN | 1:1 | | | | | | 10 | 11 |
| TARBP2 | 1:1 | 97.7 | 195.3 | 100000 | 200000 | 2.7 | 7 | 19 |
| CERT1 | 1:1 | | | | | | 10 | 13 |
| WASF3 | 1:1 | | | | | | 6 | 9 |
| TNF | 1:1 | 1.5 | 6.1 | 6250 | 200000 | 3.0 | 10 | 16 |
| NOS1 | 1:1 | 6.1 | 6.1 | 25000 | 50000 | 3.6 | 11 | 21 |
| DUSP3 | 1:1 | | | | | | 9 | 10 |
| CETN2 | 1:1 | 3125.0 | 3125.0 | 200000 | 800000 | 1.8 | 11 | 20 |
| PLIN1 | 1:1 | | | | | | 6 | 9 |
| STK24 | 1:1 | 24.4 | 195.3 | 50000 | 200000 | 2.4 | 5 | 18 |
| PBLD | 1:1 | 390.6 | 781.3 | 400000 | 800000 | 2.7 | 7 | 12 |
| PTS | 1:1 | 24.4 | 48.8 | 25000 | 200000 | 2.7 | 8 | 11 |
| XRCC4 | 1:1 | 390.6 | 781.3 | 100000 | 200000 | 2.1 | 7 | 11 |
| GDNF | 1:1 | 0.4 | 0.8 | 1563 | 12500 | 3.3 | 7 | 22 |
| ING1 | 1:1 | 48.8 | 97.7 | 100000 | 200000 | 3.0 | 8 | 15 |
| GHRHR | 1:1 | 3125.0 | 3125.0 | 400000 | 800000 | 2.1 | 9 | 17 |
| CARHSP1 | 1:1 | 781.3 | 1562.5 | 100000 | 400000 | 1.8 | 11 | 17 |
| ANXA10 | 1:1 | 48.8 | 97.7 | 25000 | 200000 | 2.4 | 8 | 19 |
| DRAXIN | 1:1 | 97.7 | 195.3 | 50000 | 200000 | 2.4 | 8 | 9 |
| SERPINB9 | 1:1 | 1562.5 | 3125.0 | 400000 | 800000 | 2.1 | 8 | 7 |
| EFNA4 | 1:1 | 6.1 | 6.1 | 3125 | 12500 | 2.7 | 7 | 5 |
| PTPRN2 | 1:1 | 12.2 | 48.8 | 25000 | 50000 | 2.7 | 9 | 7 |
| NAAA | 1:1 | 97.7 | 195.3 | 100000 | 800000 | 2.7 | 7 | 7 |
| CDCP1 | 1:1 | 48.8 | 97.7 | 6250 | 12500 | 1.8 | 7 | 6 |
| CLPP | 1:1 | 48.8 | 195.3 | 100000 | 800000 | 2.7 | 7 | 10 |

| | | | | | | | | |
|---|---|---|---|---|---|---|---|---|
| CRIP2 | 1:1 | 781.3 | 1562.5 | 200000 | 800000 | 2.1 | 7 | 6 |
| GGT5 | 1:1 | 781.3 | 1562.5 | 400000 | 800000 | 2.4 | 9 | 6 |
| MAD1L1 | 1:1 | | | | | | 9 | 5 |
| PPP3R1 | 1:1 | | | | | | 8 | 7 |
| IL18RAP | 1:1 | 12.2 | 12.2 | 6250 | 50000 | 2.7 | 9 | 5 |
| NMNAT1 | 1:1 | | | | | | 7 | 7 |
| ABHD14B | 1:1 | | | | | | 8 | 16 |
| FLRT2 | 1:1 | 12.2 | 48.8 | 25000 | 100000 | 2.7 | 9 | 8 |
| APRT | 1:1 | 390.6 | 781.3 | 400000 | 800000 | 2.7 | 10 | 6 |
| TNFRSF10A | 1:1 | 6.1 | 12.2 | 6250 | 25000 | 2.7 | 10 | 6 |
| GBP4 | 1:1 | 1562.5 | 6250.0 | 800000 | 3200000 | 2.1 | 9 | 10 |
| GPKOW | 1:1 | 3125.0 | 6250.0 | 400000 | 3200000 | 1.8 | 8 | 10 |
| CLEC10A | 1:1 | 12.2 | 48.8 | 12500 | 50000 | 2.4 | 7 | 8 |
| AKT1S1 | 1:1 | 546.9 | 1093.8 | 70000 | 280000 | 1.8 | 9 | 16 |
| CASP10 | 1:1 | 97.7 | 195.3 | 50000 | 200000 | 2.4 | 7 | 12 |
| BAG3 | 1:1 | 97.7 | 97.7 | 50000 | 200000 | 2.7 | 9 | 6 |
| STAMBP | 1:1 | 24.4 | 48.8 | 25000 | 200000 | 2.7 | 10 | 7 |
| VWC2 | 1:1 | 97.7 | 195.3 | 25000 | 50000 | 2.1 | 8 | 6 |
| DKK4 | 1:1 | | | | | | 9 | 5 |
| WWP2 | 1:1 | 97.7 | 390.6 | 400000 | 800000 | 3.0 | 11 | 10 |
| PAEP | 1:1 | 46.4 | 92.8 | 47500 | 380000 | 2.7 | 10 | 13 |
| SPOCK1 | 1:1 | 390.6 | 390.6 | 200000 | 800000 | 2.7 | 9 | 5 |
| LAYN | 1:1 | 3.1 | 6.1 | 6250 | 50000 | 3.0 | 9 | 6 |
| SETMAR | 1:1 | 390.6 | 1562.5 | 400000 | 800000 | 2.4 | 8 | 4 |
| FHIT | 1:1 | 6.1 | 12.2 | 1563 | 6250 | 2.1 | 10 | 14 |
| IL6 | 1:1 | 0.8 | 1.5 | 1563 | 12500 | 3.0 | 10 | 8 |
| PTPN1 | 1:1 | | | | | | 8 | 12 |
| IMPA1 | 1:1 | 195.3 | 781.3 | 200000 | 800000 | 2.4 | 9 | 9 |
| DARS1 | 1:1 | 48.8 | 97.7 | 100000 | 400000 | 3.0 | 10 | 13 |
| CRTAM | 1:1 | 12.2 | 48.8 | 12500 | 200000 | 2.4 | 9 | 8 |
| NGF | 1:1 | 48.8 | 97.7 | 12500 | 25000 | 2.1 | 7 | 4 |
| ULBP2 | 1:1 | 24.4 | 48.8 | 100000 | 200000 | 3.3 | 9 | 8 |
| PHOSPHO1 | 1:1 | 3.1 | 6.1 | 781 | 6250 | 2.1 | 8 | 6 |
| RSPO1 | 1:1 | 3.1 | 3.1 | 1563 | 12500 | 2.7 | 8 | 7 |
| TIGAR | 1:1 | 24.4 | 195.3 | 100000 | 400000 | 2.7 | 7 | 24 |
| FKBP7 | 1:1 | 97.7 | 195.3 | 100000 | 800000 | 2.7 | 7 | 10 |
| SKAP1 | 1:1 | 48.8 | 195.3 | 50000 | 200000 | 2.4 | 8 | 8 |
| EREG | 1:1 | 6.1 | 6.1 | 1563 | 6250 | 2.4 | 6 | 6 |
| FKBP5 | 1:1 | 97.7 | 195.3 | 25000 | 50000 | 2.1 | 9 | 6 |
| MAP4K5 | 1:1 | 12.2 | 48.8 | 6250 | 50000 | 2.1 | 6 | 10 |
| DPEP1 | 1:1 | | | | | | 8 | 5 |
| SNCG | 1:1 | 24.4 | 48.8 | 50000 | 800000 | 3.0 | 9 | 8 |
| TBCC | 1:1 | 781.3 | 781.3 | 400000 | 800000 | 2.7 | 8 | 8 |
| CD63 | 1:1 | 24.4 | 48.8 | 12500 | 50000 | 2.4 | 9 | 6 |
| MATN3 | 1:1 | 12.2 | 12.2 | 25000 | 200000 | 3.3 | 7 | 6 |
| MASP1 | 1:1 | 390.6 | 781.3 | 100000 | 400000 | 2.1 | 10 | 6 |
| IFNGR2 | 1:1 | 24.4 | 48.8 | 12500 | 100000 | 2.4 | 9 | 4 |
| LAIR2 | 1:1 | 0.8 | 1.5 | 781 | 6250 | 2.7 | 10 | 6 |
| CC2D1A | 1:1 | 97.7 | 195.3 | 100000 | 800000 | 2.7 | 9 | 6 |
| TNFRSF9 | 1:1 | 0.8 | 1.5 | 3125 | 6250 | 3.3 | 8 | 6 |
| IPCEF1 | 1:1 | 12.2 | 97.7 | 25000 | 50000 | 2.4 | 11 | 6 |
| EPHB6 | 1:1 | 12.2 | 48.8 | 25000 | 50000 | 2.7 | 9 | 6 |

| | | | | | | | | |
|---|---|---|---|---|---|---|---|---|
| OBP2B | 1:1 | 0.8 | 3.1 | 3125 | 12500 | 3.0 | 9 | 6 |
| CXCL8 | 1:1 | 0.2 | 0.4 | 1563 | 6250 | 3.6 | 7 | 5 |
| ADAM22 | 1:1 | 97.7 | 195.3 | 50000 | 200000 | 2.4 | 9 | 6 |
| RBKS | 1:1 | 6.1 | 6.1 | 12500 | 50000 | 3.3 | 7 | 11 |
| SFRP1 | 1:1 | 24.4 | 48.8 | 50000 | 200000 | 3.0 | 18 | 20 |
| TXLNA | 1:1 | 3.1 | 6.1 | 1563 | 12500 | 2.4 | 9 | 8 |
| AFP | 1:1 | 12.2 | 24.4 | 25000 | 100000 | 3.0 | 9 | 4 |
| FKBP4 | 1:1 | 48.8 | 195.3 | 25000 | 50000 | 2.1 | 8 | 7 |
| BCAN | 1:1 | 781.3 | 1562.5 | 200000 | 800000 | 2.1 | 7 | 5 |
| CD274 | 1:1 | 3.1 | 3.1 | 3125 | 50000 | 3.0 | 9 | 6 |
| CALCA | 1:1 | 24.4 | 48.8 | 25000 | 50000 | 2.7 | 8 | 4 |
| SCARB2 | 1:1 | 3.1 | 6.1 | 6250 | 50000 | 3.0 | 8 | 6 |
| TCL1A | 1:1 | 97.7 | 195.3 | 50000 | 200000 | 2.4 | 10 | 6 |
| PSME2 | 1:1 | 48.8 | 195.3 | 25000 | 200000 | 2.1 | 7 | 7 |
| LXN | 1:1 | 390.6 | 390.6 | 200000 | 800000 | 2.7 | 10 | 12 |
| EIF4B | 1:1 | 390.6 | 390.6 | 100000 | 200000 | 2.4 | 8 | 20 |
| RHOC | 1:1 | 24.4 | 48.8 | 50000 | 200000 | 3.0 | 8 | 8 |
| MDGA1 | 1:1 | 48.8 | 97.7 | 25000 | 200000 | 2.4 | 8 | 4 |
| LPO | 1:1 | 3.1 | 12.2 | 25000 | 200000 | 3.3 | 7 | 8 |
| TNFSF14 | 1:1 | 48.8 | 195.3 | 50000 | 200000 | 2.4 | 7 | 6 |
| DNMBP | 1:1 | 12.2 | 24.4 | 50000 | 200000 | 3.3 | 9 | 6 |
| ACVRL1 | 1:1 | 3.1 | 6.1 | 3125 | 12500 | 2.7 | 9 | 5 |
| TNFRSF6B | 1:1 | 97.7 | 195.3 | 50000 | 200000 | 2.4 | 10 | 8 |
| PRDX1 | 1:1 | 97.7 | 195.3 | 25000 | 200000 | 2.1 | 10 | 12 |
| MITD1 | 1:1 | 3.1 | 6.1 | 25000 | 50000 | 3.6 | 9 | 8 |
| TNR | 1:1 | 97.7 | 195.3 | 50000 | 200000 | 2.4 | 9 | 7 |
| TBCB | 1:1 | 24.4 | 97.7 | 100000 | 400000 | 3.0 | 7 | 10 |
| BMP4 | 1:1 | 6.1 | 48.8 | 12500 | 50000 | 2.4 | 18 | 49 |
| FUT8 | 1:1 | | | | | | 7 | 6 |
| CXCL13 | 1:10 | 3.1 | 6.1 | 1563 | 6250 | 2.4 | 6 | 10 |
| PVR | 1:10 | 97.7 | 195.3 | 200000 | 800000 | 3.0 | 9 | 8 |
| DDR1 | 1:10 | 12.2 | 12.2 | 50000 | 400000 | 3.6 | 7 | 5 |
| ENO1 | 1:10 | | | | | | 7 | 11 |
| SCARF2 | 1:10 | 48.8 | 97.7 | 200000 | 400000 | 3.3 | 6 | 7 |
| CPM | 1:10 | 12.2 | 24.4 | 25000 | 200000 | 3.0 | 5 | 5 |
| NCAN | 1:10 | 3.1 | 6.1 | 6250 | 50000 | 3.0 | 6 | 8 |
| RGMA | 1:10 | 24.4 | 48.8 | 100000 | 400000 | 3.3 | 7 | 7 |
| GOLM2 | 1:10 | 24.4 | 48.8 | 50000 | 100000 | 3.0 | 7 | 5 |
| CXCL11 | 1:10 | 1.5 | 6.1 | 6250 | 12500 | 3.0 | 7 | 9 |
| SERPINB6 | 1:10 | 24.4 | 97.7 | 50000 | 400000 | 2.7 | 7 | 7 |
| STC1 | 1:10 | | | | | | 7 | 7 |
| CD300C | 1:10 | 3.1 | 6.1 | 6250 | 25000 | 3.0 | 7 | 9 |
| OXT | 1:10 | 6.1 | 24.4 | 25000 | 200000 | 3.0 | 5 | 6 |
| SMPD1 | 1:10 | 97.7 | 195.3 | 400000 | 800000 | 3.3 | 6 | 9 |
| SULT1A1 | 1:10 | 97.7 | 195.3 | 100000 | 400000 | 2.7 | 5 | 14 |
| EZR | 1:10 | 390.6 | 781.3 | 100000 | 400000 | 2.1 | 6 | 4 |
| VCAN | 1:10 | 1.5 | 3.1 | 6250 | 25000 | 3.3 | 7 | 6 |
| SUMF2 | 1:10 | 24.4 | 48.8 | 6250 | 200000 | 2.1 | 6 | 5 |
| TXNDC5 | 1:10 | 3125.0 | 6250.0 | 800000 | 3200000 | 2.1 | 10 | 9 |
| CDH15 | 1:10 | 1562.5 | 3125.0 | 400000 | 800000 | 2.1 | 7 | 6 |
| CDH3 | 1:10 | 97.7 | 195.3 | 100000 | 200000 | 2.7 | 9 | 5 |
| PDCD5 | 1:10 | 6.1 | 24.4 | 12500 | 100000 | 2.7 | 8 | 13 |

| | | | | | | | | |
|---|---|---|---|---|---|---|---|---|
| JAM2 | 1:10 | 0.4 | 1.5 | 6250 | 25000 | 3.6 | 6 | 5 |
| MSR1 | 1:10 | 12.2 | 24.4 | 12500 | 100000 | 2.7 | 6 | 8 |
| VTA1 | 1:10 | 195.3 | 390.6 | 400000 | 800000 | 3.0 | 9 | 15 |
| CHGB | 1:10 | 390.6 | 781.3 | 400000 | 800000 | 2.7 | 7 | 7 |
| ADAM8 | 1:10 | 3.1 | 6.1 | 12500 | 25000 | 3.3 | 5 | 7 |
| DKK1 | 1:10 | 12.2 | 24.4 | 25000 | 50000 | 3.0 | 7 | 8 |
| LRPAP1 | 1:10 | 6.1 | 12.2 | 6250 | 12500 | 2.7 | 5 | 6 |
| CLEC1B | 1:10 | 12.2 | 24.4 | 12500 | 50000 | 2.7 | 11 | 12 |
| BST2 | 1:10 | 6.1 | 12.2 | 25000 | 50000 | 3.3 | 7 | 4 |
| PRL | 1:10 | 24.4 | 97.7 | 100000 | 400000 | 3.0 | 6 | 11 |
| SEMA4D | 1:10 | 97.7 | 195.3 | 50000 | 400000 | 2.4 | 6 | 7 |
| CD34 | 1:10 | 97.7 | 195.3 | 200000 | 400000 | 3.0 | 7 | 12 |
| LBR | 1:10 | 390.6 | 781.3 | 400000 | 800000 | 2.7 | 6 | 7 |
| CD8A | 1:10 | | | | | | 6 | 10 |
| CCL19 | 1:10 | | | | | | 5 | 12 |
| RWDD1 | 1:10 | 24.4 | 48.8 | 100000 | 200000 | 3.3 | 7 | 11 |
| NSFL1C | 1:10 | 48.8 | 97.7 | 100000 | 800000 | 3.0 | 8 | 8 |
| FUT3_FUT5 | 1:10 | 1.5 | 6.1 | 6250 | 50000 | 3.0 | 5 | 7 |
| CPPED1 | 1:10 | 24.4 | 48.8 | 50000 | 200000 | 3.0 | 5 | 13 |
| FOLR2 | 1:10 | 3.1 | 6.1 | 6250 | 25000 | 3.0 | 6 | 5 |
| MMP8 | 1:10 | 97.7 | 390.6 | 200000 | 800000 | 2.7 | 5 | 5 |
| BST1 | 1:10 | 48.8 | 97.7 | 50000 | 800000 | 2.7 | 6 | 7 |
| ENO2 | 1:10 | 3125.0 | 6250.0 | 3200000 | 12800000 | 2.7 | 6 | 13 |
| SCARA5 | 1:10 | 48.8 | 97.7 | 25000 | 100000 | 2.4 | 6 | 6 |
| NTRK3 | 1:10 | 12.2 | 24.4 | 25000 | 100000 | 3.0 | 6 | 5 |
| RGMB | 1:10 | 48.8 | 195.3 | 100000 | 200000 | 2.7 | 6 | 5 |
| FABP5 | 1:10 | 24.4 | 97.7 | 100000 | 800000 | 3.0 | 14 | 34 |
| LGALS8 | 1:10 | 12.2 | 48.8 | 25000 | 100000 | 2.7 | 5 | 6 |
| ROBO2 | 1:10 | 12.2 | 24.4 | 25000 | 200000 | 3.0 | 6 | 6 |
| LAMP2 | 1:10 | 97.7 | 195.3 | 200000 | 800000 | 3.0 | 5 | 6 |
| ANXA3 | 1:10 | 195.3 | 390.6 | 100000 | 200000 | 2.4 | 7 | 8 |
| PPCDC | 1:10 | 12.2 | 24.4 | 25000 | 200000 | 3.0 | 5 | 6 |
| TNFRSF8 | 1:10 | 12.2 | 48.8 | 50000 | 200000 | 3.0 | 5 | 6 |
| CPA2 | 1:10 | 97.7 | 195.3 | 400000 | 800000 | 3.3 | 6 | 6 |
| CTSS | 1:10 | 1.5 | 12.2 | 12500 | 50000 | 3.0 | 7 | 6 |
| CNTN5 | 1:10 | 24.4 | 48.8 | 25000 | 100000 | 2.7 | 5 | 6 |
| TNFRSF21 | 1:10 | 24.4 | 48.8 | 25000 | 100000 | 2.7 | 6 | 6 |
| THY1 | 1:10 | 0.4 | 0.8 | 6250 | 25000 | 3.9 | 6 | 6 |
| CD74 | 1:10 | 97.7 | 195.3 | 25000 | 100000 | 2.1 | 5 | 5 |
| AHSP | 1:100 | 6.1 | 6.1 | 6250 | 25000 | 3.0 | 7 | 5 |
| NID2 | 1:100 | 1562.5 | 3125.0 | 800000 | 800000 | 2.4 | 10 | 10 |
| CGA | 1:100 | 0.8 | 1.5 | 6250 | 50000 | 3.6 | 7 | 6 |
| FCRL5 | 1:100 | 6.1 | 12.2 | 25000 | 800000 | 3.3 | 7 | 3 |
| IL1RAP | 1:100 | 6.1 | 12.2 | 12500 | 50000 | 3.0 | 7 | 3 |
| IL7R | 1:100 | 6.1 | 12.2 | 12500 | 200000 | 3.0 | 7 | 15 |
| CNTN3 | 1:100 | 6.1 | 24.4 | 12500 | 50000 | 2.7 | 6 | 4 |
| CD177 | 1:100 | 0.8 | 1.5 | 6250 | 25000 | 3.6 | 8 | 4 |
| FCER2 | 1:100 | 1.5 | 3.1 | 6250 | 25000 | 3.3 | 7 | 6 |
| PLAU | 1:100 | 0.4 | 0.8 | 6250 | 12500 | 3.9 | 5 | 3 |
| CLEC11A | 1:100 | 12.2 | 24.4 | 25000 | 100000 | 3.0 | 8 | 6 |
| CD99L2 | 1:100 | 0.2 | 0.8 | 3125 | 6250 | 3.6 | 6 | 7 |
| OGN | 1:100 | 6.1 | 24.4 | 25000 | 50000 | 3.0 | 6 | 4 |

| | | | | | | | | |
|---|---|---|---|---|---|---|---|---|
| PIK3IP1 | 1:100 | 6.1 | 12.2 | 6250 | 25000 | 2.7 | 5 | 4 |
| PAMR1 | 1:100 | 24.4 | 48.8 | 25000 | 50000 | 2.7 | 7 | 5 |
| RELT | 1:100 | 3.1 | 12.2 | 6250 | 25000 | 2.7 | 8 | 4 |
| SPINK5 | 1:100 | 97.7 | 195.3 | 25000 | 50000 | 2.1 | 7 | 3 |
| TNFRSF1B | 1:100 | 3.1 | 12.2 | 12500 | 50000 | 3.0 | 7 | 4 |
| MIF | 1:100 | 6.1 | 12.2 | 3125 | 100000 | 2.4 | 7 | 14 |
| BIN2 | 1:100 | 3125.0 | 6250.0 | 400000 | 800000 | 1.8 | 8 | 12 |
| HARS1 | 1:100 | 48.8 | 195.3 | 100000 | 200000 | 2.7 | 6 | 16 |
| CLPS | 1:100 | 24.4 | 48.8 | 50000 | 400000 | 3.0 | 9 | 15 |
| SERPINB1 | 1:100 | 97.7 | 390.6 | 400000 | 800000 | 3.0 | 7 | 8 |
| WARS1 | 1:100 | 48.8 | 97.7 | 200000 | 400000 | 3.3 | 7 | 17 |
| CLEC14A | 1:100 | 48.8 | 97.7 | 200000 | 800000 | 3.3 | 5 | 5 |
| MYOC | 1:100 | | | | | | 6 | 8 |
| NOMO1 | 1:100 | 6.1 | 12.2 | 25000 | 200000 | 3.3 | 5 | 5 |
| NPTX1 | 1:100 | 6.1 | 12.2 | 25000 | 200000 | 3.3 | 7 | 5 |
| SUSD2 | 1:100 | 3.1 | 6.1 | 12500 | 50000 | 3.3 | 7 | 6 |
| MPO | 1:100 | 97.7 | 195.3 | 12500 | 25000 | 1.8 | 6 | 6 |
| INHBC | 1:100 | 24.4 | 24.4 | 12500 | 50000 | 2.7 | 8 | 7 |
| IGFBP4 | 1:100 | 390.6 | 390.6 | 100000 | 400000 | 2.4 | 8 | 5 |
| ITGAM | 1:100 | 97.7 | 195.3 | 200000 | 400000 | 3.0 | 8 | 6 |
| APOH | 1:100 | | | | | | 9 | 12 |
| BCAM | 1:100 | 24.4 | 48.8 | 6250 | 12500 | 2.1 | 6 | 6 |
| GGT1 | 1:100 | 6.1 | 24.4 | 50000 | 400000 | 3.3 | 6 | 5 |
| STIP1 | 1:100 | 24.4 | 97.7 | 25000 | 200000 | 2.4 | 9 | 7 |
| CCN5 | 1:100 | | | | | | 11 | 13 |
| PEBP1 | 1:100 | 48.8 | 97.7 | 50000 | 400000 | 2.7 | 9 | 10 |
| IL17RA | 1:100 | 3.1 | 6.1 | 12500 | 25000 | 3.3 | 8 | 5 |
| MMP3 | 1:100 | 6.1 | 12.2 | 6250 | 25000 | 2.7 | 6 | 4 |
| IL1R1 | 1:100 | 0.4 | 0.4 | 6250 | 12500 | 4.2 | 5 | 5 |
| CD164 | 1:100 | 24.4 | 48.8 | 12500 | 25000 | 2.4 | 8 | 5 |
| DSC2 | 1:100 | 6.1 | 24.4 | 25000 | 200000 | 3.0 | 8 | 5 |
| NRP2 | 1:100 | 24.4 | 48.8 | 25000 | 200000 | 2.7 | 10 | 7 |
| SIGLEC5 | 1:100 | 97.7 | 195.3 | 100000 | 200000 | 2.7 | 8 | 6 |
| APP | 1:100 | 195.3 | 390.6 | 200000 | 400000 | 2.7 | 5 | 7 |
| CD109 | 1:100 | 48.8 | 97.7 | 200000 | 800000 | 3.3 | 5 | 8 |
| SOD2 | 1:100 | 390.6 | 1562.5 | 800000 | 800000 | 2.7 | 7 | 4 |
| ALDH1A1 | 1:100 | 48.8 | 97.7 | 100000 | 200000 | 3.0 | 6 | 6 |
| SPINT1 | 1:100 | 6.1 | 24.4 | 25000 | 400000 | 3.0 | 5 | 3 |
| STC2 | 1:100 | 1.5 | 3.1 | 6250 | 25000 | 3.3 | 6 | 5 |
| DSG2 | 1:100 | 195.3 | 390.6 | 25000 | 50000 | 1.8 | 7 | 5 |
| VSIG4 | 1:100 | 3.1 | 6.1 | 6250 | 25000 | 3.0 | 6 | 5 |
| IGF2R | 1:100 | 97.7 | 97.7 | 50000 | 200000 | 2.7 | 7 | 6 |
| ARSA | 1:100 | 24.4 | 97.7 | 25000 | 50000 | 2.4 | 5 | 6 |
| TIMP4 | 1:100 | 3.1 | 6.1 | 3125 | 12500 | 2.7 | 7 | 5 |
| GUCA2A | 1:100 | 1.5 | 3.1 | 6250 | 25000 | 3.3 | 5 | 6 |
| FYB1 | 1:100 | 48.8 | 97.7 | 50000 | 200000 | 2.7 | 8 | 8 |
| GP6 | 1:100 | 3.1 | 3.1 | 6250 | 25000 | 3.3 | 7 | 6 |
| TREML2 | 1:100 | 3.1 | 6.1 | 6250 | 25000 | 3.0 | 7 | 5 |
| TNXB | 1:100 | 97.7 | 390.6 | 50000 | 400000 | 2.1 | 6 | 3 |
| PECAM1 | 1:100 | 6.1 | 12.2 | 50000 | 100000 | 3.6 | 8 | 4 |
| F11R | 1:100 | 0.4 | 0.8 | 3125 | 6250 | 3.6 | 11 | 6 |
| BLVRB | 1:100 | 97.7 | 390.6 | 200000 | 400000 | 2.7 | 8 | 6 |

| | | | | | | | | |
|---|---|---|---|---|---|---|---|---|
| CD99 | 1:100 | 1.5 | 3.1 | 6250 | 25000 | 3.3 | 6 | 3 |
| PIGR | 1:100 | 48.8 | 97.7 | 50000 | 200000 | 2.7 | 6 | 4 |
| CNTN4 | 1:100 | 48.8 | 97.7 | 50000 | 400000 | 2.7 | 6 | 5 |
| GNLY | 1:100 | 3.1 | 6.1 | 12500 | 50000 | 3.3 | 8 | 9 |
| MMP9 | 1:100 | 97.7 | 195.3 | 50000 | 400000 | 2.4 | 7 | 5 |
| TFF1 | 1:100 | 3.1 | 6.1 | 6250 | 25000 | 3.0 | 7 | 5 |
| GRN | 1:100 | 12.2 | 48.8 | 12500 | 25000 | 2.4 | 7 | 5 |
| CA2 | 1:100 | 97.7 | 195.3 | 200000 | 800000 | 3.0 | 8 | 6 |
| PARK7 | 1:100 | 6.1 | 12.2 | 3125 | 12500 | 2.4 | 7 | 9 |
| CTRB1 | 1:100 | 1.5 | 3.1 | 12500 | 25000 | 3.6 | 6 | 4 |
| NUDT5 | 1:100 | 48.8 | 97.7 | 25000 | 50000 | 2.4 | 7 | 10 |
| MFGE8 | 1:100 | 48.8 | 97.7 | 200000 | 800000 | 3.3 | 8 | 5 |
| PILRA | 1:100 | 0.8 | 1.5 | 12500 | 25000 | 3.9 | 5 | 4 |
| TXNRD1 | 1:100 | 1.5 | 6.1 | 6250 | 25000 | 3.0 | 8 | 4 |
| MESD | 1:100 | 1562.5 | 3125.0 | 800000 | 800000 | 2.4 | 8 | 13 |
| THBS2 | 1:100 | 24.4 | 48.8 | 100000 | 800000 | 3.3 | 6 | 4 |
| HAVCR2 | 1:100 | 1.5 | 3.1 | 6250 | 25000 | 3.3 | 6 | 4 |
| PLA2G7 | 1:100 | 195.3 | 390.6 | 200000 | 800000 | 2.7 | 7 | 5 |
| ITGA5 | 1:100 | 24.4 | 48.8 | 50000 | 400000 | 3.0 | 10 | 9 |
| TMSB10 | 1:100 | 48.8 | 97.7 | 12500 | 25000 | 2.1 | 7 | 6 |
| EFNA1 | 1:100 | 97.7 | 195.3 | 12500 | 25000 | 1.8 | 8 | 7 |
| SPINK1 | 1:100 | 24.4 | 48.8 | 12500 | 25000 | 2.4 | 8 | 5 |
| LILRA2 | 1:100 | 24.4 | 48.8 | 12500 | 50000 | 2.4 | 6 | 4 |
| DBI | 1:100 | 12.2 | 24.4 | 12500 | 25000 | 2.7 | 7 | 7 |
| TNFRSF1A | 1:100 | 1.5 | 6.1 | 12500 | 25000 | 3.3 | 7 | 3 |
| CA6 | 1:100 | 6.1 | 12.2 | 12500 | 25000 | 3.0 | 9 | 7 |
| CD300LG | 1:100 | 24.4 | 48.8 | 6250 | 25000 | 2.1 | 7 | 6 |
| B4GAT1 | 1:100 | 24.4 | 97.7 | 50000 | 400000 | 2.7 | 7 | 6 |
| NCAM2 | 1:100 | 24.4 | 48.8 | 25000 | 200000 | 2.7 | 7 | 5 |
| DNLZ | 1:1 | 3125.0 | 3125.0 | 200000 | 800000 | 1.8 | 19 | 26 |
| KIF20B | 1:1 | 12.2 | 48.8 | 12500 | 100000 | 2.4 | 18 | 12 |
| CNGB3 | 1:1 | 781.3 | 781.3 | 50000 | 200000 | 1.8 | 15 | 17 |
| CNTF | 1:1 | 195.3 | 390.6 | 400000 | 800000 | 3.0 | | |
| DCUN1D1 | 1:1 | 1562.5 | 3125.0 | 200000 | 800000 | 1.8 | 18 | |
| GABRA4 | 1:1 | 3125.0 | 3125.0 | 100000 | 400000 | 1.5 | | |
| RANBP2 | 1:1 | 3125.0 | 3125.0 | 400000 | 800000 | 2.1 | | |
| KCNC4 | 1:1 | 195.3 | 390.6 | 25000 | 100000 | 1.8 | 6 | 10 |
| ART5 | 1:1 | 390.6 | 1562.5 | 100000 | 400000 | 1.8 | 13 | |
| CEND1 | 1:1 | 6.1 | 24.4 | 6250 | 25000 | 2.4 | 15 | 13 |
| GPR101 | 1:1 | 48.8 | 97.7 | 12500 | 50000 | 2.1 | 9 | 7 |
| CTNNA1 | 1:1 | 6250.0 | 6250.0 | 200000 | 800000 | 1.5 | 7 | 20 |
| CACNA1C | 1:1 | 781.3 | 781.3 | 100000 | 800000 | 2.1 | 11 | 36 |
| LYPLA2 | 1:1 | | | | | | 8 | 32 |
| CD3D | 1:1 | 390.6 | 390.6 | 50000 | 400000 | 2.1 | 4 | |
| BRD2 | 1:1 | 1562.5 | 3125.0 | 200000 | 400000 | 1.8 | 7 | 6 |
| TJP3 | 1:1 | 1562.5 | 3125.0 | 400000 | 800000 | 2.1 | 12 | 20 |
| HTR1A | 1:1 | 1562.5 | 1562.5 | 100000 | 800000 | 1.8 | 11 | 20 |
| DEFB116 | 1:1 | 195.3 | 195.3 | 12500 | 100000 | 1.8 | 10 | 4 |
| TUBB3 | 1:1 | 1562.5 | 1562.5 | 200000 | 800000 | 2.1 | 7 | |
| IGDCC3 | 1:1 | 1562.5 | 1562.5 | 100000 | 800000 | 1.8 | 13 | |
| LMOD2 | 1:1 | 781.3 | 1562.5 | 50000 | 400000 | 1.5 | 19 | 35 |
| CLIC5 | 1:1 | 12.2 | 24.4 | 6250 | 25000 | 2.4 | 8 | 6 |

| | | | | | | | | |
|---|---|---|---|---|---|---|---|---|
| CLEC2L | 1:1 | 195.3 | 195.3 | 50000 | 200000 | 2.4 | 13 | |
| DYNLT1 | 1:1 | 390.6 | 781.3 | 50000 | 100000 | 1.8 | 14 | 19 |
| CACNB1 | 1:1 | 781.3 | 781.3 | 50000 | 400000 | 1.8 | 7 | |
| CRYM | 1:1 | | | | | | 13 | 17 |
| CDKL5 | 1:1 | 1562.5 | 1562.5 | 100000 | 800000 | 1.8 | 7 | 33 |
| OTOA | 1:1 | 97.7 | 195.3 | 50000 | 400000 | 2.4 | 10 | 20 |
| AGBL2 | 1:1 | 390.6 | 781.3 | 50000 | 800000 | 1.8 | 21 | 22 |
| FH | 1:1 | | | | | | 10 | 27 |
| C2orf69 | 1:1 | 1562.5 | 3125.0 | 400000 | 800000 | 2.1 | 14 | 15 |
| TP73 | 1:1 | 1562.5 | 1562.5 | 800000 | 800000 | 2.7 | 4 | 7 |
| FZD10 | 1:1 | 200000.0 | 200000.0 | 6400000 | 12800000 | 1.5 | | |
| BOLA1 | 1:1 | 1562.5 | 3125.0 | 400000 | 800000 | 2.1 | 19 | 11 |
| DOCK9 | 1:1 | 195.3 | 390.6 | 100000 | 400000 | 2.4 | 12 | 34 |
| OMP | 1:1 | 6.1 | 24.4 | 3125 | 25000 | 2.1 | 5 | 10 |
| ITPRIP | 1:1 | 195.3 | 195.3 | 12500 | 800000 | 1.8 | 13 | 15 |
| CBLN1 | 1:1 | 6250.0 | 6250.0 | 400000 | 1600000 | 1.8 | 14 | |
| ATXN2 | 1:1 | 781.3 | 781.3 | 50000 | 400000 | 1.8 | 12 | 28 |
| DEFB104A_DEFB | 1:1 | 97.7 | 390.6 | 25000 | 100000 | 1.8 | 10 | 21 |
| CGN | 1:1 | 781.3 | 1562.5 | 100000 | 200000 | 1.8 | 9 | 8 |
| DGCR6 | 1:1 | 390.6 | 781.3 | 50000 | 100000 | 1.8 | 13 | 13 |
| GRIK2 | 1:1 | 12.2 | 24.4 | 50000 | 100000 | 3.3 | 12 | 17 |
| MAP1LC3A | 1:1 | 3125.0 | 3125.0 | 800000 | 800000 | 2.4 | 10 | |
| BLOC1S2 | 1:1 | 195.3 | 390.6 | 25000 | 100000 | 1.8 | 24 | 5 |
| ELAVL4 | 1:1 | 24.4 | 195.3 | 12500 | 50000 | 1.8 | 15 | 24 |
| TMPRSS11B | 1:1 | 390.6 | 781.3 | 50000 | 800000 | 1.8 | 4 | |
| CHP1 | 1:1 | 12500.0 | 12500.0 | 400000 | 800000 | 1.5 | 13 | 22 |
| CORO6 | 1:1 | 3125.0 | 12500.0 | 800000 | 800000 | 1.8 | 14 | 21 |
| ARHGEF5 | 1:1 | 390.6 | 390.6 | 25000 | 50000 | 1.8 | 16 | 25 |
| LEO1 | 1:1 | 781.3 | 781.3 | 100000 | 800000 | 2.1 | 8 | 13 |
| PPP1R14D | 1:1 | 781.3 | 781.3 | 50000 | 200000 | 1.8 | 10 | 19 |
| MRPL28 | 1:1 | 1562.5 | 1562.5 | 800000 | 800000 | 2.7 | 9 | 6 |
| NMT1 | 1:1 | | | | | | 8 | 32 |
| LRTM2 | 1:1 | 24.4 | 48.8 | 12500 | 50000 | 2.4 | 9 | 9 |
| IFNL2 | 1:1 | 3125.0 | 3125.0 | 400000 | 800000 | 2.1 | 6 | 10 |
| RAB3GAP1 | 1:1 | 1562.5 | 1562.5 | 50000 | 400000 | 1.5 | 8 | 11 |
| SYNGAP1 | 1:1 | 48.8 | 97.7 | 12500 | 100000 | 2.1 | 5 | 13 |
| TFAP2A | 1:1 | 3125.0 | 6250.0 | 200000 | 800000 | 1.5 | 13 | |
| BRME1 | 1:1 | 25000.0 | 25000.0 | 1600000 | 12800000 | 1.8 | 11 | 27 |
| HTR1B | 1:1 | | | | | | 8 | 4 |
| BCAT2 | 1:1 | 195.3 | 390.6 | 50000 | 400000 | 2.1 | | |
| STEAP4 | 1:1 | 48.8 | 48.8 | 12500 | 100000 | 2.4 | 11 | 17 |
| LELP1 | 1:1 | | | | | | 4 | |
| PNMA2 | 1:1 | 12500.0 | 12500.0 | 800000 | 800000 | 1.8 | 7 | 22 |
| CSPG5 | 1:1 | 390.6 | 390.6 | 50000 | 800000 | 2.1 | 16 | 23 |
| CNTNAP4 | 1:1 | 390.6 | 390.6 | 25000 | 100000 | 1.8 | 7 | 17 |
| ARHGEF10 | 1:1 | 390.6 | 1562.5 | 100000 | 200000 | 1.8 | 6 | 22 |
| FCER1A | 1:1 | 390.6 | 781.3 | 50000 | 200000 | 1.8 | 12 | 11 |
| CACNB3 | 1:1 | 781.3 | 1562.5 | 100000 | 200000 | 1.8 | 11 | 35 |
| RBM17 | 1:1 | | | | | | 13 | 25 |
| AMPD3 | 1:1 | | | | | | 4 | 21 |
| ZHX2 | 1:1 | 390.6 | 390.6 | 50000 | 100000 | 2.1 | 6 | 8 |
| DNAJA4 | 1:1 | | | | | | 8 | 16 |

| | | | | | | | | |
|---|---|---|---|---|---|---|---|---|
| HLA-A | 1:1 | | | | | | 9 | 9 |
| GID8 | 1:1 | 781.3 | 781.3 | 25000 | 100000 | 1.5 | 6 | |
| LDLRAP1 | 1:1 | 12500.0 | 12500.0 | 800000 | 1600000 | 1.8 | | |
| VGF | 1:1 | | | | | | 9 | 10 |
| NEO1 | 1:1 | 97.7 | 97.7 | 25000 | 50000 | 2.4 | 8 | 12 |
| IL13RA2 | 1:1 | 1562.5 | 1562.5 | 800000 | 800000 | 2.7 | 10 | |
| RRP15 | 1:1 | 781.3 | 781.3 | 100000 | 800000 | 2.1 | | |
| MYLPF | 1:1 | 3125.0 | 3125.0 | 100000 | 800000 | 1.5 | 14 | 25 |
| CABP2 | 1:1 | 1562.5 | 3125.0 | 200000 | 800000 | 1.8 | 16 | 25 |
| CADPS | 1:1 | 195.3 | 390.6 | 100000 | 200000 | 2.4 | 10 | 13 |
| OTUD7B | 1:1 | 390.6 | 781.3 | 200000 | 800000 | 2.4 | 8 | 19 |
| PDLIM5 | 1:1 | 390.6 | 781.3 | 100000 | 400000 | 2.1 | 7 | 37 |
| MRI1 | 1:1 | | | | | | 9 | 36 |
| DOC2B | 1:1 | 195.3 | 390.6 | 100000 | 200000 | 2.4 | 8 | 15 |
| SLC1A4 | 1:1 | 12500.0 | 25000.0 | 1600000 | 6400000 | 1.8 | 9 | 20 |
| DUSP29 | 1:1 | 390.6 | 390.6 | 50000 | 200000 | 2.1 | 16 | 22 |
| MYL6B | 1:1 | 1562.5 | 1562.5 | 100000 | 400000 | 1.8 | 12 | 16 |
| PGM2 | 1:1 | 390.6 | 781.3 | 200000 | 800000 | 2.4 | 11 | 28 |
| SCN3A | 1:1 | 781.3 | 781.3 | 50000 | 800000 | 1.8 | | |
| CRTAP | 1:1 | | | | | | 4 | |
| TARM1 | 1:1 | 195.3 | 390.6 | 50000 | 800000 | 2.1 | | |
| NRXN3 | 1:1 | 390.6 | 390.6 | 25000 | 800000 | 1.8 | 6 | 31 |
| ARMCX2 | 1:1 | 781.3 | 1562.5 | 100000 | 800000 | 1.8 | | |
| INPP5J | 1:1 | 195.3 | 195.3 | 50000 | 100000 | 2.4 | 16 | 29 |
| SOX2 | 1:1 | 781.3 | 1562.5 | 100000 | 400000 | 1.8 | 2 | |
| HJV | 1:1 | 781.3 | 1562.5 | 100000 | 200000 | 1.8 | 12 | 10 |
| RGS10 | 1:1 | 195.3 | 195.3 | 25000 | 100000 | 2.1 | 8 | 24 |
| GIGYF2 | 1:1 | 390.6 | 390.6 | 50000 | 200000 | 2.1 | 9 | 16 |
| PIBF1 | 1:1 | 195.3 | 195.3 | 50000 | 200000 | 2.4 | 11 | 31 |
| RGCC | 1:1 | 1562.5 | 3125.0 | 200000 | 800000 | 1.8 | 8 | 31 |
| OCLN | 1:1 | 195.3 | 195.3 | 25000 | 100000 | 2.1 | 9 | 12 |
| IGBP1 | 1:1 | 1562.5 | 1562.5 | 100000 | 400000 | 1.8 | 11 | 21 |
| TOM1L2 | 1:1 | | | | | | 12 | 37 |
| AZI2 | 1:1 | 781.3 | 1562.5 | 400000 | 800000 | 2.4 | 10 | 25 |
| MYOM1 | 1:1 | 195.3 | 781.3 | 50000 | 200000 | 1.8 | 12 | |
| SCN2A | 1:1 | 195.3 | 390.6 | 25000 | 100000 | 1.8 | 9 | 23 |
| UPK3A | 1:1 | 781.3 | 781.3 | 50000 | 200000 | 1.8 | | |
| KIRREL1 | 1:1 | 1562.5 | 1562.5 | 200000 | 800000 | 2.1 | 9 | 14 |
| HDAC9 | 1:1 | | | | | | 8 | 22 |
| ECSCR | 1:1 | | | | | | 0.1 | |
| LRFN2 | 1:1 | | | | | | 21 | |
| SNAPIN | 1:1 | 781.3 | 781.3 | 100000 | 400000 | 2.1 | 11 | 23 |
| FAM171A2 | 1:1 | 781.3 | 1562.5 | 100000 | 400000 | 1.8 | 9 | 28 |
| ASPSCR1 | 1:1 | 390.6 | 781.3 | 200000 | 800000 | 2.4 | 14 | 22 |
| STXBP1 | 1:1 | 1562.5 | 3125.0 | 400000 | 800000 | 2.1 | | |
| CSNK2A1 | 1:1 | | | | | | 7 | 22 |
| GRIN2B | 1:1 | 1562.5 | 6250.0 | 400000 | 800000 | 1.8 | 9 | 12 |
| ZPR1 | 1:1 | 781.3 | 781.3 | 50000 | 800000 | 1.8 | 13 | 23 |
| SNAP25 | 1:1 | 97.7 | 390.6 | 50000 | 400000 | 2.1 | 14 | 23 |
| PCARE | 1:1 | 390.6 | 781.3 | 50000 | 100000 | 1.8 | 13 | 8 |
| KRT8 | 1:1 | 24.4 | 195.3 | 400000 | 800000 | 3.3 | 13 | 28 |
| ARID3A | 1:1 | | | | | | 12 | |

| | | | | | | | | |
|---|---|---|---|---|---|---|---|---|
| DEFB118 | 1:1 | 3125.0 | 6250.0 | 200000 | 800000 | 1.5 | 3 | 1 |
| SMS | 1:1 | 3125.0 | 3125.0 | 200000 | 400000 | 1.8 | 9 | 21 |
| SCIN | 1:1 | 48.8 | 48.8 | 25000 | 100000 | 2.7 | 12 | 16 |
| FARSA | 1:1 | 97.7 | 781.3 | 100000 | 200000 | 2.1 | 10 | 19 |
| EPB41L5 | 1:1 | 781.3 | 1562.5 | 100000 | 400000 | 1.8 | 19 | 31 |
| KLRC1 | 1:1 | 195.3 | 390.6 | 50000 | 100000 | 2.1 | 2 | |
| TRIM40 | 1:1 | | | | | | | |
| ADGRV1 | 1:1 | 781.3 | 781.3 | 100000 | 400000 | 2.1 | 19 | 34 |
| SPTBN2 | 1:1 | 195.3 | 390.6 | 25000 | 100000 | 1.8 | 7 | |
| CDH4 | 1:1 | 390.6 | 781.3 | 50000 | 200000 | 1.8 | 9 | 2 |
| SAG | 1:1 | 3125.0 | 3125.0 | 200000 | 400000 | 1.8 | | |
| BHLHE40 | 1:1 | 195.3 | 390.6 | 50000 | 400000 | 2.1 | 11 | 26 |
| TXNL1 | 1:1 | | | | | | 11 | 32 |
| MICALL2 | 1:1 | 1562.5 | 1562.5 | 50000 | 800000 | 1.5 | 16 | 15 |
| SOWAHA | 1:1 | 781.3 | 1562.5 | 200000 | 800000 | 2.1 | 14 | 38 |
| TDO2 | 1:1 | 12.2 | 48.8 | 12500 | 50000 | 2.4 | 12 | 16 |
| OSTN | 1:1 | 3125.0 | 3125.0 | 100000 | 200000 | 1.5 | 11 | 21 |
| MN1 | 1:1 | 781.3 | 781.3 | 50000 | 800000 | 1.8 | 11 | 24 |
| NLGN1 | 1:1 | 1562.5 | 1562.5 | 100000 | 800000 | 1.8 | 13 | 16 |
| KCNH2 | 1:1 | 1562.5 | 1562.5 | 50000 | 400000 | 1.5 | 6 | 7 |
| PFDN6 | 1:1 | 3125.0 | 6250.0 | 800000 | 800000 | 2.1 | 18 | 18 |
| STX1B | 1:1 | 390.6 | 1562.5 | 200000 | 800000 | 2.1 | 13 | 10 |
| CUZD1 | 1:1 | 6.1 | 24.4 | 6250 | 50000 | 2.4 | 8 | 33 |
| TPBGL | 1:1 | 25000.0 | 25000.0 | 1600000 | 6400000 | 1.8 | 16 | 34 |
| CHRM1 | 1:1 | 1562.5 | 3125.0 | 200000 | 800000 | 1.8 | 12 | 5 |
| LRRC38 | 1:1 | 1562.5 | 3125.0 | 400000 | 800000 | 2.1 | 9 | 10 |
| TBR1 | 1:1 | 1562.5 | 1562.5 | 200000 | 800000 | 2.1 | 6 | |
| FRMD7 | 1:1 | 1562.5 | 1562.5 | 50000 | 800000 | 1.5 | 7 | 3 |
| CD164L2 | 1:1 | 390.6 | 781.3 | 50000 | 200000 | 1.8 | | |
| GNGT1 | 1:1 | 390.6 | 390.6 | 25000 | 100000 | 1.8 | 11 | 14 |
| HNF1A | 1:1 | 6250.0 | 6250.0 | 400000 | 800000 | 1.8 | 9 | 8 |
| PSMD5 | 1:1 | 390.6 | 390.6 | 100000 | 800000 | 2.4 | 11 | 13 |
| MFAP3L | 1:1 | | | | | | 6 | 17 |
| CASC3 | 1:1 | | | | | | 10 | 21 |
| DTX2 | 1:1 | 1562.5 | 3125.0 | 200000 | 800000 | 1.8 | 13 | 9 |
| AMIGO1 | 1:1 | 781.3 | 781.3 | 25000 | 800000 | 1.5 | 11 | 21 |
| ERC2 | 1:1 | 97.7 | 195.3 | 100000 | 200000 | 2.7 | 16 | 23 |
| SLC44A4 | 1:1 | 195.3 | 390.6 | 100000 | 200000 | 2.4 | 5 | |
| NLGN2 | 1:1 | 3125.0 | 3125.0 | 200000 | 800000 | 1.8 | 10 | |
| CLNS1A | 1:1 | 1562.5 | 1562.5 | 100000 | 800000 | 1.8 | 10 | 23 |
| NAA80 | 1:1 | 781.3 | 3125.0 | 200000 | 400000 | 1.8 | 11 | 30 |
| DDX25 | 1:1 | | | | | | | |
| TMCO5A | 1:1 | 390.6 | 1562.5 | 100000 | 200000 | 1.8 | 14 | 19 |
| BATF | 1:1 | 48.8 | 97.7 | 25000 | 50000 | 2.4 | 11 | 9 |
| MSLNL | 1:1 | 781.3 | 1562.5 | 100000 | 800000 | 1.8 | 11 | 16 |
| PRKAG3 | 1:1 | 12500.0 | 12500.0 | 400000 | 1600000 | 1.5 | | |
| SEZ6 | 1:1 | 781.3 | 781.3 | 50000 | 200000 | 1.8 | 15 | 21 |
| GBA | 1:1 | 50000.0 | 50000.0 | 1600000 | 6400000 | 1.5 | | |
| SV2A | 1:1 | 195.3 | 390.6 | 100000 | 400000 | 2.4 | 19 | 29 |
| PTH | 1:1 | | | | | | | |
| IFT20 | 1:1 | 781.3 | 781.3 | 25000 | 100000 | 1.5 | 12 | 13 |
| RRAS | 1:1 | 390.6 | 390.6 | 100000 | 200000 | 2.4 | 16 | 30 |

| | | | | | | | | |
|---|---|---|---|---|---|---|---|---|
| LRP2 | 1:1 | 1562.5 | 1562.5 | 100000 | 800000 | 1.8 | 23 | 34 |
| RPGR | 1:1 | 781.3 | 781.3 | 50000 | 200000 | 1.8 | 15 | 16 |
| CLGN | 1:1 | 6.1 | 48.8 | 25000 | 50000 | 2.7 | 16 | 22 |
| FZD8 | 1:1 | | | | | | 22 | 13 |
| KLF4 | 1:1 | 781.3 | 1562.5 | 200000 | 800000 | 2.1 | 12 | 14 |
| SCN3B | 1:1 | 6250.0 | 6250.0 | 800000 | 800000 | 2.1 | | |
| GLP1R | 1:1 | | | | | | | |
| TSPAN7 | 1:1 | | | | | | 11 | |
| IMPG1 | 1:1 | 781.3 | 1562.5 | 100000 | 400000 | 1.8 | 10 | 10 |
| VAV3 | 1:1 | 97.7 | 195.3 | 25000 | 100000 | 2.1 | 7 | 19 |
| SH3BGRL2 | 1:1 | 6250.0 | 12500.0 | 800000 | 3200000 | 1.8 | 13 | 28 |
| AIDA | 1:1 | | | | | | | |
| AKR1B10 | 1:1 | 6250.0 | 6250.0 | 200000 | 800000 | 1.5 | 16 | |
| EFNB2 | 1:1 | 390.6 | 781.3 | 100000 | 800000 | 2.1 | 22 | 17 |
| DCC | 1:1 | | | | | | 11 | 11 |
| CAMLG | 1:1 | 781.3 | 1562.5 | 100000 | 200000 | 1.8 | 12 | 26 |
| SCT | 1:1 | | | | | | 9 | 26 |
| LYSMD3 | 1:1 | 97.7 | 195.3 | 100000 | 400000 | 2.7 | 14 | 39 |
| CHM | 1:1 | 195.3 | 390.6 | 25000 | 100000 | 1.8 | | |
| SPACA5 | 1:1 | 12.2 | 24.4 | 6250 | 25000 | 2.4 | 9 | 11 |
| MYO6 | 1:1 | | | | | | 8 | 23 |
| NFIC | 1:1 | 3125.0 | 3125.0 | 800000 | 800000 | 2.4 | 10 | 19 |
| S100A14 | 1:1 | 195.3 | 390.6 | 50000 | 400000 | 2.1 | 10 | 17 |
| KLHL41 | 1:1 | 390.6 | 390.6 | 200000 | 400000 | 2.7 | 14 | 20 |
| CDK5RAP3 | 1:1 | | | | | | 12 | 16 |
| RBFOX3 | 1:1 | 24.4 | 48.8 | 6250 | 100000 | 2.1 | 9 | 10 |
| LRP2BP | 1:1 | | | | | | 10 | 11 |
| LRTM1 | 1:1 | 3125.0 | 3125.0 | 100000 | 200000 | 1.5 | 9 | 20 |
| UROS | 1:1 | 195.3 | 390.6 | 25000 | 800000 | 1.8 | | |
| COL28A1 | 1:1 | 781.3 | 781.3 | 100000 | 400000 | 2.1 | 13 | 20 |
| NRN1 | 1:1 | | | | | | 6 | |
| TEX101 | 1:1 | 12.2 | 24.4 | 6250 | 25000 | 2.4 | 12 | 19 |
| TPSD1 | 1:1 | 195.3 | 390.6 | 50000 | 200000 | 2.1 | 12 | 14 |
| SOX9 | 1:1 | 781.3 | 1562.5 | 100000 | 400000 | 1.8 | 16 | |
| PDRG1 | 1:1 | 3125.0 | 3125.0 | 100000 | 800000 | 1.5 | 14 | 19 |
| RBP1 | 1:1 | 390.6 | 781.3 | 50000 | 800000 | 1.8 | 12 | 20 |
| DIPK1C | 1:1 | 3125.0 | 3125.0 | 200000 | 800000 | 1.8 | 10 | 22 |
| ATXN2L | 1:1 | 3125.0 | 3125.0 | 400000 | 800000 | 2.1 | 7 | 11 |
| SPAG1 | 1:1 | | | | | | 11 | 15 |
| SORBS1 | 1:1 | 1562.5 | 1562.5 | 50000 | 200000 | 1.5 | 6 | 17 |
| DLGAP5 | 1:1 | 195.3 | 195.3 | 50000 | 400000 | 2.4 | 6 | 23 |
| FBN2 | 1:1 | | | | | | | |
| CCDC28A | 1:1 | | | | | | | |
| TTF2 | 1:1 | 390.6 | 390.6 | 25000 | 100000 | 1.8 | | |
| SEPTIN3 | 1:1 | 781.3 | 1562.5 | 800000 | 800000 | 2.7 | 7 | 18 |
| ARFIP1 | 1:1 | 390.6 | 1562.5 | 400000 | 800000 | 2.4 | 13 | 29 |
| RAC3 | 1:1 | | | | | | 9 | 11 |
| AP2B1 | 1:1 | | | | | | 12 | 20 |
| CIT | 1:1 | 48.8 | 48.8 | 25000 | 100000 | 2.7 | 15 | 30 |
| SCN2B | 1:1 | 781.3 | 1562.5 | 100000 | 800000 | 1.8 | | |
| MAG | 1:1 | | | | | | 19 | 22 |
| GTPBP2 | 1:1 | 781.3 | 781.3 | 50000 | 200000 | 1.8 | 12 | 24 |

| | | | | | | | | |
|---|---|---|---|---|---|---|---|---|
| DDX53 | 1:1 | 390.6 | 781.3 | 100000 | 800000 | 2.1 | 12 | |
| PAFAH1B3 | 1:1 | 6250.0 | 6250.0 | 800000 | 800000 | 2.1 | 10 | |
| PMS1 | 1:1 | 48.8 | 97.7 | 12500 | 50000 | 2.1 | 21 | 24 |
| PPT1 | 1:1 | 3125.0 | 3125.0 | 200000 | 800000 | 1.8 | 9 | 26 |
| MDM1 | 1:1 | 1562.5 | 1562.5 | 100000 | 200000 | 1.8 | 11 | 19 |
| LETM1 | 1:1 | 781.3 | 781.3 | 50000 | 200000 | 1.8 | 11 | 22 |
| DNM3 | 1:1 | 390.6 | 390.6 | 100000 | 800000 | 2.4 | 9 | 22 |
| SLA2 | 1:1 | 3125.0 | 6250.0 | 800000 | 800000 | 2.1 | 10 | 35 |
| TSC22D1 | 1:1 | 390.6 | 390.6 | 25000 | 100000 | 1.8 | 13 | 19 |
| CCDC50 | 1:1 | 781.3 | 1562.5 | 400000 | 800000 | 2.4 | 10 | 13 |
| TMEM25 | 1:1 | 97.7 | 97.7 | 25000 | 100000 | 2.4 | 9 | 11 |
| LMOD1 | 1:1 | 24.4 | 48.8 | 12500 | 50000 | 2.4 | 8 | 23 |
| FTCD | 1:1 | 781.3 | 3125.0 | 100000 | 800000 | 1.5 | 5 | 19 |
| GPR158 | 1:1 | 781.3 | 781.3 | 50000 | 200000 | 1.8 | 9 | 9 |
| VWC2L | 1:1 | 48.8 | 97.7 | 12500 | 50000 | 2.1 | 9 | 16 |
| SAFB2 | 1:1 | 781.3 | 1562.5 | 100000 | 400000 | 1.8 | 9 | 10 |
| ARHGEF1 | 1:1 | 12500.0 | 12500.0 | 6400000 | 12800000 | 2.7 | 6 | 23 |
| PPP1R14A | 1:1 | | | | | | 11 | 32 |
| PCBP2 | 1:1 | 25000.0 | 25000.0 | 800000 | 3200000 | 1.5 | 7 | 23 |
| EDF1 | 1:1 | | | | | | 13 | 22 |
| PHACTR2 | 1:1 | 781.3 | 781.3 | 100000 | 800000 | 2.1 | | |
| GNAS | 1:1 | | | | | | 7 | 25 |
| GART | 1:1 | 781.3 | 1562.5 | 50000 | 200000 | 1.5 | 5 | 23 |
| C1QBP | 1:1 | 48.8 | 195.3 | 50000 | 100000 | 2.4 | 8 | 12 |
| GOLGA3 | 1:1 | 781.3 | 781.3 | 400000 | 800000 | 2.7 | 10 | 17 |
| RNASEH2A | 1:1 | 1562.5 | 3125.0 | 400000 | 800000 | 2.1 | 9 | 21 |
| EIF4G3 | 1:1 | 1562.5 | 6250.0 | 400000 | 800000 | 1.8 | 11 | 21 |
| HNRNPUL1 | 1:1 | 97.7 | 195.3 | 25000 | 50000 | 2.1 | 7 | 15 |
| MCEE | 1:1 | 6250.0 | 6250.0 | 400000 | 800000 | 1.8 | 11 | 12 |
| LCN15 | 1:1 | 97.7 | 390.6 | 100000 | 400000 | 2.4 | 8 | 12 |
| SUOX | 1:1 | 97.7 | 390.6 | 50000 | 100000 | 2.1 | 9 | 18 |
| SATB1 | 1:1 | 195.3 | 390.6 | 100000 | 400000 | 2.4 | 7 | 7 |
| RLN1 | 1:1 | 1562.5 | 6250.0 | 400000 | 800000 | 1.8 | 5 | 26 |
| RTN4IP1 | 1:1 | 781.3 | 781.3 | 200000 | 800000 | 2.4 | 8 | 35 |
| REEP4 | 1:1 | 195.3 | 390.6 | 100000 | 800000 | 2.4 | 6 | 9 |
| DSCAM | 1:1 | 195.3 | 195.3 | 50000 | 200000 | 2.4 | 9 | 7 |
| MYCBP2 | 1:1 | 390.6 | 390.6 | 100000 | 800000 | 2.4 | 9 | 20 |
| ELAC1 | 1:1 | 390.6 | 390.6 | 50000 | 400000 | 2.1 | 9 | 25 |
| MRPL58 | 1:1 | 781.3 | 1562.5 | 50000 | 800000 | 1.5 | 12 | 22 |
| FGD3 | 1:1 | 97.7 | 195.3 | 25000 | 100000 | 2.1 | 15 | 30 |
| NARS1 | 1:1 | | | | | | 11 | 10 |
| AKT2 | 1:1 | 6250.0 | 12500.0 | 800000 | 800000 | 1.8 | | |
| S100G | 1:1 | 97.7 | 195.3 | 25000 | 200000 | 2.1 | 10 | 14 |
| IDO1 | 1:1 | 195.3 | 390.6 | 200000 | 400000 | 2.7 | 14 | 12 |
| PSMD1 | 1:1 | | | | | | 15 | 26 |
| C1QTNF6 | 1:1 | 1562.5 | 1562.5 | 200000 | 800000 | 2.1 | 14 | 30 |
| CAMSAP1 | 1:1 | 97.7 | 195.3 | 25000 | 100000 | 2.1 | 11 | 29 |
| DHRS4L2 | 1:1 | 781.3 | 781.3 | 200000 | 800000 | 2.4 | 6 | 11 |
| STX3 | 1:1 | 1562.5 | 1562.5 | 200000 | 800000 | 2.1 | 7 | 5 |
| SYT1 | 1:1 | 195.3 | 195.3 | 50000 | 800000 | 2.4 | 7 | 10 |
| PLEKHO1 | 1:1 | 1562.5 | 6250.0 | 400000 | 800000 | 1.8 | 8 | 16 |
| OPHN1 | 1:1 | 3125.0 | 3125.0 | 200000 | 800000 | 1.8 | 9 | 21 |

| | | | | | | | | |
|---|---|---|---|---|---|---|---|---|
| SPTLC1 | 1:1 | 1562.5 | 3125.0 | 800000 | 800000 | 2.4 | 7 | 12 |
| OSBPL2 | 1:1 | 781.3 | 1562.5 | 200000 | 800000 | 2.1 | 5 | 7 |
| USP28 | 1:1 | 195.3 | 390.6 | 100000 | 400000 | 2.4 | 8 | 8 |
| GAST | 1:1 | | | | | | 6 | 30 |
| DLG4 | 1:1 | 195.3 | 390.6 | 25000 | 100000 | 1.8 | 8 | 11 |
| KLRK1 | 1:1 | 48.8 | 97.7 | 25000 | 50000 | 2.4 | 10 | 13 |
| DXO | 1:1 | 97.7 | 97.7 | 6250 | 25000 | 1.8 | 6 | 16 |
| SEPTIN8 | 1:1 | 195.3 | 390.6 | 200000 | 800000 | 2.7 | 5 | 9 |
| MTHFD2 | 1:1 | 195.3 | 195.3 | 12500 | 50000 | 1.8 | 6 | 15 |
| CCAR2 | 1:1 | 781.3 | 1562.5 | 400000 | 800000 | 2.4 | 9 | 16 |
| CDAN1 | 1:1 | 50000.0 | 50000.0 | 3200000 | 12800000 | 1.8 | | |
| TAX1BP1 | 1:1 | 24.4 | 24.4 | 12500 | 25000 | 2.7 | 6 | 11 |
| CDH23 | 1:1 | 1562.5 | 3125.0 | 100000 | 800000 | 1.5 | 8 | 13 |
| MORC3 | 1:1 | 1562.5 | 3125.0 | 800000 | 800000 | 2.4 | 5 | 5 |
| CRYBB2 | 1:1 | 97.7 | 195.3 | 100000 | 400000 | 2.7 | 6 | 14 |
| ALDH5A1 | 1:1 | 3125.0 | 6250.0 | 200000 | 400000 | 1.5 | 11 | 15 |
| FKBP14 | 1:1 | 781.3 | 781.3 | 25000 | 100000 | 1.5 | 9 | 33 |
| SCRIB | 1:1 | 390.6 | 781.3 | 50000 | 400000 | 1.8 | 10 | 24 |
| EFHD1 | 1:1 | 195.3 | 390.6 | 50000 | 200000 | 2.1 | 9 | 15 |
| DDHD2 | 1:1 | 195.3 | 390.6 | 25000 | 100000 | 1.8 | 7 | 16 |
| BRSK2 | 1:1 | 3125.0 | 3125.0 | 100000 | 400000 | 1.5 | 8 | 21 |
| PCDH12 | 1:1 | | | | | | 6 | 8 |
| GFRAL | 1:1 | 12.2 | 24.4 | 12500 | 50000 | 2.7 | 7 | 17 |
| FNDC1 | 1:1 | | | | | | 8 | 17 |
| CAPS | 1:1 | 48.8 | 48.8 | 12500 | 200000 | 2.4 | 7 | 9 |
| EIF1AX | 1:1 | | | | | | 12 | 25 |
| PRND | 1:1 | 48.8 | 48.8 | 12500 | 25000 | 2.4 | 3 | 13 |
| PNLIPRP1 | 1:1 | | | | | | 9 | 22 |
| PTPRR | 1:1 | 48.8 | 48.8 | 25000 | 100000 | 2.7 | 6 | 10 |
| TXNDC9 | 1:1 | 6250.0 | 6250.0 | 200000 | 800000 | 1.5 | 5 | 17 |
| TNIP1 | 1:1 | 97.7 | 195.3 | 25000 | 200000 | 2.1 | 4 | 12 |
| PRKAR2A | 1:1 | 781.3 | 1562.5 | 400000 | 800000 | 2.4 | 4 | 13 |
| CALCB | 1:1 | 195.3 | 195.3 | 200000 | 800000 | 3.0 | 8 | 10 |
| DNPEP | 1:1 | 390.6 | 390.6 | 50000 | 400000 | 2.1 | 3 | 19 |
| AP3S2 | 1:1 | 25000.0 | 25000.0 | 3200000 | 12800000 | 2.1 | 7 | 11 |
| CHCHD10 | 1:1 | 195.3 | 195.3 | 6250 | 25000 | 1.5 | 10 | 19 |
| NGRN | 1:1 | 781.3 | 781.3 | 200000 | 400000 | 2.4 | 6 | 7 |
| PLXDC2 | 1:1 | 781.3 | 781.3 | 50000 | 400000 | 1.8 | 10 | 19 |
| NAPRT | 1:1 | | | | | | 6 | 9 |
| YAP1 | 1:1 | 781.3 | 1562.5 | 50000 | 100000 | 1.5 | 6 | 9 |
| NT5C1A | 1:1 | 97.7 | 97.7 | 6250 | 25000 | 1.8 | 6 | 11 |
| IMPACT | 1:1 | 390.6 | 781.3 | 100000 | 400000 | 2.1 | 3 | 11 |
| DOK1 | 1:1 | 6250.0 | 6250.0 | 200000 | 800000 | 1.5 | | |
| PLB1 | 1:1 | 97.7 | 97.7 | 12500 | 100000 | 2.1 | 9 | 21 |
| TMPRSS11D | 1:1 | 12.2 | 24.4 | 6250 | 25000 | 2.4 | 8 | 8 |
| LMNB2 | 1:1 | 1562.5 | 1562.5 | 800000 | 800000 | 2.7 | 6 | 13 |
| FGFBP3 | 1:1 | 390.6 | 390.6 | 12500 | 50000 | 1.5 | 7 | 8 |
| DNAJC21 | 1:1 | 390.6 | 390.6 | 25000 | 100000 | 1.8 | 11 | 17 |
| HHEX | 1:1 | 6.1 | 6.1 | 6250 | 25000 | 3.0 | 7 | 26 |
| BLOC1S3 | 1:1 | 781.3 | 1562.5 | 50000 | 200000 | 1.5 | 7 | 17 |
| HSD17B14 | 1:1 | 390.6 | 781.3 | 200000 | 800000 | 2.4 | 6 | 22 |
| GBP1 | 1:1 | 3125.0 | 3125.0 | 3200000 | 6400000 | 3.0 | 14 | 25 |

| | | | | | | | | |
|---|---|---|---|---|---|---|---|---|
| WASHC3 | 1:1 | 195.3 | 390.6 | 25000 | 200000 | 1.8 | 5 | 17 |
| VSNL1 | 1:1 | 195.3 | 390.6 | 25000 | 100000 | 1.8 | 7 | 17 |
| PRSS53 | 1:1 | 3.1 | 6.1 | 12500 | 25000 | 3.3 | 7 | 10 |
| CASP7 | 1:1 | | | | | | 7 | 19 |
| INSL3 | 1:1 | | | | | | | |
| KIAA0319 | 1:1 | 195.3 | 195.3 | 25000 | 100000 | 2.1 | 7 | 11 |
| ESYT2 | 1:1 | 6250.0 | 12500.0 | 800000 | 800000 | 1.8 | 9 | 18 |
| MAP2 | 1:1 | 25000.0 | 50000.0 | 1600000 | 6400000 | 1.5 | 5 | 10 |
| C7orf50 | 1:1 | | | | | | 5 | 7 |
| HDGFL2 | 1:1 | | | | | | 10 | 14 |
| SNRPB2 | 1:1 | 390.6 | 781.3 | 50000 | 400000 | 1.8 | 9 | 13 |
| CLSTN3 | 1:1 | 3125.0 | 6250.0 | 200000 | 800000 | 1.5 | 6 | 15 |
| SPRR3 | 1:1 | | | | | | 12 | 18 |
| GSTT2B | 1:1 | 48.8 | 48.8 | 12500 | 100000 | 2.4 | 5 | 39 |
| PALM2 | 1:1 | 390.6 | 781.3 | 50000 | 400000 | 1.8 | 5 | 9 |
| HIP1R | 1:1 | | | | | | 7 | 10 |
| PLSCR3 | 1:1 | 1562.5 | 1562.5 | 200000 | 800000 | 2.1 | 7 | 16 |
| GCC1 | 1:1 | 97.7 | 195.3 | 50000 | 200000 | 2.4 | 4 | 36 |
| KCTD5 | 1:1 | 97.7 | 195.3 | 50000 | 100000 | 2.4 | 8 | 11 |
| CRYGD | 1:1 | 3.1 | 24.4 | 3125 | 25000 | 2.1 | 5 | 15 |
| ATXN3 | 1:1 | 1562.5 | 3125.0 | 100000 | 800000 | 1.5 | 6 | 13 |
| BAP18 | 1:1 | 195.3 | 390.6 | 50000 | 200000 | 2.1 | 7 | 22 |
| HEPACAM2 | 1:1 | 390.6 | 390.6 | 50000 | 400000 | 2.1 | 7 | 26 |
| TIMM8A | 1:1 | 390.6 | 781.3 | 50000 | 100000 | 1.8 | 10 | 27 |
| GIPC2 | 1:1 | 195.3 | 195.3 | 25000 | 100000 | 2.1 | 7 | 8 |
| SMTN | 1:1 | 1562.5 | 1562.5 | 50000 | 400000 | 1.5 | 5 | 12 |
| BCL2L1 | 1:1 | | | | | | | |
| PLCB2 | 1:1 | 390.6 | 781.3 | 50000 | 200000 | 1.8 | | |
| CTRL | 1:1 | 97.7 | 97.7 | 12500 | 25000 | 2.1 | 7 | 22 |
| MINK1 | 1:1 | | | | | | | |
| AK2 | 1:1 | | | | | | 11 | 37 |
| SH3GLB2 | 1:1 | | | | | | 6 | 16 |
| ACAA1 | 1:1 | 1562.5 | 3125.0 | 400000 | 800000 | 2.1 | 14 | 20 |
| ITGB1BP1 | 1:1 | | | | | | 11 | 14 |
| BAMBI | 1:1 | | | | | | 7 | 33 |
| LYAR | 1:1 | | | | | | 9 | 32 |
| VNN2 | 1:1 | 781.3 | 781.3 | 200000 | 800000 | 2.4 | 6 | 10 |
| LRP1 | 1:1 | 12.2 | 24.4 | 12500 | 50000 | 2.7 | 9 | 11 |
| AARSD1 | 1:1 | 48.8 | 195.3 | 200000 | 400000 | 3.0 | 8 | 16 |
| CES3 | 1:1 | 195.3 | 390.6 | 200000 | 400000 | 2.7 | 9 | 53 |
| LHB | 1:1 | 6250.0 | 12500.0 | 800000 | 1600000 | 1.8 | 10 | 16 |
| CALB1 | 1:1 | 0.8 | 1.5 | 6250 | 25000 | 3.6 | 8 | 13 |
| IGF1R | 1:1 | 97.7 | 195.3 | 100000 | 800000 | 2.7 | 6 | 12 |
| ABL1 | 1:1 | 12.2 | 24.4 | 50000 | 200000 | 3.3 | 9 | 11 |
| KAZALD1 | 1:1 | 97.7 | 195.3 | 100000 | 200000 | 2.7 | 7 | 11 |
| ERP44 | 1:1 | 97.7 | 195.3 | 100000 | 400000 | 2.7 | 7 | 10 |
| ELOA | 1:1 | 3.1 | 6.1 | 12500 | 50000 | 3.3 | 7 | 10 |
| GRPEL1 | 1:1 | 12.2 | 24.4 | 50000 | 200000 | 3.3 | 7 | 13 |
| UBAC1 | 1:1 | 48.8 | 195.3 | 25000 | 800000 | 2.1 | 10 | 11 |
| SEMA4C | 1:1 | 24.4 | 48.8 | 50000 | 800000 | 3.0 | 10 | 8 |
| PODXL | 1:1 | 97.7 | 195.3 | 12500 | 100000 | 1.8 | 8 | 8 |
| DKKL1 | 1:1 | 48.8 | 97.7 | 50000 | 800000 | 2.7 | 8 | 11 |

| | | | | | | | | |
|---|---|---|---|---|---|---|---|---|
| APBB1IP | 1:1 | 195.3 | 390.6 | 100000 | 400000 | 2.4 | 10 | 10 |
| PRDX6 | 1:1 | | | | | | 6 | 15 |
| RBP5 | 1:1 | 97.7 | 195.3 | 12500 | 50000 | 1.8 | 8 | 11 |
| XCL1 | 1:1 | | | | | | 4 | 7 |
| FGF21 | 1:1 | 12.2 | 24.4 | 25000 | 200000 | 3.0 | 4 | 5 |
| CBLN4 | 1:1 | 97.7 | 195.3 | 200000 | 800000 | 3.0 | 4 | 27 |
| EBI3_IL27 | 1:1 | 97.7 | 390.6 | 400000 | 800000 | 3.0 | 3 | 8 |
| F3 | 1:1 | 0.8 | 1.5 | 6250 | 25000 | 3.6 | 7 | 4 |
| ITGAV | 1:1 | 48.8 | 97.7 | 12500 | 100000 | 2.1 | 3 | 4 |
| POLR2F | 1:1 | 12.2 | 24.4 | 6250 | 12500 | 2.4 | 10 | 11 |
| RILP | 1:1 | 24.4 | 48.8 | 25000 | 50000 | 2.7 | 5 | 4 |
| METAP2 | 1:1 | 781.3 | 1562.5 | 200000 | 800000 | 2.1 | 4 | 8 |
| CRNN | 1:1 | 1.5 | 3.1 | 6250 | 12500 | 3.3 | 3 | 4 |
| CASP8 | 1:1 | 0.8 | 1.5 | 6250 | 12500 | 3.6 | 6 | 13 |
| FUS | 1:1 | 100000.0 | 100000.0 | 6400000 | 12800000 | 1.8 | 8 | 16 |
| BIRC2 | 1:1 | 24.4 | 195.3 | 50000 | 200000 | 2.4 | 11 | 24 |
| BAIAP2 | 1:1 | | | | | | 9 | 11 |
| CLEC6A | 1:1 | 1.5 | 3.1 | 6250 | 12500 | 3.3 | 10 | 18 |
| CCL8 | 1:1 | 0.8 | 1.5 | 1563 | 6250 | 3.0 | 4 | 5 |
| LEFTY2 | 1:1 | | | | | | 4 | 10 |
| RTBDN | 1:1 | 97.7 | 195.3 | 50000 | 200000 | 2.4 | 9 | 14 |
| HBEGF | 1:1 | 0.8 | 1.5 | 3125 | 12500 | 3.3 | 4 | 4 |
| WIF1 | 1:1 | 48.8 | 48.8 | 25000 | 400000 | 2.7 | 4 | 6 |
| DPP6 | 1:1 | 97.7 | 195.3 | 200000 | 800000 | 3.0 | 9 | 13 |
| SMAD5 | 1:1 | 195.3 | 390.6 | 25000 | 200000 | 1.8 | 8 | 12 |
| SIGLEC6 | 1:1 | 12.2 | 24.4 | 25000 | 200000 | 3.0 | 3 | 5 |
| DSG4 | 1:1 | 12.2 | 24.4 | 6250 | 25000 | 2.4 | 6 | 11 |
| ADM | 1:1 | 781.3 | 1562.5 | 400000 | 800000 | 2.4 | 4 | 8 |
| PPM1A | 1:1 | 390.6 | 781.3 | 50000 | 200000 | 1.8 | 8 | 14 |
| MAEA | 1:1 | | | | | | 9 | 42 |
| TNF | 1:1 | 6.1 | 12.2 | 12500 | 100000 | 3.0 | 7 | 20 |
| RUVBL1 | 1:1 | 781.3 | 3125.0 | 400000 | 800000 | 2.1 | 11 | 15 |
| ARHGAP25 | 1:1 | 195.3 | 390.6 | 100000 | 800000 | 2.4 | 10 | 16 |
| CRH | 1:1 | 1562.5 | 3125.0 | 400000 | 800000 | 2.1 | 12 | 27 |
| PLXDC1 | 1:1 | 48.8 | 97.7 | 25000 | 200000 | 2.4 | 12 | 14 |
| CRACR2A | 1:1 | 24.4 | 195.3 | 25000 | 200000 | 2.1 | 11 | 16 |
| CCN4 | 1:1 | 12.2 | 24.4 | 6250 | 25000 | 2.4 | 3 | 4 |
| PRTG | 1:1 | 6.1 | 12.2 | 6250 | 100000 | 2.7 | 5 | 6 |
| SORD | 1:1 | | | | | | 6 | 11 |
| DDX58 | 1:1 | 1562.5 | 3125.0 | 800000 | 800000 | 2.4 | 8 | 13 |
| EGFL7 | 1:1 | | | | | | 11 | 18 |
| GFAP | 1:1 | 97.7 | 195.3 | 400000 | 800000 | 3.3 | 14 | 23 |
| FCGR2B | 1:1 | | | | | | 12 | 27 |
| GFER | 1:1 | 781.3 | 1562.5 | 100000 | 800000 | 1.8 | 9 | 17 |
| CDKN2D | 1:1 | | | | | | 8 | 23 |
| AKT3 | 1:1 | 48.8 | 97.7 | 50000 | 200000 | 2.7 | 9 | 24 |
| SCP2 | 1:1 | 24.4 | 195.3 | 100000 | 200000 | 2.7 | 12 | 12 |
| KIR2DL3 | 1:1 | 24.4 | 195.3 | 25000 | 200000 | 2.1 | 9 | 18 |
| RRM2 | 1:1 | 390.6 | 781.3 | 50000 | 400000 | 1.8 | 11 | 20 |
| FEN1 | 1:1 | | | | | | 11 | 25 |
| HGS | 1:1 | 3125.0 | 6250.0 | 800000 | 800000 | 2.1 | 11 | 19 |
| PDGFC | 1:1 | 195.3 | 781.3 | 100000 | 800000 | 2.1 | 10 | 16 |

| | | | | | | | | |
|---|---|---|---|---|---|---|---|---|
| ZBTB16 | 1:1 | 24.4 | 48.8 | 50000 | 200000 | 3.0 | 9 | 18 |
| CCT5 | 1:1 | | | | | | 9 | 38 |
| RANGAP1 | 1:1 | 6250.0 | 6250.0 | 1600000 | 12800000 | 2.4 | 10 | 11 |
| PRKRA | 1:1 | 3125.0 | 6250.0 | 400000 | 800000 | 1.8 | 11 | 15 |
| LYPD8 | 1:1 | 24.4 | 48.8 | 25000 | 200000 | 2.7 | 10 | 16 |
| MAP3K5 | 1:1 | 6250.0 | 6250.0 | 1600000 | 12800000 | 2.4 | 13 | 21 |
| GH2 | 1:1 | 390.6 | 781.3 | 100000 | 200000 | 2.1 | 14 | 19 |
| MZT1 | 1:1 | 48.8 | 195.3 | 50000 | 100000 | 2.4 | 9 | 27 |
| SLAMF6 | 1:1 | 390.6 | 781.3 | 50000 | 800000 | 1.8 | 13 | 13 |
| KIR3DL1 | 1:1 | 6.1 | 12.2 | 50000 | 100000 | 3.6 | 15 | 35 |
| STX6 | 1:1 | 390.6 | 781.3 | 100000 | 800000 | 2.1 | 11 | 13 |
| PQBP1 | 1:1 | 195.3 | 390.6 | 50000 | 200000 | 2.1 | 8 | 22 |
| BTC | 1:1 | 3.1 | 6.1 | 3125 | 12500 | 2.7 | 10 | 20 |
| SFTPA1 | 1:1 | 3125.0 | 3125.0 | 100000 | 400000 | 1.5 | 10 | 29 |
| VTCN1 | 1:1 | 24.4 | 48.8 | 50000 | 200000 | 3.0 | 11 | 41 |
| NTF4 | 1:1 | 6.1 | 12.2 | 6250 | 200000 | 2.7 | 7 | 26 |
| KLK12 | 1:1 | 24.4 | 48.8 | 12500 | 100000 | 2.4 | 8 | 19 |
| ANKRD54 | 1:1 | 97.7 | 195.3 | 100000 | 200000 | 2.7 | 9 | 15 |
| RAD23B | 1:1 | 1562.5 | 3125.0 | 400000 | 800000 | 2.1 | 12 | 16 |
| FCRLB | 1:1 | 97.7 | 195.3 | 50000 | 200000 | 2.4 | 11 | 17 |
| NINJ1 | 1:1 | 97.7 | 195.3 | 100000 | 800000 | 2.7 | 11 | 20 |
| UXS1 | 1:1 | 1562.5 | 3125.0 | 800000 | 800000 | 2.4 | 8 | 22 |
| ADCYAP1R1 | 1:1 | 24.4 | 48.8 | 6250 | 800000 | 2.1 | 8 | 9 |
| ATP6AP2 | 1:1 | | | | | | 12 | 14 |
| SMAD1 | 1:1 | 48.8 | 195.3 | 400000 | 800000 | 3.3 | 14 | 24 |
| CEP20 | 1:1 | 24.4 | 48.8 | 100000 | 800000 | 3.3 | 10 | 23 |
| ARHGAP1 | 1:1 | 24.4 | 97.7 | 12500 | 50000 | 2.1 | 8 | 15 |
| RASSF2 | 1:1 | 195.3 | 390.6 | 100000 | 800000 | 2.4 | 10 | 18 |
| PSRC1 | 1:1 | | | | | | 9 | 16 |
| CEACAM5 | 1:1 | | | | | | 15 | 23 |
| SFTPA2 | 1:1 | 195.3 | 781.3 | 100000 | 200000 | 2.1 | 12 | 15 |
| STX16 | 1:1 | | | | | | 10 | 20 |
| RABEPK | 1:1 | 1562.5 | 3125.0 | 800000 | 6400000 | 2.4 | 10 | 11 |
| SH2B3 | 1:1 | 781.3 | 1562.5 | 100000 | 800000 | 1.8 | 9 | 16 |
| PSMA1 | 1:1 | 6250.0 | 12500.0 | 400000 | 800000 | 1.5 | 15 | 35 |
| RNF41 | 1:1 | 312.5 | 625.0 | 320000 | 640000 | 2.7 | 10 | 11 |
| CA11 | 1:1 | 1562.5 | 3125.0 | 400000 | 800000 | 2.1 | 11 | 18 |
| LTBP3 | 1:1 | 97.7 | 195.3 | 25000 | 100000 | 2.1 | 11 | 27 |
| GSAP | 1:1 | | | | | | 10 | 18 |
| RARRES1 | 1:1 | 97.7 | 781.3 | 100000 | 800000 | 2.1 | 10 | 14 |
| BGN | 1:1 | 24.4 | 48.8 | 25000 | 100000 | 2.7 | 14 | 21 |
| RRM2B | 1:1 | | | | | | 12 | 28 |
| PDP1 | 1:1 | | | | | | 8 | 16 |
| MAGED1 | 1:1 | 3.1 | 3.1 | 1563 | 12500 | 2.7 | 9 | 54 |
| CAMKK1 | 1:1 | 97.7 | 195.3 | 25000 | 200000 | 2.1 | 11 | 21 |
| MAVS | 1:1 | 390.6 | 781.3 | 100000 | 800000 | 2.1 | 10 | 13 |
| FES | 1:1 | 6250.0 | 12500.0 | 400000 | 800000 | 1.5 | 5 | |
| LSM1 | 1:1 | 48.8 | 195.3 | 25000 | 100000 | 2.1 | 9 | 43 |
| NAMPT | 1:1 | 3125.0 | 3125.0 | 800000 | 1600000 | 2.4 | 13 | 16 |
| NDUFS6 | 1:1 | 48.8 | 97.7 | 25000 | 100000 | 2.4 | 11 | 20 |
| TBL1X | 1:1 | 24.4 | 48.8 | 12500 | 100000 | 2.4 | 11 | 19 |
| FLI1 | 1:1 | 1.5 | 3.1 | 1563 | 3125 | 2.7 | 7 | 37 |

| | | | | | | | | |
|---|---|---|---|---|---|---|---|---|
| IKZF2 | 1:1 | 6.1 | 6.1 | 6250 | 25000 | 3.0 | 8 | 21 |
| FMR1 | 1:1 | 97.7 | 97.7 | 200000 | 800000 | 3.3 | 8 | 18 |
| TP53 | 1:1 | 781.3 | 1562.5 | 400000 | 800000 | 2.4 | 10 | 23 |
| CHAC2 | 1:1 | 97.7 | 195.3 | 50000 | 200000 | 2.4 | 7 | 19 |
| TACC3 | 1:1 | 24.4 | 195.3 | 25000 | 800000 | 2.1 | 8 | 25 |
| MPI | 1:1 | 195.3 | 390.6 | 200000 | 800000 | 2.7 | 9 | 12 |
| ATG4A | 1:1 | 1562.5 | 3125.0 | 800000 | 800000 | 2.4 | 8 | 15 |
| SORCS2 | 1:1 | 1562.5 | 1562.5 | 200000 | 800000 | 2.1 | 10 | 24 |
| AKR1B1 | 1:1 | 1562.5 | 1562.5 | 400000 | 800000 | 2.4 | 11 | 18 |
| FLT3 | 1:1 | 48.8 | 97.7 | 25000 | 100000 | 2.4 | 10 | 11 |
| HS3ST3B1 | 1:1 | 390.6 | 781.3 | 400000 | 800000 | 2.7 | 8 | 12 |
| ADAMTS15 | 1:1 | 195.3 | 781.3 | 200000 | 800000 | 2.4 | 9 | 14 |
| CDHR2 | 1:1 | 3125.0 | 6250.0 | 800000 | 800000 | 2.1 | 7 | 12 |
| IL6 | 1:1 | 0.8 | 1.5 | 3125 | 12500 | 3.3 | 9 | 8 |
| LPCAT2 | 1:1 | | | | | | 8 | 15 |
| AIF1 | 1:1 | | | | | | 11 | 20 |
| CEACAM3 | 1:1 | | | | | | 16 | 31 |
| GNE | 1:1 | 781.3 | 1562.5 | 400000 | 800000 | 2.4 | 8 | 11 |
| ENTPD2 | 1:1 | 6.1 | 12.2 | 6250 | 25000 | 2.7 | 9 | 27 |
| KRT18 | 1:1 | 6.1 | 12.2 | 12500 | 100000 | 3.0 | 7 | 11 |
| AGR3 | 1:1 | 50000.0 | 50000.0 | 6400000 | 12800000 | 2.1 | 11 | 13 |
| SUGT1 | 1:1 | | | | | | 11 | 23 |
| SLAMF8 | 1:1 | 6.1 | 48.8 | 6250 | 25000 | 2.1 | 10 | 18 |
| TPMT | 1:1 | 48.8 | 195.3 | 50000 | 200000 | 2.4 | 10 | 14 |
| FOXO3 | 1:1 | 1562.5 | 6250.0 | 200000 | 400000 | 1.5 | 13 | 13 |
| RP2 | 1:1 | 3125.0 | 6250.0 | 400000 | 800000 | 1.8 | 8 | 10 |
| PSMD9 | 1:1 | 48.8 | 195.3 | 100000 | 400000 | 2.7 | 9 | 9 |
| LAT2 | 1:1 | 781.3 | 1562.5 | 400000 | 800000 | 2.4 | 7 | 16 |
| VPS53 | 1:1 | 97.7 | 195.3 | 50000 | 200000 | 2.4 | 8 | 9 |
| CD207 | 1:1 | 48.8 | 48.8 | 50000 | 400000 | 3.0 | 10 | 13 |
| SERPINA9 | 1:1 | 6.1 | 12.2 | 50000 | 200000 | 3.6 | 7 | 16 |
| SF3B4 | 1:1 | 97.7 | 195.3 | 25000 | 100000 | 2.1 | 9 | 14 |
| COX5B | 1:1 | 97.7 | 195.3 | 50000 | 100000 | 2.4 | 5 | 14 |
| STAT5B | 1:1 | 24.4 | 48.8 | 200000 | 400000 | 3.6 | 14 | 20 |
| CFC1 | 1:1 | 97.7 | 195.3 | 100000 | 200000 | 2.7 | 10 | 29 |
| DPEP2 | 1:1 | 97.7 | 390.6 | 50000 | 400000 | 2.1 | 10 | 11 |
| MME | 1:1 | 24.4 | 48.8 | 25000 | 100000 | 2.7 | 9 | 9 |
| TBC1D23 | 1:1 | 48.8 | 97.7 | 12500 | 25000 | 2.1 | 10 | 26 |
| FLT1 | 1:1 | 781.3 | 1562.5 | 200000 | 400000 | 2.1 | 8 | 9 |
| CD28 | 1:1 | 195.3 | 390.6 | 100000 | 800000 | 2.4 | 11 | 15 |
| GSTA3 | 1:1 | 24.4 | 48.8 | 25000 | 100000 | 2.7 | 10 | 13 |
| MED18 | 1:1 | | | | | | 10 | 14 |
| STXBP3 | 1:1 | | | | | | 8 | 24 |
| GFOD2 | 1:1 | 100000.0 | 400000.0 | 12800000 | 12800000 | 1.5 | 8 | 16 |
| CEP85 | 1:1 | 87.9 | 175.8 | 45000 | 180000 | 2.4 | 9 | 31 |
| NFKBIE | 1:1 | 24.4 | 48.8 | 25000 | 200000 | 2.7 | 10 | 15 |
| PDCD1LG2 | 1:1 | 24.4 | 97.7 | 6250 | 400000 | 1.8 | 9 | 12 |
| DDAH1 | 1:1 | 97.7 | 390.6 | 100000 | 800000 | 2.4 | 11 | 11 |
| SEPTIN9 | 1:1 | | | | | | 7 | 9 |
| CDC27 | 1:1 | 6.1 | 12.2 | 50000 | 200000 | 3.6 | 10 | 29 |
| NPTN | 1:1 | 390.6 | 781.3 | 50000 | 200000 | 1.8 | 12 | 15 |
| AIFM1 | 1:1 | 97.7 | 195.3 | 25000 | 800000 | 2.1 | 11 | 8 |

| | | | | | | | | |
|---|---|---|---|---|---|---|---|---|
| SEZ6L2 | 1:1 | 48.8 | 195.3 | 100000 | 400000 | 2.7 | 7 | 7 |
| IGSF3 | 1:1 | 6.1 | 12.2 | 25000 | 200000 | 3.3 | 8 | 6 |
| PPME1 | 1:1 | 390.6 | 781.3 | 400000 | 800000 | 2.7 | 9 | 17 |
| YES1 | 1:1 | 24.4 | 48.8 | 12500 | 50000 | 2.4 | 11 | 16 |
| ARSB | 1:1 | 48.8 | 195.3 | 100000 | 800000 | 2.7 | 8 | 12 |
| ST3GAL1 | 1:1 | 3125.0 | 6250.0 | 800000 | 800000 | 2.1 | 9 | 8 |
| DRG2 | 1:1 | 3125.0 | 6250.0 | 800000 | 800000 | 2.1 | 7 | 12 |
| CD302 | 1:1 | 195.3 | 390.6 | 25000 | 800000 | 1.8 | 10 | 9 |
| STX4 | 1:1 | 195.3 | 390.6 | 100000 | 800000 | 2.4 | 9 | 10 |
| ERBIN | 1:1 | 97.7 | 195.3 | 12500 | 25000 | 1.8 | 7 | 13 |
| INPPL1 | 1:1 | 97.7 | 195.3 | 50000 | 100000 | 2.4 | 6 | 19 |
| OMG | 1:1 | 6.1 | 12.2 | 12500 | 25000 | 3.0 | 12 | 12 |
| CD38 | 1:1 | 6.1 | 12.2 | 6250 | 25000 | 2.7 | 7 | 14 |
| CTSV | 1:1 | 24.4 | 48.8 | 6250 | 12500 | 2.1 | 4 | 6 |
| TAFA5 | 1:1 | 48.8 | 97.7 | 6250 | 25000 | 1.8 | 9 | 14 |
| CPE | 1:1 | 97.7 | 97.7 | 50000 | 800000 | 2.7 | 5 | 4 |
| HPGDS | 1:1 | 781.3 | 1562.5 | 400000 | 800000 | 2.4 | 10 | 12 |
| KIFBP | 1:1 | 390.6 | 781.3 | 400000 | 800000 | 2.7 | 11 | 19 |
| DTX3 | 1:1 | 97.7 | 195.3 | 400000 | 800000 | 3.3 | 10 | 9 |
| PFKFB2 | 1:1 | 97.7 | 195.3 | 400000 | 800000 | 3.3 | 8 | 13 |
| OPTC | 1:1 | 12.2 | 24.4 | 25000 | 100000 | 3.0 | 10 | 12 |
| USO1 | 1:1 | 195.3 | 390.6 | 100000 | 400000 | 2.4 | 11 | 23 |
| RET | 1:1 | 24.4 | 48.8 | 25000 | 400000 | 2.7 | 8 | 10 |
| LRRC25 | 1:1 | 6.1 | 24.4 | 12500 | 50000 | 2.7 | 9 | 11 |
| VPS37A | 1:1 | | | | | | 8 | 10 |
| AREG | 1:1 | 1.5 | 3.1 | 6250 | 25000 | 3.3 | 8 | 10 |
| LAG3 | 1:1 | | | | | | 10 | 8 |
| GALNT7 | 1:1 | 781.3 | 781.3 | 50000 | 800000 | 1.8 | 9 | 9 |
| CCN1 | 1:1 | 48.8 | 97.7 | 25000 | 200000 | 2.4 | 9 | 9 |
| ICOSLG | 1:1 | 48.8 | 97.7 | 12500 | 800000 | 2.1 | 9 | 13 |
| IQGAP2 | 1:1 | | | | | | 10 | 17 |
| CES2 | 1:1 | | | | | | 10 | 13 |
| FAM3B | 1:1 | 97.7 | 195.3 | 12500 | 800000 | 1.8 | 6 | 10 |
| ITGB7 | 1:1 | 12.2 | 48.8 | 50000 | 200000 | 3.0 | 4 | 6 |
| KLK13 | 1:1 | 24.4 | 48.8 | 6250 | 50000 | 2.1 | 8 | 11 |
| INPP1 | 1:1 | 1562.5 | 3125.0 | 400000 | 800000 | 2.1 | 11 | 29 |
| CA12 | 1:1 | 24.4 | 48.8 | 12500 | 100000 | 2.4 | 9 | 11 |
| SRP14 | 1:1 | 390.6 | 781.3 | 50000 | 800000 | 1.8 | 9 | 11 |
| CDKN1A | 1:1 | 390.6 | 781.3 | 400000 | 800000 | 2.7 | 9 | 12 |
| DCTN2 | 1:1 | | | | | | 11 | 9 |
| SCAMP3 | 1:1 | 195.3 | 195.3 | 50000 | 400000 | 2.4 | 10 | 9 |
| LYN | 1:1 | 6.1 | 24.4 | 3125 | 6250 | 2.1 | 10 | 11 |
| OGFR | 1:1 | 48.8 | 97.7 | 200000 | 800000 | 3.3 | 10 | 10 |
| GCG | 1:1 | | | | | | 15 | 31 |
| NUDT2 | 1:1 | 97.7 | 195.3 | 12500 | 800000 | 1.8 | 8 | 13 |
| ADGRG1 | 1:1 | 195.3 | 390.6 | 100000 | 400000 | 2.4 | 7 | 14 |
| CNPY4 | 1:1 | 97.7 | 195.3 | 12500 | 800000 | 1.8 | 10 | 7 |
| CDNF | 1:1 | 12.2 | 12.2 | 12500 | 50000 | 3.0 | 9 | 10 |
| EPHA2 | 1:1 | 12.2 | 24.4 | 25000 | 200000 | 3.0 | 7 | 9 |
| ATP6V1D | 1:1 | | | | | | 2 | 8 |
| GFRA2 | 1:1 | | | | | | 9 | 9 |
| IL12A_IL12B | 1:1 | 24.4 | 48.8 | 12500 | 100000 | 2.4 | 8 | 12 |

| | | | | | | | | |
|---|---|---|---|---|---|---|---|---|
| GPA33 | 1:1 | 6.1 | 12.2 | 6250 | 12500 | 2.7 | 9 | 10 |
| VAT1 | 1:1 | | | | | | 9 | 12 |
| FGF23 | 1:1 | 97.7 | 195.3 | 50000 | 200000 | 2.4 | 5 | 6 |
| SLITRK2 | 1:1 | 24.4 | 48.8 | 200000 | 800000 | 3.6 | 4 | 6 |
| TMPRSS15 | 1:1 | 48.8 | 195.3 | 25000 | 100000 | 2.1 | 9 | 9 |
| KLK14 | 1:1 | | | | | | 7 | 5 |
| TRIAP1 | 1:1 | 48.8 | 97.7 | 25000 | 100000 | 2.4 | 4 | 3 |
| CA14 | 1:1 | 6.1 | 12.2 | 50000 | 400000 | 3.6 | 4 | 4 |
| GALNT10 | 1:1 | 781.3 | 1562.5 | 400000 | 800000 | 2.4 | 11 | 15 |
| DPY30 | 1:1 | 48.8 | 97.7 | 6250 | 25000 | 1.8 | 5 | 4 |
| MSLN | 1:1 | | | | | | 9 | 9 |
| PDCD1 | 1:1 | 3.1 | 6.1 | 12500 | 25000 | 3.3 | 4 | 10 |
| HAO1 | 1:1 | 97.7 | 195.3 | 400000 | 800000 | 3.3 | 4 | 7 |
| ALPP | 1:1 | 3.1 | 6.1 | 12500 | 50000 | 3.3 | 8 | 11 |
| DNAJB1 | 1:1 | 3125.0 | 6250.0 | 800000 | 1600000 | 2.1 | 8 | 15 |
| MUC16 | 1:1 | | | | | | 14 | 22 |
| CXCL8 | 1:1 | 0.2 | 0.4 | 1563 | 12500 | 3.6 | 5 | 4 |
| ANGPTL7 | 1:1 | 48.8 | 97.7 | 25000 | 800000 | 2.4 | 3 | 5 |
| MMP12 | 1:1 | 3.1 | 6.1 | 12500 | 100000 | 3.3 | 5 | 5 |
| HBQ1 | 1:1 | | | | | | 4 | 9 |
| SCG2 | 1:1 | 97.7 | 195.3 | 200000 | 400000 | 3.0 | 4 | 5 |
| RBP2 | 1:1 | 48.8 | 195.3 | 12500 | 25000 | 1.8 | 8 | 8 |
| LTA4H | 1:1 | 12500.0 | 25000.0 | 1600000 | 3200000 | 1.8 | 4 | 19 |
| EDA2R | 1:1 | 0.4 | 0.8 | 3125 | 12500 | 3.6 | 4 | 5 |
| ARG1 | 1:1 | 390.6 | 781.3 | 50000 | 200000 | 1.8 | 4 | 8 |
| SMOC1 | 1:1 | 781.3 | 1562.5 | 200000 | 800000 | 2.1 | 4 | 7 |
| GCNT1 | 1:1 | 781.3 | 781.3 | 400000 | 800000 | 2.7 | 10 | 9 |
| HSPB6 | 1:1 | 48.8 | 97.7 | 25000 | 400000 | 2.4 | 3 | 6 |
| DCTN1 | 1:1 | 195.3 | 781.3 | 400000 | 800000 | 2.7 | 4 | 9 |
| SIGLEC9 | 1:1 | 12.2 | 24.4 | 6250 | 25000 | 2.4 | 5 | 3 |
| NPY | 1:1 | 1562.5 | 3125.0 | 800000 | 800000 | 2.4 | 6 | 12 |
| CLMP | 1:1 | 24.4 | 48.8 | 12500 | 400000 | 2.4 | 3 | 8 |
| LGALS7_LGALS7 | 1:1 | 25000.0 | 25000.0 | 800000 | 1600000 | 1.5 | 6 | 13 |
| SPINK6 | 1:1 | 12.2 | 24.4 | 12500 | 100000 | 2.7 | 3 | 5 |
| CAPG | 1:1 | 781.3 | 781.3 | 400000 | 800000 | 2.7 | 3 | 8 |
| FXN | 1:1 | 3125.0 | 6250.0 | 800000 | 800000 | 2.1 | 4 | 5 |
| SIRT2 | 1:1 | 24.4 | 48.8 | 50000 | 100000 | 3.0 | 10 | 12 |
| S100A12 | 1:1 | 781.3 | 1562.5 | 400000 | 800000 | 2.4 | 9 | 17 |
| AMIGO2 | 1:1 | 48.8 | 195.3 | 12500 | 50000 | 1.8 | 9 | 7 |
| DAB2 | 1:1 | 195.3 | 390.6 | 50000 | 200000 | 2.1 | 4 | 12 |
| RSPO3 | 1:1 | 24.4 | 48.8 | 50000 | 200000 | 3.0 | 3 | 6 |
| CD5 | 1:1 | 0.8 | 1.5 | 3125 | 12500 | 3.3 | 5 | 6 |
| NUCB2 | 1:1 | 97.7 | 195.3 | 12500 | 400000 | 1.8 | 4 | 5 |
| TNFRSF19 | 1:1 | 3.1 | 6.1 | 3125 | 12500 | 2.7 | 4 | 5 |
| EPS8L2 | 1:1 | 97.7 | 195.3 | 100000 | 400000 | 2.7 | 4 | 4 |
| DEFB4A_DEFB4B | 1:1 | | | | | | 12 | 12 |
| KLK4 | 1:1 | 0.2 | 0.4 | 6250 | 25000 | 4.2 | 4 | 5 |
| CALCOCO1 | 1:1 | 195.3 | 390.6 | 100000 | 400000 | 2.4 | 5 | 7 |
| MOG | 1:1 | 1.5 | 3.1 | 1563 | 3125 | 2.7 | 3 | 7 |
| HMBS | 1:1 | 390.6 | 781.3 | 200000 | 800000 | 2.4 | 5 | 8 |
| VMO1 | 1:1 | 1562.5 | 3125.0 | 200000 | 800000 | 1.8 | 13 | 11 |
| LRIG1 | 1:1 | 48.8 | 97.7 | 100000 | 800000 | 3.0 | 4 | 5 |

| | | | | | | | | |
|---|---|---|---|---|---|---|---|---|
| CA9 | 1:1 | 6.1 | 6.1 | 6250 | 50000 | 3.0 | 4 | 6 |
| TACSTD2 | 1:1 | 1.5 | 3.1 | 6250 | 25000 | 3.3 | 5 | 5 |
| HS6ST1 | 1:1 | 390.6 | 781.3 | 200000 | 400000 | 2.4 | 8 | 8 |
| ACP6 | 1:1 | 12.2 | 24.4 | 50000 | 400000 | 3.3 | 5 | 5 |
| SCLY | 1:1 | 97.7 | 195.3 | 100000 | 800000 | 2.7 | 3 | 5 |
| CRISP2 | 1:1 | 6.1 | 12.2 | 12500 | 200000 | 3.0 | 4 | 6 |
| APEX1 | 1:1 | 6.1 | 12.2 | 6250 | 200000 | 2.7 | 3 | 5 |
| CNTN2 | 1:1 | 24.4 | 48.8 | 12500 | 50000 | 2.4 | 4 | 5 |
| RTN4R | 1:1 | 48.8 | 97.7 | 200000 | 800000 | 3.3 | 4 | 7 |
| NCS1 | 1:1 | 12.2 | 24.4 | 25000 | 400000 | 3.0 | 4 | 5 |
| GALNT2 | 1:1 | 195.3 | 390.6 | 50000 | 400000 | 2.1 | 3 | 6 |
| WFDC12 | 1:1 | 48.8 | 48.8 | 6250 | 12500 | 2.1 | 4 | 16 |
| VWA1 | 1:1 | 48.8 | 195.3 | 400000 | 800000 | 3.3 | 4 | 4 |
| HDGF | 1:1 | 24.4 | 24.4 | 6250 | 25000 | 2.4 | 5 | 10 |
| HAGH | 1:1 | 781.3 | 781.3 | 200000 | 800000 | 2.4 | 6 | 14 |
| CD300LF | 1:1 | 12.2 | 48.8 | 12500 | 25000 | 2.4 | 6 | 6 |
| AMBP | 1:1 | 1562.5 | 12500.0 | 12800000 | 12800000 | 3.0 | 5 | 6 |
| CPVL | 1:1 | 97.7 | 390.6 | 200000 | 800000 | 2.7 | 4 | 7 |
| CD300E | 1:1 | 390.6 | 781.3 | 400000 | 800000 | 2.7 | 3 | 4 |
| GPC1 | 1:1 | 195.3 | 390.6 | 100000 | 400000 | 2.4 | 3 | 3 |
| GFRA1 | 1:1 | 24.4 | 48.8 | 50000 | 200000 | 3.0 | 3 | 4 |
| IDUA | 1:1 | 12.2 | 24.4 | 3125 | 12500 | 2.1 | 4 | 9 |
| DSG3 | 1:1 | 3.1 | 6.1 | 6250 | 50000 | 3.0 | 6 | 5 |
| HAVCR1 | 1:1 | 3.1 | 6.1 | 12500 | 50000 | 3.3 | 3 | 4 |
| PVALB | 1:1 | 12.2 | 12.2 | 6250 | 25000 | 2.7 | 9 | 9 |
| ANGPT2 | 1:1 | 195.3 | 390.6 | 200000 | 800000 | 2.7 | 5 | 6 |
| CD1C | 1:10 | 97.7 | 195.3 | 25000 | 400000 | 2.1 | 8 | 12 |
| WFDC2 | 1:10 | 12.2 | 24.4 | 25000 | 400000 | 3.0 | 7 | 6 |
| XPNPEP2 | 1:10 | 12.2 | 48.8 | 50000 | 200000 | 3.0 | 6 | 6 |
| CTSF | 1:10 | 97.7 | 195.3 | 50000 | 400000 | 2.4 | 7 | 8 |
| P4HB | 1:10 | 24.4 | 24.4 | 12500 | 50000 | 2.7 | 8 | 13 |
| CIAPIN1 | 1:10 | 195.3 | 390.6 | 200000 | 400000 | 2.7 | 6 | 8 |
| FOLR3 | 1:10 | 0.2 | 0.4 | 12500 | 50000 | 4.5 | 6 | 10 |
| TJAP1 | 1:10 | 195.3 | 390.6 | 400000 | 800000 | 3.0 | 6 | 6 |
| PPP1R12A | 1:10 | 97.7 | 195.3 | 25000 | 100000 | 2.1 | 9 | 22 |
| TGFBR2 | 1:10 | 1.5 | 3.1 | 12500 | 25000 | 3.6 | 7 | 7 |
| ERBB4 | 1:10 | 3.1 | 6.1 | 12500 | 50000 | 3.3 | 7 | 4 |
| NELL1 | 1:10 | 48.8 | 97.7 | 200000 | 800000 | 3.3 | 7 | 7 |
| NECTIN4 | 1:10 | 1.5 | 3.1 | 3125 | 12500 | 3.0 | 7 | 6 |
| TXNDC15 | 1:10 | 48.8 | 97.7 | 25000 | 400000 | 2.4 | 8 | 10 |
| MANSC1 | 1:10 | 48.8 | 97.7 | 50000 | 400000 | 2.7 | 6 | 9 |
| MDK | 1:10 | | | | | | 8 | 14 |
| SEZ6L | 1:10 | 195.3 | 390.6 | 100000 | 400000 | 2.4 | 7 | 7 |
| CDC37 | 1:10 | 390.6 | 781.3 | 100000 | 400000 | 2.1 | 8 | 23 |
| PODXL2 | 1:10 | 48.8 | 97.7 | 25000 | 100000 | 2.4 | 8 | 13 |
| C4BPB | 1:10 | 6.1 | 12.2 | 12500 | 100000 | 3.0 | 8 | 6 |
| NBL1 | 1:10 | 24.4 | 48.8 | 12500 | 25000 | 2.4 | 7 | 8 |
| FLT4 | 1:10 | 781.3 | 1562.5 | 400000 | 800000 | 2.4 | 7 | 5 |
| CD33 | 1:10 | 12.2 | 48.8 | 12500 | 50000 | 2.4 | 7 | 9 |
| CPXM1 | 1:10 | 195.3 | 390.6 | 200000 | 400000 | 2.7 | 8 | 10 |
| KDR | 1:10 | 3.1 | 6.1 | 25000 | 50000 | 3.6 | 6 | 10 |
| TEK | 1:10 | 48.8 | 97.7 | 100000 | 400000 | 3.0 | 7 | 9 |

| | | | | | | | | |
|---|---|---|---|---|---|---|---|---|
| NT5E | 1:10 | 6.1 | 12.2 | 25000 | 100000 | 3.3 | 7 | 8 |
| SNAP29 | 1:10 | 48.8 | 97.7 | 100000 | 400000 | 3.0 | 12 | 13 |
| SPARC | 1:10 | 195.3 | 390.6 | 200000 | 800000 | 2.7 | 8 | 10 |
| ADAMTS8 | 1:10 | 48.8 | 97.7 | 50000 | 200000 | 2.7 | 7 | 8 |
| L1CAM | 1:10 | 97.7 | 195.3 | 100000 | 400000 | 2.7 | 6 | 8 |
| HTRA2 | 1:10 | 12.2 | 24.4 | 12500 | 25000 | 2.7 | 10 | 10 |
| DCXR | 1:10 | 195.3 | 390.6 | 100000 | 800000 | 2.4 | 5 | 8 |
| SIAE | 1:10 | 195.3 | 390.6 | 50000 | 400000 | 2.1 | 6 | 5 |
| PLA2G15 | 1:10 | 24.4 | 48.8 | 50000 | 400000 | 3.0 | 7 | 6 |
| FGFR2 | 1:10 | 12.2 | 24.4 | 25000 | 400000 | 3.0 | 6 | 6 |
| FGFBP1 | 1:10 | | | | | | 7 | 8 |
| TNFRSF12A | 1:10 | 97.7 | 195.3 | 200000 | 800000 | 3.0 | 9 | 11 |
| ATOX1 | 1:10 | 48.8 | 97.7 | 12500 | 800000 | 2.1 | 9 | 15 |
| DCBLD2 | 1:10 | 6.1 | 12.2 | 12500 | 50000 | 3.0 | 8 | 7 |
| IL13RA1 | 1:10 | 24.4 | 48.8 | 12500 | 25000 | 2.4 | 6 | 9 |
| PPY | 1:10 | 97.7 | 97.7 | 12500 | 50000 | 2.1 | 7 | 12 |
| CEACAM1 | 1:10 | 6.1 | 12.2 | 25000 | 400000 | 3.3 | 8 | 8 |
| S100A4 | 1:10 | | | | | | 8 | 12 |
| CREG1 | 1:10 | 97.7 | 195.3 | 25000 | 400000 | 2.1 | 6 | 12 |
| FURIN | 1:10 | 48.8 | 97.7 | 50000 | 100000 | 2.7 | 7 | 7 |
| TFPI2 | 1:10 | 0.8 | 1.5 | 6250 | 12500 | 3.6 | 7 | 8 |
| ITGB5 | 1:10 | 97.7 | 195.3 | 100000 | 400000 | 2.7 | 8 | 14 |
| FOLR1 | 1:10 | 0.8 | 1.5 | 3125 | 12500 | 3.3 | 8 | 9 |
| KLK8 | 1:10 | 3.1 | 6.1 | 6250 | 25000 | 3.0 | 7 | 11 |
| MSRA | 1:10 | 0.8 | 1.5 | 391 | 1563 | 2.4 | 7 | 9 |
| CD27 | 1:10 | 24.4 | 48.8 | 25000 | 100000 | 2.7 | 7 | 8 |
| KLK1 | 1:10 | 1.5 | 3.1 | 6250 | 25000 | 3.3 | 7 | 8 |
| SRC | 1:10 | 24.4 | 48.8 | 12500 | 25000 | 2.4 | 7 | 14 |
| KLK10 | 1:10 | 24.4 | 48.8 | 25000 | 50000 | 2.7 | 8 | 25 |
| LYPD3 | 1:10 | 6.1 | 12.2 | 6250 | 25000 | 2.7 | 6 | 15 |
| DLL1 | 1:10 | 6.1 | 12.2 | 50000 | 100000 | 3.6 | 7 | 9 |
| VEGFC | 1:10 | 97.7 | 97.7 | 12500 | 400000 | 2.1 | 11 | 13 |
| ERBB2 | 1:10 | 1.5 | 3.1 | 12500 | 50000 | 3.6 | 7 | 8 |
| MIA | 1:10 | 31.7 | 63.5 | 65000 | 130000 | 3.0 | 6 | 6 |
| KLK11 | 1:10 | 48.8 | 97.7 | 25000 | 100000 | 2.4 | 7 | 40 |
| KLK6 | 1:10 | 1.5 | 12.2 | 3125 | 12500 | 2.4 | 7 | 5 |
| GAGE2A | 1:1 | 97.7 | 195.3 | 12500 | 400000 | 1.8 | 10 | 4 |
| CLASP1 | 1:1 | | | | | | | |
| LYZL2 | 1:1 | 6.1 | 12.2 | 1563 | 25000 | 2.1 | 9 | 23 |
| MAPRE3 | 1:1 | | | | | | 2 | 3 |
| SLC28A1 | 1:1 | | | | | | 5 | 11 |
| GATA3 | 1:1 | 100000.0 | 200000.0 | 6400000 | 12800000 | 1.5 | | |
| MANEAL | 1:1 | | | | | | 16 | 28 |
| KIR3DL2 | 1:1 | 390.6 | 781.3 | 100000 | 400000 | 2.1 | 8 | 19 |
| MYH7B | 1:1 | 390.6 | 781.3 | 50000 | 400000 | 1.8 | 15 | 4 |
| CDC25A | 1:1 | 390.6 | 781.3 | 50000 | 100000 | 1.8 | 6 | |
| UNC79 | 1:1 | 781.3 | 781.3 | 100000 | 400000 | 2.1 | 14 | 20 |
| PDE1C | 1:1 | | | | | | 9 | 21 |
| PCNA | 1:1 | 6250.0 | 6250.0 | 400000 | 3200000 | 1.8 | 3 | |
| DTNB | 1:1 | 97.7 | 97.7 | 12500 | 50000 | 2.1 | 12 | 13 |
| LAMTOR5 | 1:1 | | | | | | 9 | 13 |
| TSPAN8 | 1:1 | 3.1 | 12.2 | 6250 | 25000 | 2.7 | 4 | 25 |

| | | | | | | | | |
|---|---|---|---|---|---|---|---|---|
| OGA | 1:1 | 1562.5 | 3125.0 | 200000 | 800000 | 1.8 | 8 | 15 |
| ADAM9 | 1:1 | 25000.0 | 25000.0 | 1600000 | 3200000 | 1.8 | 9 | 19 |
| CD101 | 1:1 | 390.6 | 781.3 | 200000 | 800000 | 2.4 | 6 | 15 |
| CDC26 | 1:1 | 1562.5 | 1562.5 | 50000 | 800000 | 1.5 | 6 | 40 |
| DDX1 | 1:1 | | | | | | 9 | 15 |
| PALM | 1:1 | 195.3 | 195.3 | 12500 | 50000 | 1.8 | 6 | 6 |
| GPR15L | 1:1 | | | | | | 10 | 18 |
| NFU1 | 1:1 | 48.8 | 97.7 | 100000 | 400000 | 3.0 | | |
| PBK | 1:1 | 97.7 | 195.3 | 100000 | 400000 | 2.7 | 8 | 8 |
| GIMAP8 | 1:1 | 195.3 | 390.6 | 25000 | 200000 | 1.8 | 16 | 21 |
| HMMR | 1:1 | 3125.0 | 6250.0 | 800000 | 800000 | 2.1 | 9 | 13 |
| CLINT1 | 1:1 | 6250.0 | 12500.0 | 800000 | 800000 | 1.8 | 9 | 21 |
| SLIRP | 1:1 | | | | | | 12 | 22 |
| BEX3 | 1:1 | 195.3 | 390.6 | 25000 | 100000 | 1.8 | | |
| PHLDB2 | 1:1 | | | | | | 13 | 21 |
| NMRK2 | 1:1 | | | | | | | |
| ANKRA2 | 1:1 | | | | | | | |
| CREB3 | 1:1 | 195.3 | 390.6 | 25000 | 800000 | 1.8 | 12 | 7 |
| GJA8 | 1:1 | 3125.0 | 3125.0 | 200000 | 800000 | 1.8 | | |
| SMC3 | 1:1 | | | | | | 9 | |
| MAPK13 | 1:1 | 6250.0 | 6250.0 | 800000 | 800000 | 2.1 | 0.2 | |
| CDK1 | 1:1 | | | | | | 15 | 8 |
| JMJD1C | 1:1 | 1562.5 | 3125.0 | 200000 | 800000 | 1.8 | 4 | 14 |
| CYB5A | 1:1 | 195.3 | 390.6 | 50000 | 400000 | 2.1 | 11 | 18 |
| SLC34A3 | 1:1 | 1562.5 | 3125.0 | 200000 | 800000 | 1.8 | 11 | |
| CPLX2 | 1:1 | 781.3 | 1562.5 | 100000 | 400000 | 1.8 | | |
| SPRING1 | 1:1 | | | | | | 13 | 19 |
| PRAME | 1:1 | | | | | | 5 | |
| MUCL3 | 1:1 | 390.6 | 390.6 | 25000 | 100000 | 1.8 | 10 | 18 |
| STOML2 | 1:1 | | | | | | 11 | 8 |
| ZP4 | 1:1 | 24.4 | 97.7 | 12500 | 50000 | 2.1 | 1 | |
| IGLON5 | 1:1 | 781.3 | 1562.5 | 200000 | 800000 | 2.1 | 8 | 14 |
| BTNL10 | 1:1 | 3125.0 | 3125.0 | 200000 | 800000 | 1.8 | | |
| GAD1 | 1:1 | 6250.0 | 12500.0 | 800000 | 800000 | 1.8 | | |
| PDCL2 | 1:1 | 195.3 | 390.6 | 50000 | 200000 | 2.1 | 11 | 8 |
| BSND | 1:1 | 195.3 | 390.6 | 25000 | 200000 | 1.8 | | |
| ATF4 | 1:1 | 48.8 | 48.8 | 6250 | 400000 | 2.1 | 12 | |
| VWA5A | 1:1 | 781.3 | 1562.5 | 100000 | 400000 | 1.8 | 10 | 17 |
| DNAJA1 | 1:1 | | | | | | 0.3 | |
| FGF9 | 1:1 | | | | | | 1 | |
| IZUMO1 | 1:1 | 97.7 | 195.3 | 200000 | 800000 | 3.0 | | |
| ARHGAP5 | 1:1 | | | | | | 9 | 14 |
| TRIM26 | 1:1 | | | | | | | |
| MORF4L2 | 1:1 | 781.3 | 781.3 | 50000 | 400000 | 1.8 | 11 | 11 |
| VCPKMT | 1:1 | | | | | | 8 | 23 |
| ESPL1 | 1:1 | 390.6 | 781.3 | 50000 | 400000 | 1.8 | 11 | 18 |
| PSMC3 | 1:1 | | | | | | | |
| CREBZF | 1:1 | 6250.0 | 6250.0 | 800000 | 800000 | 2.1 | | |
| CYTH3 | 1:1 | 781.3 | 781.3 | 50000 | 400000 | 1.8 | | |
| IL25 | 1:1 | 195.3 | 390.6 | 100000 | 400000 | 2.4 | 4 | |
| KHDC3L | 1:1 | | | | | | 12 | 21 |
| BCL7A | 1:1 | 781.3 | 781.3 | 50000 | 800000 | 1.8 | | |

| | | | | | | | | |
|---|---|---|---|---|---|---|---|---|
| TMOD4 | 1:1 | 1562.5 | 3125.0 | 200000 | 800000 | 1.8 | 7 | |
| CDC123 | 1:1 | 1562.5 | 1562.5 | 100000 | 800000 | 1.8 | 16 | |
| MORN4 | 1:1 | 390.6 | 390.6 | 100000 | 400000 | 2.4 | 10 | 20 |
| SSH3 | 1:1 | 6250.0 | 12500.0 | 800000 | 3200000 | 1.8 | 9 | 3 |
| REST | 1:1 | | | | | | 8 | 28 |
| CEP152 | 1:1 | 97.7 | 195.3 | 25000 | 50000 | 2.1 | 16 | 36 |
| PITHD1 | 1:1 | 1562.5 | 1562.5 | 100000 | 400000 | 1.8 | 7 | 20 |
| NEK7 | 1:1 | 6250.0 | 6250.0 | 800000 | 800000 | 2.1 | 9 | 15 |
| CEACAM18 | 1:1 | 195.3 | 390.6 | 50000 | 200000 | 2.1 | 10 | 12 |
| EPN1 | 1:1 | | | | | | 11 | 20 |
| CEP350 | 1:1 | | | | | | 18 | |
| FOS | 1:1 | 48.8 | 97.7 | 50000 | 200000 | 2.7 | | |
| FGF7 | 1:1 | 195.3 | 195.3 | 12500 | 800000 | 1.8 | 14 | 13 |
| SHC1 | 1:1 | | | | | | | |
| IL3 | 1:1 | 195.3 | 781.3 | 200000 | 800000 | 2.4 | | |
| EVI2B | 1:1 | 781.3 | 1562.5 | 100000 | 400000 | 1.8 | 8 | 16 |
| PDXDC1 | 1:1 | 3125.0 | 3125.0 | 200000 | 400000 | 1.8 | | |
| ATP1B4 | 1:1 | | | | | | 14 | 12 |
| DCUN1D2 | 1:1 | 1562.5 | 1562.5 | 100000 | 800000 | 1.8 | 2 | |
| TAGLN3 | 1:1 | | | | | | 15 | |
| H2AP | 1:1 | 97.7 | 195.3 | 25000 | 50000 | 2.1 | | |
| TOP1MT | 1:1 | 3125.0 | 6250.0 | 400000 | 800000 | 1.8 | 9 | 10 |
| TPRKB | 1:1 | 3125.0 | 6250.0 | 400000 | 3200000 | 1.8 | 11 | |
| GRSF1 | 1:1 | 390.6 | 781.3 | 50000 | 100000 | 1.8 | 16 | 21 |
| DCDC2C | 1:1 | 48.8 | 97.7 | 25000 | 100000 | 2.4 | | |
| RBM25 | 1:1 | 3125.0 | 3125.0 | 200000 | 800000 | 1.8 | 4 | |
| BTLA | 1:1 | 195.3 | 195.3 | 12500 | 50000 | 1.8 | 18 | 33 |
| THAP12 | 1:1 | 48.8 | 97.7 | 25000 | 100000 | 2.4 | 14 | 26 |
| MRPL52 | 1:1 | | | | | | 8 | |
| ERCC1 | 1:1 | 781.3 | 3125.0 | 200000 | 800000 | 1.8 | | |
| DCLRE1C | 1:1 | | | | | | 12 | |
| PDIA2 | 1:1 | 781.3 | 1562.5 | 200000 | 800000 | 2.1 | 13 | 18 |
| GABARAPL1 | 1:1 | | | | | | | |
| PAIP2B | 1:1 | | | | | | 19 | 18 |
| RASGRF1 | 1:1 | 97.7 | 195.3 | 25000 | 50000 | 2.1 | | |
| PFDN4 | 1:1 | | | | | | 14 | 22 |
| RAD51 | 1:1 | 24.4 | 97.7 | 50000 | 200000 | 2.7 | 7 | |
| MORF4L1 | 1:1 | | | | | | | |
| PDIA5 | 1:1 | | | | | | 14 | 31 |
| CD82 | 1:1 | 195.3 | 390.6 | 25000 | 50000 | 1.8 | 13 | 15 |
| MLLT1 | 1:1 | 781.3 | 781.3 | 100000 | 800000 | 2.1 | 21 | |
| STAU1 | 1:1 | | | | | | 9 | 17 |
| COL9A2 | 1:1 | 3125.0 | 6250.0 | 400000 | 800000 | 1.8 | 12 | 25 |
| PPIE | 1:1 | 1562.5 | 3125.0 | 400000 | 800000 | 2.1 | 14 | 6 |
| NFKB2 | 1:1 | 195.3 | 390.6 | 25000 | 200000 | 1.8 | | |
| SHPK | 1:1 | 1562.5 | 3125.0 | 200000 | 800000 | 1.8 | 12 | 11 |
| TXK | 1:1 | 6250.0 | 12500.0 | 800000 | 800000 | 1.8 | 8 | 25 |
| RNF43 | 1:1 | 390.6 | 781.3 | 100000 | 400000 | 2.1 | 12 | 18 |
| TLR2 | 1:1 | 195.3 | 390.6 | 100000 | 400000 | 2.4 | 16 | 30 |
| HRAS | 1:1 | 97.7 | 195.3 | 25000 | 50000 | 2.1 | 6 | 9 |
| TAP1 | 1:1 | | | | | | 10 | 3 |
| CEP290 | 1:1 | 390.6 | 781.3 | 50000 | 100000 | 1.8 | 12 | 10 |

| | | | | | | | | |
|---|---|---|---|---|---|---|---|---|
| PAFAH2 | 1:1 | 3125.0 | 6250.0 | 400000 | 800000 | 1.8 | 14 | 17 |
| GTF2IRD1 | 1:1 | 781.3 | 1562.5 | 100000 | 400000 | 1.8 | 8 | 13 |
| GAS2 | 1:1 | 1562.5 | 1562.5 | 200000 | 800000 | 2.1 | 10 | 15 |
| ID4 | 1:1 | 390.6 | 781.3 | 100000 | 200000 | 2.1 | 8 | 14 |
| IFIT3 | 1:1 | 781.3 | 781.3 | 100000 | 200000 | 2.1 | 8 | 14 |
| SLK | 1:1 | | | | | | 14 | 17 |
| MAPKAPK2 | 1:1 | 1562.5 | 1562.5 | 200000 | 800000 | 2.1 | 13 | 16 |
| UPK3BL1 | 1:1 | 195.3 | 390.6 | 25000 | 50000 | 1.8 | 9 | 16 |
| CA7 | 1:1 | 1562.5 | 3125.0 | 400000 | 800000 | 2.1 | | |
| TMED1 | 1:1 | 12500.0 | 25000.0 | 800000 | 6400000 | 1.5 | 20 | |
| ACADM | 1:1 | | | | | | 14 | 27 |
| CASP4 | 1:1 | 97.7 | 195.3 | 200000 | 400000 | 3.0 | 10 | 24 |
| TET2 | 1:1 | 195.3 | 390.6 | 25000 | 50000 | 1.8 | 12 | 12 |
| ALMS1 | 1:1 | 390.6 | 390.6 | 25000 | 50000 | 1.8 | 14 | 34 |
| TPPP2 | 1:1 | | | | | | 12 | 9 |
| TCP11 | 1:1 | 1562.5 | 1562.5 | 200000 | 800000 | 2.1 | 8 | |
| VPS28 | 1:1 | 3125.0 | 6250.0 | 400000 | 800000 | 1.8 | 12 | 18 |
| VSTM2B | 1:1 | | | | | | 5 | 21 |
| OFD1 | 1:1 | | | | | | 7 | 11 |
| TRIM24 | 1:1 | 195.3 | 390.6 | 50000 | 100000 | 2.1 | 14 | 21 |
| LSM8 | 1:1 | 6250.0 | 12500.0 | 800000 | 12800000 | 1.8 | 8 | 9 |
| TEX33 | 1:1 | | | | | | | |
| TK1 | 1:1 | | | | | | 10 | 3 |
| SEPTIN7 | 1:1 | | | | | | 14 | 21 |
| OGT | 1:1 | 12500.0 | 25000.0 | 800000 | 800000 | 1.5 | 3 | 11 |
| KIAA1549 | 1:1 | | | | | | 20 | 14 |
| SAT2 | 1:1 | 97.7 | 97.7 | 25000 | 50000 | 2.4 | 13 | 17 |
| MTHFSD | 1:1 | | | | | | 15 | 34 |
| PPP1R12B | 1:1 | 781.3 | 781.3 | 50000 | 200000 | 1.8 | 17 | 20 |
| ZNF75D | 1:1 | 1562.5 | 3125.0 | 200000 | 800000 | 1.8 | 6 | 17 |
| BRD1 | 1:1 | 1562.5 | 1562.5 | 100000 | 800000 | 1.8 | 13 | |
| CD3E | 1:1 | 97.7 | 195.3 | 100000 | 800000 | 2.7 | 15 | |
| PARD3 | 1:1 | | | | | | 12 | |
| FUT1 | 1:1 | 3125.0 | 3125.0 | 200000 | 800000 | 1.8 | 11 | 14 |
| ARAF | 1:1 | 1562.5 | 1562.5 | 100000 | 800000 | 1.8 | 12 | 7 |
| CTAG1A | 1:1 | 48.8 | 97.7 | 25000 | 50000 | 2.4 | 17 | |
| PRUNE2 | 1:1 | 195.3 | 390.6 | 50000 | 200000 | 2.1 | 14 | 17 |
| SPRR1B | 1:1 | | | | | | 11 | 18 |
| RGL2 | 1:1 | 390.6 | 390.6 | 25000 | 50000 | 1.8 | 13 | 33 |
| IL9 | 1:1 | 48.8 | 97.7 | 12500 | 50000 | 2.1 | 11 | 12 |
| GADD45B | 1:1 | 3125.0 | 3125.0 | 200000 | 3200000 | 1.8 | 9 | 16 |
| CEACAM20 | 1:1 | 48.8 | 97.7 | 25000 | 100000 | 2.4 | 11 | 16 |
| SAP18 | 1:1 | 195.3 | 390.6 | 100000 | 400000 | 2.4 | 14 | 28 |
| EFCAB2 | 1:1 | | | | | | 13 | 13 |
| RAPGEF2 | 1:1 | | | | | | 15 | 8 |
| DYNC1H1 | 1:1 | 195.3 | 390.6 | 100000 | 400000 | 2.4 | 6 | 11 |
| GUCY2C | 1:1 | 390.6 | 390.6 | 50000 | 200000 | 2.1 | 14 | |
| LRRFIP1 | 1:1 | | | | | | 11 | 21 |
| ARHGAP30 | 1:1 | 3125.0 | 3125.0 | 200000 | 800000 | 1.8 | 16 | |
| SERPINH1 | 1:1 | 6250.0 | 6250.0 | 400000 | 800000 | 1.8 | | |
| ZNF830 | 1:1 | 6250.0 | 6250.0 | 200000 | 800000 | 1.5 | 7 | 6 |
| NENF | 1:1 | 1562.5 | 3125.0 | 200000 | 800000 | 1.8 | 8 | 20 |

| | | | | | | | | |
|---|---|---|---|---|---|---|---|---|
| MTUS1 | 1:1 | 12.2 | 24.4 | 6250 | 50000 | 2.4 | 20 | 15 |
| MAGEA3 | 1:1 | 781.3 | 1562.5 | 100000 | 800000 | 1.8 | | |
| RNF4 | 1:1 | | | | | | | |
| AMOTL2 | 1:1 | 1562.5 | 3125.0 | 400000 | 800000 | 2.1 | 9 | 10 |
| TIGIT | 1:1 | 97.7 | 195.3 | 12500 | 200000 | 1.8 | 15 | 18 |
| TRDMT1 | 1:1 | 97.7 | 195.3 | 100000 | 200000 | 2.7 | 14 | 26 |
| CCND2 | 1:1 | 1562.5 | 3125.0 | 400000 | 800000 | 2.1 | 11 | 14 |
| TARS1 | 1:1 | 3125.0 | 6250.0 | 800000 | 800000 | 2.1 | 7 | 19 |
| CDH22 | 1:1 | 12500.0 | 12500.0 | 800000 | 3200000 | 1.8 | 10 | 36 |
| ERI1 | 1:1 | | | | | | 11 | 35 |
| S100A3 | 1:1 | 97.7 | 195.3 | 25000 | 50000 | 2.1 | 9 | 25 |
| TAB2 | 1:1 | 3125.0 | 6250.0 | 400000 | 800000 | 1.8 | 14 | 22 |
| COL4A4 | 1:1 | | | | | | 19 | |
| MAMDC4 | 1:1 | 48.8 | 97.7 | 25000 | 50000 | 2.4 | 6 | 18 |
| SMAD2 | 1:1 | 25000.0 | 50000.0 | 3200000 | 12800000 | 1.8 | 8 | 13 |
| TNPO1 | 1:1 | | | | | | 12 | 9 |
| KIAA1549L | 1:1 | 1562.5 | 1562.5 | 100000 | 200000 | 1.8 | | |
| CENPJ | 1:1 | 195.3 | 195.3 | 25000 | 50000 | 2.1 | | |
| ENSA | 1:1 | 12500.0 | 12500.0 | 800000 | 1600000 | 1.8 | 12 | 25 |
| ZNRF4 | 1:1 | 781.3 | 781.3 | 100000 | 200000 | 2.1 | 7 | 23 |
| MNAT1 | 1:1 | 390.6 | 781.3 | 200000 | 800000 | 2.4 | 7 | 8 |
| DHPS | 1:1 | 781.3 | 781.3 | 25000 | 200000 | 1.5 | 13 | 20 |
| EVPL | 1:1 | 390.6 | 390.6 | 100000 | 200000 | 2.4 | 14 | 14 |
| ARG2 | 1:1 | 100000.0 | 100000.0 | 6400000 | 12800000 | 1.8 | 9 | 14 |
| FAM171B | 1:1 | 195.3 | 390.6 | 100000 | 400000 | 2.4 | 9 | 8 |
| GPRC5C | 1:1 | 781.3 | 1562.5 | 400000 | 800000 | 2.4 | 11 | 20 |
| IFIT1 | 1:1 | | | | | | | |
| PTTG1 | 1:1 | 390.6 | 781.3 | 100000 | 800000 | 2.1 | 14 | 6 |
| RAB44 | 1:1 | 1562.5 | 3125.0 | 200000 | 800000 | 1.8 | 12 | 19 |
| CINP | 1:1 | 6250.0 | 12500.0 | 1600000 | 6400000 | 2.1 | 8 | 19 |
| PRC1 | 1:1 | 1562.5 | 3125.0 | 200000 | 800000 | 1.8 | 15 | 15 |
| RCC1 | 1:1 | 390.6 | 781.3 | 50000 | 200000 | 1.8 | 8 | 12 |
| ITGAX | 1:1 | 1562.5 | 1562.5 | 100000 | 800000 | 1.8 | 9 | 37 |
| SH3GL3 | 1:1 | 6250.0 | 6250.0 | 400000 | 800000 | 1.8 | | |
| RPE | 1:1 | | | | | | 12 | 18 |
| CENPF | 1:1 | 390.6 | 390.6 | 50000 | 200000 | 2.1 | 13 | 26 |
| PKN3 | 1:1 | 1562.5 | 1562.5 | 200000 | 800000 | 2.1 | 17 | 12 |
| AP1G2 | 1:1 | 12.2 | 48.8 | 25000 | 200000 | 2.7 | 8 | 15 |
| ENOPH1 | 1:1 | | | | | | 9 | 18 |
| GPD1 | 1:1 | 3125.0 | 3125.0 | 200000 | 800000 | 1.8 | 8 | 25 |
| WDR46 | 1:1 | 200000.0 | 200000.0 | 6400000 | 12800000 | 1.5 | 9 | 9 |
| COMMD9 | 1:1 | 195.3 | 390.6 | 50000 | 200000 | 2.1 | 11 | 18 |
| INSL4 | 1:1 | 97.7 | 195.3 | 25000 | 100000 | 2.1 | 14 | 17 |
| ERN1 | 1:1 | 195.3 | 390.6 | 100000 | 200000 | 2.4 | 9 | 13 |
| KIAA2013 | 1:1 | 12500.0 | 12500.0 | 800000 | 800000 | 1.8 | | |
| NDST1 | 1:1 | 195.3 | 390.6 | 50000 | 200000 | 2.1 | 9 | 14 |
| VSIG2 | 1:1 | 195.3 | 195.3 | 6250 | 50000 | 1.5 | 11 | 13 |
| DUSP13 | 1:1 | 781.3 | 781.3 | 50000 | 400000 | 1.8 | 10 | 29 |
| LACRT | 1:1 | 6250.0 | 6250.0 | 400000 | 3200000 | 1.8 | 10 | 25 |
| WFDC1 | 1:1 | | | | | | 11 | 12 |
| IMMT | 1:1 | 781.3 | 781.3 | 50000 | 800000 | 1.8 | 13 | 11 |
| EDDM3B | 1:1 | 24.4 | 48.8 | 3125 | 12500 | 1.8 | 11 | 33 |

| | | | | | | | | |
|---|---|---|---|---|---|---|---|---|
| UBE2B | 1:1 | | | | | | 7 | 11 |
| SUSD4 | 1:1 | 48.8 | 48.8 | 25000 | 100000 | 2.7 | 14 | 13 |
| SLC13A1 | 1:1 | | | | | | 9 | 12 |
| SMPDL3B | 1:1 | 12500.0 | 12500.0 | 800000 | 3200000 | 1.8 | 9 | 16 |
| FNTA | 1:1 | 24.4 | 48.8 | 12500 | 50000 | 2.4 | 9 | 16 |
| ALDH2 | 1:1 | 781.3 | 1562.5 | 100000 | 400000 | 1.8 | 5 | 27 |
| RALY | 1:1 | 781.3 | 781.3 | 100000 | 800000 | 2.1 | 11 | 13 |
| AHSA1 | 1:1 | | | | | | 11 | 9 |
| NMI | 1:1 | 195.3 | 390.6 | 200000 | 800000 | 2.7 | 12 | 22 |
| LTB | 1:1 | 390.6 | 390.6 | 50000 | 200000 | 2.1 | 9 | 18 |
| LARP1 | 1:1 | 781.3 | 1562.5 | 200000 | 800000 | 2.1 | 13 | 21 |
| PCDHB15 | 1:1 | 1562.5 | 3125.0 | 200000 | 800000 | 1.8 | 8 | 16 |
| RFC4 | 1:1 | 781.3 | 1562.5 | 200000 | 800000 | 2.1 | 7 | 9 |
| ACRBP | 1:1 | 195.3 | 390.6 | 25000 | 100000 | 1.8 | 12 | 12 |
| THTPA | 1:1 | 195.3 | 390.6 | 25000 | 200000 | 1.8 | 7 | 25 |
| SNX2 | 1:1 | | | | | | 8 | 17 |
| VPS4B | 1:1 | 781.3 | 1562.5 | 100000 | 800000 | 1.8 | 7 | 32 |
| EPGN | 1:1 | 195.3 | 390.6 | 25000 | 100000 | 1.8 | 2 | 8 |
| CAPN3 | 1:1 | 195.3 | 390.6 | 100000 | 800000 | 2.4 | 12 | 21 |
| MINDY1 | 1:1 | | | | | | | |
| NACC1 | 1:1 | 781.3 | 781.3 | 50000 | 200000 | 1.8 | 12 | 12 |
| LUZP2 | 1:1 | 195.3 | 390.6 | 25000 | 800000 | 1.8 | 11 | 10 |
| RAB2B | 1:1 | 3125.0 | 3125.0 | 200000 | 800000 | 1.8 | 12 | 21 |
| TDP1 | 1:1 | 195.3 | 390.6 | 25000 | 200000 | 1.8 | 19 | 36 |
| PXDNL | 1:1 | 781.3 | 781.3 | 50000 | 400000 | 1.8 | 12 | 21 |
| GLYR1 | 1:1 | | | | | | 10 | 17 |
| REG3G | 1:1 | | | | | | 18 | 13 |
| TG | 1:1 | 6.1 | 12.2 | 3125 | 12500 | 2.4 | 8 | 16 |
| IL22 | 1:1 | 12.2 | 24.4 | 6250 | 25000 | 2.4 | 13 | 15 |
| VSIG10 | 1:1 | 195.3 | 390.6 | 100000 | 400000 | 2.4 | 11 | 14 |
| BTN1A1 | 1:1 | 3.1 | 6.1 | 3125 | 25000 | 2.7 | 13 | 14 |
| IL2RG | 1:1 | 390.6 | 781.3 | 50000 | 800000 | 1.8 | 12 | 13 |
| CMIP | 1:1 | 6250.0 | 12500.0 | 800000 | 3200000 | 1.8 | 8 | 16 |
| YJU2 | 1:1 | 781.3 | 781.3 | 50000 | 200000 | 1.8 | 12 | 13 |
| PPP1CC | 1:1 | 1562.5 | 3125.0 | 200000 | 800000 | 1.8 | 6 | 20 |
| SCRIB | 1:1 | 195.3 | 390.6 | 50000 | 800000 | 2.1 | 6 | 20 |
| LMOD1 | 1:1 | | | | | | 14 | 24 |
| MTSS2 | 1:1 | | | | | | 14 | 32 |
| CEACAM19 | 1:1 | 48.8 | 97.7 | 6250 | 50000 | 1.8 | 17 | 23 |
| GABARAP | 1:1 | 1562.5 | 1562.5 | 100000 | 400000 | 1.8 | 11 | 23 |
| EIF2AK2 | 1:1 | | | | | | 6 | 17 |
| SLMAP | 1:1 | 97.7 | 195.3 | 12500 | 50000 | 1.8 | 7 | 21 |
| LRRC37A2 | 1:1 | 781.3 | 1562.5 | 100000 | 400000 | 1.8 | 5 | 19 |
| SH3BP1 | 1:1 | 1562.5 | 3125.0 | 200000 | 800000 | 1.8 | 6 | 29 |
| TADA3 | 1:1 | | | | | | 13 | 21 |
| GORASP2 | 1:1 | 6250.0 | 6250.0 | 400000 | 1600000 | 1.8 | 7 | 25 |
| GPHA2 | 1:1 | 97.7 | 97.7 | 6250 | 50000 | 1.8 | 5 | 7 |
| TGFB2 | 1:1 | 12500.0 | 12500.0 | 400000 | 1600000 | 1.5 | 15 | 23 |
| SDHB | 1:1 | 781.3 | 1562.5 | 200000 | 800000 | 2.1 | 7 | 35 |
| PBXIP1 | 1:1 | | | | | | 11 | 9 |
| DNAJB14 | 1:1 | 48.8 | 97.7 | 25000 | 100000 | 2.4 | 8 | 13 |
| PTPRH | 1:1 | 12.2 | 48.8 | 25000 | 200000 | 2.7 | 7 | 16 |

| | | | | | | | | |
|---|---|---|---|---|---|---|---|---|
| SMAD3 | 1:1 | 12500.0 | 12500.0 | 800000 | 800000 | 1.8 | 13 | 17 |
| KIR2DS4 | 1:1 | 48.8 | 97.7 | 12500 | 200000 | 2.1 | 7 | 17 |
| KIR2DL2 | 1:1 | 97.7 | 97.7 | 6250 | 50000 | 1.8 | 15 | 27 |
| ITIH5 | 1:1 | 781.3 | 1562.5 | 100000 | 400000 | 1.8 | 17 | 37 |
| TRIM58 | 1:1 | 390.6 | 390.6 | 25000 | 200000 | 1.8 | 7 | 11 |
| USP25 | 1:1 | 97.7 | 195.3 | 12500 | 50000 | 1.8 | 7 | 14 |
| UNG | 1:1 | | | | | | 12 | 28 |
| KLK15 | 1:1 | 97.7 | 97.7 | 12500 | 100000 | 2.1 | 13 | 20 |
| CHCHD6 | 1:1 | 781.3 | 781.3 | 100000 | 400000 | 2.1 | 9 | 10 |
| STAM | 1:1 | | | | | | 10 | 14 |
| HSPA2 | 1:1 | 195.3 | 781.3 | 50000 | 200000 | 1.8 | 11 | 13 |
| MPRIP | 1:1 | | | | | | 15 | 18 |
| SDCCAG8 | 1:1 | 6.1 | 24.4 | 12500 | 50000 | 2.7 | 10 | 19 |
| CCDC134 | 1:1 | 1562.5 | 3125.0 | 200000 | 800000 | 1.8 | 16 | 10 |
| MCTS1 | 1:1 | | | | | | 4 | 8 |
| DYNLT3 | 1:1 | | | | | | 14 | 28 |
| SLC9A3R2 | 1:1 | | | | | | 9 | 20 |
| DNM1 | 1:1 | 1562.5 | 3125.0 | 200000 | 800000 | 1.8 | 6 | 40 |
| DUT | 1:1 | 1562.5 | 3125.0 | 400000 | 800000 | 2.1 | 13 | 29 |
| PCDH7 | 1:1 | 3125.0 | 3125.0 | 200000 | 800000 | 1.8 | 5 | 12 |
| PCSK7 | 1:1 | 1562.5 | 3125.0 | 200000 | 800000 | 1.8 | 18 | 22 |
| BCL2 | 1:1 | 195.3 | 195.3 | 50000 | 200000 | 2.4 | 11 | 19 |
| SSNA1 | 1:1 | | | | | | 6 | 18 |
| ZFYVE19 | 1:1 | 1562.5 | 3125.0 | 400000 | 800000 | 2.1 | 5 | 29 |
| ATRAID | 1:1 | 48.8 | 48.8 | 12500 | 50000 | 2.4 | 7 | 9 |
| ZNRD2 | 1:1 | 1562.5 | 3125.0 | 200000 | 800000 | 1.8 | 6 | 13 |
| TOP1 | 1:1 | | | | | | 6 | 21 |
| TMEM106A | 1:1 | 97.7 | 97.7 | 12500 | 50000 | 2.1 | 8 | 33 |
| BCL7B | 1:1 | 390.6 | 781.3 | 50000 | 800000 | 1.8 | 14 | |
| INSL5 | 1:1 | 97.7 | 97.7 | 12500 | 50000 | 2.1 | | |
| TNFAIP2 | 1:1 | 48.8 | 48.8 | 12500 | 50000 | 2.4 | 5 | 7 |
| NUBP1 | 1:1 | | | | | | 5 | |
| HDDC2 | 1:1 | 195.3 | 390.6 | 50000 | 800000 | 2.1 | 6 | 8 |
| IDO1 | 1:1 | 195.3 | 390.6 | 100000 | 400000 | 2.4 | 7 | 14 |
| MEP1A | 1:1 | 195.3 | 195.3 | 50000 | 800000 | 2.4 | 5 | 17 |
| CWC15 | 1:1 | 195.3 | 781.3 | 50000 | 200000 | 1.8 | 6 | 17 |
| AP3B1 | 1:1 | 195.3 | 781.3 | 50000 | 800000 | 1.8 | 7 | 26 |
| C9orf40 | 1:1 | 195.3 | 390.6 | 25000 | 200000 | 1.8 | 5 | 19 |
| SMNDC1 | 1:1 | | | | | | 8 | 14 |
| NCR3LG1 | 1:1 | 195.3 | 390.6 | 100000 | 400000 | 2.4 | 6 | 12 |
| SWAP70 | 1:1 | 781.3 | 781.3 | 50000 | 800000 | 1.8 | 6 | 8 |
| CIRBP | 1:1 | | | | | | 7 | 37 |
| IFNAR1 | 1:1 | 195.3 | 195.3 | 25000 | 800000 | 2.1 | 5 | 7 |
| ATG16L1 | 1:1 | 781.3 | 1562.5 | 200000 | 800000 | 2.1 | 5 | 10 |
| SNX18 | 1:1 | 12500.0 | 12500.0 | 800000 | 1600000 | 1.8 | 6 | 11 |
| PPP2R5A | 1:1 | | | | | | | |
| PHLDB1 | 1:1 | | | | | | 21 | 22 |
| MTSS1 | 1:1 | | | | | | | |
| LMNB1 | 1:1 | | | | | | 17 | 16 |
| MAP2K1 | 1:1 | 195.3 | 390.6 | 200000 | 800000 | 2.7 | 4 | 28 |
| NAP1L4 | 1:1 | 1562.5 | 1562.5 | 100000 | 800000 | 1.8 | 4 | 12 |
| PSCA | 1:1 | | | | | | 5 | 8 |

| | | | | | | | | |
|---|---|---|---|---|---|---|---|---|
| PSAPL1 | 1:1 | 12.2 | 97.7 | 6250 | 25000 | 1.8 | 5 | 12 |
| YARS1 | 1:1 | | | | | | | |
| CEACAM16 | 1:1 | 12.2 | 24.4 | 6250 | 50000 | 2.4 | 5 | 8 |
| CD86 | 1:1 | 97.7 | 97.7 | 12500 | 200000 | 2.1 | 6 | 7 |
| DNAJC9 | 1:1 | | | | | | 7 | 14 |
| GPIHBP1 | 1:1 | | | | | | 23 | 25 |
| EGFLAM | 1:1 | 781.3 | 1562.5 | 200000 | 800000 | 2.1 | 7 | 6 |
| FKBPL | 1:1 | 195.3 | 390.6 | 100000 | 400000 | 2.4 | 8 | 20 |
| ENDOU | 1:1 | 97.7 | 195.3 | 25000 | 50000 | 2.1 | 7 | 11 |
| SERPINI2 | 1:1 | 195.3 | 390.6 | 25000 | 200000 | 1.8 | 6 | 14 |
| ADGRE1 | 1:1 | 1562.5 | 1562.5 | 100000 | 800000 | 1.8 | 8 | 10 |
| NUDT16 | 1:1 | | | | | | 5 | 31 |
| CDC42BPB | 1:1 | | | | | | 6 | 14 |
| EFCAB14 | 1:1 | 25000.0 | 25000.0 | 1600000 | 3200000 | 1.8 | 7 | 6 |
| GIPC3 | 1:1 | 195.3 | 390.6 | 25000 | 200000 | 1.8 | 7 | 36 |
| PTPRK | 1:1 | 3125.0 | 6250.0 | 800000 | 3200000 | 2.1 | 7 | 12 |
| TRIM25 | 1:1 | 781.3 | 781.3 | 50000 | 200000 | 1.8 | 6 | 16 |
| IFI30 | 1:1 | | | | | | 5 | 11 |
| KAZN | 1:1 | 781.3 | 1562.5 | 400000 | 800000 | 2.4 | 5 | 23 |
| UFD1 | 1:1 | 781.3 | 781.3 | 200000 | 400000 | 2.4 | | |
| TMED8 | 1:1 | 97.7 | 195.3 | 25000 | 200000 | 2.1 | 5 | 32 |
| BRAP | 1:1 | 48.8 | 97.7 | 25000 | 800000 | 2.4 | 5 | 27 |
| OTUD6B | 1:1 | | | | | | 7 | 26 |
| IST1 | 1:1 | | | | | | | |
| FAM13A | 1:1 | 97.7 | 390.6 | 25000 | 200000 | 1.8 | 6 | 25 |
| COL24A1 | 1:1 | | | | | | 24 | 30 |
| SPINT3 | 1:1 | 24.4 | 24.4 | 6250 | 25000 | 2.4 | | |
| PCYT2 | 1:1 | 781.3 | 1562.5 | 100000 | 400000 | 1.8 | 6 | 22 |
| FAM3D | 1:1 | 48.8 | 97.7 | 12500 | 50000 | 2.1 | 4 | 18 |
| JPT2 | 1:1 | | | | | | 8 | 39 |
| NDUFB7 | 1:1 | | | | | | 9 | 33 |
| MTIF3 | 1:1 | 781.3 | 3125.0 | 100000 | 800000 | 1.5 | | |
| PCDH9 | 1:1 | 3125.0 | 6250.0 | 800000 | 800000 | 2.1 | 6 | 9 |
| ACOT13 | 1:1 | 781.3 | 781.3 | 25000 | 200000 | 1.5 | 8 | 37 |
| CYB5R2 | 1:1 | | | | | | 6 | 9 |
| ARF6 | 1:1 | | | | | | | |
| SUGP1 | 1:1 | 3125.0 | 6250.0 | 200000 | 800000 | 1.5 | 11 | 17 |
| GLIPR1 | 1:1 | | | | | | 14 | 19 |
| RBP7 | 1:1 | 781.3 | 1562.5 | 100000 | 200000 | 1.8 | 7 | 14 |
| CETN3 | 1:1 | 781.3 | 781.3 | 50000 | 400000 | 1.8 | 4 | 18 |

**Table S3.** Differential expression of proteins in plasma samples of cancers patients compared to healthy individuals.

| Prorein | Male | | Females | |
|---|---|---|---|---|
| | npx_dif | pvalue | npx_dif | pvalue |
| MYBPC2 | -0.53755 | 0.14878 | 0.20023 | 0.52796 |
| PSMC3 | 0.16656 | 0.40771 | 0.09687 | 0.63534 |
| LSM1 | 0.11167 | 0.63633 | 0.66939 | 0.01222 |
| MELTF | -0.12089 | 0.44802 | -0.21311 | 0.06270 |
| FCRL3 | 0.20374 | 0.30450 | -0.14799 | 0.44826 |
| FXYD5 | 0.28553 | 0.32044 | 1.29673 | 0.00002 |
| EFCAB2 | 0.24969 | 0.25550 | 0.67410 | 0.00094 |
| CD101 | 0.15190 | 0.34783 | 0.06182 | 0.71795 |
| NXPH3 | -0.05456 | 0.67390 | 0.04685 | 0.74823 |
| SHBG | 0.52716 | 0.00025 | 0.12102 | 0.44859 |
| NEDD4L | -0.17566 | 0.23970 | 1.06923 | 0.00000 |
| ARSA | -0.19961 | 0.13432 | -0.15709 | 0.14906 |
| LPP | -0.07109 | 0.74897 | 0.72533 | 0.00031 |
| WARS1 | 0.53443 | 0.00996 | 1.09616 | 0.00000 |
| ANK2 | 0.45513 | 0.11625 | -0.08243 | 0.74855 |
| JUN | 0.21414 | 0.41711 | 0.69756 | 0.00695 |
| XRCC4 | -0.05471 | 0.76992 | 0.23520 | 0.17814 |
| EPCAM | -0.09225 | 0.76451 | -0.25068 | 0.40228 |
| BST1 | -0.12302 | 0.33341 | -0.20325 | 0.10697 |
| SULT2A1 | 0.50271 | 0.22244 | 0.81810 | 0.06311 |
| APRT | -0.14449 | 0.56707 | 1.47594 | 0.00000 |
| REXO2 | 0.26703 | 0.25800 | 0.09488 | 0.70954 |
| CBX2 | 0.15008 | 0.34432 | 0.12393 | 0.36114 |
| CNTNAP2 | 0.00806 | 0.95288 | -0.14341 | 0.28538 |
| PF4 | 0.24024 | 0.39048 | -1.18087 | 0.00000 |
| CDH4 | 0.59623 | 0.03489 | 0.00385 | 0.99099 |
| CD3E | 0.63667 | 0.05407 | 0.05011 | 0.88847 |
| ANXA5 | 0.72769 | 0.00389 | 0.50998 | 0.04268 |
| MSLNL | -0.25176 | 0.37905 | 0.11452 | 0.67187 |
| VEGFB | 0.08362 | 0.46896 | 0.00934 | 0.93912 |
| VSIG4 | 0.70959 | 0.02098 | 0.64574 | 0.04060 |
| AMFR | 0.03470 | 0.76433 | 0.14209 | 0.14609 |
| CD79B | 0.35879 | 0.10883 | 0.05886 | 0.82133 |
| SPINK8 | 0.18598 | 0.15238 | -0.37221 | 0.07850 |
| CEND1 | -0.25928 | 0.35390 | -0.06525 | 0.78842 |
| APOM | 0.01435 | 0.92202 | -0.20796 | 0.22179 |
| CLASP1 | 0.26965 | 0.36995 | 0.93202 | 0.00155 |
| PTEN | 0.21479 | 0.47976 | 0.89413 | 0.00137 |
| CRKL | -0.05651 | 0.85867 | 0.87102 | 0.00914 |
| MAP4K5 | 0.42232 | 0.27593 | 2.02107 | 0.00000 |
| BRD2 | 0.33100 | 0.24115 | 0.61185 | 0.02160 |
| TNFRSF19 | 0.21898 | 0.17521 | -0.29617 | 0.15593 |

| | | | | |
|---|---|---|---|---|
| NTRK3 | -0.06471 | 0.53182 | -0.20492 | 0.06721 |
| PSMD1 | -0.14992 | 0.67360 | 0.85853 | 0.00540 |
| GP5 | 0.10966 | 0.35284 | 0.04439 | 0.68393 |
| HEXIM1 | 0.46556 | 0.06672 | 1.57554 | 0.00000 |
| CPPED1 | -0.01927 | 0.94317 | 1.36299 | 0.00000 |
| S100A12 | 0.61446 | 0.03008 | -1.23252 | 0.00570 |
| IL12B | 0.06782 | 0.74973 | -0.23538 | 0.37474 |
| DDR1 | 0.31751 | 0.02881 | 0.04668 | 0.72480 |
| RAC3 | 0.13901 | 0.27836 | 0.05022 | 0.74701 |
| DTD1 | 0.19603 | 0.57416 | 2.28786 | 0.00000 |
| FGF6 | -0.04988 | 0.77256 | -0.00756 | 0.96226 |
| SERPING1 | 0.17274 | 0.02041 | 0.02122 | 0.78023 |
| MRPL24 | 0.23625 | 0.24745 | 0.47081 | 0.00512 |
| TIMP2 | 0.04177 | 0.70776 | -0.01399 | 0.91008 |
| ICOSLG | -0.12505 | 0.14237 | -0.57540 | 0.00157 |
| CD55 | 0.04338 | 0.72463 | -0.06933 | 0.62073 |
| DDX25 | 0.20561 | 0.36024 | 0.09787 | 0.71024 |
| KITLG | -0.24076 | 0.21608 | -0.42816 | 0.02607 |
| PAEP | 0.84083 | 0.01944 | -1.17090 | 0.01314 |
| MFAP4 | -0.07715 | 0.57220 | 0.34907 | 0.00646 |
| KYNU | -0.01507 | 0.93939 | 0.68681 | 0.00094 |
| PRDX5 | -0.07123 | 0.71783 | 1.03901 | 0.00012 |
| TPM3 | -0.05958 | 0.77228 | -0.16340 | 0.40775 |
| TAB2 | 0.10970 | 0.72679 | 0.23145 | 0.39724 |
| ADAMTS13 | -0.20752 | 0.06579 | -0.84416 | 0.00000 |
| TREH | -0.23201 | 0.44874 | -0.22641 | 0.38165 |
| PLXNA4 | 0.15645 | 0.55345 | 0.08713 | 0.74770 |
| ANXA11 | -0.09381 | 0.41030 | 0.30751 | 0.02195 |
| DSC2 | 0.52960 | 0.00230 | 0.10348 | 0.57025 |
| LIFR | 0.11094 | 0.26523 | -0.18294 | 0.14582 |
| CXCL12 | -0.12895 | 0.43873 | -0.06488 | 0.74852 |
| BLVRB | 0.03486 | 0.85720 | 0.83903 | 0.00003 |
| SCIN | 0.25119 | 0.35538 | 0.22680 | 0.40086 |
| C2 | 0.11978 | 0.23442 | -0.41982 | 0.00300 |
| TNFRSF1A | 0.50379 | 0.01782 | 0.36082 | 0.10172 |
| NRXN3 | -0.20803 | 0.14567 | -0.15665 | 0.19391 |
| ADAMTS15 | -0.11988 | 0.58302 | 0.87998 | 0.00395 |
| INPP5D | 0.15971 | 0.51405 | 0.60006 | 0.01204 |
| FGF5 | 0.17331 | 0.19538 | 0.08490 | 0.57326 |
| NGRN | 0.22981 | 0.22366 | 0.49189 | 0.00097 |
| RNF149 | 0.28788 | 0.04909 | 0.22003 | 0.10821 |
| CBLN4 | -0.54022 | 0.00877 | -0.79497 | 0.00226 |
| ATP5PO | 0.35123 | 0.04242 | 0.22590 | 0.15923 |
| ZNF174 | 0.07571 | 0.66811 | 0.27659 | 0.24766 |
| SH3GLB2 | 0.14101 | 0.49690 | 1.10753 | 0.00000 |
| MORF4L2 | 0.18319 | 0.39628 | 0.05478 | 0.80412 |
| TPD52L2 | 0.07388 | 0.81177 | 1.83805 | 0.00000 |
| ARHGAP30 | 0.13173 | 0.57557 | 0.35286 | 0.05568 |

| | | | | |
|---|---|---|---|---|
| PTN | 0.85480 | 0.08049 | 1.09051 | 0.02271 |
| CDC37 | 0.24921 | 0.40762 | 1.83394 | 0.00000 |
| CCL27 | 0.62306 | 0.00016 | -0.00431 | 0.97809 |
| NELL2 | -0.05586 | 0.69427 | -0.15739 | 0.27214 |
| CACNA1H | -0.15055 | 0.59162 | 0.37254 | 0.18752 |
| IL17C | 0.22218 | 0.47394 | 0.42190 | 0.16943 |
| TMEM132A | 0.04759 | 0.87028 | 0.24923 | 0.30668 |
| CDH15 | 0.15806 | 0.41115 | -0.16563 | 0.33604 |
| ADAM8 | -0.11352 | 0.48841 | -0.17450 | 0.28603 |
| EIF4B | 0.19202 | 0.41468 | 2.45061 | 0.00000 |
| KLRF1 | 0.37352 | 0.09194 | 0.10676 | 0.62871 |
| CD14 | 0.37622 | 0.04123 | 0.40662 | 0.04773 |
| NECTIN1 | 0.03954 | 0.72074 | -0.03465 | 0.72865 |
| ATP5IF1 | 0.17107 | 0.65841 | 2.14878 | 0.00000 |
| HTR1A | -0.12018 | 0.74581 | -0.16121 | 0.60402 |
| RAB2B | -0.51485 | 0.02742 | -0.39417 | 0.06020 |
| FNDC1 | -0.27700 | 0.20777 | -0.47928 | 0.01432 |
| CRIP2 | 0.35635 | 0.05272 | -0.27613 | 0.23874 |
| MMP15 | 0.01838 | 0.90145 | 0.14039 | 0.39008 |
| FCGR3B | -0.00196 | 0.98895 | -0.12414 | 0.40822 |
| SELENOP | -0.05851 | 0.61900 | -0.17819 | 0.17118 |
| KLRC1 | 0.12565 | 0.65020 | -0.08711 | 0.77004 |
| LRRC59 | 0.22024 | 0.53717 | 0.81075 | 0.00530 |
| LY75 | -0.03810 | 0.78615 | -0.21410 | 0.18689 |
| GART | -0.06052 | 0.74359 | 0.03641 | 0.85462 |
| SERPINB1 | -0.37699 | 0.35173 | 1.05191 | 0.01769 |
| GPC5 | -0.52548 | 0.02346 | -0.79586 | 0.00016 |
| PALLD | 0.56932 | 0.00668 | -0.18210 | 0.39955 |
| CPQ | 0.05409 | 0.71475 | 0.15021 | 0.24337 |
| CCDC50 | 0.40990 | 0.02588 | 1.36540 | 0.00000 |
| ATP1B2 | -0.13238 | 0.43821 | -0.30415 | 0.07468 |
| SERPINB8 | -0.01422 | 0.97026 | 1.09448 | 0.00634 |
| ECI2 | 0.72939 | 0.03775 | 0.54739 | 0.07933 |
| CNTN3 | -0.21035 | 0.04553 | -0.02385 | 0.86638 |
| CCL24 | -0.11288 | 0.62153 | 0.19100 | 0.40911 |
| CEACAM1 | 0.12257 | 0.31584 | 0.03936 | 0.77761 |
| CBLN1 | 0.03943 | 0.93670 | -0.37747 | 0.46546 |
| LMNB1 | 0.28925 | 0.40107 | -0.07571 | 0.82910 |
| EDIL3 | 0.03640 | 0.81617 | -1.54333 | 0.00000 |
| CD36 | 0.01355 | 0.91890 | -0.56985 | 0.00007 |
| ECSCR | 0.29523 | 0.13087 | 0.13497 | 0.51580 |
| MASP1 | -0.08847 | 0.42240 | -0.12275 | 0.24503 |
| M6PR | 0.07544 | 0.59423 | -0.34836 | 0.01069 |
| PROCR | 0.06670 | 0.72022 | -0.09868 | 0.35988 |
| FAM3C | 0.15321 | 0.23731 | -0.26666 | 0.08908 |
| NCAM1 | -0.26507 | 0.03068 | -0.18563 | 0.17170 |
| NEB | 0.02106 | 0.92924 | -0.04500 | 0.80860 |
| HMGCS1 | -0.02739 | 0.88852 | 0.42999 | 0.10846 |

| | | | | |
|---|---|---|---|---|
| SART1 | 0.58917 | 0.15332 | 0.95556 | 0.01373 |
| SYNGAP1 | -0.05538 | 0.82864 | -0.12273 | 0.63552 |
| SIAE | 0.03561 | 0.84485 | -0.57990 | 0.00081 |
| TOR1AIP1 | -0.08996 | 0.84997 | 1.90498 | 0.00003 |
| GIMAP7 | 0.06955 | 0.73820 | 0.20380 | 0.28589 |
| GDF15 | 0.94431 | 0.00182 | 1.17266 | 0.00016 |
| AKR1B10 | 0.41511 | 0.33256 | 0.45416 | 0.29296 |
| CNST | 0.66082 | 0.03089 | 1.04594 | 0.00011 |
| TPP1 | 0.10680 | 0.48560 | -0.28931 | 0.05604 |
| BOC | 0.02509 | 0.81287 | -0.41106 | 0.00199 |
| KLK10 | -0.17368 | 0.36921 | -0.26781 | 0.11137 |
| IL25 | 0.58527 | 0.00955 | 0.19802 | 0.29977 |
| GZMH | 1.26414 | 0.00897 | 0.51275 | 0.14485 |
| TRIAP1 | 0.56523 | 0.03402 | 1.44770 | 0.00000 |
| VSIG2 | 0.45345 | 0.10102 | 0.10743 | 0.62911 |
| IDI2 | 0.08376 | 0.69091 | 0.09248 | 0.68610 |
| CD5L | 0.35129 | 0.02570 | 0.24589 | 0.11528 |
| CDH17 | 0.00988 | 0.97508 | -0.37504 | 0.14725 |
| HJV | 0.11503 | 0.45564 | -0.16209 | 0.35524 |
| TBC1D5 | -0.66426 | 0.05481 | -0.28087 | 0.40216 |
| VMO1 | 0.52096 | 0.00657 | 0.08491 | 0.72468 |
| HBQ1 | 0.09822 | 0.62860 | 1.31510 | 0.00000 |
| REG3G | 0.30430 | 0.17066 | -0.05631 | 0.79489 |
| RBP7 | 0.45254 | 0.24661 | 1.13848 | 0.00187 |
| PTGES2 | 0.55658 | 0.07508 | 0.68218 | 0.02124 |
| FCRL5 | 0.04590 | 0.82174 | -0.31792 | 0.09598 |
| CA12 | 0.33803 | 0.07803 | -0.13742 | 0.52853 |
| APCS | -0.03482 | 0.80545 | -0.05567 | 0.72022 |
| PROS1 | -0.02520 | 0.83164 | -0.34980 | 0.00035 |
| CSTB | 0.14003 | 0.50027 | 0.89941 | 0.00003 |
| VEGFD | 0.13662 | 0.43097 | -0.35645 | 0.09823 |
| MERTK | 0.25213 | 0.04797 | -0.00690 | 0.97008 |
| PPP2R5A | -0.42743 | 0.34725 | 2.76423 | 0.00000 |
| PIK3IP1 | 0.38958 | 0.01388 | 0.23802 | 0.13600 |
| HIF1A | -0.32221 | 0.34797 | 0.25407 | 0.41027 |
| PTTG1 | 1.20231 | 0.00954 | 0.96766 | 0.02919 |
| NPHS1 | -0.23158 | 0.06690 | -0.46114 | 0.00173 |
| SUGP1 | -0.36668 | 0.16260 | 0.42677 | 0.09051 |
| ISLR2 | -0.20436 | 0.13877 | -0.30586 | 0.04695 |
| CSF2 | -0.04860 | 0.82658 | -0.13526 | 0.62171 |
| GPR37 | 0.29817 | 0.36518 | 1.61632 | 0.00002 |
| CNTN5 | -0.47347 | 0.00131 | -0.46970 | 0.00166 |
| HNF1A | -0.00959 | 0.96997 | 0.35445 | 0.16267 |
| NENF | 0.08865 | 0.73976 | -0.23146 | 0.42934 |
| GALNT10 | 0.08978 | 0.36887 | -0.33162 | 0.04042 |
| DPY30 | 0.42988 | 0.21391 | 0.82684 | 0.01403 |
| CHAD | -0.74028 | 0.02303 | -0.46123 | 0.12149 |
| CEMIP2 | -0.01704 | 0.90294 | -0.12916 | 0.34986 |

| | | | | |
|---|---|---|---|---|
| IFNGR2 | -0.03780 | 0.83635 | -0.23976 | 0.25287 |
| BRME1 | 0.09988 | 0.58340 | 0.17745 | 0.30781 |
| IL6ST | 0.06822 | 0.44132 | -0.07342 | 0.48628 |
| FCRLB | 0.52111 | 0.03760 | 0.07935 | 0.77040 |
| APOA4 | -0.06283 | 0.76536 | -0.67907 | 0.00136 |
| CDA | -0.22521 | 0.35890 | 0.64602 | 0.00768 |
| ABHD14B | -0.28495 | 0.34045 | 0.03050 | 0.91560 |
| REG1B | 0.63547 | 0.02533 | 0.02071 | 0.94213 |
| CD200R1 | -0.09171 | 0.46560 | -0.25126 | 0.15196 |
| SAA4 | 0.07727 | 0.39475 | -0.22539 | 0.00412 |
| DNAJC9 | 0.49356 | 0.09286 | 2.56753 | 0.00000 |
| CDHR2 | 0.30005 | 0.25335 | 0.52464 | 0.06665 |
| RPE | -0.21399 | 0.41017 | 1.13974 | 0.00000 |
| PPY | 0.92072 | 0.02388 | 0.27666 | 0.41889 |
| TGFBR1 | 0.51658 | 0.04549 | 0.06911 | 0.77805 |
| DNAJA2 | -0.49272 | 0.06262 | -0.00063 | 0.99838 |
| NOS3 | 0.72892 | 0.00703 | 0.65304 | 0.01385 |
| CD274 | 0.36229 | 0.13913 | 0.13755 | 0.57465 |
| CMC1 | 0.29267 | 0.38314 | 1.55945 | 0.00000 |
| CA6 | -0.90942 | 0.00087 | -1.12010 | 0.00003 |
| CTRB1 | 0.29382 | 0.25189 | -0.24456 | 0.34132 |
| PADI4 | -0.25746 | 0.58179 | 0.11474 | 0.81417 |
| TMCO5A | 0.47160 | 0.03076 | 0.90314 | 0.00592 |
| DKKL1 | 0.41715 | 0.13115 | -0.39369 | 0.10802 |
| CHP1 | 0.59068 | 0.20166 | 0.32628 | 0.48755 |
| MUCL3 | 0.46256 | 0.08035 | -0.15850 | 0.47941 |
| SLAMF1 | 0.06449 | 0.73763 | -0.21858 | 0.25368 |
| BANK1 | -0.05689 | 0.85043 | 0.60394 | 0.08726 |
| PKD2 | 0.15797 | 0.65646 | -0.28650 | 0.41074 |
| CELSR2 | -0.43121 | 0.01347 | 0.19268 | 0.28924 |
| CX3CL1 | 0.11793 | 0.53322 | 0.11180 | 0.55978 |
| INSR | 0.12409 | 0.39524 | 0.06427 | 0.61145 |
| C8B | -0.14326 | 0.12487 | -0.20898 | 0.03055 |
| RALY | 0.33906 | 0.40244 | 0.38748 | 0.31414 |
| NEXN | 0.17434 | 0.59690 | 1.85486 | 0.00000 |
| BSND | 0.53750 | 0.05272 | -0.77104 | 0.00829 |
| ACOX1 | 0.30225 | 0.37498 | 1.21862 | 0.00034 |
| AHSP | 0.00634 | 0.98155 | 1.69813 | 0.00000 |
| CXCL14 | 0.36994 | 0.02363 | 0.26022 | 0.11966 |
| HNRNPUL1 | 0.24840 | 0.36140 | 1.04593 | 0.00013 |
| NFATC3 | 0.28143 | 0.24712 | 0.17070 | 0.34221 |
| PAG1 | -0.15248 | 0.66890 | 0.66744 | 0.05510 |
| DYNLT1 | 0.21154 | 0.50614 | 0.42851 | 0.15963 |
| COMMD1 | -0.15325 | 0.54516 | 0.81210 | 0.00110 |
| BSG | 0.18344 | 0.07994 | -0.30965 | 0.02848 |
| PAK4 | 0.22032 | 0.46091 | 1.04771 | 0.00030 |
| ARMCX2 | 0.18827 | 0.34207 | -0.11709 | 0.64175 |
| CD46 | 0.00345 | 0.98063 | -0.10306 | 0.49203 |

| | | | | |
|---|---|---|---|---|
| CDH6 | 0.33362 | 0.06248 | -0.37931 | 0.01038 |
| PILRA | 0.30623 | 0.05329 | 0.20914 | 0.23792 |
| C1R | -0.02714 | 0.65524 | -0.09009 | 0.15235 |
| MMP9 | 0.85726 | 0.00209 | -0.06628 | 0.80429 |
| BNIP2 | 0.09617 | 0.73928 | 0.52873 | 0.06021 |
| HAVCR2 | 0.52028 | 0.01753 | 0.28889 | 0.21389 |
| VTCN1 | 0.26518 | 0.29737 | -0.05404 | 0.83688 |
| ENTPD2 | 0.52397 | 0.09842 | 0.03889 | 0.84865 |
| MAD1L1 | 0.24115 | 0.50714 | 0.84541 | 0.02468 |
| GTF2IRD1 | 0.07626 | 0.72005 | -0.15197 | 0.62997 |
| AFM | -0.28834 | 0.03671 | -0.25651 | 0.04224 |
| FOLH1 | 0.61868 | 0.06232 | 0.47530 | 0.10011 |
| DCBLD2 | 0.49376 | 0.00045 | 0.22766 | 0.15624 |
| CBLIF | -0.30808 | 0.31484 | -0.66563 | 0.04310 |
| RABEP1 | -0.45076 | 0.11624 | 0.20997 | 0.41088 |
| CCND2 | -0.42796 | 0.36631 | -0.08851 | 0.77653 |
| GZMA | 0.18000 | 0.35949 | 0.07979 | 0.69927 |
| PADI2 | 0.34387 | 0.26205 | 0.58304 | 0.09967 |
| IL22RA1 | 0.13691 | 0.50501 | -0.31885 | 0.04991 |
| DNM1 | 0.40380 | 0.29256 | 2.85623 | 0.00000 |
| GAS2 | 0.35025 | 0.29775 | 0.36712 | 0.19631 |
| LRG1 | 0.63010 | 0.00140 | 0.36870 | 0.03735 |
| SNX5 | -0.18679 | 0.29928 | 0.51589 | 0.00092 |
| CASP4 | -0.03735 | 0.94036 | 0.31772 | 0.51321 |
| CLEC1B | 0.10717 | 0.72880 | -0.77639 | 0.00415 |
| LCP1 | -0.34432 | 0.05457 | 0.31953 | 0.03166 |
| NHLRC3 | 0.16881 | 0.20832 | -0.25317 | 0.02420 |
| C4BPB | 0.03889 | 0.83396 | 0.14233 | 0.46120 |
| HARS1 | 0.06615 | 0.80301 | 1.75592 | 0.00000 |
| ZBTB16 | -0.02648 | 0.92397 | 0.37969 | 0.13271 |
| ENPP2 | -0.05894 | 0.59709 | 0.01165 | 0.92042 |
| GALNT3 | -0.12436 | 0.50428 | -0.35174 | 0.06851 |
| SPART | -0.62160 | 0.04349 | -0.35059 | 0.22835 |
| VASH1 | 0.27892 | 0.37790 | 1.36599 | 0.00001 |
| LCN15 | 0.46731 | 0.08290 | 0.59642 | 0.00384 |
| COCH | 0.23006 | 0.10500 | -0.09134 | 0.54782 |
| LATS1 | 0.38776 | 0.30896 | 1.98742 | 0.00000 |
| RPS10 | 0.01411 | 0.94415 | 0.17060 | 0.39434 |
| DCXR | -0.09488 | 0.70800 | 0.91040 | 0.00006 |
| TNIP1 | -0.35462 | 0.30514 | 0.02214 | 0.94986 |
| ITIH3 | 0.64595 | 0.00002 | 0.15819 | 0.28538 |
| NPPC | -0.55808 | 0.01031 | -0.40308 | 0.13896 |
| ITGB6 | -0.06452 | 0.72547 | -0.53182 | 0.00294 |
| UROS | -0.19669 | 0.48311 | 0.79209 | 0.00554 |
| MECR | 0.65256 | 0.11491 | 2.14546 | 0.00000 |
| HGFAC | 0.11421 | 0.37388 | -0.13736 | 0.24940 |
| IL7R | -0.12910 | 0.39951 | -0.22836 | 0.16416 |
| RTN4R | 0.05333 | 0.67874 | -0.13123 | 0.43847 |

| | | | | |
|---|---|---|---|---|
| ANKMY2 | -0.14391 | 0.63709 | 0.83310 | 0.00417 |
| TMOD4 | -0.17326 | 0.51259 | 0.31982 | 0.13363 |
| CSF1R | 0.07508 | 0.66725 | 0.04397 | 0.77099 |
| SPINT1 | 0.25655 | 0.17818 | -0.03820 | 0.81937 |
| MTPN | -0.29095 | 0.20143 | -0.70607 | 0.00001 |
| DEFB104A; DEFB104E | 0.07521 | 0.68135 | -0.05076 | 0.84090 |
| FLT1 | 0.12167 | 0.44179 | 0.24571 | 0.19758 |
| RNF41 | -0.56354 | 0.01933 | 0.73886 | 0.01365 |
| APOF | -0.04398 | 0.73063 | 0.13951 | 0.23295 |
| PRG2 | 0.53237 | 0.01671 | 0.26866 | 0.22604 |
| GORASP2 | 0.18139 | 0.46842 | 0.99874 | 0.00003 |
| CDH5 | -0.05083 | 0.66150 | -0.24923 | 0.06597 |
| CA13 | -0.12734 | 0.72356 | 0.92850 | 0.01052 |
| PECAM1 | 0.17352 | 0.20670 | -0.08674 | 0.53131 |
| SLITRK6 | -0.01005 | 0.93638 | -0.21295 | 0.13009 |
| TGM2 | 0.20215 | 0.40765 | 2.61904 | 0.00000 |
| TJP3 | 0.22840 | 0.08998 | 0.08358 | 0.68929 |
| PIKFYVE | 0.14559 | 0.63922 | 0.75908 | 0.00555 |
| PRCP | -0.05454 | 0.65087 | 0.08560 | 0.48105 |
| GUK1 | 0.25504 | 0.19075 | -0.20745 | 0.23247 |
| GBP2 | -0.65788 | 0.06293 | -0.00895 | 0.97904 |
| VPS37A | -0.15376 | 0.57326 | 0.62773 | 0.01033 |
| ATRAID | 0.31887 | 0.05969 | 0.04766 | 0.75994 |
| RHOC | 0.72345 | 0.07491 | 2.16282 | 0.00000 |
| PER3 | 0.15092 | 0.78077 | 2.16723 | 0.00003 |
| GLYR1 | 0.09939 | 0.75537 | 0.16024 | 0.60456 |
| SOST | 0.04070 | 0.77494 | 0.36776 | 0.04664 |
| CLIP2 | -0.01807 | 0.95991 | 1.44919 | 0.00027 |
| CLEC4A | -0.43856 | 0.00162 | -0.29024 | 0.05373 |
| PCYT2 | 0.36241 | 0.21606 | 2.07533 | 0.00000 |
| PRKAG3 | 0.32733 | 0.40217 | 0.79309 | 0.05685 |
| TPSD1 | 0.04834 | 0.74018 | -0.00094 | 0.99555 |
| AMIGO2 | -0.01431 | 0.84257 | -0.23699 | 0.19663 |
| ADGRG2 | -0.10663 | 0.44271 | -0.44314 | 0.00181 |
| PSMA1 | 0.00738 | 0.97447 | -0.14603 | 0.55017 |
| EDDM3B | -0.25216 | 0.21513 | -0.08896 | 0.57552 |
| HMBS | 0.06387 | 0.67965 | 1.34440 | 0.00000 |
| CLEC2L | 0.54953 | 0.00703 | 0.33469 | 0.13030 |
| TNXB | -0.31806 | 0.00430 | -0.41746 | 0.00177 |
| LILRA6 | 0.48303 | 0.02771 | -0.05835 | 0.79055 |
| BRSK2 | -0.06639 | 0.80133 | -0.16784 | 0.40464 |
| REST | 0.30520 | 0.19633 | 0.19776 | 0.45204 |
| AKT3 | -0.02889 | 0.91138 | 0.31668 | 0.25646 |
| CIAPIN1 | -0.14115 | 0.57462 | 0.33465 | 0.16002 |
| CTSB | 0.40857 | 0.10465 | 0.40769 | 0.10339 |
| ACADSB | 0.24023 | 0.54121 | 1.57176 | 0.00003 |
| COPE | 0.10730 | 0.63494 | 0.23845 | 0.42767 |
| FABP9 | -0.32899 | 0.08664 | -0.62495 | 0.00136 |

| | | | | |
|---|---|---|---|---|
| CTSL | 0.33213 | 0.06532 | 0.28386 | 0.13779 |
| CASP9 | -0.13314 | 0.74972 | 0.14770 | 0.69655 |
| FGF19 | 0.23791 | 0.35405 | -0.52893 | 0.06519 |
| NGF | 0.07216 | 0.23466 | -0.04835 | 0.21119 |
| LRP2BP | -0.12243 | 0.69377 | 0.21138 | 0.59202 |
| AMOT | -0.28391 | 0.11368 | -0.57431 | 0.00223 |
| USP25 | 0.04035 | 0.86239 | 0.32964 | 0.14748 |
| NDUFS6 | -0.05527 | 0.88000 | 0.65449 | 0.08774 |
| KIT | -0.36643 | 0.00487 | -0.37156 | 0.00743 |
| CXCL5 | -0.10417 | 0.78575 | -0.78189 | 0.01713 |
| DAPP1 | 0.19695 | 0.66142 | 3.77985 | 0.00000 |
| RETN | -0.00496 | 0.98661 | 0.41242 | 0.11595 |
| IGFBP2 | 1.32805 | 0.00000 | 0.66070 | 0.00749 |
| IL1RL2 | -0.16845 | 0.21408 | 0.13819 | 0.43297 |
| MAMDC4 | -0.15473 | 0.49294 | -0.86495 | 0.00105 |
| BOLA2B; BOLA2 | -0.02000 | 0.90481 | 1.17527 | 0.00000 |
| ITPRIP | 0.20747 | 0.09948 | -0.17729 | 0.18237 |
| MAPK13 | 0.27499 | 0.47556 | 0.62510 | 0.06992 |
| NOTCH2 | 0.01538 | 0.85430 | -0.17511 | 0.06577 |
| PGLYRP2 | -0.13975 | 0.38587 | -0.12621 | 0.29107 |
| ADAMTSL2 | 0.05466 | 0.70796 | -0.04560 | 0.71702 |
| CIRBP | 0.15172 | 0.60784 | 1.65281 | 0.00000 |
| CHIT1 | 0.50944 | 0.11845 | 0.74645 | 0.05292 |
| SLURP1 | -0.03295 | 0.82286 | -0.06192 | 0.62137 |
| RNASEH2A | -0.45932 | 0.09672 | 0.28404 | 0.29924 |
| HLA-E | 0.17663 | 0.06033 | 0.25288 | 0.02388 |
| SNAP29 | -0.19014 | 0.55051 | 0.14021 | 0.63253 |
| ADGRB3 | -0.02403 | 0.89080 | -0.11450 | 0.57480 |
| AZU1 | -0.07175 | 0.84194 | -1.13127 | 0.00642 |
| PPBP | 0.33097 | 0.19682 | -1.02302 | 0.00001 |
| PPME1 | 0.16691 | 0.43401 | 1.58478 | 0.00000 |
| MOG | 0.34569 | 0.03413 | -0.00347 | 0.97915 |
| CHRDL2 | 0.03145 | 0.86268 | -0.45470 | 0.04694 |
| ALDH5A1 | 0.24737 | 0.49636 | 1.12109 | 0.00032 |
| CAMLG | 0.41045 | 0.16526 | 0.69355 | 0.01084 |
| SUGT1 | 0.02251 | 0.93331 | 1.62998 | 0.00000 |
| CNDP1 | -0.44160 | 0.03879 | -1.45719 | 0.00000 |
| VTI1A | -0.36500 | 0.20530 | -0.18344 | 0.48656 |
| CLEC4G | 0.07036 | 0.60651 | -0.02572 | 0.86281 |
| ESPL1 | 0.41194 | 0.07087 | -0.02547 | 0.91388 |
| CLEC4C | -0.15296 | 0.51012 | -0.60093 | 0.01587 |
| SIGLEC7 | 0.16280 | 0.14557 | 0.05784 | 0.64604 |
| PRR5 | 0.09276 | 0.61542 | 0.62707 | 0.00132 |
| CDK1 | 0.73133 | 0.00133 | 0.29995 | 0.18710 |
| BAIAP2 | 0.19076 | 0.58599 | 0.57672 | 0.04463 |
| EIF5A | 0.09495 | 0.40699 | -0.21385 | 0.03145 |
| CCN5 | 0.14580 | 0.41565 | 0.30995 | 0.10040 |
| CYB5A | 0.37430 | 0.25972 | 0.57636 | 0.05486 |

| | | | | |
|---|---|---|---|---|
| PDE1C | 0.51822 | 0.01598 | 0.12281 | 0.62591 |
| AGER | -0.16245 | 0.28228 | -0.06974 | 0.65081 |
| DGKZ | -0.43588 | 0.01843 | -0.06607 | 0.69053 |
| PFKFB2 | -0.04686 | 0.88635 | 1.38121 | 0.00060 |
| RANGAP1 | -0.13194 | 0.36029 | 0.00226 | 0.98793 |
| RNF31 | -0.53179 | 0.03945 | 0.02628 | 0.92593 |
| PZP | 0.33079 | 0.26450 | 0.05891 | 0.83879 |
| FMR1 | -0.62478 | 0.12243 | 0.86927 | 0.00723 |
| CD33 | -0.18493 | 0.40142 | 0.19063 | 0.36733 |
| PTPRB | 0.00950 | 0.93090 | 0.36308 | 0.00140 |
| TNFRSF10C | 0.10600 | 0.48073 | -0.12758 | 0.43733 |
| LETM1 | 0.99535 | 0.00516 | 1.33388 | 0.00000 |
| TSC22D1 | 0.36423 | 0.10328 | 0.84352 | 0.00014 |
| PAPPA | 0.09633 | 0.73019 | -0.04198 | 0.88563 |
| SLC9A3R2 | 0.23667 | 0.20915 | 0.25184 | 0.13589 |
| GAL | -0.15917 | 0.53875 | -1.54599 | 0.00000 |
| AGR3 | 0.00382 | 0.99187 | -0.53902 | 0.13620 |
| ZFYVE19 | -0.10520 | 0.74785 | 2.30845 | 0.00000 |
| ZCCHC8 | 0.35219 | 0.35887 | 0.68183 | 0.06412 |
| ESM1 | 0.26322 | 0.22395 | 0.91059 | 0.00001 |
| KIF1C | 0.42390 | 0.10426 | 0.58928 | 0.01550 |
| PDCD6 | -0.02828 | 0.87578 | 0.04848 | 0.78094 |
| SMARCA2 | -0.00384 | 0.99018 | -0.27754 | 0.38466 |
| RRP15 | 0.19937 | 0.75050 | -0.03761 | 0.95367 |
| ACTN2 | 0.25642 | 0.45278 | 0.61469 | 0.04358 |
| GADD45B | 0.29744 | 0.53125 | 0.25864 | 0.55498 |
| TFPI | 0.41332 | 0.00616 | 0.36085 | 0.04329 |
| LDLRAP1 | 0.42281 | 0.28663 | 2.84144 | 0.00000 |
| EFNA4 | 0.20152 | 0.36745 | -0.14826 | 0.48392 |
| SLC16A1 | 0.34412 | 0.11328 | -0.24989 | 0.29489 |
| AGBL2 | 0.32231 | 0.55141 | 0.52825 | 0.33064 |
| SLC39A5 | 0.41955 | 0.03707 | 0.60632 | 0.00258 |
| LPCAT2 | -0.25197 | 0.37623 | 0.13626 | 0.68361 |
| NMI | 0.14951 | 0.57996 | 1.05301 | 0.00031 |
| JPT2 | 0.37833 | 0.15373 | 1.57037 | 0.00000 |
| PTPN1 | -0.36303 | 0.24211 | -0.22447 | 0.43094 |
| LTBP2 | 0.60012 | 0.00419 | 0.30543 | 0.16558 |
| OTUD6B | -0.08596 | 0.70292 | 0.57446 | 0.00953 |
| LY6D | 0.13090 | 0.47552 | 0.03497 | 0.83420 |
| VASN | -0.01304 | 0.89797 | -0.06468 | 0.56010 |
| LILRB2 | 0.24970 | 0.07166 | 0.02414 | 0.86765 |
| TLR2 | 0.05851 | 0.82965 | -0.67709 | 0.02061 |
| EBAG9 | 0.35046 | 0.27821 | 1.98975 | 0.00000 |
| ATP6AP2 | 0.20282 | 0.17183 | -0.41834 | 0.00806 |
| EIF4E | 0.28199 | 0.43074 | 3.23186 | 0.00000 |
| CCL26 | -0.09370 | 0.77008 | 0.00485 | 0.98692 |
| PHYKPL | 0.06625 | 0.73675 | 0.66241 | 0.00024 |
| GSAP | 0.09244 | 0.71356 | 0.11516 | 0.71735 |

| | | | | |
|---|---|---|---|---|
| LACTB2 | 0.05976 | 0.81308 | 1.04460 | 0.00001 |
| SCARB2 | 0.25550 | 0.25526 | 0.33772 | 0.13895 |
| EIF1AX | 0.11785 | 0.73582 | 0.02780 | 0.91744 |
| SMPDL3A | -0.25746 | 0.23696 | 0.22783 | 0.25948 |
| IL32 | -0.01530 | 0.93328 | -0.42951 | 0.04865 |
| LTA4H | -0.42851 | 0.03645 | 0.33449 | 0.06791 |
| CRYGD | 0.06504 | 0.84617 | 1.14448 | 0.00015 |
| DNAJB2 | -0.24472 | 0.27176 | 0.58133 | 0.00686 |
| EGFLAM | 0.12559 | 0.53819 | -0.03868 | 0.83612 |
| CRISP3 | -0.07119 | 0.40190 | -0.26565 | 0.00120 |
| TMED4 | 0.27155 | 0.29792 | 0.40500 | 0.18542 |
| ACAN | -0.03608 | 0.74798 | -0.02620 | 0.78076 |
| NEK7 | -0.10627 | 0.73985 | 1.00709 | 0.00092 |
| GET3 | 0.15659 | 0.51807 | 0.38701 | 0.03823 |
| TNFRSF9 | 0.21782 | 0.22898 | 0.07502 | 0.71243 |
| PAXX | 0.02141 | 0.92629 | 1.02060 | 0.00000 |
| CLEC12A | -0.00382 | 0.98397 | -0.19403 | 0.29646 |
| IMPA1 | -0.20639 | 0.42389 | 1.40005 | 0.00000 |
| NUBP1 | -0.25137 | 0.68755 | -0.60248 | 0.33731 |
| A1BG | 0.14235 | 0.11839 | 0.14327 | 0.10720 |
| BCL7A | -0.03900 | 0.91641 | 0.32176 | 0.37169 |
| TREML1 | 0.16414 | 0.60062 | -1.19193 | 0.00007 |
| SERPINA11 | 0.10334 | 0.46633 | 0.14325 | 0.38219 |
| CORO1A | -0.57706 | 0.18288 | 0.46711 | 0.26062 |
| CPM | -0.15230 | 0.29459 | -0.05036 | 0.73925 |
| PTPRS | -0.14921 | 0.07789 | -0.27460 | 0.01283 |
| GSTA1 | -0.47924 | 0.32431 | 0.94097 | 0.03326 |
| S100P | 0.37221 | 0.32660 | 0.61765 | 0.09355 |
| PAGR1 | 0.16040 | 0.56552 | 0.91431 | 0.00129 |
| ERBIN | -0.02544 | 0.93788 | 1.08165 | 0.00173 |
| KLK14 | -0.33845 | 0.10327 | -0.42153 | 0.08857 |
| MAGED1 | 0.38285 | 0.04901 | 0.35832 | 0.03645 |
| FLT3 | -0.33727 | 0.02775 | -0.96402 | 0.00000 |
| CCL28 | 0.44222 | 0.11931 | -0.10886 | 0.67721 |
| LRTM1 | -0.43435 | 0.19764 | -0.38024 | 0.38564 |
| ITIH5 | -0.14081 | 0.60364 | -0.11923 | 0.61642 |
| PTH1R | 0.11987 | 0.27077 | -0.04020 | 0.74652 |
| NCS1 | 0.16130 | 0.31790 | -0.31890 | 0.19054 |
| NPC2 | 0.06609 | 0.65938 | 0.23769 | 0.09696 |
| EDAR | -0.49567 | 0.10503 | -1.32175 | 0.00002 |
| KLRB1 | 0.26981 | 0.05666 | -0.00674 | 0.97457 |
| FST | 0.39359 | 0.04300 | 0.40506 | 0.09008 |
| PDGFB | 0.21961 | 0.39639 | -0.72618 | 0.00254 |
| REPS1 | 0.27015 | 0.20222 | 0.75877 | 0.00109 |
| IL20RB | 0.01822 | 0.85959 | -0.17378 | 0.07050 |
| BTN3A2 | 0.10258 | 0.48632 | -0.16720 | 0.35656 |
| RASA1 | 0.14556 | 0.14071 | -0.07712 | 0.41620 |
| DNMBP | -0.31221 | 0.35059 | -0.48147 | 0.16249 |

| | | | | |
|---|---|---|---|---|
| IL2RA | 0.39434 | 0.05282 | 0.04499 | 0.83416 |
| PCBP2 | 0.27422 | 0.35099 | 2.17909 | 0.00000 |
| POLR2F | 0.34450 | 0.30517 | 0.90559 | 0.00546 |
| CD300LG | 0.05543 | 0.66937 | 0.10333 | 0.43872 |
| WAS | -0.13403 | 0.73837 | -0.52882 | 0.15709 |
| COL15A1 | -0.35804 | 0.01288 | -0.11637 | 0.36929 |
| TFRC | 0.34186 | 0.07928 | 1.13166 | 0.00000 |
| SELL | -0.27091 | 0.00614 | -0.30485 | 0.00234 |
| APOA2 | 0.28258 | 0.32627 | -1.90699 | 0.00000 |
| CD40LG | 0.66315 | 0.01203 | -0.90930 | 0.00144 |
| BAG4 | -0.10537 | 0.71654 | 0.89034 | 0.00113 |
| SIRT1 | 0.01103 | 0.94859 | 0.06502 | 0.74272 |
| ALCAM | 0.07166 | 0.49092 | -0.01689 | 0.89602 |
| CCL25 | 0.26255 | 0.12502 | -0.43810 | 0.07577 |
| INHBB | 0.60888 | 0.00963 | 0.40514 | 0.06672 |
| GCLM | -0.02045 | 0.92100 | 0.62831 | 0.00188 |
| LRRC25 | 0.16707 | 0.32112 | 0.00733 | 0.97253 |
| OGA | -0.15765 | 0.49214 | 0.74541 | 0.00152 |
| GABARAP | 0.13270 | 0.59965 | -0.16077 | 0.45461 |
| CCL22 | -0.16028 | 0.43326 | -0.32044 | 0.09343 |
| ASPN | -0.14472 | 0.40932 | -0.60112 | 0.00007 |
| DDX58 | -0.76664 | 0.03809 | 0.54594 | 0.15080 |
| PCDH7 | 0.01736 | 0.89839 | -0.11011 | 0.25997 |
| PPM1B | -0.13589 | 0.57200 | 0.88591 | 0.00007 |
| DNPH1 | 0.12527 | 0.49944 | 1.74590 | 0.00000 |
| APP | 0.20077 | 0.28947 | -1.06037 | 0.00000 |
| KHDC3L | 0.31416 | 0.24250 | -0.52301 | 0.05142 |
| STIP1 | -0.06408 | 0.81138 | 1.70133 | 0.00000 |
| SPINK5 | -0.19900 | 0.05367 | -0.16834 | 0.13821 |
| C1S | -0.04440 | 0.55759 | -0.19783 | 0.04535 |
| C19orf12 | 0.21838 | 0.51941 | 0.44528 | 0.13061 |
| TWF2 | -0.06937 | 0.83472 | 1.43655 | 0.00000 |
| ATOX1 | -0.01269 | 0.95035 | 0.63825 | 0.00555 |
| CRTAP | 0.30170 | 0.13473 | 0.61988 | 0.00054 |
| CD99 | -0.07186 | 0.46897 | -0.10700 | 0.35959 |
| MTUS1 | 0.35829 | 0.37733 | 0.74344 | 0.01685 |
| PLXDC1 | -0.25278 | 0.08058 | -0.19219 | 0.16536 |
| PON1 | -0.37689 | 0.00528 | -0.22485 | 0.05857 |
| EFEMP1 | 0.54795 | 0.00241 | 0.18719 | 0.28224 |
| CHGB | 0.47839 | 0.01781 | 0.35324 | 0.07328 |
| RNF43 | 0.14934 | 0.46317 | -0.05371 | 0.77549 |
| WIF1 | -0.00088 | 0.99446 | -0.04641 | 0.85444 |
| VTA1 | 0.24812 | 0.37929 | 2.07843 | 0.00000 |
| CENPJ | -0.06661 | 0.85247 | 0.23092 | 0.63288 |
| GDNF | 0.47876 | 0.01671 | 0.74866 | 0.00024 |
| BIN2 | -0.40469 | 0.30860 | 0.01385 | 0.96968 |
| TCTN3 | 0.07939 | 0.50389 | -0.14749 | 0.18660 |
| TSPYL1 | 0.05901 | 0.74272 | 0.50810 | 0.00122 |

| | | | | |
|---|---|---|---|---|
| FAM171B | 0.12284 | 0.16272 | -0.09232 | 0.34530 |
| FOXO1 | 0.18565 | 0.50889 | 1.29142 | 0.00002 |
| CCL7 | -0.89097 | 0.02011 | -0.10257 | 0.76890 |
| TOMM20 | 0.40287 | 0.14649 | 0.79926 | 0.00082 |
| AMBN | -0.06676 | 0.58149 | 0.03302 | 0.72568 |
| ERP29 | 0.25247 | 0.52788 | 1.97793 | 0.00000 |
| DNAJB14 | -0.18854 | 0.57010 | 0.62447 | 0.04164 |
| STOML2 | 0.36983 | 0.28507 | 0.01066 | 0.97406 |
| IGDCC4 | -0.29678 | 0.03018 | -0.39003 | 0.00211 |
| OBP2B | -0.24120 | 0.36427 | -0.45224 | 0.06950 |
| NEFL | 1.11864 | 0.00257 | 0.80773 | 0.02203 |
| P4HB | 0.13197 | 0.42105 | 0.27899 | 0.07312 |
| CEP350 | 0.39724 | 0.20435 | 0.04470 | 0.87113 |
| ANKRA2 | 0.50743 | 0.05451 | -0.20119 | 0.47838 |
| TFPI2 | 0.34821 | 0.16458 | 0.72278 | 0.00033 |
| GZMB | 0.71353 | 0.03833 | 0.44827 | 0.16465 |
| SCN3B | 0.19608 | 0.30918 | -0.00748 | 0.97243 |
| HRC | -0.06770 | 0.61227 | 0.02459 | 0.85098 |
| BHMT2 | 0.11655 | 0.49706 | -0.01137 | 0.93316 |
| ITIH4 | 0.23279 | 0.06962 | -0.18128 | 0.10902 |
| OXCT1 | 0.84293 | 0.00361 | 1.56202 | 0.00000 |
| EHD3 | 0.26650 | 0.48574 | 2.18771 | 0.00000 |
| RNASE6 | 0.22830 | 0.12735 | 0.14312 | 0.25366 |
| SFRP1 | 0.44270 | 0.27925 | 1.07549 | 0.01594 |
| ARHGEF1 | -0.13470 | 0.64179 | 0.98543 | 0.00070 |
| IFNW1 | 0.27924 | 0.11329 | 0.09862 | 0.54057 |
| LRIG3 | 0.07531 | 0.44099 | -0.08140 | 0.35745 |
| AMN | 0.13086 | 0.51153 | 0.12468 | 0.51602 |
| SPINT2 | 0.18247 | 0.23649 | 0.07255 | 0.66555 |
| COL6A3 | 0.26350 | 0.18960 | -0.06614 | 0.74253 |
| CDC27 | 0.54323 | 0.04978 | 0.65712 | 0.01157 |
| APLP1 | -0.39994 | 0.03825 | -0.56391 | 0.00149 |
| SNAP23 | 0.00682 | 0.98268 | 1.10713 | 0.00214 |
| LRFN2 | -0.20776 | 0.24681 | -0.45352 | 0.02183 |
| CNTN1 | -0.22282 | 0.11795 | -0.38102 | 0.01303 |
| CELA2A | 0.06249 | 0.82636 | -0.20542 | 0.38730 |
| RCC1 | 0.08125 | 0.79262 | 0.64160 | 0.02712 |
| HSD17B14 | 0.54242 | 0.08863 | 0.60757 | 0.05197 |
| TMEM25 | -0.14327 | 0.38157 | -0.00922 | 0.95535 |
| CD58 | -0.08392 | 0.34314 | -0.11762 | 0.29971 |
| GATA3 | 0.34344 | 0.02449 | -0.07066 | 0.69665 |
| TNFRSF11B | 0.31572 | 0.04877 | 0.30360 | 0.07944 |
| MDK | 0.91781 | 0.01313 | 0.91909 | 0.00969 |
| GAMT | 0.09159 | 0.44420 | 0.31851 | 0.01084 |
| IST1 | 0.08654 | 0.76798 | 1.06519 | 0.00042 |
| PARD3 | 0.49962 | 0.22572 | 1.96204 | 0.00008 |
| EPO | 0.68912 | 0.16656 | 0.59216 | 0.17600 |
| NPY | -0.42433 | 0.06259 | 0.45974 | 0.10851 |

| | | | | |
|---|---|---|---|---|
| PRSS27 | -0.04622 | 0.74387 | -0.38462 | 0.00976 |
| UPB1 | 0.06489 | 0.89115 | 1.06801 | 0.01227 |
| TGOLN2 | 0.16574 | 0.12666 | 0.09520 | 0.35810 |
| CCER2 | 0.20914 | 0.31600 | -0.37021 | 0.05010 |
| CST6 | -0.13222 | 0.46174 | -0.60719 | 0.00113 |
| KLK7 | -0.51589 | 0.01180 | -0.53190 | 0.00985 |
| PYDC1 | 0.03177 | 0.82693 | 0.04258 | 0.77749 |
| NUMB | -0.14182 | 0.61710 | 1.80477 | 0.00000 |
| IGFBPL1 | 0.30193 | 0.15014 | 0.05123 | 0.75306 |
| TFF3 | 0.72294 | 0.00275 | 0.36972 | 0.04431 |
| MNAT1 | -0.04834 | 0.81734 | 0.14495 | 0.55666 |
| ABL1 | 0.00422 | 0.98800 | 0.49722 | 0.07770 |
| C3 | -0.23721 | 0.19018 | -1.40835 | 0.00000 |
| SPACA5B; SPACA5 | 0.25110 | 0.09641 | -0.03324 | 0.76671 |
| FAS | 0.31956 | 0.02227 | -0.13261 | 0.37867 |
| CSF3R | -0.03447 | 0.82675 | -0.31207 | 0.01118 |
| NUDT15 | 0.09507 | 0.74369 | 0.42936 | 0.03788 |
| ERN1 | -0.14607 | 0.29683 | -0.24014 | 0.04856 |
| BAX | 0.00924 | 0.98333 | 1.30981 | 0.00143 |
| TYMP | 0.12366 | 0.58438 | 0.63934 | 0.00724 |
| NLGN1 | 0.00008 | 0.99975 | 0.08071 | 0.78163 |
| OGFR | 0.10413 | 0.55699 | 0.17177 | 0.42879 |
| CD7 | 0.24395 | 0.11028 | 0.10739 | 0.44938 |
| WFDC2 | 0.81665 | 0.00211 | 0.28034 | 0.24722 |
| KCNIP4 | -0.40990 | 0.06973 | -0.64822 | 0.00465 |
| MLN | 0.94418 | 0.00422 | 0.28444 | 0.35695 |
| VASP | 0.08890 | 0.76724 | 1.91633 | 0.00000 |
| ZNRF4 | 0.50329 | 0.02199 | 0.69673 | 0.00015 |
| TTF2 | -0.03910 | 0.90161 | 0.59222 | 0.05931 |
| HBEGF | 0.09283 | 0.71828 | -0.76185 | 0.01215 |
| CDNF | -0.20266 | 0.17207 | -0.18331 | 0.25494 |
| TIGIT | 0.53926 | 0.01204 | 0.25538 | 0.16405 |
| LIPF | -0.14446 | 0.50882 | -0.59343 | 0.01273 |
| MANSC1 | 0.29317 | 0.02428 | 0.00379 | 0.97512 |
| FCN1 | 0.31531 | 0.11819 | 0.59224 | 0.00027 |
| PPM1F | 0.00270 | 0.98946 | 1.16186 | 0.00000 |
| RGS10 | 0.15861 | 0.55566 | 1.00622 | 0.00057 |
| AKR1B1 | -0.00428 | 0.98568 | 1.70664 | 0.00000 |
| ST6GAL1 | 0.20236 | 0.17471 | -0.35036 | 0.02263 |
| SUOX | 0.38054 | 0.22999 | 1.04316 | 0.00078 |
| PBXIP1 | -0.31728 | 0.01605 | 0.11580 | 0.35104 |
| ANG | -0.09302 | 0.58848 | 0.04890 | 0.76443 |
| IL3 | 0.85185 | 0.04917 | 0.18340 | 0.65801 |
| ENTR1 | 0.02484 | 0.91336 | 0.41105 | 0.06413 |
| GAD1 | 0.15872 | 0.28520 | -0.10367 | 0.45322 |
| MYBPC1 | 0.06392 | 0.87405 | 0.49709 | 0.18274 |
| TPSG1 | -0.10105 | 0.66161 | 0.18112 | 0.36987 |
| BDNF | -0.03961 | 0.88679 | -0.86815 | 0.00069 |

| | | | | |
|---|---|---|---|---|
| NFKB1 | 0.11694 | 0.65458 | 0.19631 | 0.40186 |
| CNP | 0.01383 | 0.95449 | 1.84812 | 0.00000 |
| FGF2 | 0.28012 | 0.28135 | 0.44875 | 0.02852 |
| DBNL | -0.27665 | 0.36807 | 0.84117 | 0.00622 |
| SLC27A4 | 0.15692 | 0.66147 | -0.25658 | 0.44095 |
| SERPINA3 | 0.27681 | 0.02438 | 0.12890 | 0.28713 |
| SH3BP1 | -0.91589 | 0.01381 | -0.05012 | 0.90353 |
| METAP2 | 0.11321 | 0.55012 | 1.02568 | 0.00011 |
| VSIG10L | 0.11059 | 0.55275 | -0.17798 | 0.22475 |
| SERPINA4 | -0.24817 | 0.12321 | -0.20330 | 0.15988 |
| CLEC4D | 0.75430 | 0.01860 | 0.09541 | 0.76170 |
| BECN1 | 0.19057 | 0.47562 | 1.24022 | 0.00000 |
| GAS6 | 0.17537 | 0.20650 | -0.06160 | 0.69217 |
| TPPP3 | -0.03489 | 0.89103 | 0.14800 | 0.54948 |
| DOCK9 | -0.67457 | 0.06483 | -0.09121 | 0.77507 |
| CA1 | 0.08997 | 0.68138 | 1.72926 | 0.00000 |
| MB | -0.40812 | 0.18381 | 0.58278 | 0.07162 |
| RP2 | -0.25032 | 0.18754 | 0.11178 | 0.52764 |
| KIR3DL1 | -0.15849 | 0.80059 | 0.17491 | 0.78143 |
| TDP1 | -0.20449 | 0.61072 | 0.50041 | 0.22811 |
| CBS | 0.18433 | 0.65819 | 0.67700 | 0.06068 |
| GAD2 | -0.08116 | 0.66928 | -0.03084 | 0.85609 |
| ADAM15 | 0.20296 | 0.08340 | -0.16117 | 0.30604 |
| CALB1 | 0.53308 | 0.05973 | 0.17571 | 0.58382 |
| USP28 | 0.55558 | 0.03858 | 0.73353 | 0.00223 |
| NUDC | 0.06128 | 0.75045 | 1.33073 | 0.00000 |
| LGALS3 | 0.31448 | 0.06040 | 0.37322 | 0.00995 |
| GP1BB | 0.45911 | 0.10098 | -1.00158 | 0.00040 |
| RBKS | -0.19596 | 0.50249 | 1.13825 | 0.00011 |
| STX6 | 0.07369 | 0.79447 | 0.90771 | 0.00069 |
| DEFB4A; DEFB4B | 0.68905 | 0.11892 | -1.22363 | 0.00592 |
| TEK | -0.02964 | 0.77424 | -0.02157 | 0.84145 |
| COL3A1 | 0.00823 | 0.96440 | -0.14217 | 0.41209 |
| ATXN2 | 0.54402 | 0.10348 | 0.89254 | 0.00730 |
| IL17D | 0.14969 | 0.27864 | 0.25985 | 0.04731 |
| ANXA4 | -0.32278 | 0.34291 | 0.96062 | 0.00296 |
| LXN | 0.15868 | 0.52717 | 0.58603 | 0.01996 |
| EXOSC10 | 0.03382 | 0.86122 | 0.08575 | 0.64441 |
| ERCC1 | -0.03982 | 0.89955 | -0.05977 | 0.85408 |
| ASAH1 | -0.04780 | 0.79659 | -0.42559 | 0.01410 |
| HRAS | -0.40333 | 0.21692 | -0.62901 | 0.07423 |
| TIMP3 | 0.94745 | 0.01545 | -0.44828 | 0.23206 |
| PDGFC | 0.04130 | 0.72390 | -0.00537 | 0.96419 |
| GC | -0.05524 | 0.59813 | -0.17902 | 0.10482 |
| SMS | -0.28853 | 0.32809 | 0.86626 | 0.00498 |
| TPMT | -0.38936 | 0.21992 | 0.68141 | 0.03145 |
| SRC | 0.16172 | 0.69014 | 1.13237 | 0.00541 |
| NCR3LG1 | 0.58387 | 0.01176 | 0.33445 | 0.09264 |

| | | | | |
|---|---|---|---|---|
| TIMM10 | 0.28786 | 0.15963 | 0.56474 | 0.00011 |
| PDIA5 | -0.15855 | 0.59177 | -0.12381 | 0.62249 |
| CCL16 | 0.40812 | 0.02922 | 0.12202 | 0.55501 |
| ARHGAP1 | 0.02247 | 0.88122 | 1.22171 | 0.00000 |
| RAD23B | 0.05292 | 0.72481 | 0.84442 | 0.00001 |
| RAB39B | -0.57057 | 0.03650 | 0.53281 | 0.05023 |
| CPTP | -0.03469 | 0.84837 | 0.30594 | 0.07648 |
| CADPS | -0.12601 | 0.70471 | -0.55362 | 0.12591 |
| MESD | 0.54220 | 0.21967 | 1.92242 | 0.00002 |
| CTSO | -0.02684 | 0.80295 | 0.09078 | 0.57123 |
| MAPRE3 | 0.66209 | 0.06714 | 0.13879 | 0.66332 |
| EIF4G3 | 0.12675 | 0.75787 | 0.50784 | 0.13992 |
| C1QL2 | -0.21499 | 0.23220 | -0.67757 | 0.00017 |
| ENAH | 0.37862 | 0.09030 | 0.23792 | 0.26917 |
| PTX3 | 0.62599 | 0.00068 | 0.79355 | 0.00034 |
| S100A11 | 0.16588 | 0.57278 | 0.87643 | 0.00211 |
| PGD | -0.13429 | 0.62549 | 0.76643 | 0.00311 |
| SHMT1 | 0.08574 | 0.79227 | 2.18725 | 0.00000 |
| MIF | -0.17747 | 0.51787 | 1.75815 | 0.00000 |
| AIFM1 | 0.63629 | 0.15662 | 2.93446 | 0.00000 |
| CD164 | -0.02457 | 0.87413 | -0.24052 | 0.08146 |
| SERPINA9 | -0.49575 | 0.07102 | -0.30178 | 0.26631 |
| PRDX2 | 0.05714 | 0.80704 | 1.58259 | 0.00000 |
| PPP3R1 | -0.28870 | 0.11084 | 0.08692 | 0.65594 |
| IL18 | 0.23651 | 0.46212 | 0.58467 | 0.02730 |
| FOXJ3 | -0.26942 | 0.31288 | 0.57139 | 0.04216 |
| INHBC | 0.10998 | 0.50496 | -0.03049 | 0.85380 |
| BTC | 0.06342 | 0.83739 | -0.58490 | 0.02354 |
| PROK1 | -0.17169 | 0.40531 | 0.07674 | 0.75666 |
| ISM2 | 0.35130 | 0.06469 | 0.18416 | 0.19159 |
| AIDA | -0.05249 | 0.86213 | 0.77846 | 0.00682 |
| SMNDC1 | 0.49460 | 0.32089 | 0.91223 | 0.04398 |
| SIGLEC9 | 0.15700 | 0.22130 | -0.06136 | 0.74178 |
| IFNG | 0.52594 | 0.23130 | 0.07040 | 0.87852 |
| SDCCAG8 | -0.29547 | 0.42360 | 0.03283 | 0.92454 |
| OGN | 0.51154 | 0.01508 | 0.12300 | 0.58085 |
| PSAP | 0.21841 | 0.06851 | 0.08047 | 0.49418 |
| SPON2 | 0.33309 | 0.07125 | -0.03958 | 0.82732 |
| ITGB1BP2 | 0.41290 | 0.29108 | 3.06483 | 0.00000 |
| CXCL9 | 0.76728 | 0.00206 | 0.23421 | 0.36909 |
| TNFSF11 | -0.78179 | 0.00923 | -0.94932 | 0.00159 |
| MINDY1 | -0.17714 | 0.60596 | 0.21717 | 0.50230 |
| ENTPD6 | 0.00456 | 0.96329 | -0.11891 | 0.19748 |
| DKK1 | 0.37999 | 0.03731 | -0.57354 | 0.00151 |
| EGFL7 | 0.37743 | 0.01028 | -0.29409 | 0.10008 |
| IL1R1 | 0.27612 | 0.01461 | 0.01489 | 0.90809 |
| CLEC1A | 0.20088 | 0.20152 | -0.02605 | 0.87479 |
| WFIKKN2 | 0.08855 | 0.49042 | -0.15993 | 0.28461 |

| | | | | |
|---|---|---|---|---|
| KIRREL1 | -0.00675 | 0.97544 | -0.01372 | 0.96339 |
| DTNB | 0.15289 | 0.52384 | 0.71622 | 0.00555 |
| HDGFL2 | 0.12314 | 0.71106 | 1.28788 | 0.00005 |
| SYAP1 | 0.16885 | 0.54504 | 0.67003 | 0.00240 |
| IL10RB | 0.16305 | 0.17311 | 0.06128 | 0.67095 |
| GPKOW | 0.15633 | 0.52713 | 0.30316 | 0.17961 |
| GFRA1 | 0.41453 | 0.02129 | 0.33721 | 0.15929 |
| SHPK | 0.14987 | 0.50478 | 0.56637 | 0.02838 |
| FABP6 | 0.12843 | 0.62102 | 0.03927 | 0.84619 |
| CDCP1 | 0.70677 | 0.00314 | 0.35662 | 0.13827 |
| DMP1 | -0.30702 | 0.21144 | 0.57737 | 0.02017 |
| PPIB | 0.04859 | 0.87990 | 0.15926 | 0.57919 |
| S100A13 | 0.31314 | 0.07498 | -0.16748 | 0.27325 |
| CLEC6A | 0.61886 | 0.00624 | -0.17513 | 0.41921 |
| GOLGA3 | 0.62091 | 0.01452 | 1.11898 | 0.00001 |
| TGFBI | 0.02286 | 0.84994 | -0.11698 | 0.37029 |
| IFNL1 | 0.38930 | 0.16220 | 0.28154 | 0.40440 |
| IL1RAP | 0.04901 | 0.70057 | 0.01674 | 0.91059 |
| DUT | 0.90568 | 0.01542 | 0.24930 | 0.45650 |
| SLC28A1 | -0.05935 | 0.81297 | 0.17817 | 0.47264 |
| TG | 0.37757 | 0.23883 | 0.11930 | 0.74955 |
| NMNAT1 | -0.13402 | 0.75085 | 0.34301 | 0.42156 |
| TLR4 | 0.14181 | 0.26736 | -0.02370 | 0.84059 |
| CDH1 | 0.42908 | 0.00384 | -0.48189 | 0.00196 |
| CNTF | -0.16652 | 0.71196 | -0.33282 | 0.47503 |
| DLK1 | -0.00543 | 0.97775 | -0.23850 | 0.16058 |
| SLMAP | 0.36019 | 0.23942 | 0.35092 | 0.23189 |
| ADA | -0.20643 | 0.25206 | 0.52559 | 0.01567 |
| TXNDC15 | 0.29698 | 0.00979 | -0.01249 | 0.92191 |
| CA8 | 0.05958 | 0.66440 | -0.10881 | 0.20472 |
| AFP | 0.18610 | 0.50608 | 0.04613 | 0.84806 |
| PMM2 | 1.00602 | 0.00708 | 0.86573 | 0.00514 |
| DPP4 | -0.31897 | 0.01768 | -0.28198 | 0.05549 |
| BLNK | 0.31949 | 0.06131 | 0.77768 | 0.00064 |
| CSF2RB | -0.35459 | 0.05539 | -0.15166 | 0.39511 |
| COMP | -0.37630 | 0.03817 | -0.73468 | 0.00029 |
| TMPRSS11D | 0.30067 | 0.11296 | -0.13828 | 0.31733 |
| STK24 | -0.11207 | 0.65807 | 0.50713 | 0.04189 |
| HS3ST3B1 | 0.11047 | 0.51696 | 0.04869 | 0.83950 |
| AARSD1 | 0.31625 | 0.17740 | 1.34445 | 0.00000 |
| TSPAN15 | 0.15217 | 0.52532 | 0.02902 | 0.91675 |
| FIS1 | -0.20709 | 0.49986 | 0.51453 | 0.07009 |
| ADAMTSL5 | -0.11608 | 0.43276 | -0.14811 | 0.28037 |
| FGFR4 | 0.02440 | 0.87680 | -0.26854 | 0.10605 |
| IVD | 0.40664 | 0.21854 | 0.31375 | 0.32396 |
| PCDHB15 | -0.08505 | 0.64570 | -0.72341 | 0.00001 |
| SNX18 | -0.05464 | 0.72515 | 0.29390 | 0.05136 |
| FCAMR | 0.32588 | 0.23897 | 0.19277 | 0.42568 |

| | | | | |
|---|---|---|---|---|
| HAVCR1 | 0.89446 | 0.00616 | 0.64746 | 0.06497 |
| IL2RG | 0.25967 | 0.08207 | 0.07839 | 0.55457 |
| SESTD1 | -0.29928 | 0.28054 | -0.24969 | 0.28781 |
| TGFA | 1.01247 | 0.00066 | -0.20542 | 0.48278 |
| CFH | 0.01467 | 0.86357 | 0.00064 | 0.99390 |
| GNLY | 0.25091 | 0.21103 | 0.07572 | 0.68192 |
| IMMT | 0.73060 | 0.00868 | 0.79453 | 0.00171 |
| MENT | -0.26439 | 0.01287 | -0.22439 | 0.04875 |
| PRKRA | 0.06017 | 0.81319 | 0.56407 | 0.02427 |
| PGA4 | -0.37464 | 0.31490 | -0.35291 | 0.20743 |
| ADAM9 | 0.17420 | 0.23499 | 0.05660 | 0.68930 |
| NAMPT | -0.24715 | 0.51711 | 0.96284 | 0.02099 |
| EPB41L5 | 0.13006 | 0.70893 | 0.25586 | 0.39691 |
| MMP13 | -0.00806 | 0.97792 | -0.75224 | 0.03281 |
| DNAJA4 | -0.04308 | 0.83709 | 1.23135 | 0.00000 |
| B4GALT1 | 0.17024 | 0.21313 | 0.18393 | 0.24094 |
| ATP2B4 | 0.54655 | 0.05774 | -0.10225 | 0.74903 |
| CNTN4 | -0.08983 | 0.36164 | -0.24997 | 0.02774 |
| NADK | -0.02855 | 0.93734 | 0.50470 | 0.16158 |
| ACADM | 1.01600 | 0.00701 | -0.11596 | 0.75422 |
| NAA10 | -0.13342 | 0.65990 | 0.43043 | 0.13963 |
| PKLR | 0.21545 | 0.38312 | 1.56415 | 0.00000 |
| SIT1 | 0.58660 | 0.02531 | 0.96072 | 0.00023 |
| WASHC3 | -0.25708 | 0.46067 | 1.15110 | 0.00103 |
| NDST1 | -0.14438 | 0.52063 | -0.38786 | 0.07724 |
| LAMA1 | -0.08653 | 0.68277 | -0.02555 | 0.90563 |
| BCR | 0.31256 | 0.27184 | 1.31418 | 0.00000 |
| MRPL58 | 0.39877 | 0.28190 | 1.17590 | 0.00061 |
| IGFBP3 | -0.51220 | 0.00633 | -0.48720 | 0.01420 |
| PLCB2 | 0.04463 | 0.90786 | 1.78238 | 0.00001 |
| TBL1X | 0.03567 | 0.89848 | 1.13477 | 0.00015 |
| COL9A2 | 0.04519 | 0.86300 | -0.40009 | 0.27337 |
| LBP | 0.26053 | 0.30077 | 0.13217 | 0.57272 |
| SLITRK2 | -0.02077 | 0.90864 | -0.18170 | 0.37476 |
| RYR1 | -0.24289 | 0.25217 | -0.09214 | 0.62212 |
| ARG1 | -0.14591 | 0.57410 | 0.82009 | 0.00149 |
| SIGLEC6 | -0.21027 | 0.16005 | -0.28832 | 0.19777 |
| CLNS1A | 0.08252 | 0.68646 | 0.54978 | 0.00823 |
| MANSC4 | 0.41290 | 0.17603 | -0.32592 | 0.25066 |
| MSMB | 0.22024 | 0.37982 | -0.07830 | 0.72633 |
| TBCA | -0.15289 | 0.62918 | 1.77952 | 0.00000 |
| DDAH1 | 0.00222 | 0.99456 | 0.92042 | 0.00241 |
| TRAF2 | -0.47704 | 0.01918 | 0.46547 | 0.03104 |
| NEO1 | -0.09033 | 0.19292 | -0.04688 | 0.43981 |
| OXT | -0.02697 | 0.94100 | 0.41543 | 0.21760 |
| C1QBP | 0.54520 | 0.04433 | 0.68002 | 0.00080 |
| DOK2 | 0.38359 | 0.33861 | 0.61254 | 0.11279 |
| NPTX1 | 0.14609 | 0.41643 | 0.12484 | 0.43260 |

| | | | | |
|---|---|---|---|---|
| ARHGEF10 | 0.52462 | 0.09022 | 0.54783 | 0.05007 |
| PLA2G4A | 0.34598 | 0.26980 | 1.77765 | 0.00000 |
| RNASET2 | 0.11976 | 0.33332 | -0.14858 | 0.23785 |
| NUCB2 | 0.09499 | 0.49759 | 0.02688 | 0.88718 |
| ART3 | -0.01030 | 0.94331 | -0.18076 | 0.23608 |
| GRSF1 | 0.81547 | 0.03609 | 1.92466 | 0.00000 |
| RNASE10 | 0.31805 | 0.17273 | -0.05671 | 0.79521 |
| THRAP3 | -0.04766 | 0.90724 | 0.13546 | 0.69626 |
| IL5 | -0.39506 | 0.43456 | -0.62205 | 0.16667 |
| TRIM21 | -0.73306 | 0.01012 | 0.75433 | 0.02140 |
| MYOM1 | 0.08417 | 0.67524 | 0.20479 | 0.31056 |
| PDE5A | 0.25901 | 0.41605 | 1.85651 | 0.00000 |
| ERVV-1 | 0.31044 | 0.20154 | -0.10694 | 0.59471 |
| LTB | -0.18294 | 0.27423 | -0.22598 | 0.27260 |
| GSR | 0.17108 | 0.25957 | 0.29124 | 0.00752 |
| CSF1 | 0.38884 | 0.03268 | 0.30096 | 0.09811 |
| CLGN | 0.23900 | 0.31384 | 0.19266 | 0.33899 |
| SCN2B | 0.16722 | 0.32365 | 0.19891 | 0.29015 |
| FMNL1 | -0.25178 | 0.52168 | 0.16389 | 0.69960 |
| SORCS2 | 0.44165 | 0.01857 | 0.32320 | 0.15217 |
| CHI3L1 | 0.80934 | 0.03589 | 0.59391 | 0.10834 |
| PDZK1 | 0.20530 | 0.62077 | 0.52406 | 0.16193 |
| TTR | -0.48195 | 0.00995 | -0.41340 | 0.01721 |
| CALCA | -0.09842 | 0.82370 | 0.48649 | 0.28236 |
| EFNB2 | 0.39877 | 0.10545 | 0.05654 | 0.80795 |
| AMY2A | -0.02523 | 0.90218 | -0.27894 | 0.14582 |
| PSMG4 | 0.16930 | 0.48263 | 1.96395 | 0.00000 |
| PTPRR | 0.00184 | 0.99028 | -0.13950 | 0.28377 |
| SLA2 | 0.32747 | 0.38333 | 2.74764 | 0.00000 |
| FABP5 | -0.03665 | 0.90812 | 1.49661 | 0.00001 |
| TIA1 | 0.18243 | 0.69065 | 2.60324 | 0.00000 |
| EPHA1 | 0.31626 | 0.01313 | 0.19309 | 0.16669 |
| MANF | 0.21820 | 0.54814 | 0.96675 | 0.00824 |
| DMD | 0.32483 | 0.22734 | 0.66679 | 0.01284 |
| CR1 | 0.01046 | 0.94273 | -0.10188 | 0.48054 |
| CPXM1 | 0.36108 | 0.05848 | -1.20023 | 0.00000 |
| PLAU | -0.17233 | 0.19584 | -0.14066 | 0.30767 |
| MEGF11 | 0.04601 | 0.72143 | -0.31279 | 0.05871 |
| S100A4 | -0.40074 | 0.09145 | 0.32382 | 0.10458 |
| ATP6V1F | -0.36397 | 0.18176 | -0.25705 | 0.32709 |
| CCL19 | -0.45456 | 0.09814 | -0.31803 | 0.25291 |
| INSL5 | 0.49594 | 0.09249 | 0.50858 | 0.10436 |
| MMP8 | 1.17010 | 0.02000 | -0.36667 | 0.43534 |
| PDE4D | -0.19676 | 0.53978 | 0.21008 | 0.54009 |
| MDM1 | -0.34593 | 0.20853 | 0.60350 | 0.04957 |
| CCL17 | 0.19928 | 0.51171 | -1.09445 | 0.00003 |
| EPS8L2 | 0.32750 | 0.21112 | 0.49441 | 0.04542 |
| ST13 | 0.16221 | 0.42287 | 1.70615 | 0.00000 |

| | | | | |
|---|---|---|---|---|
| TXLNA | 0.26285 | 0.35437 | 1.39422 | 0.00000 |
| PLA2G2A | -0.02854 | 0.94270 | 0.15189 | 0.64874 |
| FASLG | -0.10756 | 0.44660 | -0.23160 | 0.25407 |
| TACSTD2 | 0.11282 | 0.50111 | -0.14672 | 0.42018 |
| CLIC5 | 0.45748 | 0.01441 | 0.13355 | 0.45468 |
| TNC | 0.14084 | 0.52422 | 0.21003 | 0.33647 |
| ENTPD5 | -0.15421 | 0.07046 | -0.08565 | 0.39358 |
| MRPL28 | 0.20062 | 0.11239 | 0.04934 | 0.59860 |
| CDC42BPB | 0.44658 | 0.16832 | 0.73067 | 0.01316 |
| SCN4B | -0.27150 | 0.16111 | -0.39502 | 0.02175 |
| MDH1 | 0.03030 | 0.49270 | 0.03913 | 0.47428 |
| TBC1D17 | -0.00447 | 0.97981 | 0.49446 | 0.00200 |
| IGF2R | 0.17170 | 0.07545 | 0.09745 | 0.35030 |
| CST3 | 0.22159 | 0.12565 | -0.01658 | 0.91235 |
| VPS53 | -0.07755 | 0.76523 | 0.73578 | 0.00224 |
| SUSD2 | 0.10046 | 0.37696 | -0.21734 | 0.10222 |
| PTP4A3 | 0.19368 | 0.23891 | 0.65256 | 0.00001 |
| GSTM4 | -0.01738 | 0.95703 | -0.17902 | 0.56596 |
| FN1 | -0.01318 | 0.91888 | 0.55874 | 0.00008 |
| DNER | -0.41028 | 0.00143 | -0.37561 | 0.00368 |
| TNFSF9 | 0.28334 | 0.12619 | -0.07494 | 0.64029 |
| TPBGL | -0.06164 | 0.73087 | -0.08700 | 0.64013 |
| NIT2 | -0.20638 | 0.53628 | 1.78592 | 0.00000 |
| MATN3 | -0.04947 | 0.85472 | 0.09945 | 0.72549 |
| EIF2AK3 | 0.06314 | 0.85122 | 0.41408 | 0.14634 |
| GP6 | 0.14273 | 0.54235 | -0.55294 | 0.02104 |
| LUZP2 | 0.30556 | 0.01847 | 0.07162 | 0.55027 |
| CLPS | 0.22321 | 0.48299 | -0.01240 | 0.96379 |
| PLAT | 0.22590 | 0.33669 | 0.52913 | 0.04428 |
| GSTA3 | -0.14743 | 0.68230 | 0.64195 | 0.05232 |
| NRCAM | 0.14487 | 0.37170 | -0.15349 | 0.26536 |
| AGXT | 0.25144 | 0.50908 | 1.17207 | 0.00019 |
| RILPL2 | -0.08412 | 0.79337 | 1.76877 | 0.00000 |
| CFB | 0.16570 | 0.20577 | 0.15440 | 0.20612 |
| LSM8 | 0.16272 | 0.62792 | -0.16764 | 0.62820 |
| ANGPTL3 | 0.09725 | 0.41496 | -0.10923 | 0.45919 |
| TNFSF13 | 0.42288 | 0.01179 | 0.23318 | 0.20793 |
| SPAG1 | -0.25220 | 0.34151 | 0.44216 | 0.09686 |
| OLR1 | 0.79773 | 0.04811 | -0.48206 | 0.21983 |
| TSLP | -0.16102 | 0.42418 | -0.14425 | 0.59322 |
| PDRG1 | -0.20233 | 0.50586 | 0.57955 | 0.04263 |
| DRG2 | -0.30021 | 0.14423 | 0.33999 | 0.07940 |
| ZP3 | 0.42913 | 0.49066 | 0.24454 | 0.66719 |
| CRYZL1 | 0.26591 | 0.34641 | 2.08134 | 0.00000 |
| AXL | 0.15008 | 0.21356 | 0.18353 | 0.16567 |
| MAP3K5 | -0.40545 | 0.29986 | -1.64218 | 0.00001 |
| CRIM1 | 0.20290 | 0.12456 | 0.17839 | 0.18668 |
| PREB | 0.34380 | 0.02882 | 0.43544 | 0.00359 |

| | | | | |
|---|---|---|---|---|
| SCGB3A1 | 0.01209 | 0.93131 | -0.22211 | 0.08893 |
| NRN1 | 0.22557 | 0.13608 | -0.19846 | 0.25614 |
| SMOC1 | 0.62690 | 0.00961 | 0.20478 | 0.47787 |
| KYAT1 | -0.20759 | 0.44162 | 1.18047 | 0.00001 |
| PRDX3 | 0.43678 | 0.17444 | 1.47522 | 0.00000 |
| CASP2 | -0.48148 | 0.05766 | -1.04347 | 0.00002 |
| PLTP | 0.05044 | 0.74065 | -0.68214 | 0.00004 |
| GLT8D2 | 0.01523 | 0.93914 | -0.07241 | 0.66110 |
| NAAA | -0.08421 | 0.72028 | -0.06061 | 0.78791 |
| MFAP5 | 0.14729 | 0.50599 | -0.60231 | 0.00390 |
| LGALS7B; LGALS7 | 0.28983 | 0.12989 | 0.16310 | 0.36120 |
| WFDC1 | 0.04519 | 0.71867 | 0.33883 | 0.01618 |
| LRRFIP1 | 0.06755 | 0.78910 | 0.02803 | 0.91742 |
| PLEKHO1 | 0.49961 | 0.10946 | 1.76432 | 0.00000 |
| LYPD8 | 0.20655 | 0.35388 | 0.02896 | 0.90222 |
| TMED10 | -0.11237 | 0.67404 | 0.10401 | 0.73358 |
| NUDT10 | -0.21327 | 0.18073 | -0.02370 | 0.87484 |
| AAMDC | -0.05077 | 0.78319 | 0.34852 | 0.05476 |
| BHLHE40 | 0.10492 | 0.80545 | -0.75112 | 0.07873 |
| ANP32C | -0.05770 | 0.51648 | -0.40704 | 0.00578 |
| COL9A1 | -0.60595 | 0.00043 | -0.10679 | 0.62564 |
| SMPD3 | 0.41696 | 0.14413 | 0.33143 | 0.15891 |
| GKN1 | -0.10451 | 0.51033 | -0.09208 | 0.52103 |
| ADGRE2 | -0.08567 | 0.47891 | -0.20053 | 0.13964 |
| POF1B | 0.92148 | 0.00110 | 0.17773 | 0.43396 |
| CD1C | -0.30425 | 0.04516 | -0.28170 | 0.04543 |
| GLP1R | 0.67735 | 0.00713 | -0.26798 | 0.27055 |
| RASSF2 | -0.13867 | 0.74505 | 0.45543 | 0.26322 |
| PBK | -0.00974 | 0.94155 | -0.07609 | 0.51094 |
| GNGT1 | 0.14314 | 0.54220 | -0.41173 | 0.09030 |
| MAMDC2 | 0.23691 | 0.30027 | 1.55560 | 0.00000 |
| GPNMB | 0.13988 | 0.21152 | -0.07210 | 0.60687 |
| SEPTIN3 | 0.09376 | 0.71530 | 0.28988 | 0.22041 |
| BLMH | 0.24292 | 0.18785 | 0.36489 | 0.01747 |
| KDM3A | 0.01765 | 0.91429 | -0.49736 | 0.00567 |
| L1CAM | -0.14613 | 0.30103 | -0.29438 | 0.02335 |
| DCUN1D2 | -0.06608 | 0.86133 | 0.19001 | 0.54558 |
| PSTPIP2 | -0.06572 | 0.83938 | 1.52152 | 0.00001 |
| DOC2B | -0.21455 | 0.58348 | 0.51626 | 0.12768 |
| DSG4 | -0.51331 | 0.00325 | -1.05266 | 0.00000 |
| KLB | 0.01465 | 0.94550 | -0.30806 | 0.13534 |
| HDDC2 | -0.05614 | 0.73499 | 0.72970 | 0.00000 |
| CARHSP1 | 0.05768 | 0.74184 | 1.64064 | 0.00000 |
| KIAA0319 | 0.29788 | 0.06141 | -0.06026 | 0.60586 |
| PPP1R14A | 0.13669 | 0.53714 | 1.05354 | 0.00010 |
| REG4 | 0.60142 | 0.00235 | 0.18082 | 0.38200 |
| IL20RA | 0.02226 | 0.91283 | 0.01814 | 0.91294 |
| MYH9 | 0.34779 | 0.33212 | -0.01369 | 0.96308 |

| | | | | |
|---|---|---|---|---|
| KIAA1549 | 0.78611 | 0.20379 | 0.68600 | 0.25356 |
| CKAP4 | 0.46142 | 0.04938 | 0.23021 | 0.41090 |
| KHK | 0.04393 | 0.88329 | 1.19298 | 0.00004 |
| NOP56 | 0.37832 | 0.32442 | -0.25286 | 0.41926 |
| CD163 | 0.25626 | 0.18102 | 0.25506 | 0.18691 |
| GRHPR | 0.10029 | 0.75175 | 2.37118 | 0.00000 |
| REG3A | 1.00889 | 0.00011 | 0.40666 | 0.12861 |
| MDGA1 | -0.44929 | 0.07273 | -0.43545 | 0.09919 |
| GPA33 | -0.46079 | 0.19246 | -0.65644 | 0.10770 |
| CAPS | 0.32797 | 0.29106 | 1.16008 | 0.00000 |
| GHRL | -0.11049 | 0.75271 | -0.39835 | 0.29620 |
| CD59 | 0.19934 | 0.18540 | 0.12088 | 0.41095 |
| SCLY | 0.16317 | 0.51367 | 0.96658 | 0.00133 |
| MITD1 | 0.22743 | 0.49910 | 1.64738 | 0.00000 |
| LAMP3 | 0.28907 | 0.12909 | -0.14188 | 0.56833 |
| PEPD | -0.06390 | 0.61264 | -0.19806 | 0.06343 |
| MYOM3 | -0.37320 | 0.20196 | -0.19407 | 0.43797 |
| SPRED2 | 0.00409 | 0.97164 | -0.00458 | 0.95653 |
| SCN2A | 0.08797 | 0.79117 | 0.77811 | 0.01444 |
| BCL7B | 0.14086 | 0.42010 | -0.13444 | 0.40990 |
| FAP | -0.15244 | 0.21887 | -0.25148 | 0.04950 |
| CD300C | 0.30999 | 0.04867 | 0.18082 | 0.28393 |
| NOTCH1 | -0.04583 | 0.50266 | -0.21388 | 0.03957 |
| GGH | -0.06935 | 0.60294 | 0.00532 | 0.96832 |
| PTPRC | -0.01143 | 0.91015 | -0.07607 | 0.43824 |
| CORO6 | -0.31037 | 0.07982 | -0.20258 | 0.21296 |
| UBQLN3 | -0.11516 | 0.69531 | -0.20112 | 0.44345 |
| CCL11 | 0.37872 | 0.00592 | -0.19920 | 0.39636 |
| RSPO3 | 0.39240 | 0.04154 | 0.46226 | 0.06056 |
| RRAS | 0.50198 | 0.30059 | 2.47476 | 0.00000 |
| CCL23 | 0.42585 | 0.05002 | 0.24395 | 0.22615 |
| APPL2 | -0.33843 | 0.34786 | 0.36724 | 0.27219 |
| PHACTR2 | -0.24387 | 0.51309 | 1.69139 | 0.00001 |
| DSG2 | 0.14403 | 0.17561 | -0.19504 | 0.08153 |
| SERPINE1 | 0.10139 | 0.61080 | -0.48914 | 0.01821 |
| CSPG4 | -0.01857 | 0.90075 | 0.12314 | 0.36774 |
| PLXDC2 | -0.01777 | 0.90070 | 1.14825 | 0.00000 |
| ASS1 | -0.02300 | 0.95635 | 0.94096 | 0.00793 |
| ART5 | 0.15941 | 0.41755 | -0.01572 | 0.94747 |
| LY96 | 0.07636 | 0.56843 | -0.02601 | 0.84995 |
| TNFRSF13C | 0.04957 | 0.75283 | 0.02541 | 0.89688 |
| SLC34A3 | 0.50767 | 0.22132 | -0.00851 | 0.98126 |
| RBPMS2 | -0.24285 | 0.56098 | -1.08936 | 0.00873 |
| PIBF1 | -0.32602 | 0.29827 | 1.48301 | 0.00000 |
| HMOX1 | -0.13222 | 0.61036 | 0.42220 | 0.06353 |
| CLTA | -0.02967 | 0.89453 | 0.94640 | 0.00005 |
| LMNB2 | 0.74510 | 0.00706 | 0.97211 | 0.00013 |
| DCTN2 | -0.42280 | 0.21797 | 0.57909 | 0.11569 |

| | | | | |
|---|---|---|---|---|
| HMGCL | 0.31712 | 0.35505 | 1.19182 | 0.00037 |
| SOWAHA | 0.52106 | 0.03592 | 0.15994 | 0.63886 |
| MEGF9 | -0.14498 | 0.13890 | -0.14688 | 0.22330 |
| CD83 | 0.00822 | 0.95581 | -0.07709 | 0.63034 |
| NAP1L4 | -0.13916 | 0.28111 | 1.23823 | 0.00000 |
| LBR | -0.12125 | 0.73912 | 0.74870 | 0.04995 |
| DAND5 | 1.34642 | 0.01133 | -0.25613 | 0.62461 |
| EDA2R | 0.90307 | 0.00102 | 0.43059 | 0.10238 |
| SF3B4 | -0.45581 | 0.16147 | 1.16336 | 0.00079 |
| IL13 | 0.13779 | 0.62282 | -0.04050 | 0.89059 |
| RBM25 | 0.85420 | 0.01238 | 0.51295 | 0.14673 |
| DHRS4L2 | 0.24612 | 0.45601 | 1.31922 | 0.00001 |
| DNM3 | 0.12167 | 0.60180 | 0.47128 | 0.04588 |
| DDX1 | -0.29133 | 0.24913 | 0.36654 | 0.12091 |
| CAT | -0.03568 | 0.84883 | 1.27751 | 0.00000 |
| ADCYAP1R1 | 0.23729 | 0.11348 | -0.30606 | 0.01915 |
| LGALS1 | 0.29612 | 0.09030 | 0.61484 | 0.00068 |
| CSRP3 | -0.79774 | 0.06486 | 0.50554 | 0.28250 |
| VNN2 | 0.07413 | 0.66315 | -0.57341 | 0.00717 |
| FCER1A | -0.28889 | 0.00143 | -0.07871 | 0.26317 |
| CTHRC1 | 0.17824 | 0.30372 | -0.07005 | 0.66841 |
| NTF3 | -0.42448 | 0.06156 | 0.53311 | 0.01584 |
| IL1RN | 0.35406 | 0.36380 | 1.20546 | 0.00185 |
| LDLR | -0.14274 | 0.38774 | 0.14492 | 0.47090 |
| KLK6 | 0.12162 | 0.44261 | -0.12004 | 0.41344 |
| PNLIPRP1 | 0.27712 | 0.38886 | 0.02524 | 0.93411 |
| SIRT2 | 0.06514 | 0.82280 | 1.90817 | 0.00000 |
| EDN1 | 0.05469 | 0.76604 | 0.79795 | 0.00000 |
| UNC79 | -0.18919 | 0.51921 | -0.15941 | 0.56423 |
| TCL1A | -0.10916 | 0.79860 | 1.01461 | 0.01308 |
| GABRA4 | 0.54306 | 0.05504 | -0.40400 | 0.12424 |
| CCL20 | 0.58001 | 0.14369 | 0.72037 | 0.07277 |
| MRC1 | 0.47374 | 0.00884 | 0.31284 | 0.07712 |
| CD69 | 0.03707 | 0.90958 | -0.22681 | 0.51197 |
| VCAN | 0.43118 | 0.00623 | 0.31101 | 0.05243 |
| ELAC1 | 0.30694 | 0.28741 | 1.92130 | 0.00000 |
| GH2 | 0.39765 | 0.38711 | -0.40335 | 0.41413 |
| FRZB | 0.03008 | 0.83509 | 0.41272 | 0.01863 |
| CASQ2 | 0.12675 | 0.71019 | 0.31547 | 0.16505 |
| BNIP3L | 0.55446 | 0.04981 | 0.88865 | 0.00123 |
| RCOR1 | 0.42683 | 0.23249 | 0.39806 | 0.16144 |
| CEP112 | 0.31802 | 0.04881 | 0.32413 | 0.06183 |
| SORT1 | 0.21477 | 0.14132 | -0.46396 | 0.00218 |
| DPEP2 | 0.24864 | 0.05003 | 0.94782 | 0.00002 |
| RET | -0.65864 | 0.00041 | -1.39323 | 0.00000 |
| SEPTIN8 | 0.32428 | 0.09455 | 0.41330 | 0.01042 |
| TIMM8A | 0.13748 | 0.64377 | 1.89794 | 0.00000 |
| SRPK2 | 0.26129 | 0.33346 | 0.78402 | 0.00117 |

| | | | | |
|---|---|---|---|---|
| CPB2 | -0.03300 | 0.75433 | -0.05598 | 0.56485 |
| SH3BGRL2 | 0.20830 | 0.35456 | -0.79907 | 0.00094 |
| USO1 | 0.17868 | 0.54665 | 1.64332 | 0.00000 |
| PTH | 0.09524 | 0.80689 | 0.78017 | 0.04380 |
| C1QTNF1 | -0.03744 | 0.85332 | 0.83190 | 0.00059 |
| TNFSF8 | -0.00128 | 0.99328 | -0.01734 | 0.90133 |
| GIT1 | 0.13313 | 0.61511 | 1.12170 | 0.00009 |
| NID1 | 0.24546 | 0.07860 | 0.09390 | 0.51210 |
| MCAM | -0.07056 | 0.61971 | -0.21347 | 0.14278 |
| SPON1 | 0.33521 | 0.03912 | 0.25323 | 0.14363 |
| PRSS2 | 0.61981 | 0.04340 | 0.03664 | 0.89884 |
| CREBZF | 0.11644 | 0.18182 | -0.04405 | 0.68632 |
| MZT1 | -0.43291 | 0.09456 | -0.15271 | 0.49717 |
| CETN2 | -0.13607 | 0.65580 | 0.31141 | 0.30836 |
| OMG | -0.19516 | 0.43500 | -0.40289 | 0.13396 |
| UBE2L6 | 0.32164 | 0.20372 | 1.21779 | 0.00000 |
| VSTM2B | 0.18847 | 0.35986 | -0.01365 | 0.95171 |
| PDCL2 | 0.13867 | 0.57252 | 0.05295 | 0.83046 |
| PXDNL | 0.06701 | 0.74514 | -0.45570 | 0.00411 |
| ITGAV | -0.24052 | 0.02010 | -0.17686 | 0.30301 |
| HEBP1 | 0.18942 | 0.47956 | -0.42465 | 0.04849 |
| CNPY4 | 0.15105 | 0.50038 | 0.50958 | 0.04523 |
| CRACR2A | -0.57381 | 0.17201 | -1.76458 | 0.00014 |
| IL4R | 0.80670 | 0.00299 | 0.43822 | 0.11609 |
| TLR1 | 0.00111 | 0.99470 | -0.11796 | 0.43732 |
| NRTN | 0.12417 | 0.57193 | 0.03580 | 0.88548 |
| TNR | -0.45119 | 0.00715 | -0.05861 | 0.72374 |
| ATXN3 | -0.07133 | 0.75642 | 1.62058 | 0.00000 |
| CXCL13 | 0.84124 | 0.00228 | -0.33004 | 0.18668 |
| PXN | 0.18083 | 0.64977 | 0.22780 | 0.57641 |
| FKBP5 | -0.16910 | 0.55208 | 0.27467 | 0.39348 |
| SERPINC1 | -0.06079 | 0.39117 | -0.02153 | 0.75763 |
| LRRN1 | -0.55789 | 0.00198 | -0.76256 | 0.00001 |
| F11R | 0.50742 | 0.01345 | 0.18637 | 0.36281 |
| CRYBB2 | 0.33829 | 0.24638 | 0.35433 | 0.11284 |
| PHOSPHO1 | 0.04231 | 0.71178 | -0.07679 | 0.57134 |
| GLB1 | -0.47750 | 0.01571 | -1.04151 | 0.00000 |
| PCDH1 | 0.04580 | 0.25245 | -0.11689 | 0.42067 |
| ICAM3 | 0.08759 | 0.34580 | -0.21799 | 0.06476 |
| ARID4B | -0.17040 | 0.52242 | 0.62418 | 0.04697 |
| GIPC3 | -0.09790 | 0.73422 | 0.47971 | 0.12804 |
| MFAP3 | 0.11267 | 0.54922 | -0.02156 | 0.90844 |
| F7 | -0.01760 | 0.89997 | 0.14853 | 0.32973 |
| ICA1 | -0.02501 | 0.93627 | 0.57600 | 0.03634 |
| GSTT2B | -0.14488 | 0.80852 | 1.80109 | 0.00173 |
| PLXNB3 | -0.14796 | 0.36383 | -0.23442 | 0.16440 |
| HNMT | 0.30799 | 0.39554 | 1.01848 | 0.00399 |
| FZD10 | 0.55642 | 0.32355 | 0.67656 | 0.15468 |

| | | | | |
|---|---|---|---|---|
| PPP1R2 | 0.25129 | 0.37751 | 1.63333 | 0.00000 |
| ARFIP1 | 0.11423 | 0.70734 | 2.01060 | 0.00000 |
| SERPINF1 | -0.09243 | 0.08749 | -0.04235 | 0.44924 |
| GMFG | -0.15147 | 0.60995 | 1.31511 | 0.00001 |
| SPINT3 | -0.48509 | 0.20377 | -0.21134 | 0.61793 |
| CD63 | 0.17592 | 0.45207 | -0.55709 | 0.02504 |
| CDH2 | 0.48108 | 0.00055 | 0.48568 | 0.00175 |
| C1QTNF5 | -0.29135 | 0.01967 | -0.19910 | 0.13302 |
| SPOCK1 | -0.07608 | 0.62877 | -0.02874 | 0.84460 |
| TNFAIP8L2 | -0.24115 | 0.47022 | 1.35520 | 0.00001 |
| GNPDA2 | 0.03961 | 0.81136 | 0.16034 | 0.26083 |
| SERPINA6 | -0.02217 | 0.79890 | -0.12408 | 0.19893 |
| UPK3A | -0.25149 | 0.33804 | -0.26351 | 0.30245 |
| HS1BP3 | 0.14689 | 0.53800 | 1.77063 | 0.00000 |
| LAIR1 | 0.48554 | 0.01977 | 0.27005 | 0.20468 |
| AKT2 | 0.41263 | 0.39435 | 3.90856 | 0.00000 |
| CD226 | -0.00329 | 0.98413 | -0.60864 | 0.00034 |
| C1QA | 0.47817 | 0.00101 | -0.01115 | 0.94794 |
| BMP4 | -0.23011 | 0.30347 | 0.10310 | 0.63309 |
| SCAMP3 | 0.18189 | 0.55292 | -0.21524 | 0.51680 |
| GLA | 0.01560 | 0.91010 | 0.03243 | 0.79559 |
| ADH1B | -0.18766 | 0.69529 | 1.14792 | 0.00806 |
| SUMF1 | 0.10940 | 0.60160 | 0.36978 | 0.11055 |
| PDGFRB | 0.16216 | 0.14227 | -0.17578 | 0.15000 |
| ARL2BP | -0.04953 | 0.84730 | 1.32023 | 0.00000 |
| CEP85 | -0.26847 | 0.24675 | 0.12814 | 0.57801 |
| COMMD9 | -0.13927 | 0.46809 | 0.18832 | 0.23947 |
| YOD1 | 0.00442 | 0.98613 | 0.77683 | 0.00187 |
| IGLON5 | 0.09837 | 0.73492 | -0.15025 | 0.55967 |
| PGR | 0.08867 | 0.69687 | 0.24182 | 0.25533 |
| RBPMS | -0.08904 | 0.85235 | -0.37545 | 0.34779 |
| CCL15 | 0.29440 | 0.09872 | 0.08617 | 0.65809 |
| NTproBNP | 1.67983 | 0.00077 | 0.71582 | 0.10008 |
| GADD45GIP1 | 0.51573 | 0.07668 | 1.14407 | 0.00001 |
| ODAM | -0.08205 | 0.74317 | -0.34213 | 0.13408 |
| IL2RB | 0.36953 | 0.06737 | 0.06883 | 0.72125 |
| TNFRSF11A | 0.51178 | 0.02013 | 0.18323 | 0.53944 |
| ITGBL1 | 0.22179 | 0.18035 | 0.26410 | 0.04288 |
| CXADR | 0.51863 | 0.04669 | 0.26123 | 0.24395 |
| FZD8 | 0.21917 | 0.14721 | 0.01506 | 0.94296 |
| IMPACT | 0.45220 | 0.02563 | 2.00738 | 0.00000 |
| CFI | 0.03336 | 0.57270 | -0.02661 | 0.63827 |
| STAT2 | 0.45907 | 0.12329 | 1.03095 | 0.00021 |
| CTSS | 0.08750 | 0.30186 | 0.11873 | 0.21921 |
| F2 | -0.13580 | 0.05504 | -0.39070 | 0.00000 |
| ADGRE5 | -0.07470 | 0.52824 | -0.16555 | 0.23680 |
| STX16 | -0.00485 | 0.98590 | -0.67756 | 0.01395 |
| MGMT | 0.02779 | 0.94060 | 2.72750 | 0.00000 |

| | | | | |
|---|---|---|---|---|
| DDA1 | 0.30595 | 0.16471 | 0.59068 | 0.01057 |
| EPHA2 | 0.49105 | 0.01385 | 0.23704 | 0.31528 |
| TNFAIP8 | -0.21408 | 0.54551 | -0.51238 | 0.15989 |
| BRDT | 0.24799 | 0.26114 | 1.23827 | 0.00000 |
| ECM1 | 0.02290 | 0.85248 | 0.50863 | 0.00046 |
| PRL | 0.07590 | 0.79267 | 0.45832 | 0.16461 |
| NBN | 0.30106 | 0.44348 | 0.53715 | 0.14855 |
| NOTCH3 | 0.01636 | 0.90893 | -0.19257 | 0.21637 |
| NT5E | 0.49649 | 0.07693 | 0.09899 | 0.68621 |
| TNFRSF4 | 0.28789 | 0.06769 | -0.01484 | 0.94694 |
| ITGA11 | -0.65954 | 0.00022 | 0.00216 | 0.99032 |
| PCARE | 0.14648 | 0.60016 | 0.20004 | 0.49992 |
| IL12RB1 | 0.06007 | 0.71860 | 0.02753 | 0.86222 |
| MIA | 0.17580 | 0.34499 | -0.23903 | 0.17464 |
| CD209 | -0.09925 | 0.36208 | -0.24359 | 0.06337 |
| MAP2K6 | -0.26931 | 0.37171 | -0.29828 | 0.34638 |
| APOL1 | -0.02812 | 0.89807 | -0.45490 | 0.07599 |
| GNE | 0.05102 | 0.83945 | 1.16863 | 0.00000 |
| DNLZ | 0.61955 | 0.07853 | 0.56093 | 0.16272 |
| DPEP1 | -0.02246 | 0.89883 | -0.41526 | 0.03181 |
| AGR2 | 1.86716 | 0.00176 | 0.97394 | 0.06897 |
| SEZ6 | 0.19937 | 0.22630 | -0.08902 | 0.56956 |
| VWC2 | 0.17636 | 0.39063 | -0.08190 | 0.70413 |
| ACY1 | 0.16135 | 0.58987 | 0.32031 | 0.23881 |
| NFKB2 | 0.06361 | 0.81463 | -0.03430 | 0.89480 |
| CHCHD10 | 0.65975 | 0.02555 | 0.96118 | 0.00104 |
| CRX | 0.16559 | 0.67387 | -0.25295 | 0.51375 |
| LAYN | 0.35619 | 0.09378 | 0.06033 | 0.78369 |
| FYB1 | 0.20566 | 0.54885 | 3.07409 | 0.00000 |
| TAX1BP1 | 0.06177 | 0.81746 | 0.08105 | 0.74563 |
| MRPS16 | 0.24379 | 0.03408 | 0.14609 | 0.17493 |
| MUC2 | 0.04760 | 0.88792 | 0.06893 | 0.83387 |
| CLMP | -0.08235 | 0.55877 | -0.23958 | 0.23355 |
| TNFRSF17 | 0.27481 | 0.03174 | -0.21804 | 0.05182 |
| IL12A_IL12B | 0.13430 | 0.64496 | -0.27137 | 0.39270 |
| SCARB1 | 0.29894 | 0.31403 | 0.13401 | 0.60656 |
| PGM2 | -0.22670 | 0.42365 | 0.34978 | 0.25287 |
| NMRK2 | 0.50256 | 0.04579 | -0.28447 | 0.18284 |
| SKIV2L | -0.12908 | 0.66046 | -0.17121 | 0.45584 |
| SLC13A1 | -0.11833 | 0.59357 | 0.29641 | 0.10203 |
| SIGLEC5 | 0.23344 | 0.63212 | 0.93041 | 0.04303 |
| MCTS1 | -0.22682 | 0.46508 | 0.16384 | 0.49434 |
| SAT1 | -0.16619 | 0.47965 | -0.09757 | 0.62606 |
| UNC5D | 0.16196 | 0.22043 | 0.00131 | 0.99034 |
| METAP1D | 0.32511 | 0.38914 | 1.95284 | 0.00000 |
| SCRG1 | 0.30806 | 0.07251 | -0.03928 | 0.80073 |
| PCDH17 | -0.03893 | 0.84795 | 0.17942 | 0.35247 |
| CTBS | -0.07275 | 0.49753 | -0.10440 | 0.27507 |

| | | | | |
|---|---|---|---|---|
| IL13RA1 | -0.02797 | 0.75573 | -0.17762 | 0.05197 |
| LYN | 0.14136 | 0.58952 | 2.39681 | 0.00000 |
| TNFRSF1B | 0.48533 | 0.01661 | 0.31972 | 0.13454 |
| NACC1 | -0.06657 | 0.82017 | 1.07983 | 0.00009 |
| CSNK1D | 0.34372 | 0.39090 | 2.11893 | 0.00000 |
| LAP3 | 0.18505 | 0.54733 | 1.24953 | 0.00002 |
| TP73 | 0.24255 | 0.62544 | 0.44166 | 0.38761 |
| SBSN | -0.13907 | 0.25260 | -0.08386 | 0.49327 |
| LGALS9 | 0.42512 | 0.00759 | 0.48024 | 0.00816 |
| RAB27B | -0.12791 | 0.64050 | -0.49905 | 0.06938 |
| CRELD2 | 0.32779 | 0.06958 | 0.33622 | 0.08919 |
| SPARCL1 | -0.00386 | 0.97105 | -0.09054 | 0.44280 |
| DKK3 | 0.09798 | 0.45093 | -0.06011 | 0.69618 |
| NCAM2 | -0.07708 | 0.41855 | -0.26003 | 0.06346 |
| PLB1 | -0.29091 | 0.11970 | -0.83792 | 0.00018 |
| ORM1 | 0.23647 | 0.10165 | 0.22720 | 0.13632 |
| DCC | -0.07825 | 0.30633 | -0.21416 | 0.00530 |
| SEMA4C | -0.47592 | 0.04924 | -1.48354 | 0.00000 |
| CRYM | 0.44483 | 0.21092 | 0.80047 | 0.02188 |
| COL28A1 | -0.30039 | 0.16906 | 0.16926 | 0.46027 |
| USP47 | -0.08544 | 0.76177 | -0.08361 | 0.72772 |
| EXTL1 | -0.39378 | 0.14152 | -0.69957 | 0.00248 |
| IGFBP1 | 1.23886 | 0.00486 | 1.45702 | 0.00057 |
| SEL1L | 0.29821 | 0.10804 | 0.11361 | 0.49692 |
| IKBKG | -0.28602 | 0.30194 | -0.14214 | 0.62341 |
| APBB1IP | 0.16733 | 0.63388 | 0.32331 | 0.31188 |
| CDC26 | 0.29600 | 0.28254 | 0.95097 | 0.00037 |
| COL18A1 | 0.32623 | 0.00136 | -0.01851 | 0.86967 |
| CPVL | -0.38093 | 0.00693 | -0.18892 | 0.36198 |
| SUSD4 | 0.27726 | 0.12889 | 0.07343 | 0.69395 |
| EEF1D | -0.34309 | 0.37155 | -0.19211 | 0.58352 |
| HCG22 | 0.29314 | 0.25176 | -0.32353 | 0.16616 |
| NFASC | 0.04878 | 0.69591 | -0.21961 | 0.07072 |
| F3 | 0.04847 | 0.64871 | -0.08608 | 0.69687 |
| OMP | 0.12600 | 0.54641 | 0.52498 | 0.01004 |
| LYZL2 | 0.04525 | 0.76877 | -0.29260 | 0.03709 |
| HCLS1 | 0.14442 | 0.51333 | 1.37321 | 0.00000 |
| SCT | -0.12555 | 0.34610 | -0.39944 | 0.00373 |
| CFP | -0.16828 | 0.01911 | -0.10133 | 0.19074 |
| KLHL41 | -0.89254 | 0.00119 | -0.14164 | 0.51120 |
| SIRPB1 | 0.43008 | 0.01329 | 0.32638 | 0.08408 |
| UHRF2 | -0.01065 | 0.95779 | 0.19463 | 0.30203 |
| CD207 | -0.04012 | 0.80633 | -0.37684 | 0.03585 |
| BACH1 | 0.21222 | 0.36284 | 0.04597 | 0.82723 |
| TXNDC5 | 0.51791 | 0.03649 | 0.85088 | 0.00083 |
| MYL1 | -0.08882 | 0.75417 | 0.17217 | 0.43811 |
| ACSL1 | -0.12031 | 0.55942 | -0.27845 | 0.16270 |
| SHC1 | 0.40833 | 0.43229 | -0.34315 | 0.52905 |

| | | | | |
|---|---|---|---|---|
| ADAMTS4 | 0.53989 | 0.00455 | 0.55001 | 0.00921 |
| PPIE | 0.11236 | 0.78242 | -0.63539 | 0.12729 |
| WASL | 1.14875 | 0.01667 | 0.70050 | 0.16268 |
| PRDX1 | -0.18059 | 0.47371 | 0.74339 | 0.00317 |
| ANPEP | 0.03258 | 0.85114 | -0.07227 | 0.62732 |
| ADAM23 | -0.10256 | 0.48503 | 0.25354 | 0.28996 |
| BCAN | -0.10244 | 0.52814 | -0.24902 | 0.11270 |
| RWDD1 | -0.05300 | 0.82893 | 1.38307 | 0.00000 |
| EIF2S2 | 0.17667 | 0.60737 | 2.09484 | 0.00000 |
| ROBO4 | 0.11850 | 0.05323 | -0.05180 | 0.38516 |
| BABAM1 | -0.06933 | 0.68735 | 0.37570 | 0.05365 |
| MILR1 | 0.33608 | 0.10617 | 0.25163 | 0.20867 |
| ADAMTS8 | -0.32669 | 0.04491 | -0.46891 | 0.00605 |
| FKBPL | 0.15505 | 0.53971 | 1.40090 | 0.00000 |
| DYNLT3 | -0.08118 | 0.73116 | 0.61639 | 0.00676 |
| MCEMP1 | 0.06430 | 0.87519 | -0.19763 | 0.62313 |
| LEP | -1.25607 | 0.00156 | -0.57951 | 0.12973 |
| SARG | 0.34372 | 0.34879 | 2.51398 | 0.00000 |
| NAGK | -0.27748 | 0.29072 | 0.81711 | 0.00200 |
| CDC123 | 0.04766 | 0.88839 | -0.25179 | 0.39913 |
| CCNE1 | 0.37639 | 0.03937 | 0.26626 | 0.09255 |
| RICTOR | 0.09219 | 0.58644 | -0.06410 | 0.75441 |
| TFF1 | 1.15340 | 0.00017 | 0.62482 | 0.03079 |
| FKBP1B | -0.41768 | 0.35402 | -0.57573 | 0.18578 |
| ENOPH1 | -0.37113 | 0.04705 | -0.03085 | 0.89323 |
| TP53I3 | 0.08980 | 0.69936 | 1.15306 | 0.00000 |
| IL31RA | -0.15988 | 0.24778 | -0.36917 | 0.00865 |
| IL16 | -0.15960 | 0.42415 | 0.11483 | 0.63168 |
| PTGDS | 0.20072 | 0.18721 | -0.04569 | 0.75251 |
| ICAM5 | 0.20933 | 0.12195 | -0.15040 | 0.34130 |
| VWC2L | -0.14316 | 0.42155 | -0.47219 | 0.00034 |
| CEACAM19 | -0.05269 | 0.75615 | -0.15866 | 0.26375 |
| KEL | 0.34063 | 0.03739 | 0.29140 | 0.07344 |
| GGT1 | 0.16235 | 0.55589 | 0.20229 | 0.38165 |
| PLA2G10 | 0.53774 | 0.03122 | -0.58299 | 0.01822 |
| NCAN | -0.28482 | 0.04401 | -0.22079 | 0.19294 |
| COPB2 | 0.17325 | 0.58209 | -0.00831 | 0.97392 |
| CXCL1 | -0.02677 | 0.91548 | -0.33921 | 0.16282 |
| SMAD3 | 0.56467 | 0.11798 | 1.59422 | 0.00000 |
| FETUB | 0.08771 | 0.66559 | -0.15914 | 0.37088 |
| RELT | 0.24405 | 0.13140 | -0.10927 | 0.51482 |
| RNASE4 | 0.08199 | 0.58933 | 0.22554 | 0.08320 |
| GIGYF2 | -0.18638 | 0.55644 | 0.11375 | 0.69302 |
| FKBP7 | 0.13075 | 0.54219 | 0.62958 | 0.00352 |
| NFIC | 0.01196 | 0.94409 | 0.51814 | 0.00018 |
| TARS1 | 0.12869 | 0.35863 | 1.19679 | 0.00000 |
| LZTFL1 | -0.08156 | 0.76330 | 1.72239 | 0.00000 |
| MOCS2 | 0.20553 | 0.39595 | 0.98605 | 0.00000 |

| | | | | |
|---|---|---|---|---|
| VSIG10 | -0.00108 | 0.99374 | -0.13720 | 0.25844 |
| AREG | 0.74423 | 0.05940 | 0.48883 | 0.22101 |
| PTPRN2 | 0.38750 | 0.11023 | -0.02942 | 0.87056 |
| PSME2 | 0.14405 | 0.38844 | 1.11230 | 0.00000 |
| JAM3 | 0.51684 | 0.00736 | 0.20623 | 0.24473 |
| PON2 | 0.17947 | 0.26080 | 0.76453 | 0.00001 |
| PVALB | 0.15001 | 0.61656 | -0.08971 | 0.80436 |
| AKAP12 | -0.08984 | 0.32729 | -0.30829 | 0.00108 |
| LRIG1 | 0.22094 | 0.18348 | 0.23667 | 0.30078 |
| NIT1 | -0.05194 | 0.70179 | 1.54420 | 0.00000 |
| EIF2AK2 | -0.24393 | 0.60542 | 1.07388 | 0.01218 |
| SLITRK1 | -0.04274 | 0.75812 | -0.25104 | 0.07031 |
| GP2 | 0.59960 | 0.08950 | 0.41605 | 0.30356 |
| FSHB | 0.23011 | 0.20315 | 0.86442 | 0.00220 |
| ROR1 | 0.20568 | 0.10670 | -0.14676 | 0.33120 |
| RRM2B | -0.20996 | 0.44608 | 0.24638 | 0.34348 |
| COX6B1 | 0.20767 | 0.50038 | 1.01478 | 0.00075 |
| CPOX | 0.03817 | 0.79499 | 0.36232 | 0.00279 |
| TPK1 | 0.03323 | 0.82616 | 0.31210 | 0.01958 |
| EGF | 0.26742 | 0.44728 | -1.39657 | 0.00008 |
| SSBP1 | 0.16071 | 0.61191 | 0.72420 | 0.01171 |
| SERPINE2 | 0.16178 | 0.52236 | -1.36609 | 0.00000 |
| ROBO2 | -0.11904 | 0.28752 | -0.20342 | 0.10267 |
| TPSAB1 | 0.06874 | 0.69423 | 0.13523 | 0.55719 |
| MYCBP2 | 0.22040 | 0.36933 | -0.40431 | 0.07484 |
| IL31 | 0.23241 | 0.34897 | 0.21282 | 0.39263 |
| UBAC1 | 0.00991 | 0.93792 | 0.58888 | 0.00055 |
| MAP2K1 | 0.00338 | 0.99203 | 1.00446 | 0.00127 |
| SFTPA2 | 0.02755 | 0.90693 | -0.13871 | 0.59735 |
| SFTPD | 0.06547 | 0.73620 | -0.45236 | 0.01986 |
| EVI5 | -0.56531 | 0.07551 | 0.20464 | 0.51768 |
| MTR | 0.13111 | 0.52559 | 0.41853 | 0.02594 |
| STAU1 | 0.41351 | 0.17703 | 0.18904 | 0.46284 |
| BAG6 | -0.17386 | 0.19802 | -0.03412 | 0.82201 |
| LAT2 | -0.25653 | 0.50935 | 1.44488 | 0.00025 |
| EDF1 | -0.08609 | 0.83444 | 1.00515 | 0.00884 |
| TCN1 | 0.22210 | 0.31915 | 0.04409 | 0.79677 |
| FLT4 | 0.33269 | 0.01346 | -0.11576 | 0.34643 |
| IL22 | 0.85752 | 0.00289 | -0.10203 | 0.72179 |
| CD3G | 0.18876 | 0.56071 | 0.31059 | 0.34897 |
| ITPA | -0.00798 | 0.97830 | 1.35280 | 0.00000 |
| KLK11 | 0.10664 | 0.50544 | -0.00803 | 0.95410 |
| ACE2 | 0.79024 | 0.01692 | 0.19914 | 0.50061 |
| GPR158 | 0.22294 | 0.18678 | 0.33551 | 0.05112 |
| NCF2 | 0.57327 | 0.25528 | 0.56449 | 0.25288 |
| UXS1 | 0.14164 | 0.40680 | -0.08093 | 0.67464 |
| CSPG5 | 0.34363 | 0.06260 | 0.07463 | 0.68380 |
| CTNNA1 | 0.14680 | 0.45035 | -0.16244 | 0.32928 |

| | | | | |
|---|---|---|---|---|
| HMCN2 | -0.56909 | 0.02255 | -0.45274 | 0.05691 |
| FGA | 0.29417 | 0.30528 | 5.95859 | 0.00000 |
| FGF9 | -0.17739 | 0.62844 | -0.10763 | 0.74900 |
| TNFRSF10B | 0.67491 | 0.03887 | 0.40745 | 0.14665 |
| UROD | 0.14736 | 0.49362 | 1.80104 | 0.00000 |
| TOP1 | -0.26191 | 0.43487 | -0.62806 | 0.04389 |
| CEP20 | -0.43340 | 0.13546 | 0.72233 | 0.02051 |
| PRKG1 | 0.00936 | 0.98328 | 0.64631 | 0.11349 |
| AIF1L | 0.28150 | 0.05681 | 0.16600 | 0.26672 |
| HS6ST2 | -0.14323 | 0.42648 | -0.27361 | 0.10640 |
| FOSB | 0.23405 | 0.41708 | 0.33952 | 0.22290 |
| PMCH | 0.01468 | 0.92907 | 0.18110 | 0.32212 |
| NUDT5 | 0.27442 | 0.23184 | 1.21770 | 0.00000 |
| CXCL11 | 1.05061 | 0.00054 | 0.13154 | 0.64548 |
| PAFAH1B3 | -0.02920 | 0.91042 | 0.15663 | 0.47225 |
| RIDA | 0.12364 | 0.71830 | 1.24311 | 0.00006 |
| ACRV1 | 0.43786 | 0.22669 | 0.00862 | 0.96577 |
| GIPR | 0.41486 | 0.38364 | 0.30212 | 0.54334 |
| ABCA2 | 0.20300 | 0.25580 | 0.07924 | 0.62086 |
| KIRREL2 | 0.16322 | 0.31238 | -0.18985 | 0.17944 |
| HSPB6 | 0.08746 | 0.69621 | 0.05483 | 0.83928 |
| C9 | 0.32010 | 0.03324 | 0.33365 | 0.02433 |
| NPR1 | 0.15267 | 0.17207 | -0.00811 | 0.94218 |
| RGS8 | -0.20316 | 0.21377 | 0.00865 | 0.96378 |
| CD160 | 0.33781 | 0.05680 | -0.14947 | 0.49823 |
| ITGB7 | -0.25177 | 0.15262 | -0.25894 | 0.23062 |
| LAMA4 | -0.04140 | 0.76022 | -0.08153 | 0.54978 |
| IL1A | -0.34723 | 0.08783 | -0.16236 | 0.45309 |
| IL18BP | 0.23743 | 0.13072 | 0.22794 | 0.17444 |
| FABP2 | 0.23627 | 0.41459 | 0.18726 | 0.49464 |
| SORBS1 | 0.29461 | 0.24526 | 1.17819 | 0.00004 |
| GSTP1 | -0.33353 | 0.21592 | -0.38509 | 0.08901 |
| HNRNPK | 0.31025 | 0.47069 | 3.20060 | 0.00000 |
| FCRL2 | -0.20387 | 0.16783 | -0.39492 | 0.04967 |
| GJA8 | 0.50654 | 0.04004 | -0.29775 | 0.28507 |
| CCL18 | 0.51463 | 0.01410 | 0.23590 | 0.27982 |
| PECR | -0.05231 | 0.47265 | 0.10848 | 0.20368 |
| S100A3 | 0.00128 | 0.99436 | 0.22213 | 0.11316 |
| RBP5 | 0.27861 | 0.37129 | 0.59986 | 0.05663 |
| QPCT | 0.00540 | 0.95623 | 0.08979 | 0.40010 |
| ARNT | 0.19044 | 0.35989 | -0.02352 | 0.84138 |
| BCL2L15 | 0.39685 | 0.36852 | 0.24284 | 0.54694 |
| PI3 | 0.52808 | 0.02455 | -0.06117 | 0.77228 |
| SKAP2 | -0.44200 | 0.16448 | -0.17461 | 0.58762 |
| C7 | 0.24689 | 0.07582 | 0.06090 | 0.65679 |
| CCN3 | 0.22300 | 0.14064 | -0.03838 | 0.82955 |
| RBFOX3 | 0.96437 | 0.00483 | 1.14179 | 0.00022 |
| SIGLEC10 | 0.39435 | 0.11519 | 0.28230 | 0.23963 |

| | | | | |
|---|---|---|---|---|
| FDX2 | 0.82746 | 0.03288 | 1.10987 | 0.00012 |
| KIF22 | 0.57196 | 0.11726 | 1.97073 | 0.00000 |
| SERPINI1 | -0.00904 | 0.93918 | -0.71916 | 0.00000 |
| APOC1 | -0.14303 | 0.45359 | -0.17509 | 0.26146 |
| CTLA4 | 0.01153 | 0.96231 | -0.26128 | 0.34192 |
| BCAT1 | 0.11127 | 0.49464 | 0.46864 | 0.00529 |
| CASP3 | 0.05960 | 0.83845 | 1.40196 | 0.00000 |
| IL17A | 1.06570 | 0.00325 | -0.07521 | 0.80660 |
| CD84 | -0.06539 | 0.56833 | -0.47722 | 0.00256 |
| EREG | 0.35686 | 0.49221 | -1.77536 | 0.00019 |
| HGS | 0.11224 | 0.71666 | 1.73002 | 0.00000 |
| CHL1 | -0.02651 | 0.83693 | -0.24404 | 0.08027 |
| ELOA | 0.21695 | 0.59781 | 0.72275 | 0.07942 |
| EPHA10 | -0.04052 | 0.87604 | 0.57165 | 0.04831 |
| DBH | -0.55180 | 0.12097 | -0.07151 | 0.79328 |
| HK2 | -0.17340 | 0.67951 | 0.08991 | 0.82450 |
| PPT1 | 0.30162 | 0.19846 | -0.28230 | 0.16444 |
| TFF2 | 0.32606 | 0.23604 | -0.13963 | 0.57051 |
| ITGA5 | 0.07457 | 0.45842 | 0.86421 | 0.00000 |
| AFAP1 | -0.58218 | 0.33432 | 0.68639 | 0.16325 |
| CRNN | -0.11541 | 0.61572 | -0.19279 | 0.38024 |
| SSH3 | -0.11135 | 0.28657 | 0.05261 | 0.63586 |
| LRRC37A2 | 0.13994 | 0.35086 | -0.19888 | 0.12714 |
| CENPF | 0.10127 | 0.69170 | 1.28899 | 0.00000 |
| IL2 | 0.15071 | 0.62957 | -0.11148 | 0.67298 |
| ZPR1 | -0.24790 | 0.34819 | 0.03309 | 0.89505 |
| UMOD | -0.12115 | 0.42945 | -0.45040 | 0.00606 |
| ACP6 | 0.33184 | 0.06964 | 0.54196 | 0.04728 |
| CDKL5 | 0.25393 | 0.24287 | 0.08524 | 0.68241 |
| SKAP1 | 0.25250 | 0.48234 | 0.62142 | 0.08970 |
| OSBPL2 | -0.21469 | 0.21604 | 0.13918 | 0.47333 |
| RELB | 0.35769 | 0.28041 | 0.45565 | 0.29624 |
| MAPT | 0.53313 | 0.28171 | 0.85445 | 0.10594 |
| EPHB4 | 0.25086 | 0.06892 | 0.21707 | 0.14855 |
| NCK2 | 0.52794 | 0.14655 | 2.21467 | 0.00000 |
| NXPE4 | 0.10588 | 0.67125 | 0.16972 | 0.46448 |
| ANXA10 | 0.02395 | 0.94350 | 0.79190 | 0.04434 |
| DHPS | -0.01434 | 0.96092 | 0.61425 | 0.02217 |
| STX3 | -0.15458 | 0.16700 | 0.23938 | 0.08501 |
| IFNGR1 | 0.10030 | 0.36428 | 0.07278 | 0.48914 |
| MAP1LC3B2 | 0.09114 | 0.63779 | -0.20067 | 0.37020 |
| TPRKB | 0.15874 | 0.57782 | -0.03146 | 0.89437 |
| HS6ST1 | -0.16816 | 0.26920 | -0.11701 | 0.57224 |
| MUC16 | 0.89145 | 0.01914 | 0.50734 | 0.22395 |
| IL1B | 0.12453 | 0.78957 | 0.09202 | 0.84182 |
| CEACAM6 | -0.02289 | 0.93264 | 0.02099 | 0.91256 |
| IGBP1 | 0.00399 | 0.98784 | 2.08756 | 0.00000 |
| HGF | 0.51044 | 0.06920 | 0.30756 | 0.35696 |

| | | | | |
|---|---|---|---|---|
| RABEPK | -0.07081 | 0.75334 | 0.37719 | 0.09982 |
| BAMBI | 0.25952 | 0.09057 | -0.02014 | 0.85952 |
| CTRC | -0.35675 | 0.28415 | -0.90931 | 0.00761 |
| VSNL1 | 0.10177 | 0.71912 | 0.12104 | 0.62747 |
| NPTXR | -0.15628 | 0.36201 | -0.03229 | 0.80807 |
| CD48 | 0.04427 | 0.71196 | -0.01958 | 0.88539 |
| NRP1 | 0.16980 | 0.12188 | -0.08847 | 0.43086 |
| MYLPF | -0.32186 | 0.40366 | -0.05310 | 0.87380 |
| ACP1 | -0.42960 | 0.10398 | -0.20302 | 0.42992 |
| SSB | 0.18453 | 0.54265 | 0.28865 | 0.31042 |
| FHIP2A | -0.11772 | 0.59621 | -0.10050 | 0.59720 |
| SNCA | 0.95782 | 0.01659 | 2.47289 | 0.00000 |
| OSCAR | 0.39770 | 0.00275 | 0.06881 | 0.62346 |
| PODXL2 | -0.04126 | 0.75945 | 0.00527 | 0.97100 |
| RAB11FIP3 | -0.24492 | 0.51460 | 1.01246 | 0.00269 |
| RAB44 | 0.21159 | 0.68028 | 0.54345 | 0.26440 |
| TXNL1 | 0.11864 | 0.58427 | -0.45726 | 0.05839 |
| VAMP8 | -0.39989 | 0.22395 | -0.04131 | 0.89316 |
| LYPD1 | 0.24853 | 0.48661 | -0.23931 | 0.42105 |
| TRIM5 | -0.83225 | 0.00147 | -0.42997 | 0.09967 |
| RSPO1 | 0.36064 | 0.20817 | 0.23720 | 0.42337 |
| IL9 | 0.18254 | 0.39444 | -0.51848 | 0.02513 |
| KLKB1 | -0.08664 | 0.30038 | -0.15318 | 0.09285 |
| SPINK2 | 0.05504 | 0.70613 | 0.65324 | 0.00007 |
| SIGLEC15 | 0.00014 | 0.99938 | -0.21851 | 0.35842 |
| MBL2 | -0.03402 | 0.91420 | -0.22554 | 0.46204 |
| ADA2 | 0.22767 | 0.16000 | 0.01270 | 0.93727 |
| PRKAR2A | -0.17873 | 0.51581 | 0.47755 | 0.08446 |
| RAB10 | -0.03568 | 0.91658 | 0.47588 | 0.10867 |
| CHCHD6 | 0.64661 | 0.00610 | 0.50702 | 0.00276 |
| CHMP6 | 0.11250 | 0.56625 | 0.48898 | 0.00674 |
| SPTLC1 | -0.08858 | 0.71432 | 0.36511 | 0.12634 |
| PTPRH | 0.10616 | 0.64261 | 0.32145 | 0.16939 |
| CRH | -0.46990 | 0.21654 | -1.52452 | 0.00025 |
| CREG1 | -0.31578 | 0.06246 | -0.09643 | 0.56762 |
| PRTN3 | -0.22384 | 0.41681 | -0.32805 | 0.29605 |
| ESYT2 | -0.04719 | 0.87957 | 0.57857 | 0.06296 |
| GRPEL1 | 0.92932 | 0.01065 | 1.39006 | 0.00006 |
| MNDA | 0.51312 | 0.27271 | -0.09809 | 0.82692 |
| GASK1A | 0.03081 | 0.88996 | -0.52657 | 0.01358 |
| CYTH3 | 0.38257 | 0.14719 | 0.13769 | 0.62525 |
| FAM171A2 | 0.18247 | 0.67658 | -0.54399 | 0.20026 |
| EIF4EBP1 | 0.43632 | 0.10376 | 2.17157 | 0.00000 |
| MAN2B2 | -0.38773 | 0.05889 | -0.05430 | 0.77391 |
| SERPINF2 | -0.04639 | 0.57666 | 0.37911 | 0.00066 |
| GHR | -0.56163 | 0.00021 | -0.18382 | 0.23582 |
| CD248 | -0.09608 | 0.56351 | -0.15408 | 0.35274 |
| IGSF21 | -0.49947 | 0.00318 | -0.75468 | 0.00001 |

| | | | | |
|---|---|---|---|---|
| STAB2 | 0.07157 | 0.44487 | -0.04848 | 0.61651 |
| ZHX2 | 0.03058 | 0.91445 | 0.44487 | 0.15319 |
| EVI2B | 0.50708 | 0.06294 | 0.21552 | 0.33950 |
| SNED1 | 0.00598 | 0.97050 | 0.64445 | 0.00046 |
| PMS1 | 0.76986 | 0.02638 | 0.49547 | 0.11197 |
| FABP1 | 0.71229 | 0.15619 | 1.40681 | 0.00230 |
| CLC | -0.02052 | 0.95632 | -0.03475 | 0.91124 |
| LPA | 0.94896 | 0.02044 | 0.65666 | 0.11111 |
| CTRL | 0.07214 | 0.81337 | -0.15535 | 0.60713 |
| GPIHBP1 | 0.18544 | 0.09951 | 0.26642 | 0.03054 |
| GPI | -0.18681 | 0.52195 | 1.26149 | 0.00000 |
| SCRN1 | -0.31942 | 0.31505 | 0.30783 | 0.32508 |
| FGFBP3 | 0.19187 | 0.29324 | 0.34085 | 0.02521 |
| GBP6 | 0.10886 | 0.72705 | 0.27442 | 0.45091 |
| SOX2 | 0.29997 | 0.27076 | 0.19210 | 0.34440 |
| RAB33A | -0.49360 | 0.09964 | -0.94693 | 0.00053 |
| BTD | -0.21672 | 0.02653 | -0.14264 | 0.14894 |
| SHD | 0.35564 | 0.38996 | 0.61033 | 0.05289 |
| WASF3 | -0.15712 | 0.72060 | 1.64497 | 0.00055 |
| BTN1A1 | 0.25255 | 0.11309 | -0.26810 | 0.12303 |
| OSTN | -0.01330 | 0.94178 | -0.03543 | 0.87124 |
| BLOC1S3 | -0.00492 | 0.98145 | 0.54221 | 0.00205 |
| DEFB118 | -0.05086 | 0.87392 | -0.21808 | 0.46814 |
| IL3RA | -0.07476 | 0.47415 | -0.12503 | 0.14976 |
| CA11 | 0.04718 | 0.67686 | -0.03457 | 0.76444 |
| FGFBP2 | -0.43630 | 0.03814 | -0.34015 | 0.08515 |
| HDAC8 | -0.60022 | 0.02659 | 0.17788 | 0.55816 |
| FADD | -0.29706 | 0.36412 | 1.28377 | 0.00002 |
| THY1 | 0.08347 | 0.57601 | -0.10760 | 0.47388 |
| CACYBP | -0.20892 | 0.53484 | 0.74341 | 0.01938 |
| SMAD1 | -0.11677 | 0.67253 | 0.77093 | 0.00221 |
| GFAP | 0.37840 | 0.12933 | 0.37449 | 0.12039 |
| ITGA2 | -0.19578 | 0.26583 | -0.48282 | 0.00804 |
| NAGPA | -0.14695 | 0.12791 | -0.31096 | 0.00158 |
| IL36G | 0.05888 | 0.78896 | -0.00894 | 0.96526 |
| HSD17B3 | 1.24172 | 0.00147 | 0.77160 | 0.00876 |
| BLOC1S2 | -0.55105 | 0.26759 | -0.50169 | 0.18655 |
| IMPG1 | 0.11537 | 0.41408 | 0.05363 | 0.61488 |
| RNASE3 | -0.80599 | 0.12457 | -1.93968 | 0.00101 |
| LONP1 | 0.70687 | 0.08463 | 1.72095 | 0.00000 |
| ARID3A | -0.10990 | 0.16655 | -0.07636 | 0.40575 |
| PTPRZ1 | 0.00233 | 0.98854 | -0.13068 | 0.38027 |
| IGDCC3 | -0.09302 | 0.57716 | -0.44537 | 0.00716 |
| IL1R2 | -0.15225 | 0.13006 | -0.13831 | 0.19277 |
| GFER | 0.74713 | 0.01803 | 1.86644 | 0.00000 |
| THBS2 | 0.28855 | 0.27065 | 0.06460 | 0.78688 |
| PHLDB2 | -0.82709 | 0.06855 | -0.09086 | 0.82925 |
| MAP1LC3A | 0.11153 | 0.52894 | 0.15100 | 0.35070 |

| | | | | |
|---|---|---|---|---|
| UBE2B | -0.02406 | 0.88215 | 0.64809 | 0.00014 |
| PRELP | 0.01728 | 0.88529 | 0.01775 | 0.89525 |
| DAPK2 | -0.14393 | 0.77399 | 1.30229 | 0.00626 |
| SUSD5 | 0.42208 | 0.00048 | 0.22140 | 0.06589 |
| RBP2 | 0.33620 | 0.24328 | 0.45850 | 0.08432 |
| DBI | -0.05242 | 0.85767 | 0.74325 | 0.01137 |
| DTX3 | 0.32348 | 0.02685 | 0.28025 | 0.05407 |
| ALDH3A1 | 0.35655 | 0.34896 | 1.04256 | 0.00615 |
| LRRC38 | -0.01094 | 0.96222 | 0.20895 | 0.40768 |
| KIR3DL2 | 0.00236 | 0.99272 | -0.12267 | 0.61048 |
| DSCAM | 0.01169 | 0.93963 | 0.10833 | 0.43998 |
| NECTIN2 | 0.38952 | 0.01310 | 0.16603 | 0.33391 |
| NINJ1 | 0.33966 | 0.12555 | 0.68502 | 0.00170 |
| S100G | -0.00583 | 0.97940 | -0.29893 | 0.11850 |
| PPCDC | -0.02916 | 0.87540 | 1.17723 | 0.00000 |
| STK11 | 0.64161 | 0.01640 | 1.91400 | 0.00000 |
| C5 | 0.06560 | 0.32778 | -0.14772 | 0.02779 |
| FBN2 | 0.33949 | 0.18123 | -0.10883 | 0.64797 |
| GPR101 | -0.33388 | 0.26869 | -0.29210 | 0.30916 |
| CCN4 | 0.37136 | 0.03054 | -0.23212 | 0.30325 |
| HMOX2 | -0.02656 | 0.92651 | 0.44197 | 0.10369 |
| SEZ6L | 0.06142 | 0.66136 | -0.13432 | 0.25517 |
| CEP43 | 0.25090 | 0.32975 | 0.86160 | 0.00062 |
| DLL4 | 0.32007 | 0.13711 | 0.39851 | 0.07663 |
| MORF4L1 | 0.52262 | 0.26994 | 1.10517 | 0.01206 |
| SPINK1 | 0.50995 | 0.12910 | 0.33805 | 0.26320 |
| MPHOSPH8 | 0.01719 | 0.95897 | 1.23926 | 0.00008 |
| ECHS1 | 0.72465 | 0.10318 | 2.08080 | 0.00000 |
| KLRK1 | 0.02164 | 0.88007 | -0.11928 | 0.34547 |
| FAM3D | 0.58774 | 0.00843 | -0.37085 | 0.06222 |
| HSDL2 | -0.14363 | 0.57270 | 0.38530 | 0.07366 |
| PGF | 0.31979 | 0.06549 | 0.04524 | 0.85561 |
| GHRHR | -0.43293 | 0.05155 | -0.00415 | 0.98796 |
| ACTA2 | 0.84120 | 0.00015 | 0.72853 | 0.00400 |
| IL15RA | 0.28703 | 0.07114 | 0.04096 | 0.79160 |
| PRG3 | -0.07803 | 0.64904 | -0.04664 | 0.76255 |
| CSNK2A1 | 0.13969 | 0.47514 | 0.84667 | 0.00004 |
| ARHGEF12 | 0.28627 | 0.33560 | 0.26147 | 0.33149 |
| CCS | -0.32390 | 0.14579 | 1.64153 | 0.00000 |
| CD8A | 0.18753 | 0.36255 | 0.18600 | 0.37224 |
| EPN1 | -0.22107 | 0.39839 | 0.60288 | 0.03720 |
| MPO | -0.13253 | 0.60653 | -0.70352 | 0.01504 |
| PDCD5 | 0.27025 | 0.23944 | 1.58342 | 0.00000 |
| PAM | -0.06415 | 0.59322 | -0.32153 | 0.00864 |
| CXCL6 | 0.10333 | 0.65436 | -0.30094 | 0.32096 |
| AXIN1 | 0.16075 | 0.58423 | 0.85602 | 0.00190 |
| HHEX | -0.05384 | 0.85725 | 0.70334 | 0.00738 |
| TMPRSS5 | -0.09457 | 0.61132 | -0.21242 | 0.25172 |

| | | | | |
|---|---|---|---|---|
| PTS | 0.35754 | 0.38589 | 1.62751 | 0.00003 |
| AGRN | 0.31728 | 0.10491 | 0.10725 | 0.55402 |
| PQBP1 | 0.25813 | 0.43888 | 0.86688 | 0.00396 |
| NME1 | 0.23670 | 0.05444 | 0.00316 | 0.97734 |
| USP8 | 0.11983 | 0.72136 | 1.15947 | 0.00030 |
| PFDN2 | 0.07356 | 0.68947 | 0.07018 | 0.68353 |
| PRSS53 | -0.24123 | 0.07137 | -0.60868 | 0.00000 |
| CCL13 | 0.09565 | 0.62273 | -0.42553 | 0.12176 |
| SEMA4D | 0.08420 | 0.44725 | -0.55197 | 0.00000 |
| PLA2G7 | -0.02895 | 0.80593 | 0.01775 | 0.90209 |
| ARF6 | -1.02033 | 0.00722 | 0.96567 | 0.00897 |
| GLI2 | 0.08531 | 0.30102 | -0.06742 | 0.40685 |
| GGACT | -0.14066 | 0.61818 | 0.91507 | 0.00031 |
| ALPP | 0.26925 | 0.54111 | -0.95393 | 0.00968 |
| ADGRD1 | 0.27105 | 0.11867 | -0.06360 | 0.65886 |
| TNFRSF6B | 0.54582 | 0.08627 | 0.08662 | 0.79239 |
| YES1 | 0.17564 | 0.64071 | 0.59605 | 0.09395 |
| CST5 | 0.17997 | 0.44024 | -0.37657 | 0.15391 |
| PPP1R12B | 0.44648 | 0.34225 | 1.12620 | 0.00298 |
| NRGN | 0.37185 | 0.29926 | 1.77985 | 0.00000 |
| JMJD1C | 0.00399 | 0.98388 | -0.16873 | 0.42597 |
| SLK | 0.00016 | 0.99968 | -0.20650 | 0.60729 |
| GFRAL | 0.01067 | 0.96142 | -0.01396 | 0.93685 |
| SLC39A14 | 0.28598 | 0.28970 | 0.25166 | 0.34605 |
| VPS28 | 0.21218 | 0.40198 | 0.74431 | 0.00450 |
| CHRDL1 | 0.07354 | 0.60655 | 0.78002 | 0.00014 |
| IL21R | 0.11197 | 0.56177 | -0.25084 | 0.15835 |
| CD300LF | 0.22206 | 0.33968 | 0.26677 | 0.36311 |
| CAMKK1 | 0.25015 | 0.35338 | 1.28318 | 0.00000 |
| EIF5 | 0.08347 | 0.75601 | 1.37376 | 0.00000 |
| MANEAL | 0.05969 | 0.75402 | 0.17125 | 0.32418 |
| TFAP2A | -0.06108 | 0.86424 | 0.23050 | 0.55793 |
| HSD11B1 | 0.15560 | 0.26377 | -0.34747 | 0.00966 |
| SNX9 | 0.15683 | 0.51155 | 0.83296 | 0.00057 |
| FGF20 | -0.03115 | 0.83219 | 0.07094 | 0.49499 |
| CD34 | -0.18483 | 0.09873 | -0.36851 | 0.00143 |
| CMIP | -0.00668 | 0.98398 | 0.12755 | 0.67729 |
| PODXL | -0.04320 | 0.51157 | -0.22497 | 0.30130 |
| CD99L2 | 0.06535 | 0.50852 | 0.02096 | 0.86126 |
| MAN1A2 | 0.03916 | 0.75486 | -0.29447 | 0.00979 |
| TOP2B | 0.34849 | 0.34560 | 0.49750 | 0.16827 |
| RAB6B | -0.07923 | 0.59097 | -0.59477 | 0.00025 |
| NPDC1 | 0.35237 | 0.06049 | 0.42937 | 0.02427 |
| NFKBIE | 0.22045 | 0.40397 | 0.69363 | 0.00890 |
| IL6R | 0.23402 | 0.02634 | 0.06772 | 0.55118 |
| CRHR1 | 0.47915 | 0.02662 | -0.02525 | 0.92564 |
| CALCOCO1 | -0.10349 | 0.78066 | -0.73693 | 0.04606 |
| GCG | 0.27312 | 0.54292 | -0.53795 | 0.30136 |

| | | | | |
|---|---|---|---|---|
| CEACAM5 | 1.00914 | 0.02201 | 0.88288 | 0.01751 |
| SMAD5 | -0.20099 | 0.14979 | -0.27645 | 0.08792 |
| FCRL6 | 0.35900 | 0.08088 | -0.36018 | 0.10883 |
| IGFBP7 | 0.17976 | 0.14580 | 0.00630 | 0.96364 |
| RBP1 | 0.74989 | 0.05415 | 0.82483 | 0.04704 |
| SPINK4 | 0.53518 | 0.04401 | 0.33036 | 0.16937 |
| GOPC | 0.01963 | 0.95226 | 2.22046 | 0.00000 |
| FNTA | -0.07843 | 0.66714 | 0.55998 | 0.00294 |
| ING1 | -0.06151 | 0.84577 | 0.59653 | 0.06189 |
| SMPD1 | 0.27436 | 0.07378 | 0.17665 | 0.27242 |
| SNRPB2 | 0.49344 | 0.32118 | 0.56659 | 0.25137 |
| SIRT5 | -0.34332 | 0.08199 | -0.10462 | 0.63315 |
| ARHGAP45 | 0.05560 | 0.84945 | 0.71675 | 0.00468 |
| PARK7 | 0.04761 | 0.82393 | 1.38023 | 0.00000 |
| AP3S2 | 0.09896 | 0.70774 | 0.03504 | 0.87688 |
| BAP18 | -0.01373 | 0.97286 | 0.49314 | 0.22025 |
| ATP6V1G1 | -0.44082 | 0.14804 | 1.26809 | 0.00001 |
| DNAJC6 | 0.67264 | 0.05874 | 2.02702 | 0.00000 |
| ITGB5 | 0.19753 | 0.33993 | 0.85027 | 0.00004 |
| PSMD5 | -0.04780 | 0.77818 | 0.54696 | 0.00101 |
| IFT20 | -0.14819 | 0.54010 | -0.19134 | 0.37595 |
| H2AP | 0.38230 | 0.12541 | 0.01320 | 0.94711 |
| GIPC2 | 0.59745 | 0.00904 | 0.11380 | 0.58061 |
| CDC25A | 0.05204 | 0.83627 | -0.27178 | 0.15293 |
| HSPG2 | 0.02291 | 0.87930 | -0.08160 | 0.59095 |
| PARP1 | 0.63446 | 0.10716 | 1.70586 | 0.00002 |
| OTUD7B | 0.35699 | 0.12680 | 1.33217 | 0.00000 |
| PRSS22 | 0.11265 | 0.70353 | 0.16332 | 0.35608 |
| PDCD1LG2 | 0.36810 | 0.00205 | -0.03902 | 0.79190 |
| CA7 | -0.14381 | 0.55370 | 0.05660 | 0.83130 |
| CDH23 | 0.20405 | 0.13475 | -1.20999 | 0.00000 |
| FCER2 | -0.29497 | 0.08470 | -0.28718 | 0.13843 |
| CRTAM | 0.14032 | 0.45618 | -0.16474 | 0.41417 |
| SLC51B | 0.04519 | 0.82585 | -0.08308 | 0.72503 |
| PKD1 | 0.10535 | 0.37082 | -0.10994 | 0.33111 |
| STK4 | 0.26453 | 0.41063 | 2.17926 | 0.00000 |
| MYOC | -0.54802 | 0.01377 | -0.65328 | 0.00415 |
| ITIH1 | -0.09754 | 0.23731 | -0.33259 | 0.00021 |
| NECAP2 | -0.09641 | 0.71706 | 1.39250 | 0.00000 |
| FCGR2B | -0.08004 | 0.73216 | -0.15382 | 0.52904 |
| PFDN6 | 0.23893 | 0.62957 | -0.03233 | 0.94825 |
| B3GNT7 | -0.03329 | 0.77657 | -0.24791 | 0.04338 |
| DCN | 0.19152 | 0.10889 | -0.06405 | 0.62919 |
| CEP152 | -0.26274 | 0.44896 | -0.04182 | 0.90821 |
| POMC | -0.22183 | 0.55338 | 1.02946 | 0.03127 |
| CEACAM16 | -1.56094 | 0.00000 | -0.48035 | 0.06867 |
| PCDH12 | -0.05723 | 0.57819 | -0.41102 | 0.00001 |
| TCOF1 | 0.50184 | 0.12378 | 0.84262 | 0.00222 |

| | | | | |
|---|---|---|---|---|
| FKBP4 | 0.22458 | 0.22709 | 0.95655 | 0.00000 |
| MEP1B | -1.12039 | 0.00026 | -0.03790 | 0.90462 |
| CPA4 | -0.26925 | 0.12020 | -0.44154 | 0.00546 |
| APEX1 | 0.14955 | 0.60933 | 0.57536 | 0.11020 |
| IL15 | -0.14467 | 0.35778 | -0.05280 | 0.78596 |
| DDHD2 | 0.02215 | 0.91451 | 0.84740 | 0.00001 |
| CRISP2 | 0.11711 | 0.51331 | -0.51987 | 0.06070 |
| CLPP | 0.78623 | 0.03147 | 1.82315 | 0.00000 |
| CLEC11A | -0.05519 | 0.71252 | -0.05791 | 0.72063 |
| IL18R1 | 0.21767 | 0.17479 | 0.05822 | 0.80383 |
| MAEA | -0.17146 | 0.45014 | 0.37579 | 0.08874 |
| VSTM2L | -0.06787 | 0.73140 | -0.20182 | 0.27682 |
| NECTIN4 | 0.39648 | 0.08095 | 0.05350 | 0.78018 |
| TRAF3IP2 | -0.10042 | 0.68421 | -0.15947 | 0.50097 |
| EPGN | 0.17562 | 0.24045 | 0.02502 | 0.84437 |
| TCN2 | 0.13265 | 0.23236 | 0.11160 | 0.37271 |
| PTPN9 | -0.28041 | 0.21783 | 0.46287 | 0.07066 |
| IRAK1 | -0.09236 | 0.69385 | 0.16794 | 0.44342 |
| RGMB | -0.08045 | 0.53080 | 0.03163 | 0.79242 |
| SHISA5 | 0.26664 | 0.21527 | 0.24899 | 0.18804 |
| ROBO1 | -0.00339 | 0.98178 | -0.14339 | 0.32690 |
| ZBTB17 | 0.16909 | 0.46737 | 0.91197 | 0.00024 |
| SCARA5 | 0.12863 | 0.47912 | -0.08631 | 0.62466 |
| CXCL3 | -0.26640 | 0.38326 | -0.16965 | 0.52305 |
| PDCD1 | 0.17108 | 0.24152 | -0.00169 | 0.99319 |
| VCPKMT | 0.59588 | 0.02256 | -0.07928 | 0.78682 |
| MET | 0.00085 | 0.99264 | -0.01692 | 0.87499 |
| RILP | 0.26928 | 0.22949 | 2.05158 | 0.00000 |
| TDRKH | 0.51719 | 0.09884 | 1.42505 | 0.00001 |
| FCN2 | 0.15480 | 0.40019 | -0.16191 | 0.42036 |
| CXCL17 | 0.59466 | 0.00780 | 0.37589 | 0.15601 |
| GUCA2A | 0.34424 | 0.02421 | -0.01678 | 0.92342 |
| SYTL4 | 0.19074 | 0.64243 | 0.69045 | 0.07630 |
| NDRG1 | -0.27196 | 0.21111 | 0.67527 | 0.00287 |
| SPRR1B | 1.26506 | 0.07712 | -0.36861 | 0.59235 |
| CD93 | -0.00169 | 0.98909 | -0.23649 | 0.08998 |
| NFE2 | -0.15602 | 0.60314 | 0.81604 | 0.00300 |
| GDF2 | -0.15360 | 0.36346 | -0.46491 | 0.01809 |
| PDXDC1 | 0.20259 | 0.16974 | 0.04256 | 0.78570 |
| MKI67 | 0.53937 | 0.22797 | 1.00647 | 0.02349 |
| METAP1 | -0.08884 | 0.42169 | -0.18675 | 0.05412 |
| KLK13 | -0.42479 | 0.02190 | -0.52992 | 0.01855 |
| FGF23 | 0.76626 | 0.08661 | 1.85782 | 0.00004 |
| ERBB2 | -0.09717 | 0.48840 | -0.10313 | 0.28604 |
| SNX2 | 0.49381 | 0.10741 | 2.56410 | 0.00000 |
| MARCO | 0.23069 | 0.02926 | 0.01811 | 0.88068 |
| SMOC2 | 0.06505 | 0.66961 | -0.05609 | 0.72772 |
| ANGPTL2 | -0.03537 | 0.86076 | 0.43567 | 0.02834 |

| | | | | |
|---|---|---|---|---|
| CD5 | 0.08250 | 0.54471 | -0.18695 | 0.38605 |
| PAIP2B | 0.07317 | 0.81639 | 0.27027 | 0.28065 |
| KLK15 | -0.16663 | 0.30613 | 0.02573 | 0.85686 |
| THBD | 0.02464 | 0.84905 | -0.05176 | 0.70226 |
| RAP1A | 0.00256 | 0.99376 | -0.15202 | 0.63837 |
| HADH | 0.07725 | 0.79703 | 0.18832 | 0.47748 |
| PENK | 0.00903 | 0.94988 | -0.12160 | 0.45902 |
| DRAXIN | -0.00289 | 0.98933 | 0.11296 | 0.60393 |
| CA5A | 0.46020 | 0.40962 | 1.39451 | 0.00273 |
| PCBD1 | 0.08135 | 0.80562 | 1.16977 | 0.00005 |
| GRIN2B | 0.13014 | 0.54030 | 0.13258 | 0.57369 |
| CABP2 | -0.07657 | 0.79047 | 0.09763 | 0.70662 |
| ZP4 | 0.19242 | 0.46543 | -0.47898 | 0.06613 |
| CDKN1A | 0.30124 | 0.37524 | 1.52071 | 0.00010 |
| ADAM12 | 0.84443 | 0.00511 | 0.05230 | 0.83707 |
| VAT1 | -0.17758 | 0.14732 | 0.13406 | 0.31345 |
| DENR | 0.35573 | 0.14982 | 1.45736 | 0.00000 |
| LILRA4 | 0.12733 | 0.60761 | -0.01882 | 0.93904 |
| PROC | 0.06865 | 0.57827 | -0.14781 | 0.28676 |
| MTDH | 0.63896 | 0.05781 | 2.02683 | 0.00000 |
| LGALS3BP | 0.24044 | 0.10065 | 0.25387 | 0.07604 |
| FH | -0.05743 | 0.89956 | 0.23697 | 0.55069 |
| CEP164 | -0.01489 | 0.94797 | 0.27677 | 0.15807 |
| ASPSCR1 | -0.03795 | 0.83826 | 1.15330 | 0.00000 |
| MPRIP | -0.05222 | 0.70326 | 0.10779 | 0.39606 |
| OPHN1 | 0.38952 | 0.25489 | 1.98935 | 0.00000 |
| CELA3A | 0.06173 | 0.82907 | -0.34858 | 0.19034 |
| DIPK1C | 0.26119 | 0.14176 | -0.24493 | 0.35224 |
| MSLN | 0.33528 | 0.12137 | -0.74523 | 0.00139 |
| C9orf40 | 0.16907 | 0.48836 | 2.00552 | 0.00000 |
| THBS4 | -0.10857 | 0.62454 | -0.57570 | 0.00983 |
| MED21 | 0.06835 | 0.72478 | 0.27499 | 0.07631 |
| C1RL | 0.05188 | 0.52932 | -0.11146 | 0.26525 |
| DLGAP5 | 0.49118 | 0.07512 | -0.12171 | 0.61997 |
| TNFRSF8 | 0.40756 | 0.02010 | 0.13883 | 0.50342 |
| CASP8 | -0.68019 | 0.06519 | 0.28518 | 0.44957 |
| MYH4 | 0.25509 | 0.44680 | -0.42273 | 0.09947 |
| OMD | -0.43275 | 0.00583 | 0.10801 | 0.53855 |
| GCNT1 | 0.31600 | 0.02309 | -0.07112 | 0.70600 |
| IL4 | 0.80852 | 0.16892 | -1.17483 | 0.03075 |
| CD244 | 0.01901 | 0.85969 | -0.04486 | 0.76463 |
| HTRA2 | 0.23244 | 0.35629 | 1.15863 | 0.00000 |
| CD28 | 0.14909 | 0.33942 | -0.43074 | 0.00970 |
| DAB2 | 0.34153 | 0.29207 | 2.12331 | 0.00000 |
| ARG2 | 0.66226 | 0.01470 | 0.32863 | 0.18049 |
| SOX9 | 1.00869 | 0.00786 | 1.15367 | 0.00366 |
| AHNAK | 0.17548 | 0.30220 | 0.39791 | 0.01439 |
| IFNAR1 | -0.09818 | 0.18122 | -0.00816 | 0.91050 |

| | | | | |
|---|---|---|---|---|
| F12 | -0.04370 | 0.70489 | -0.10578 | 0.41005 |
| RPA2 | -0.10113 | 0.69714 | 0.31798 | 0.17526 |
| SLAMF6 | 0.40533 | 0.02637 | 0.14935 | 0.42817 |
| POSTN | 0.38890 | 0.02034 | 0.08056 | 0.62105 |
| CA2 | 0.26780 | 0.31279 | 1.85847 | 0.00000 |
| GFOD2 | -0.24258 | 0.40734 | -0.06896 | 0.82603 |
| KLK3 | 0.46211 | 0.22879 | -0.09795 | 0.51330 |
| ENO1 | -0.02057 | 0.93514 | 2.13382 | 0.00000 |
| TEX101 | -0.32386 | 0.28017 | -0.32896 | 0.16077 |
| CSH1 | 0.07496 | 0.79899 | 0.41038 | 0.05411 |
| CTSF | 0.34370 | 0.03762 | 0.21172 | 0.22527 |
| TRIM26 | -0.27841 | 0.29031 | 0.09302 | 0.75652 |
| DSG3 | -0.16689 | 0.25805 | -0.54991 | 0.01071 |
| CD177 | 0.30188 | 0.48707 | -0.04235 | 0.92171 |
| SNAP25 | 0.28272 | 0.23650 | 0.06399 | 0.72383 |
| FAM13A | 0.41115 | 0.16542 | 0.70196 | 0.01002 |
| YJU2 | 0.12320 | 0.56961 | 0.57680 | 0.00624 |
| CD86 | -0.17867 | 0.15664 | -0.40124 | 0.00221 |
| CANT1 | 0.13819 | 0.15949 | -0.21540 | 0.05301 |
| LRTM2 | -0.01635 | 0.87485 | -0.01485 | 0.91176 |
| ARSB | -0.31506 | 0.22115 | -0.74988 | 0.01156 |
| MST1 | -0.13035 | 0.33927 | 0.03361 | 0.83900 |
| JAM2 | 0.00820 | 0.94703 | -0.03165 | 0.80851 |
| LEFTY2 | 0.30477 | 0.15542 | -0.50661 | 0.05045 |
| ICAM1 | 0.34912 | 0.03292 | 0.08476 | 0.62291 |
| SPINK6 | -0.12627 | 0.48167 | -0.02765 | 0.89259 |
| CYP24A1 | 0.07165 | 0.82335 | -0.02554 | 0.92749 |
| CRHBP | -0.10673 | 0.37543 | -0.07031 | 0.64299 |
| ITGB1BP1 | 0.26563 | 0.32974 | 0.39125 | 0.10925 |
| TK1 | 0.20914 | 0.34535 | 0.36547 | 0.18929 |
| CNTNAP4 | 0.33253 | 0.17808 | 0.18018 | 0.46383 |
| CHRM1 | -0.14124 | 0.51316 | -0.30209 | 0.19999 |
| CAPG | 0.01507 | 0.96283 | 0.63503 | 0.04073 |
| SETMAR | -0.16987 | 0.44071 | 0.52204 | 0.02021 |
| PTPRK | 0.23546 | 0.05142 | -0.01420 | 0.87665 |
| TBR1 | 0.15190 | 0.45904 | -0.03964 | 0.85781 |
| GLRX | -0.04721 | 0.83214 | 0.95791 | 0.00005 |
| TNFSF13B | 0.19543 | 0.19981 | -0.01107 | 0.94774 |
| THTPA | -0.00878 | 0.97660 | 1.26554 | 0.00000 |
| TIMP1 | 0.45710 | 0.01299 | 0.01952 | 0.92054 |
| NPHS2 | 0.16571 | 0.61900 | 0.32173 | 0.29660 |
| CCL14 | 0.30192 | 0.02319 | -0.05367 | 0.69292 |
| MTSS2 | 0.49809 | 0.13241 | 1.92095 | 0.00000 |
| SUSD1 | 0.35302 | 0.09301 | 0.05056 | 0.80865 |
| PON3 | -0.55899 | 0.01333 | -0.16194 | 0.39877 |
| PRTFDC1 | 0.12617 | 0.76260 | 2.71103 | 0.00000 |
| SCN3A | 0.03003 | 0.92564 | -0.75225 | 0.01173 |
| LAG3 | 0.19295 | 0.25018 | -0.04097 | 0.83146 |

| | | | | |
|---|---|---|---|---|
| FARSA | 0.16150 | 0.67830 | 1.17379 | 0.00076 |
| CGN | 0.77555 | 0.00926 | 0.73809 | 0.00343 |
| DUOX2 | 0.39054 | 0.11851 | -0.31653 | 0.29630 |
| RNF4 | 0.15090 | 0.79837 | -0.48419 | 0.42569 |
| IZUMO1 | 0.35426 | 0.08992 | -0.11311 | 0.55854 |
| GMPR2 | -0.09885 | 0.69965 | 1.60892 | 0.00000 |
| DEFB116 | 0.51837 | 0.03660 | -0.16412 | 0.49091 |
| TP53 | 0.22767 | 0.44006 | 0.56669 | 0.05687 |
| SUMF2 | -0.11200 | 0.51088 | 0.01720 | 0.91924 |
| FEN1 | -0.05695 | 0.91151 | -0.15925 | 0.76708 |
| SIGLEC1 | 0.46223 | 0.01798 | 0.19851 | 0.32666 |
| DCTPP1 | 0.20924 | 0.36806 | 0.16101 | 0.43609 |
| SLC44A4 | 0.12280 | 0.69268 | 0.12927 | 0.68837 |
| DIABLO | 0.17411 | 0.66850 | 1.85414 | 0.00001 |
| PDAP1 | 0.38536 | 0.25004 | 2.12147 | 0.00000 |
| TACC3 | -0.58004 | 0.13026 | 0.52331 | 0.14679 |
| SELP | -0.03554 | 0.86714 | -0.21067 | 0.32345 |
| SERPINI2 | 0.01790 | 0.93789 | -0.06547 | 0.75747 |
| SPTBN2 | 0.10050 | 0.62977 | 0.72487 | 0.00004 |
| ADAM22 | 0.03691 | 0.84580 | -0.35359 | 0.11650 |
| COL2A1 | -0.23002 | 0.42126 | -1.08005 | 0.00013 |
| FGL1 | 1.00153 | 0.00218 | 2.19805 | 0.00000 |
| SERPINB5 | -0.02385 | 0.95173 | -0.61239 | 0.07727 |
| GCC1 | 0.01262 | 0.96965 | 1.69411 | 0.00000 |
| CPE | -0.09144 | 0.45985 | -0.33330 | 0.01551 |
| ESAM | 0.20493 | 0.11172 | -0.21799 | 0.07587 |
| CXCL16 | 0.05489 | 0.62872 | -0.03098 | 0.79882 |
| MAPKAPK2 | -0.26360 | 0.36164 | 0.17820 | 0.55044 |
| CASP10 | -0.21213 | 0.54376 | 0.70607 | 0.04558 |
| ATG4A | 0.23563 | 0.30980 | 1.60342 | 0.00000 |
| RBM17 | 0.28341 | 0.38888 | 1.89001 | 0.00000 |
| NUDT2 | -0.70550 | 0.00239 | 0.23480 | 0.25891 |
| ELAVL4 | 0.58826 | 0.12973 | 0.54888 | 0.02066 |
| ENPP6 | -0.49635 | 0.00688 | -0.75190 | 0.00000 |
| ICAM4 | -0.03638 | 0.84080 | -0.38018 | 0.03863 |
| FRMD4B | -0.10734 | 0.72623 | -0.01285 | 0.96369 |
| STEAP4 | -0.01316 | 0.95917 | -0.59838 | 0.03297 |
| FLT3LG | 0.25860 | 0.14162 | -0.41268 | 0.04768 |
| IL1RL1 | 0.56894 | 0.09991 | 0.53391 | 0.13582 |
| TBCB | -0.08280 | 0.80993 | 1.52220 | 0.00004 |
| DEFA1B; DEFA1 | 0.09329 | 0.76401 | 0.23800 | 0.43616 |
| HAO1 | 0.27865 | 0.58262 | 1.60581 | 0.00042 |
| BMPER | -0.03316 | 0.76883 | -0.11436 | 0.27762 |
| UBXN1 | 0.05518 | 0.82644 | 1.88961 | 0.00000 |
| HSP90B1 | 0.07537 | 0.74248 | -0.19781 | 0.47221 |
| EFHD1 | 0.46863 | 0.04328 | 0.28244 | 0.23718 |
| TNFSF10 | -0.11962 | 0.37734 | -0.48095 | 0.03775 |
| ANGPTL1 | 0.16078 | 0.24294 | 0.12464 | 0.38651 |

| | | | | |
|---|---|---|---|---|
| TPT1 | -0.10250 | 0.78527 | -0.37237 | 0.25675 |
| CES2 | -0.17042 | 0.36902 | -1.74786 | 0.00000 |
| INSL3 | -0.04699 | 0.91360 | -0.25109 | 0.40448 |
| ISM1 | 0.11729 | 0.44793 | 0.01002 | 0.95809 |
| SMAD2 | 0.51411 | 0.16704 | 1.75560 | 0.00000 |
| HBZ | -0.16393 | 0.68604 | 0.95250 | 0.01046 |
| NT5C1A | 0.82020 | 0.03130 | 1.76421 | 0.00001 |
| MYOM2 | -0.57566 | 0.19339 | -0.13996 | 0.71301 |
| ATP5F1D | 0.95090 | 0.01027 | 0.35772 | 0.26063 |
| HPCAL1 | 0.03413 | 0.88415 | 1.25951 | 0.00003 |
| CSF2RA | 0.08119 | 0.66071 | 0.03258 | 0.86311 |
| ARTN | 0.05061 | 0.77416 | 0.12985 | 0.37071 |
| GMPR | 0.36791 | 0.09355 | 2.35376 | 0.00000 |
| RPL14 | 0.36285 | 0.10424 | 1.07874 | 0.00000 |
| BMP10 | -0.11585 | 0.31579 | -0.12031 | 0.31867 |
| CLSTN3 | -0.20244 | 0.17529 | -0.34128 | 0.00660 |
| SHH | 0.09158 | 0.53000 | -0.06525 | 0.55662 |
| FSTL1 | 0.06991 | 0.42812 | -0.01186 | 0.87720 |
| CFC1 | -0.14298 | 0.39675 | -0.07856 | 0.62154 |
| DAAM1 | 0.22779 | 0.46253 | 0.71373 | 0.01639 |
| SORD | -0.05398 | 0.89504 | 2.62302 | 0.00000 |
| LPO | -0.13862 | 0.52281 | 0.20830 | 0.38615 |
| AHCY | -0.27907 | 0.32942 | 0.90803 | 0.00125 |
| PDLIM7 | -0.40099 | 0.29720 | 0.47597 | 0.21761 |
| CEBPA | 0.03285 | 0.86683 | 0.53731 | 0.02302 |
| PILRB | 0.31259 | 0.07854 | 0.22246 | 0.23190 |
| NDUFB7 | 0.03739 | 0.90425 | 0.93969 | 0.00095 |
| NELL1 | -0.49368 | 0.00540 | -0.30972 | 0.04205 |
| NLGN2 | 0.20842 | 0.38200 | -0.13151 | 0.55986 |
| YTHDF3 | 0.33365 | 0.32594 | 2.15259 | 0.00000 |
| ADAMTSL4 | 0.45443 | 0.00781 | 0.46584 | 0.00334 |
| NPL | 0.30741 | 0.26437 | 0.46594 | 0.06306 |
| STXBP3 | -0.22468 | 0.45902 | 1.63000 | 0.00000 |
| TMED1 | -0.38550 | 0.57804 | 0.15802 | 0.81911 |
| CTSC | -0.36539 | 0.04012 | -0.64361 | 0.00051 |
| LAMP2 | -0.10834 | 0.20651 | -0.17287 | 0.07810 |
| MTSS1 | -0.24552 | 0.51357 | -1.29354 | 0.00019 |
| SSC5D | 0.23946 | 0.12498 | 0.21016 | 0.16854 |
| SCGN | 0.36359 | 0.06984 | 0.11694 | 0.44916 |
| DPP7 | -0.18218 | 0.46973 | -0.34480 | 0.10437 |
| INSL4 | 0.37954 | 0.30226 | 0.22951 | 0.36260 |
| BGLAP | -0.39882 | 0.36262 | -0.39670 | 0.36999 |
| APOD | -0.00874 | 0.94670 | -0.24951 | 0.04016 |
| SEMA3F | -0.04930 | 0.70405 | 0.12946 | 0.32650 |
| CLEC7A | 0.72662 | 0.00109 | 0.20049 | 0.35032 |
| VSIR | -0.35202 | 0.25678 | -0.42196 | 0.19932 |
| TREML2 | 0.02728 | 0.84049 | -0.51551 | 0.00015 |
| AHSA1 | 0.20202 | 0.34334 | 1.25218 | 0.00000 |

| | | | | |
|---|---|---|---|---|
| NFU1 | 0.36759 | 0.35073 | 2.03739 | 0.00000 |
| IL18RAP | -0.43943 | 0.27176 | 0.19123 | 0.67076 |
| SCP2 | 0.06964 | 0.84354 | 0.38479 | 0.19233 |
| SULT1A1 | 0.17395 | 0.64581 | 1.56704 | 0.00013 |
| DCLRE1C | 0.28392 | 0.07743 | -0.28941 | 0.11788 |
| CACNA1C | 0.39198 | 0.17389 | -0.22311 | 0.25747 |
| LILRB5 | -0.08979 | 0.70399 | 0.10131 | 0.66010 |
| CLINT1 | 0.36874 | 0.11334 | -0.05318 | 0.84648 |
| LAMTOR5 | 0.05633 | 0.85432 | 0.77219 | 0.01023 |
| ATP1B4 | 0.14635 | 0.37708 | -0.25565 | 0.13867 |
| ASGR2 | 0.38859 | 0.02740 | 0.09007 | 0.57370 |
| CLEC14A | 0.10787 | 0.43845 | 0.05597 | 0.69252 |
| BCAT2 | 0.02295 | 0.92315 | 0.08927 | 0.70061 |
| GPR15L | 0.14590 | 0.51516 | -0.04114 | 0.82787 |
| IQGAP2 | -0.92037 | 0.01191 | -0.97009 | 0.00362 |
| CPA2 | 0.20181 | 0.51887 | -0.70638 | 0.01923 |
| ANXA3 | 0.07155 | 0.81455 | 1.43661 | 0.00001 |
| KAZN | -0.06518 | 0.84996 | 0.46245 | 0.15857 |
| TXN | -0.09696 | 0.65666 | 0.74721 | 0.00006 |
| VEGFA | 0.42831 | 0.05443 | -0.22277 | 0.42975 |
| DDX53 | -0.03932 | 0.71557 | 0.03262 | 0.81521 |
| ZNRD2 | 0.69606 | 0.05122 | 0.85186 | 0.00454 |
| F13B | -0.06232 | 0.28934 | -0.00240 | 0.97775 |
| IL10 | 0.46851 | 0.37121 | 1.61800 | 0.00333 |
| FRMD7 | 0.15665 | 0.21953 | 0.00793 | 0.95230 |
| AOC3 | -0.02671 | 0.81821 | -0.31229 | 0.01383 |
| HMMR | -0.11747 | 0.71429 | -0.10946 | 0.68572 |
| ERMAP | 0.36474 | 0.09130 | 0.28023 | 0.09192 |
| CIT | 0.65783 | 0.04536 | 1.21862 | 0.00000 |
| DNAJB6 | -0.68151 | 0.02374 | -0.35910 | 0.20820 |
| COQ7 | 0.13410 | 0.55217 | 0.59954 | 0.00167 |
| RAB3GAP1 | 0.08794 | 0.70863 | -0.05939 | 0.82663 |
| DUSP3 | -0.63987 | 0.06139 | -0.05931 | 0.86692 |
| TXNDC9 | 0.38299 | 0.23556 | 1.25767 | 0.00003 |
| RTKN2 | -0.00365 | 0.99089 | -0.60433 | 0.04491 |
| NMT1 | 0.03838 | 0.90629 | 1.66080 | 0.00000 |
| STX1B | 0.13941 | 0.41786 | 0.05870 | 0.70050 |
| DPP6 | -0.09722 | 0.48293 | -0.28760 | 0.07237 |
| HEPACAM2 | 0.15609 | 0.34072 | 0.07716 | 0.62610 |
| SLAMF8 | 0.60585 | 0.03457 | 0.28178 | 0.29483 |
| POLR2A | 0.16071 | 0.26572 | -0.13011 | 0.51897 |
| PCOLCE | 0.06076 | 0.71254 | -0.19813 | 0.22222 |
| LRPAP1 | 0.26186 | 0.16141 | 0.17509 | 0.35461 |
| ITGB1 | -0.08605 | 0.35272 | -0.10765 | 0.39333 |
| RECK | -0.08490 | 0.32672 | -0.31040 | 0.00025 |
| LAMB1 | 0.16192 | 0.18855 | 0.02228 | 0.84526 |
| GPD1 | -0.20565 | 0.54724 | 0.03889 | 0.90253 |
| AKR1C4 | 0.24110 | 0.54488 | 0.76170 | 0.06342 |

| | | | | |
|---|---|---|---|---|
| ATP6V1D | 0.00698 | 0.95568 | 0.24055 | 0.01888 |
| CCDC80 | 0.26028 | 0.12414 | 0.09922 | 0.66455 |
| GIP | 0.50036 | 0.00370 | -0.02508 | 0.85773 |
| MFAP3L | -0.37369 | 0.12840 | -0.24099 | 0.32761 |
| ITGB2 | -0.23072 | 0.07117 | 0.21758 | 0.15168 |
| FGF7 | -0.04643 | 0.84884 | 0.26752 | 0.26074 |
| CDON | -0.27173 | 0.08108 | -0.37575 | 0.02914 |
| INPP1 | -0.18002 | 0.49221 | 0.96733 | 0.00267 |
| NFAT5 | 0.56030 | 0.13786 | -0.10560 | 0.71241 |
| PDGFA | 0.18999 | 0.42392 | -0.82465 | 0.00015 |
| TBC1D23 | -0.47051 | 0.12047 | -0.60766 | 0.03364 |
| AMIGO1 | -0.10492 | 0.64612 | -0.11465 | 0.67471 |
| STX7 | 0.22773 | 0.20629 | -0.04748 | 0.78630 |
| CDHR5 | 0.01103 | 0.92982 | -0.00430 | 0.97571 |
| MN1 | 0.24560 | 0.53021 | -0.30388 | 0.42320 |
| LYPLA2 | -0.23739 | 0.51269 | -0.43289 | 0.17972 |
| TSNAX | -0.11844 | 0.57970 | 0.85138 | 0.00001 |
| TIMD4 | 0.27715 | 0.18185 | 0.17077 | 0.41220 |
| HSPA1A | -0.30846 | 0.29762 | 1.40013 | 0.00000 |
| TXK | 0.03943 | 0.86196 | -0.39595 | 0.09313 |
| NPPB | 2.05783 | 0.00052 | 1.56936 | 0.00313 |
| DFFA | 0.14817 | 0.56266 | 1.30608 | 0.00003 |
| THOP1 | 0.17607 | 0.40039 | 0.49198 | 0.00700 |
| IGSF8 | 0.46169 | 0.00354 | 0.38468 | 0.01345 |
| MICB_MICA | -0.02737 | 0.94875 | 0.83853 | 0.06693 |
| CRTAC1 | -0.06248 | 0.69949 | -0.61823 | 0.00023 |
| NME3 | -0.09778 | 0.38691 | -0.09575 | 0.43426 |
| GPRC5C | 0.93391 | 0.03381 | 0.48465 | 0.23898 |
| DIPK2B | -0.17935 | 0.10852 | 0.02767 | 0.77173 |
| CWC15 | 0.16589 | 0.64208 | 1.07824 | 0.00272 |
| LILRA2 | 0.07745 | 0.55246 | 0.04497 | 0.73992 |
| GP1BA | 0.00774 | 0.94876 | -0.25541 | 0.05802 |
| PITHD1 | 0.48299 | 0.10852 | -0.08063 | 0.77264 |
| CD74 | 0.51442 | 0.00092 | 0.31398 | 0.06755 |
| CINP | -0.26667 | 0.43701 | -0.03400 | 0.92313 |
| ATP1B3 | 0.22721 | 0.24190 | 0.21907 | 0.22922 |
| ALPI | 0.41269 | 0.34954 | -0.90071 | 0.03311 |
| SEZ6L2 | 0.42258 | 0.01364 | -0.25706 | 0.22518 |
| ICAM2 | 0.26939 | 0.03528 | 0.03547 | 0.79008 |
| HIP1 | -0.33161 | 0.10086 | 0.19502 | 0.33965 |
| KLK4 | 1.59509 | 0.00010 | 0.45314 | 0.16588 |
| AHNAK2 | 0.12067 | 0.35837 | 0.14926 | 0.33621 |
| FCRL1 | -0.23596 | 0.10366 | -0.48565 | 0.00080 |
| CCN2 | 0.23061 | 0.14432 | -0.22480 | 0.15526 |
| DGCR6 | 0.25656 | 0.31735 | 0.03936 | 0.86925 |
| CEBPB | -0.00678 | 0.98109 | -0.52036 | 0.06122 |
| LEPR | 0.33141 | 0.03410 | -0.07548 | 0.58119 |
| DDT | 0.05674 | 0.78356 | 1.47109 | 0.00000 |

| | | | | |
|---|---|---|---|---|
| ITGA6 | 0.35379 | 0.05029 | 0.84691 | 0.00003 |
| CHEK2 | -0.21947 | 0.50936 | 0.21191 | 0.54978 |
| NPM1 | 0.04574 | 0.90636 | 0.52867 | 0.15085 |
| DNAJB1 | 0.15156 | 0.61439 | 2.44334 | 0.00000 |
| SEMA7A | 0.02877 | 0.84165 | -0.04082 | 0.77298 |
| SPARC | 0.25828 | 0.24692 | -0.86831 | 0.00005 |
| SCARF1 | 0.16916 | 0.36334 | -0.46713 | 0.01437 |
| PLG | -0.03682 | 0.72228 | -0.11961 | 0.30268 |
| TARBP2 | 0.17137 | 0.56821 | 0.94873 | 0.00035 |
| STAM | -0.60170 | 0.04694 | -0.85929 | 0.00173 |
| NFYA | 0.60052 | 0.05142 | 0.47061 | 0.09474 |
| RNF5 | 0.03193 | 0.90568 | 0.89805 | 0.00094 |
| ACHE | 0.23275 | 0.32711 | 0.03707 | 0.86306 |
| RASGRF1 | -0.12201 | 0.63979 | -0.49698 | 0.06751 |
| SSNA1 | 0.81951 | 0.00897 | 1.66466 | 0.00000 |
| ATXN2L | 0.14586 | 0.13124 | 0.03489 | 0.70512 |
| GBA | 0.28741 | 0.38015 | -0.07178 | 0.77777 |
| FBLN2 | 0.26646 | 0.16237 | 0.51182 | 0.00854 |
| NSFL1C | 0.02888 | 0.90214 | 0.92843 | 0.00018 |
| PDZD2 | 0.10310 | 0.72196 | -0.03404 | 0.89860 |
| CCL21 | 0.13103 | 0.56892 | 0.37451 | 0.09465 |
| GAGE2A | 0.46540 | 0.10857 | -0.10088 | 0.72860 |
| PGLYRP4 | 0.19227 | 0.49603 | -0.15905 | 0.53391 |
| CLEC5A | 0.35348 | 0.12388 | 0.19021 | 0.36257 |
| PTK7 | 0.75652 | 0.00066 | -0.18278 | 0.43161 |
| ENG | -0.01464 | 0.84698 | -0.13829 | 0.16553 |
| TP53INP1 | 0.04155 | 0.78094 | -0.13421 | 0.38689 |
| NEDD9 | 0.30764 | 0.52069 | 0.63976 | 0.09592 |
| VGF | 0.04225 | 0.75280 | -0.21499 | 0.07866 |
| PTPRM | -0.06216 | 0.51094 | -0.21652 | 0.09017 |
| TINAGL1 | 0.32187 | 0.00673 | 0.18608 | 0.20012 |
| PSME1 | -0.03735 | 0.84362 | 0.92653 | 0.00003 |
| DUSP13 | -0.88600 | 0.00025 | -0.48034 | 0.02754 |
| MAG | 0.03869 | 0.66010 | -0.10406 | 0.31409 |
| LARP1 | 0.42095 | 0.10219 | 1.01441 | 0.00003 |
| PFDN4 | 0.46554 | 0.10363 | 0.55363 | 0.02113 |
| CAMSAP1 | 0.11294 | 0.75908 | 1.24288 | 0.00013 |
| NFATC1 | 0.02352 | 0.92153 | -0.28112 | 0.17131 |
| SEC31A | 0.05830 | 0.77934 | 1.83699 | 0.00000 |
| IL36A | 0.27282 | 0.49660 | 0.49583 | 0.24424 |
| SMTN | 0.07536 | 0.84919 | -1.51257 | 0.00041 |
| TSPAN8 | 0.60316 | 0.09524 | 0.57783 | 0.08282 |
| PRAP1 | 0.39187 | 0.00643 | 0.81231 | 0.00001 |
| OPTC | 0.22460 | 0.14542 | -0.03756 | 0.87052 |
| SIGLEC8 | 0.50518 | 0.01959 | 0.40914 | 0.04201 |
| PLA2G15 | 0.05226 | 0.61110 | -0.00040 | 0.99725 |
| TJAP1 | -0.36267 | 0.19817 | 0.15073 | 0.54985 |
| STX4 | -0.70769 | 0.03422 | -0.67906 | 0.03844 |

| | | | | |
|---|---|---|---|---|
| TNFRSF21 | 0.27020 | 0.04063 | -0.02117 | 0.89039 |
| DOK1 | 0.12391 | 0.75575 | 2.29096 | 0.00000 |
| IL10RA | 0.27889 | 0.17564 | 0.05512 | 0.73109 |
| TSPAN1 | 0.54253 | 0.08093 | -0.03391 | 0.86521 |
| PSIP1 | 0.35135 | 0.33086 | 1.06604 | 0.00234 |
| PIK3AP1 | -0.06838 | 0.80324 | 0.56521 | 0.02505 |
| ANKRD54 | -0.01170 | 0.95330 | 0.59552 | 0.00260 |
| GLO1 | 0.39733 | 0.07772 | -0.95187 | 0.00210 |
| TNFRSF13B | 0.09550 | 0.45078 | -0.17351 | 0.24028 |
| CTSD | 0.14382 | 0.33164 | 0.18526 | 0.18566 |
| PCDH9 | 0.10046 | 0.33247 | -0.31296 | 0.01727 |
| FUT8 | 0.15604 | 0.47503 | -0.88361 | 0.00005 |
| ESR1 | 0.29783 | 0.40165 | 0.23532 | 0.47711 |
| SCGB1A1 | 0.05109 | 0.75393 | -0.08671 | 0.63416 |
| HLA-A | 0.19205 | 0.20010 | 0.11039 | 0.44830 |
| ASAH2 | -0.55913 | 0.02460 | -0.80959 | 0.00105 |
| KDR | -0.05693 | 0.56834 | -0.09620 | 0.47083 |
| LCN2 | 0.33335 | 0.19417 | 0.46366 | 0.06484 |
| THPO | -0.00508 | 0.97431 | -0.65207 | 0.00029 |
| TYRO3 | -0.05075 | 0.65340 | -0.15544 | 0.20421 |
| CD27 | 0.58463 | 0.00193 | 0.23019 | 0.21574 |
| CASP1 | 0.05476 | 0.92146 | 0.67883 | 0.16445 |
| TEX33 | 0.42153 | 0.50458 | -0.25710 | 0.65621 |
| OTOA | -0.37671 | 0.15631 | -0.10574 | 0.64907 |
| SLC4A1 | 0.18377 | 0.61932 | 1.06976 | 0.00208 |
| IL34 | 0.01039 | 0.95908 | 0.26617 | 0.11357 |
| RPGR | 0.01769 | 0.94469 | -0.35447 | 0.16585 |
| IL17RB | 0.41689 | 0.01362 | 0.40774 | 0.03148 |
| TIMP4 | 0.28835 | 0.06128 | 0.22814 | 0.15374 |
| PALM | 0.08962 | 0.57565 | 0.31211 | 0.05630 |
| CDSN | 0.06396 | 0.72886 | -0.35248 | 0.04253 |
| CD2 | 0.09078 | 0.58202 | -0.14545 | 0.37011 |
| THAP12 | -0.55626 | 0.07184 | 0.50165 | 0.14115 |
| CTSH | 0.01750 | 0.93824 | -0.38089 | 0.11659 |
| DGKA | -0.07226 | 0.79182 | 0.00932 | 0.96970 |
| ENPP5 | -0.65451 | 0.00001 | -0.47821 | 0.00060 |
| PRKAR1A | -0.41819 | 0.12274 | 1.22672 | 0.00004 |
| TREM2 | 0.49885 | 0.02286 | 0.35205 | 0.08843 |
| ADGRV1 | 0.00378 | 0.99292 | 0.33418 | 0.44568 |
| VIT | -0.30606 | 0.06581 | -0.02202 | 0.89316 |
| REG1A | 0.54905 | 0.03026 | -0.11227 | 0.64777 |
| RTBDN | 0.26439 | 0.08094 | -0.06692 | 0.64633 |
| SLC1A4 | 0.54632 | 0.24795 | 1.27520 | 0.01493 |
| CDK5RAP3 | 0.32712 | 0.29165 | 1.52601 | 0.00001 |
| VWA5A | 0.40391 | 0.24779 | 0.68668 | 0.05661 |
| BGN | -0.08838 | 0.84022 | 0.99592 | 0.02762 |
| IL12RB2 | 0.05254 | 0.64678 | -0.48521 | 0.00047 |
| DNAJB8 | -0.09120 | 0.53551 | -0.38853 | 0.00042 |

| | | | | |
|---|---|---|---|---|
| XG | 0.00129 | 0.99412 | -0.03568 | 0.82751 |
| UGDH | 0.14888 | 0.67697 | 0.01878 | 0.95092 |
| C7orf50 | 0.18467 | 0.41512 | 0.35989 | 0.06787 |
| AK1 | 0.07478 | 0.75499 | 1.25260 | 0.00000 |
| LMOD2 | 0.46935 | 0.18749 | -0.14899 | 0.61661 |
| TRIM24 | 0.07847 | 0.78386 | 0.35815 | 0.19246 |
| GRAP2 | 0.65661 | 0.14512 | 2.91040 | 0.00000 |
| NPTN | 0.18627 | 0.49902 | -0.76701 | 0.00888 |
| PRND | 0.03981 | 0.85605 | -0.44570 | 0.03463 |
| KIR2DL3 | 0.41796 | 0.08628 | 0.23170 | 0.37983 |
| CD200 | -0.11176 | 0.25765 | -0.25760 | 0.04939 |
| CCL8 | 0.02843 | 0.89610 | -0.52065 | 0.07989 |
| SLIRP | 0.77389 | 0.01945 | 0.58200 | 0.08400 |
| GH1 | 1.66162 | 0.00020 | -0.04857 | 0.91124 |
| SCARF2 | 0.52794 | 0.00042 | 0.13188 | 0.39277 |
| GNAS | -0.35896 | 0.15820 | 0.65881 | 0.00492 |
| ITM2A | 0.48421 | 0.00306 | 0.48468 | 0.00941 |
| BCAM | 0.08494 | 0.54031 | 0.08447 | 0.49426 |
| HIP1R | 0.31720 | 0.35734 | 0.57964 | 0.07143 |
| ENO2 | 0.27485 | 0.30836 | 1.89694 | 0.00000 |
| SERPINA7 | 0.03096 | 0.75198 | -0.04599 | 0.64640 |
| PLSCR3 | -0.47322 | 0.02342 | -0.48515 | 0.02342 |
| CSF3 | -0.11068 | 0.81313 | 0.39256 | 0.38641 |
| DNPEP | -0.08550 | 0.72490 | 2.25159 | 0.00000 |
| LAT | 0.40540 | 0.08723 | 1.77312 | 0.00000 |
| PAFAH2 | 0.15428 | 0.61425 | 1.19604 | 0.00004 |
| ITGAL | -0.40080 | 0.15318 | 0.37529 | 0.13625 |
| BOLA1 | 0.32077 | 0.16566 | 1.24696 | 0.00000 |
| CCT5 | 0.05794 | 0.71084 | 0.86759 | 0.00000 |
| PRKAB1 | -0.24329 | 0.27150 | -0.54623 | 0.01058 |
| SELE | -0.06054 | 0.75501 | -0.20232 | 0.26280 |
| PLCB1 | 0.07727 | 0.48120 | -0.11088 | 0.21952 |
| INPP5J | 0.23640 | 0.06592 | 0.53071 | 0.00005 |
| SPP1 | 0.54923 | 0.00582 | 0.82658 | 0.00062 |
| NTF4 | 0.20481 | 0.20613 | 0.11643 | 0.55361 |
| PCNA | 0.18575 | 0.34967 | -0.34122 | 0.14582 |
| JCHAIN | 0.27233 | 0.04829 | -0.27782 | 0.05400 |
| COLEC12 | 0.24974 | 0.13369 | -0.01690 | 0.91432 |
| SIRPA | 0.31581 | 0.01411 | 0.04439 | 0.74764 |
| PACS2 | 0.11987 | 0.62448 | 1.28941 | 0.00000 |
| MAP2 | 0.83617 | 0.00549 | 0.83511 | 0.00115 |
| CDAN1 | 0.15247 | 0.03510 | -0.04097 | 0.64218 |
| CETN3 | -0.10879 | 0.69675 | 0.97845 | 0.00025 |
| B2M | 0.32821 | 0.04757 | 0.03804 | 0.80689 |
| QSOX1 | 0.12061 | 0.15965 | -0.18905 | 0.04130 |
| IFNLR1 | 0.09496 | 0.42566 | -0.20405 | 0.17824 |
| SLC12A2 | 0.18258 | 0.31146 | 0.35460 | 0.02692 |
| RGL2 | -0.78049 | 0.03496 | 0.15326 | 0.68329 |

| | | | | |
|---|---|---|---|---|
| ACOT13 | 0.34398 | 0.43910 | 2.98159 | 0.00000 |
| FGD3 | -0.47413 | 0.16188 | 1.04857 | 0.00175 |
| MRI1 | 0.03144 | 0.87257 | 2.04583 | 0.00000 |
| DCTN6 | 0.11409 | 0.61697 | 0.83963 | 0.00028 |
| NTRK2 | -0.36392 | 0.00552 | -0.34488 | 0.00580 |
| TRDMT1 | 0.67911 | 0.02646 | 1.60255 | 0.00000 |
| ADRA2A | 0.23901 | 0.42381 | 0.04534 | 0.88347 |
| C2orf69 | -0.14896 | 0.55930 | 0.71522 | 0.00325 |
| ASRGL1 | -0.12330 | 0.72537 | 0.97267 | 0.00347 |
| LEO1 | 0.66236 | 0.04241 | 0.72491 | 0.00532 |
| SAMD9L | -0.27491 | 0.37042 | 0.28877 | 0.35437 |
| DECR1 | 0.87220 | 0.02085 | 1.88197 | 0.00000 |
| WNT9A | 0.66300 | 0.00004 | 1.19669 | 0.00000 |
| IL24 | 0.37012 | 0.19506 | 0.09305 | 0.74561 |
| KCTD5 | -0.23969 | 0.36417 | 0.69635 | 0.00815 |
| FAM172A | -0.32529 | 0.19275 | 0.99031 | 0.00007 |
| DCTD | 0.07784 | 0.80277 | 1.27953 | 0.00004 |
| MEPE | -0.19365 | 0.09995 | -0.01910 | 0.90652 |
| STAMBP | 0.14073 | 0.58496 | 1.41897 | 0.00000 |
| MSTN | -1.18586 | 0.00036 | -0.86253 | 0.00477 |
| CDKN2D | -0.23503 | 0.43263 | 0.69565 | 0.05209 |
| CD72 | 0.16393 | 0.35402 | 0.52890 | 0.00581 |
| VPS4B | -0.35513 | 0.31067 | 1.07827 | 0.00244 |
| IPCEF1 | -0.05213 | 0.89684 | 0.92025 | 0.01117 |
| DPT | -0.28025 | 0.06637 | 0.22112 | 0.21244 |
| ARHGAP5 | 0.46183 | 0.14975 | 0.33896 | 0.23463 |
| LILRB4 | 0.48810 | 0.03058 | 0.79830 | 0.00122 |
| KCNC4 | 0.37452 | 0.10856 | -0.12509 | 0.57469 |
| TNFAIP2 | -0.31282 | 0.20886 | 0.54883 | 0.04884 |
| OLFM4 | 0.73127 | 0.12692 | -1.31002 | 0.00567 |
| BRAP | 0.10161 | 0.73759 | 1.62692 | 0.00000 |
| SERPINA5 | -0.16704 | 0.40766 | -0.46017 | 0.02272 |
| TCL1B | -0.00334 | 0.98862 | 0.11489 | 0.65126 |
| DUSP29 | -0.08013 | 0.78180 | 0.25011 | 0.44497 |
| CD3D | 0.06274 | 0.66490 | 0.02585 | 0.88443 |
| LGMN | -0.05459 | 0.69739 | -0.26450 | 0.04497 |
| PTPRF | 0.06212 | 0.53439 | 0.09577 | 0.45468 |
| HSBP1 | 0.16006 | 0.57878 | 1.75211 | 0.00000 |
| PNPT1 | 0.02946 | 0.91936 | -0.06375 | 0.81361 |
| VIPR1 | 0.15463 | 0.14157 | -0.26923 | 0.01915 |
| REEP4 | 0.60077 | 0.02583 | 0.80528 | 0.00142 |
| SAT2 | -0.09886 | 0.77577 | 1.16799 | 0.00018 |
| WFIKKN1 | -0.39803 | 0.02583 | -0.38793 | 0.02371 |
| TARM1 | -0.44496 | 0.29329 | -0.07585 | 0.84525 |
| PDP1 | 0.59844 | 0.08703 | 1.13200 | 0.00049 |
| WASF1 | 0.43003 | 0.10535 | 0.72936 | 0.00099 |
| IGSF3 | -0.09905 | 0.66030 | 0.02504 | 0.91086 |
| IL5RA | 0.20492 | 0.33882 | -0.17701 | 0.45650 |

| | | | | |
|---|---|---|---|---|
| TADA3 | 0.02372 | 0.93014 | 1.45405 | 0.00000 |
| IL17RA | 0.15723 | 0.17313 | -0.00676 | 0.95320 |
| ILKAP | 0.20162 | 0.47253 | 0.66583 | 0.01716 |
| FGFR2 | 0.08766 | 0.41090 | 0.04444 | 0.65241 |
| IL33 | 0.78078 | 0.00042 | 0.67007 | 0.00121 |
| MZB1 | 0.62348 | 0.00349 | 0.11581 | 0.60409 |
| IGFBP6 | -0.12743 | 0.44880 | -0.19272 | 0.23591 |
| AMBP | 0.04780 | 0.59268 | -0.13807 | 0.34922 |
| ITGAM | -0.19596 | 0.28219 | 0.19240 | 0.22478 |
| TNFRSF12A | 0.07790 | 0.71149 | 0.20909 | 0.32256 |
| STX8 | -0.21972 | 0.39424 | -0.42757 | 0.08797 |
| SOD2 | 0.29724 | 0.25323 | 0.52319 | 0.01493 |
| EDNRB | 0.14880 | 0.42451 | 0.35604 | 0.07287 |
| MMUT | 0.12245 | 0.74390 | 1.36052 | 0.00009 |
| SNCG | 0.40381 | 0.22280 | 0.81393 | 0.02330 |
| SYT1 | 0.22620 | 0.16347 | -0.00135 | 0.99192 |
| B3GAT3 | 0.08427 | 0.62615 | -0.21634 | 0.07642 |
| LAIR2 | -0.11964 | 0.70975 | -0.26941 | 0.33266 |
| L3HYPDH | 0.15950 | 0.67489 | 1.51968 | 0.00001 |
| TNFRSF10A | 0.21455 | 0.32496 | 0.21611 | 0.31926 |
| CST1 | 0.62528 | 0.13201 | -0.30123 | 0.48593 |
| PDGFRA | 0.03417 | 0.83207 | 0.06518 | 0.70158 |
| BRK1 | 0.31625 | 0.17198 | 0.44586 | 0.04165 |
| ARHGAP25 | -0.45226 | 0.08459 | -0.26126 | 0.29841 |
| IL13RA2 | 0.08945 | 0.72049 | -0.46570 | 0.06247 |
| MORN4 | 0.11458 | 0.64979 | -0.48594 | 0.04349 |
| UBE2Z | 0.11165 | 0.61571 | 0.74305 | 0.00128 |
| CALCB | 0.30241 | 0.16981 | 0.25204 | 0.18764 |
| LPL | -0.36419 | 0.05047 | -0.34195 | 0.09184 |
| PMVK | 0.32902 | 0.37962 | 3.34795 | 0.00000 |
| GALNT7 | -0.24874 | 0.07754 | -0.05744 | 0.80903 |
| FAM3B | 0.03450 | 0.82536 | -0.56360 | 0.00997 |
| LYAR | 0.30639 | 0.49704 | 0.52788 | 0.19345 |
| PRKD2 | 0.03176 | 0.79982 | 0.27362 | 0.07453 |
| CLEC10A | -0.08419 | 0.59595 | -0.32687 | 0.04150 |
| NCR1 | 0.30434 | 0.02342 | 0.12999 | 0.36044 |
| CFHR4 | 0.26128 | 0.09701 | -0.02359 | 0.86990 |
| S100A14 | 0.03714 | 0.85087 | -0.65646 | 0.00177 |
| AP2B1 | -0.12905 | 0.64710 | 0.68802 | 0.00431 |
| PIGR | 0.48679 | 0.00387 | 0.05672 | 0.74634 |
| ERP44 | 0.01660 | 0.86311 | 0.24100 | 0.26380 |
| ACAA1 | 0.49454 | 0.22789 | 1.35622 | 0.00034 |
| CFHR5 | 0.17174 | 0.24847 | 0.11344 | 0.44847 |
| SPESP1 | -0.25268 | 0.12969 | -0.05547 | 0.69295 |
| DDX4 | 0.16598 | 0.62222 | -0.74858 | 0.03433 |
| ATG16L1 | -0.27001 | 0.28189 | 1.01475 | 0.00001 |
| HSPA2 | -0.06043 | 0.75742 | 0.68667 | 0.00019 |
| PTPN6 | 0.10974 | 0.65510 | 2.61751 | 0.00000 |

| | | | | |
|---|---|---|---|---|
| FBP1 | 0.10061 | 0.80463 | 0.93667 | 0.01316 |
| ADAMTS16 | 0.34847 | 0.01569 | 0.34424 | 0.01914 |
| MARS1 | -0.23772 | 0.38877 | 0.74493 | 0.00561 |
| MPIG6B | -0.18462 | 0.51988 | -0.96676 | 0.00107 |
| PLPBP | -0.12629 | 0.63018 | 1.43485 | 0.00000 |
| SERPINA1 | 0.24542 | 0.01620 | 0.01142 | 0.89455 |
| ULBP2 | 0.39764 | 0.07438 | 0.20212 | 0.33545 |
| YARS1 | 0.40317 | 0.31716 | 3.85343 | 0.00000 |
| KIF20B | 0.91396 | 0.02168 | 0.20361 | 0.55250 |
| ATXN10 | -0.09128 | 0.72296 | 0.73894 | 0.01402 |
| LGALS8 | 0.14645 | 0.59202 | 0.57445 | 0.01054 |
| CA3 | -0.10591 | 0.66093 | 1.63051 | 0.00000 |
| EGLN1 | 0.23137 | 0.60374 | 0.90798 | 0.02482 |
| GALNT5 | 0.44245 | 0.01303 | 0.04468 | 0.78728 |
| SELPLG | -0.03330 | 0.55863 | 0.09967 | 0.29659 |
| WDR46 | 0.36051 | 0.26746 | 1.12009 | 0.00013 |
| LELP1 | 1.18697 | 0.07686 | 0.22384 | 0.70795 |
| NOMO1 | 0.03221 | 0.72882 | -0.03129 | 0.78596 |
| ELOB | 0.02499 | 0.91197 | 0.27959 | 0.17398 |
| TYRP1 | -0.15611 | 0.31778 | -0.35972 | 0.03027 |
| DLG4 | -0.32826 | 0.37552 | -0.13981 | 0.66080 |
| PYY | 1.10990 | 0.00365 | -0.18832 | 0.63697 |
| SERPINB9 | -0.19964 | 0.14103 | 0.24971 | 0.10670 |
| TXNRD1 | -0.01238 | 0.95342 | 0.90281 | 0.00000 |
| PDIA2 | 0.36577 | 0.22647 | 0.30662 | 0.30893 |
| TRIM58 | 0.07533 | 0.81384 | 2.00111 | 0.00000 |
| CD80 | 0.29541 | 0.05279 | 0.17508 | 0.25049 |
| GGT5 | -0.03095 | 0.81384 | -0.05600 | 0.67865 |
| ENOX2 | 0.24149 | 0.14350 | 0.31921 | 0.03520 |
| XCL1 | 0.22936 | 0.19951 | 0.02132 | 0.91889 |
| RRM2 | 0.65741 | 0.07454 | 0.07771 | 0.79188 |
| CPB1 | 0.30203 | 0.25941 | 0.13282 | 0.61951 |
| PSRC1 | 0.23267 | 0.30750 | 0.44224 | 0.03298 |
| TF | -0.23567 | 0.11444 | -0.36540 | 0.01562 |
| MUC13 | 0.49951 | 0.19698 | 0.27895 | 0.33455 |
| PI16 | -0.27789 | 0.12453 | -0.42441 | 0.00495 |
| CRADD | -0.23974 | 0.43975 | 0.87082 | 0.00598 |
| MSRA | -0.29449 | 0.31575 | 0.82331 | 0.00475 |
| BCL2L1 | 0.42224 | 0.18206 | 1.94724 | 0.00000 |
| ERBB4 | -0.03337 | 0.71314 | -0.23888 | 0.03814 |
| S100A16 | 0.04375 | 0.85841 | -0.36104 | 0.14866 |
| NUP50 | -0.02896 | 0.85983 | -0.50361 | 0.01276 |
| CAPN3 | -0.33065 | 0.08810 | -0.24361 | 0.16353 |
| CD302 | 0.45569 | 0.02731 | 0.32936 | 0.19997 |
| CLUL1 | 0.04093 | 0.81648 | -0.48985 | 0.00276 |
| CFD | -0.11447 | 0.28270 | -0.11578 | 0.22371 |
| LRP1 | -0.13798 | 0.26621 | -0.73409 | 0.00026 |
| TNFRSF14 | 0.30390 | 0.03653 | -0.12992 | 0.42729 |

| | | | | |
|---|---|---|---|---|
| ACYP1 | -0.08054 | 0.73469 | 0.90114 | 0.00010 |
| EGFR | -0.21181 | 0.00956 | -0.29228 | 0.00510 |
| TSHB | 0.25682 | 0.39785 | -0.22239 | 0.47469 |
| MYO9B | -0.56095 | 0.05656 | -0.11129 | 0.66272 |
| BCL2 | 0.46364 | 0.02273 | 0.56582 | 0.02201 |
| FOS | 0.13913 | 0.72160 | -0.29939 | 0.48799 |
| SEMA3G | 0.04347 | 0.82593 | -0.21561 | 0.23701 |
| AOC1 | -0.16558 | 0.57057 | -0.74939 | 0.00313 |
| MMP3 | 0.02572 | 0.90140 | -0.94806 | 0.00013 |
| TMEM106A | 0.05244 | 0.84803 | -0.87993 | 0.00026 |
| NAA80 | 0.43113 | 0.33423 | 3.09394 | 0.00000 |
| AGRP | 0.44382 | 0.02233 | 1.17092 | 0.00001 |
| DNAJC21 | -0.07306 | 0.68459 | 0.77826 | 0.00002 |
| FUT3_FUT5 | 0.49342 | 0.02067 | -0.02013 | 0.92741 |
| TSPAN7 | 0.70659 | 0.02572 | 0.22097 | 0.48345 |
| CD38 | -0.13346 | 0.43221 | -0.16705 | 0.44008 |
| AMY1A_AMY1B_AM | 0.01722 | 0.92390 | -0.23940 | 0.12445 |
| ZNF830 | -0.15442 | 0.53045 | 0.01725 | 0.94042 |
| NFX1 | -0.00014 | 0.99953 | 0.94046 | 0.00004 |
| MMP7 | 0.18250 | 0.44249 | -0.95607 | 0.00023 |
| SDHB | 0.29760 | 0.44356 | 1.56410 | 0.00002 |
| AMPD3 | -0.03486 | 0.88930 | 1.14311 | 0.00005 |
| COL24A1 | 0.36374 | 0.03594 | -0.30629 | 0.05626 |
| GALNT2 | -0.27341 | 0.00526 | -0.36050 | 0.03869 |
| CLSPN | 0.17470 | 0.17790 | -0.06301 | 0.72409 |
| BTLA | 0.40288 | 0.20049 | 0.34699 | 0.30657 |
| DTX2 | -0.63844 | 0.05976 | 0.59885 | 0.09003 |
| ANGPT1 | 0.21330 | 0.39570 | -1.09865 | 0.00001 |
| SFTPA1 | 0.33065 | 0.11421 | -0.01168 | 0.95444 |
| PRKCQ | 0.06428 | 0.62344 | -0.34294 | 0.01130 |
| ATP1B1 | -0.00511 | 0.98117 | -0.59466 | 0.00885 |
| GRP | 0.28521 | 0.36407 | 0.58635 | 0.02490 |
| HPGDS | -0.39220 | 0.03895 | -0.38578 | 0.03884 |
| CD276 | 0.66323 | 0.00085 | 0.15126 | 0.45539 |
| SDC1 | 0.71678 | 0.02063 | 0.55051 | 0.05275 |
| TANK | 0.06429 | 0.79328 | 0.45628 | 0.05496 |
| CEP170 | -0.29699 | 0.32258 | -0.54571 | 0.05732 |
| ATRN | -0.05421 | 0.56339 | -0.21411 | 0.02475 |
| PLIN3 | -0.19685 | 0.31712 | 0.41254 | 0.03809 |
| CCAR2 | 0.21552 | 0.47703 | 0.68057 | 0.03347 |
| LTBP3 | 0.32728 | 0.13151 | 0.59144 | 0.00625 |
| CTF1 | 0.09928 | 0.74354 | 0.60921 | 0.02817 |
| AMY2B | -0.05152 | 0.80546 | -0.36086 | 0.06359 |
| FURIN | 0.00176 | 0.98914 | -0.13866 | 0.30067 |
| DARS1 | -0.24191 | 0.35058 | 0.85757 | 0.00218 |
| GRN | 0.17304 | 0.21204 | -0.00946 | 0.94144 |
| RLN2 | 0.19515 | 0.43146 | -1.79818 | 0.00001 |
| GAPDH | 0.72130 | 0.00731 | 2.48710 | 0.00000 |

| | | | | |
|---|---|---|---|---|
| RNF168 | 0.39020 | 0.11434 | 0.13396 | 0.60614 |
| C1GALT1C1 | -0.07894 | 0.59070 | -0.38037 | 0.00772 |
| UFD1 | 0.30228 | 0.47897 | 3.54240 | 0.00000 |
| EHBP1 | 0.15762 | 0.57180 | 1.23999 | 0.00000 |
| ADGRG1 | 0.24389 | 0.36551 | -0.09207 | 0.70009 |
| DNAJA1 | 0.00530 | 0.98041 | -0.07778 | 0.73984 |
| KAZALD1 | -0.56787 | 0.00630 | -0.18551 | 0.36566 |
| MAPK9 | -0.14886 | 0.47322 | 0.67758 | 0.00236 |
| SERPINH1 | 0.86573 | 0.07580 | 3.87690 | 0.00000 |
| CKB | 1.27927 | 0.00000 | 0.71803 | 0.00349 |
| FCGR2A | 0.43984 | 0.01584 | 0.11279 | 0.55833 |
| IRAK4 | 0.24683 | 0.44275 | 1.92827 | 0.00000 |
| RGMA | -0.29438 | 0.02619 | -0.36520 | 0.01565 |
| WFDC12 | 0.18997 | 0.40867 | -0.40535 | 0.06799 |
| LEG1 | -0.23648 | 0.31019 | 0.10239 | 0.68237 |
| CILP | 0.25314 | 0.22633 | 0.88752 | 0.00015 |
| CHGA | 0.89493 | 0.00256 | 1.63861 | 0.00000 |
| FKBP14 | 0.04504 | 0.89688 | 1.28081 | 0.00030 |
| CCL5 | 0.18914 | 0.52650 | 0.06987 | 0.78654 |
| PINLYP | -0.52383 | 0.02626 | -0.85686 | 0.00020 |
| BST2 | 0.63285 | 0.09359 | 0.64456 | 0.06352 |
| EPPK1 | 0.55621 | 0.06318 | 0.34719 | 0.23757 |
| TLR3 | -0.05626 | 0.76749 | -0.00646 | 0.97471 |
| CCDC134 | 0.21260 | 0.50530 | 0.80746 | 0.00713 |
| OFD1 | -0.47076 | 0.22000 | 0.52879 | 0.21556 |
| TBCC | -0.13814 | 0.67656 | 1.65395 | 0.00000 |
| THSD1 | -0.17526 | 0.26471 | -0.21323 | 0.19403 |
| FGF21 | 0.42033 | 0.37371 | 1.51158 | 0.00176 |
| NPTX2 | 0.01633 | 0.92395 | 0.28277 | 0.13283 |
| FTCD | -0.01611 | 0.96701 | 0.87585 | 0.01616 |
| HDGF | 0.28626 | 0.33861 | 1.63110 | 0.00001 |
| ATF2 | 0.26232 | 0.21461 | -0.47796 | 0.10897 |
| TP53BP1 | 0.38178 | 0.15325 | 0.90167 | 0.00250 |
| PRTG | -0.11929 | 0.29152 | -0.25769 | 0.15117 |
| NT5C | 0.22588 | 0.44986 | 1.33756 | 0.00000 |
| COL4A1 | 0.17017 | 0.43240 | -0.08248 | 0.70873 |
| MXRA8 | -0.10635 | 0.40927 | -0.38889 | 0.00190 |
| PPP1R9B | 0.13876 | 0.61660 | 1.37526 | 0.00011 |
| FDX1 | 0.57231 | 0.08622 | 1.06546 | 0.00077 |
| LTBR | 0.17912 | 0.27202 | 0.12621 | 0.47770 |
| WWP2 | -0.50109 | 0.04193 | 0.65895 | 0.00828 |
| CES3 | -0.28355 | 0.17647 | -1.25756 | 0.00000 |
| CEACAM21 | 0.01929 | 0.94860 | 0.23809 | 0.38897 |
| TGFB1 | 0.45571 | 0.00344 | -0.27615 | 0.12207 |
| VSTM1 | 0.28947 | 0.25105 | 0.17594 | 0.47281 |
| FGR | -0.21242 | 0.62713 | 0.92162 | 0.03910 |
| OSM | 0.72122 | 0.06475 | 0.09127 | 0.80920 |
| CXCL10 | 0.17577 | 0.50405 | 0.02923 | 0.91595 |

| | | | | |
|---|---|---|---|---|
| PSCA | 0.08211 | 0.89195 | -0.25082 | 0.67613 |
| BPIFA2 | -0.94263 | 0.02133 | -0.86191 | 0.01855 |
| HAGH | 0.21379 | 0.46509 | 3.82671 | 0.00000 |
| CD164L2 | -0.11827 | 0.52329 | -0.17502 | 0.30732 |
| YWHAQ | 0.01136 | 0.96691 | 1.14896 | 0.00000 |
| MCFD2 | 0.14330 | 0.33647 | 0.21229 | 0.19226 |
| RARRES1 | 0.18024 | 0.23461 | -0.12878 | 0.38466 |
| PPL | 0.50472 | 0.08147 | 0.97113 | 0.00007 |
| SV2A | 0.23848 | 0.44992 | 0.05776 | 0.84368 |
| FOLR1 | 0.39642 | 0.00594 | 0.03243 | 0.81812 |
| TUBB3 | 0.95921 | 0.00065 | 0.77984 | 0.00008 |
| FOXO3 | 0.55585 | 0.09424 | 1.85420 | 0.00000 |
| QDPR | 0.03471 | 0.85843 | 1.26736 | 0.00000 |
| CKMT1B; CKMT1A | 0.40642 | 0.10693 | 1.06637 | 0.00003 |
| ADAMTS1 | 0.26228 | 0.23918 | 0.69200 | 0.00316 |
| AKR7L | 0.11316 | 0.69610 | 0.71685 | 0.00962 |
| LYVE1 | 0.08872 | 0.56049 | -0.08423 | 0.52364 |
| CRLF1 | -0.07447 | 0.52676 | 0.08541 | 0.51075 |
| RAB37 | 0.04043 | 0.88833 | 0.47829 | 0.10956 |
| LILRB1 | 0.07306 | 0.51814 | -0.06548 | 0.59491 |
| GYS1 | 0.35642 | 0.31596 | 1.73461 | 0.00001 |
| GPC1 | -0.23266 | 0.06060 | -0.23651 | 0.25047 |
| KIFBP | 0.05542 | 0.83802 | 1.84330 | 0.00000 |
| RUVBL1 | -0.36104 | 0.30724 | -0.40610 | 0.23554 |
| DKK4 | 0.19186 | 0.45375 | -0.09913 | 0.67123 |
| PLIN1 | -0.02473 | 0.93509 | 0.35116 | 0.30426 |
| VIM | -0.11607 | 0.66807 | -0.24489 | 0.42076 |
| SIL1 | -0.07189 | 0.56283 | -0.30626 | 0.01234 |
| CDH22 | 0.11507 | 0.63159 | -0.11582 | 0.63940 |
| DLL1 | 0.31886 | 0.02980 | 0.27937 | 0.08193 |
| SEPTIN9 | -0.21306 | 0.37804 | 0.44397 | 0.06801 |
| IGSF9 | -0.10201 | 0.73545 | 0.58622 | 0.04998 |
| RFC4 | -0.11134 | 0.53806 | 0.32208 | 0.05720 |
| ACRBP | 0.14508 | 0.32355 | 0.49489 | 0.00052 |
| GIMAP8 | 0.14935 | 0.58142 | 0.42754 | 0.09157 |
| MRPL46 | 0.02397 | 0.87284 | 0.27299 | 0.01236 |
| CD22 | -0.16317 | 0.26042 | -0.36903 | 0.07711 |
| CEACAM8 | 0.20794 | 0.41263 | -0.18332 | 0.51379 |
| CCDC28A | -0.06114 | 0.65943 | 0.02679 | 0.84370 |
| RANBP2 | -0.07779 | 0.65528 | -0.07419 | 0.70285 |
| IGF1R | 0.08646 | 0.45503 | -0.07869 | 0.62685 |
| LIF | 0.55115 | 0.20198 | 0.53984 | 0.20101 |
| MAX | 0.19992 | 0.60190 | 1.88318 | 0.00000 |
| BATF | 0.09740 | 0.74026 | -0.02862 | 0.92971 |
| CA9 | 0.39873 | 0.16623 | -0.06937 | 0.80184 |
| IL11 | -0.46440 | 0.03319 | -0.04980 | 0.83758 |
| KIAA2013 | 0.10134 | 0.60624 | -0.50376 | 0.00381 |
| CNPY2 | -0.02609 | 0.89531 | 0.45516 | 0.02187 |

| | | | | |
|---|---|---|---|---|
| LHPP | 0.18640 | 0.44772 | 1.52748 | 0.00000 |
| COMT | -0.18232 | 0.51085 | 1.39286 | 0.00001 |
| SLC9A3R1 | -0.22128 | 0.47321 | 1.12498 | 0.00016 |
| LTA | -0.10878 | 0.11995 | -0.23146 | 0.00525 |
| ADGRE1 | 0.07324 | 0.76164 | -0.01602 | 0.93855 |
| TMPRSS15 | -0.24684 | 0.49700 | -0.80945 | 0.01787 |
| DCUN1D1 | 0.14954 | 0.58507 | -0.31120 | 0.24606 |
| ERC2 | 0.26180 | 0.31767 | 0.95373 | 0.00132 |
| CD70 | 0.23495 | 0.13308 | -0.64332 | 0.00141 |
| LECT2 | 0.39880 | 0.15353 | 0.71757 | 0.01421 |
| APOE | 0.38558 | 0.03505 | 0.27059 | 0.08924 |
| HYOU1 | 0.16614 | 0.03699 | 0.05194 | 0.60565 |
| SH2B3 | 0.20923 | 0.55797 | 1.65884 | 0.00001 |
| ALDH1A1 | 0.11116 | 0.67969 | 1.37497 | 0.00000 |
| LILRA5 | 0.23408 | 0.08100 | -0.03363 | 0.80235 |
| ELN | 0.78317 | 0.00002 | 0.05600 | 0.77824 |
| ENPP7 | 0.32583 | 0.30608 | 0.47102 | 0.12330 |
| CGB3; CGB5; CGB8 | 0.44072 | 0.19743 | 0.43927 | 0.03120 |
| MSR1 | 0.37389 | 0.02565 | 0.33128 | 0.07315 |
| KLRD1 | 0.44496 | 0.00483 | 0.23391 | 0.34495 |
| PDIA4 | 0.33263 | 0.25512 | 1.21582 | 0.00001 |
| CNGB3 | -0.19201 | 0.24285 | -0.04729 | 0.73015 |
| RGCC | 0.29853 | 0.19765 | 1.62307 | 0.00000 |
| SSC4D | -1.75934 | 0.00007 | -0.65611 | 0.09733 |
| HRG | -0.17657 | 0.20922 | -0.09634 | 0.41940 |
| PPP1R14D | 0.70038 | 0.00438 | 0.25219 | 0.26043 |
| CNTN2 | -0.13982 | 0.43105 | -0.44716 | 0.03073 |
| SERPIND1 | 0.05687 | 0.68197 | -0.17284 | 0.17374 |
| FABP4 | 0.19336 | 0.56382 | 1.18362 | 0.00057 |
| NXPH1 | -0.05669 | 0.41861 | -0.04138 | 0.45293 |
| MMP12 | 0.89511 | 0.00281 | -0.05739 | 0.85668 |
| PLAUR | 0.44015 | 0.01889 | 0.16965 | 0.35919 |
| LY9 | 0.09238 | 0.40208 | -0.29292 | 0.11804 |
| PCSK9 | -0.05720 | 0.72470 | -0.32235 | 0.05178 |
| HYAL1 | -0.03989 | 0.67636 | -0.22496 | 0.04251 |
| CD300A | 0.24292 | 0.08377 | 0.16614 | 0.23181 |
| XPNPEP2 | 0.03653 | 0.90088 | -0.23002 | 0.40482 |
| CRELD1 | -0.07913 | 0.52191 | 0.03459 | 0.78802 |
| APOB | 0.17504 | 0.27316 | -0.84171 | 0.00000 |
| KLK8 | -0.23886 | 0.23559 | -0.55579 | 0.00124 |
| YY1 | 0.02122 | 0.93859 | -0.06390 | 0.82188 |
| BEX3 | 0.41848 | 0.02763 | -0.18354 | 0.33756 |
| CC2D1A | -0.59494 | 0.09403 | -0.04921 | 0.88024 |
| GRK5 | -0.02945 | 0.88470 | 0.12171 | 0.54747 |
| F11 | -0.15684 | 0.15729 | -0.02979 | 0.77139 |
| RAB6A | -0.45043 | 0.12448 | -1.44431 | 0.00004 |
| SCGB3A2 | 0.05710 | 0.79191 | -0.49133 | 0.07386 |
| IGFL4 | 0.03935 | 0.88559 | 0.32829 | 0.19541 |

| | | | | |
|---|---|---|---|---|
| SEMA6C | -0.08747 | 0.36834 | -0.31828 | 0.00212 |
| IGF2BP3 | -0.60079 | 0.03905 | -0.17851 | 0.50729 |
| PRDX6 | -0.36014 | 0.09873 | 1.16255 | 0.00001 |
| ENSA | 0.28213 | 0.16110 | 1.21697 | 0.00000 |
| MED18 | -0.30826 | 0.24182 | 1.38647 | 0.00000 |
| CPA1 | 0.02887 | 0.92479 | -0.23469 | 0.37220 |
| OCLN | 0.65327 | 0.04439 | 0.71009 | 0.01089 |
| GOT1 | 0.02583 | 0.93120 | 0.71592 | 0.00319 |
| MTIF3 | 0.18640 | 0.63824 | 1.97078 | 0.00000 |
| CLEC3B | -0.46436 | 0.00010 | -0.39685 | 0.00030 |
| IFIT3 | 0.06002 | 0.87775 | 0.56617 | 0.15938 |
| DXO | -0.04602 | 0.85386 | 1.32664 | 0.00000 |
| DTYMK | -0.07894 | 0.77818 | 1.62266 | 0.00000 |
| TGFBR2 | 0.18889 | 0.17931 | 0.05907 | 0.68149 |
| LSP1 | 0.24163 | 0.29689 | 0.01684 | 0.93863 |
| DCTN1 | 0.08084 | 0.71749 | 0.44976 | 0.04868 |
| APOA1 | -0.15077 | 0.32114 | -0.75207 | 0.00000 |
| ANGPTL4 | 0.24895 | 0.13511 | 0.77257 | 0.00010 |
| HEG1 | -0.06268 | 0.62296 | -0.05173 | 0.65785 |
| GFRA3 | -0.19061 | 0.26409 | -0.38869 | 0.02818 |
| TST | -0.02028 | 0.93334 | 0.87447 | 0.00021 |
| MFGE8 | 0.15410 | 0.40695 | -0.21185 | 0.27667 |
| NAGA | -0.24100 | 0.18384 | -0.78399 | 0.00001 |
| ENPEP | -0.19700 | 0.46972 | 0.04557 | 0.85694 |
| BCHE | -0.49863 | 0.00438 | -0.37787 | 0.01857 |
| EZR | 0.16627 | 0.33650 | 0.58099 | 0.00006 |
| GLRX5 | 0.26049 | 0.49274 | 1.55484 | 0.00001 |
| ITPR1 | 0.60586 | 0.02832 | 0.40657 | 0.04811 |
| DDC | -0.04638 | 0.87508 | -0.33033 | 0.21440 |
| CRYBB1 | 0.07535 | 0.74455 | 0.72993 | 0.00090 |
| CA14 | -0.33161 | 0.08917 | -0.48689 | 0.02330 |
| VNN1 | -0.29134 | 0.18052 | 0.08353 | 0.64644 |
| TAFA5 | 0.06385 | 0.72886 | -0.02342 | 0.89837 |
| TDO2 | 0.20213 | 0.65660 | 0.92691 | 0.02320 |
| SAG | 0.42505 | 0.21541 | 0.19310 | 0.61543 |
| SPRING1 | 0.09027 | 0.58373 | -0.12163 | 0.44565 |
| CGREF1 | 0.09639 | 0.61294 | -0.05833 | 0.75956 |
| CTSE | -0.05138 | 0.81030 | -0.03317 | 0.86069 |
| TRAF3 | -0.40100 | 0.01268 | 0.14228 | 0.41363 |
| BPIFB1 | -0.02670 | 0.93866 | -0.09370 | 0.75572 |
| IL19 | 0.61362 | 0.03171 | 0.16594 | 0.51182 |
| SNU13 | -0.26196 | 0.53445 | 0.30855 | 0.42550 |
| DENND2B | 0.16650 | 0.34472 | 0.01723 | 0.93107 |
| ANXA2 | 0.62596 | 0.11911 | 1.30996 | 0.00023 |
| CD6 | 0.06069 | 0.76129 | -0.08301 | 0.74386 |
| F2R | 0.59087 | 0.00529 | 2.61994 | 0.00000 |
| BPIFB2 | -0.23653 | 0.42557 | 0.00428 | 0.98691 |
| PALM2 | 0.64463 | 0.03208 | 0.92418 | 0.00193 |

| | | | | |
|---|---|---|---|---|
| BTNL10 | 0.18273 | 0.18016 | -0.02865 | 0.87757 |
| MVK | -0.12572 | 0.73544 | 1.38305 | 0.00008 |
| C2CD2L | -0.61170 | 0.11609 | -1.38026 | 0.00001 |
| RAD51 | 0.36212 | 0.27134 | 0.53839 | 0.11611 |
| PTGR1 | -0.54824 | 0.11784 | 1.05148 | 0.00040 |
| C1QTNF6 | 0.24956 | 0.32240 | 1.20397 | 0.00000 |
| SNX15 | -0.02096 | 0.92644 | 0.94277 | 0.00000 |
| NUB1 | -0.24959 | 0.43767 | 1.04355 | 0.00098 |
| GUCY2C | 0.53981 | 0.20537 | -0.20458 | 0.65108 |
| GOLM2 | 0.00536 | 0.96525 | 0.00871 | 0.95128 |
| GID8 | 0.06831 | 0.65605 | 0.34074 | 0.04449 |
| SRP14 | 0.33205 | 0.26439 | 0.27699 | 0.34225 |
| GSN | -0.37421 | 0.00082 | -0.36072 | 0.00019 |
| MMP10 | -0.06861 | 0.72717 | -0.57003 | 0.01383 |
| UNG | 0.35919 | 0.19718 | 0.67651 | 0.00389 |
| PDIA3 | 0.30049 | 0.24155 | -0.55617 | 0.00515 |
| CLEC4M | 0.02505 | 0.84316 | -0.28816 | 0.01347 |
| PLXNB2 | 0.06501 | 0.49183 | -0.02687 | 0.81596 |
| CALCOCO2 | -0.25388 | 0.39569 | 0.36256 | 0.17854 |
| STC2 | -0.13919 | 0.33013 | 0.02716 | 0.86097 |
| SMC3 | -0.38056 | 0.06673 | -0.13411 | 0.42419 |
| LRP2 | 0.02120 | 0.89956 | -0.20617 | 0.33966 |
| ST8SIA1 | -0.03678 | 0.79078 | -0.65925 | 0.00022 |
| IFNL2 | 0.36951 | 0.14686 | 0.22602 | 0.38388 |
| CST7 | 0.20153 | 0.52432 | -0.06122 | 0.84804 |
| PCSK7 | 0.00096 | 0.99381 | 0.01710 | 0.89586 |
| NDUFA5 | 0.45747 | 0.03679 | 0.85982 | 0.00002 |
| GCHFR | 0.12078 | 0.69403 | 1.07294 | 0.00007 |
| LYSMD3 | 0.37370 | 0.21527 | 0.52290 | 0.06397 |
| PSMG3 | 0.12303 | 0.53831 | 1.63074 | 0.00000 |
| HDAC9 | -0.17644 | 0.69812 | 0.59185 | 0.24004 |
| ADGRF5 | 0.17375 | 0.02294 | -0.07996 | 0.31257 |
| REN | 0.05604 | 0.82822 | -0.18812 | 0.40126 |
| TPR | 0.14702 | 0.69698 | 0.65391 | 0.10342 |
| GRIK2 | -0.09139 | 0.54357 | -0.40227 | 0.02354 |
| PEBP1 | -0.03254 | 0.88164 | 1.46109 | 0.00000 |
| AP3B1 | -0.05857 | 0.85132 | 0.92224 | 0.00265 |
| CLU | 0.11120 | 0.22531 | -0.08453 | 0.36964 |
| CD40 | 0.35811 | 0.07464 | 0.25755 | 0.18543 |
| TNN | -0.05065 | 0.64853 | -0.08600 | 0.42910 |
| STX5 | 0.14478 | 0.71474 | 0.96463 | 0.00399 |
| CA4 | -0.15214 | 0.19640 | -0.23451 | 0.03168 |
| CERT1 | -0.26085 | 0.28067 | 0.46881 | 0.09701 |
| MMP1 | 1.20646 | 0.00000 | -0.62083 | 0.05396 |
| CREB3 | 0.38507 | 0.07211 | -0.17018 | 0.47541 |
| CASC3 | 0.10153 | 0.74996 | 1.36760 | 0.00001 |
| PLA2G1B | 0.05875 | 0.82395 | -0.30747 | 0.15838 |
| AMOTL2 | -0.06429 | 0.47910 | 0.26847 | 0.02761 |

| | | | | |
|---|---|---|---|---|
| F9 | 0.00438 | 0.95593 | 0.38099 | 0.16752 |
| MAVS | -0.07917 | 0.80344 | -0.18463 | 0.56037 |
| IL17F | 0.44029 | 0.11718 | -0.29974 | 0.26347 |
| CD2AP | 0.57143 | 0.01361 | 1.55191 | 0.00000 |
| PEAR1 | -0.20188 | 0.05155 | -0.19806 | 0.05275 |
| STAT5B | 0.15853 | 0.72073 | 2.81115 | 0.00000 |
| PKN3 | -0.23917 | 0.36337 | 0.63573 | 0.06424 |
| IGFBP4 | 0.56739 | 0.03811 | 0.21286 | 0.38987 |
| FAM20A | 0.05894 | 0.72286 | 0.21735 | 0.15548 |
| SRPX | 0.18450 | 0.40602 | 0.37639 | 0.06625 |
| CPXM2 | 0.03283 | 0.82755 | 0.09014 | 0.56665 |
| AGT | 0.00273 | 0.94896 | -0.02627 | 0.51237 |
| NOS1 | 0.03871 | 0.89823 | 0.54905 | 0.10549 |
| PRRT3 | 0.15301 | 0.33596 | 0.38360 | 0.01627 |
| MEP1A | -0.21994 | 0.49075 | 0.34378 | 0.15233 |
| LILRA3 | 0.00271 | 0.99015 | -0.01484 | 0.95660 |
| COX5B | 0.30433 | 0.40550 | 1.52677 | 0.00010 |
| OPLAH | 0.25524 | 0.23606 | 0.62996 | 0.00363 |
| PRUNE2 | 0.38988 | 0.06159 | 0.22462 | 0.32304 |
| ERBB3 | 0.11289 | 0.23413 | -0.04922 | 0.62044 |
| TNNI3 | 0.02987 | 0.95754 | -0.79223 | 0.14630 |
| MINK1 | 0.22612 | 0.49854 | 3.60180 | 0.00000 |
| PRSS8 | 0.50869 | 0.00681 | 0.18603 | 0.28624 |
| CDH3 | 0.36614 | 0.05382 | -0.18581 | 0.31113 |
| ADD1 | 0.13714 | 0.51151 | 1.29030 | 0.00000 |
| TRPV3 | 0.04897 | 0.87191 | -0.09241 | 0.72627 |
| PNMA2 | 0.22189 | 0.50319 | 0.46848 | 0.15921 |
| GBP4 | -0.27999 | 0.09489 | -0.52060 | 0.00056 |
| PNLIP | -0.07423 | 0.81580 | -0.24203 | 0.38981 |
| APOBR | 0.07304 | 0.49648 | 0.05095 | 0.63045 |
| MYO6 | 0.39989 | 0.03051 | 0.45272 | 0.02009 |
| SCG3 | -0.01211 | 0.92665 | -0.19122 | 0.17084 |
| LGALS4 | 0.88753 | 0.00176 | 0.72738 | 0.00286 |
| BAG3 | -0.24859 | 0.20551 | 0.40833 | 0.11105 |
| RARRES2 | 0.29927 | 0.10635 | -0.09596 | 0.58676 |
| BTN2A1 | 0.16914 | 0.27352 | -0.01891 | 0.87895 |
| PAMR1 | 0.07937 | 0.51478 | -0.12529 | 0.34778 |
| NOS2 | 0.58838 | 0.03300 | 0.13422 | 0.59740 |
| DAG1 | 0.10172 | 0.56136 | -0.31801 | 0.07592 |
| ACVRL1 | 0.14769 | 0.40899 | 0.01902 | 0.91732 |
| PPIF | 0.03663 | 0.94944 | 1.21447 | 0.01868 |
| CD300E | 0.30555 | 0.16879 | 0.11676 | 0.63169 |
| DDI2 | 0.21910 | 0.43244 | 1.83282 | 0.00000 |
| PRR4 | -0.13534 | 0.61638 | 0.82635 | 0.00122 |
| SERPINB6 | -0.05672 | 0.80146 | 0.73628 | 0.00100 |
| TMSB10 | 0.27474 | 0.39080 | 1.55455 | 0.00003 |
| B4GAT1 | -0.09243 | 0.35389 | -0.15764 | 0.21616 |
| DCDC2C | 0.28714 | 0.29943 | -0.16376 | 0.61861 |

| | | | | |
|---|---|---|---|---|
| SCGB2A2 | 0.26397 | 0.20848 | -0.05087 | 0.81518 |
| RTN4IP1 | 0.32322 | 0.39692 | 2.29398 | 0.00000 |
| RIPK4 | -0.07339 | 0.77527 | -0.09884 | 0.68018 |
| SCPEP1 | 0.41161 | 0.07293 | -0.10017 | 0.61293 |
| APOH | -0.19511 | 0.07218 | -0.03983 | 0.70702 |
| CFHR2 | 0.16283 | 0.33897 | 0.22438 | 0.16525 |
| IGLC2 | 0.41675 | 0.00444 | -0.01230 | 0.91347 |
| RANBP1 | 0.04507 | 0.80190 | 1.76617 | 0.00000 |
| KLK1 | -0.49353 | 0.10828 | 0.27599 | 0.37978 |
| AIF1 | -0.13816 | 0.62763 | 0.70119 | 0.03076 |
| C1QTNF9 | -0.14646 | 0.37994 | -0.66977 | 0.00000 |
| ENO3 | -0.97935 | 0.00136 | 0.82539 | 0.00528 |
| ZBP1 | 0.42502 | 0.31945 | 0.45590 | 0.20104 |
| DBN1 | 0.25494 | 0.13173 | 0.52786 | 0.00118 |
| KLF4 | 0.52711 | 0.11074 | 0.52830 | 0.06006 |
| HEPH | 0.10199 | 0.44161 | -0.16973 | 0.12828 |
| TIGAR | 0.02623 | 0.92554 | 0.61321 | 0.02167 |
| DDX39A | -0.11272 | 0.56055 | 0.13862 | 0.46278 |
| FGFBP1 | -0.00632 | 0.97885 | -0.13351 | 0.56648 |
| SLAMF7 | -0.00217 | 0.99119 | -0.21508 | 0.28373 |
| ECHDC3 | 0.61727 | 0.17935 | 1.33257 | 0.00054 |
| SOD3 | 0.11483 | 0.54379 | -0.37778 | 0.01351 |
| FGF12 | -0.00442 | 0.98420 | -0.32180 | 0.08572 |
| CYTL1 | -0.15801 | 0.38030 | -0.22958 | 0.14132 |
| CUZD1 | 0.34233 | 0.37173 | 0.11661 | 0.71780 |
| PM20D1 | -0.36994 | 0.48968 | -0.62991 | 0.20092 |
| ADM | 0.34380 | 0.11901 | 2.87954 | 0.00000 |
| EIF4G1 | 0.12537 | 0.64905 | 1.30760 | 0.00013 |
| MME | -0.35369 | 0.18190 | -0.21161 | 0.47463 |
| ARHGEF5 | 0.14265 | 0.69051 | 0.75086 | 0.04148 |
| NBL1 | 0.24929 | 0.06849 | -0.08209 | 0.53693 |
| FUCA1 | -0.38229 | 0.13233 | -0.11707 | 0.61100 |
| NCLN | 0.18485 | 0.16920 | 0.00554 | 0.97214 |
| INPPL1 | 0.25337 | 0.45421 | 1.46112 | 0.00001 |
| IL20 | 0.00025 | 0.99861 | -0.02549 | 0.80295 |
| LCAT | -0.27437 | 0.00876 | -0.13863 | 0.20402 |
| PPP1R12A | -0.05951 | 0.86726 | 1.39246 | 0.00018 |
| ANXA1 | -0.03525 | 0.91542 | 0.90224 | 0.00630 |
| BRD3 | -0.14866 | 0.65835 | 0.67713 | 0.04031 |
| COL5A1 | 0.14511 | 0.47672 | -0.23119 | 0.22619 |
| CTSV | -0.45694 | 0.02148 | -0.63716 | 0.00213 |
| PALM3 | 0.33037 | 0.36914 | 0.44225 | 0.18785 |
| TRIM40 | 0.17126 | 0.65988 | 0.81343 | 0.05937 |
| PTRHD1 | 0.08109 | 0.74205 | 1.23244 | 0.00000 |
| AK2 | 0.16953 | 0.61665 | 2.30392 | 0.00000 |
| CEACAM3 | -0.11661 | 0.61701 | -0.01144 | 0.95089 |
| AMDHD2 | 0.06189 | 0.58772 | 0.24921 | 0.01722 |
| TRIM25 | -0.93398 | 0.00346 | 0.06183 | 0.84341 |

| | | | | |
|---|---|---|---|---|
| FCAR | 0.41726 | 0.06414 | 0.13667 | 0.54846 |
| BIRC2 | -0.21161 | 0.14699 | -0.04097 | 0.71857 |
| TMPRSS11B | 0.25171 | 0.42882 | -0.22193 | 0.49992 |
| TGFBR3 | 0.07389 | 0.59680 | -0.07098 | 0.71051 |
| MYH7B | 0.10584 | 0.60892 | -0.19351 | 0.44247 |
| GAST | 1.13953 | 0.01067 | 2.38175 | 0.00000 |
| ID4 | 0.11517 | 0.61660 | 0.41015 | 0.06113 |
| TALDO1 | -0.08204 | 0.72689 | 1.22532 | 0.00000 |
| LTO1 | 0.24925 | 0.14291 | 0.06843 | 0.66190 |
| PPP1CC | -0.18357 | 0.52795 | 1.21508 | 0.00003 |
| SPRY2 | 0.29200 | 0.44911 | 1.99143 | 0.00000 |
| MYDGF | 0.16208 | 0.64253 | 0.52085 | 0.10236 |
| RABGAP1L | 0.24879 | 0.28168 | 1.55256 | 0.00000 |
| RALB | -0.46923 | 0.07532 | -0.95918 | 0.00130 |
| ABRAXAS2 | 0.34671 | 0.19614 | 0.97509 | 0.00035 |
| FHIT | -0.16368 | 0.55994 | 1.31812 | 0.00001 |
| TEF | 0.61897 | 0.04635 | 0.67234 | 0.01064 |
| EPHA4 | -0.14878 | 0.23224 | -0.29818 | 0.02439 |
| SNAPIN | -0.00405 | 0.98406 | 0.70931 | 0.00181 |
| SLIT2 | 0.41182 | 0.07905 | 0.24466 | 0.28805 |
| NID2 | 0.46850 | 0.03255 | -0.23389 | 0.25592 |
| XIAP | -0.25732 | 0.27484 | 0.92857 | 0.00009 |
| ADH4 | -0.29038 | 0.39521 | 0.87288 | 0.00644 |
| TOM1L2 | 0.29829 | 0.38935 | 3.27407 | 0.00000 |
| ECE1 | -0.16205 | 0.12208 | -0.11263 | 0.26069 |
| NT5C3A | -0.36228 | 0.21130 | 1.12076 | 0.00084 |
| ACTN4 | -0.14904 | 0.43639 | -0.19544 | 0.30760 |
| TOP1MT | 0.11886 | 0.57922 | -0.24556 | 0.28520 |
| KLK12 | 0.32225 | 0.47408 | 0.19489 | 0.65717 |
| TCP11 | 0.22379 | 0.67069 | 0.43811 | 0.39831 |
| CHAC2 | 0.18283 | 0.35549 | 0.67877 | 0.00254 |
| VEGFC | 0.15586 | 0.33989 | -0.69032 | 0.00002 |
| ST3GAL1 | 0.02723 | 0.82113 | -0.28926 | 0.08187 |
| CR2 | -0.60572 | 0.00036 | -0.65356 | 0.00138 |
| ACE | -0.01537 | 0.88169 | -0.09052 | 0.31724 |
| LACRT | -0.05333 | 0.80374 | -0.33515 | 0.18424 |
| ANGPT2 | 0.17776 | 0.30193 | -0.12933 | 0.58619 |
| ASGR1 | 0.15726 | 0.35535 | 0.14536 | 0.40535 |
| SH2D1A | -0.06356 | 0.81399 | 0.58740 | 0.02770 |
| FUS | 0.21599 | 0.39148 | 0.85905 | 0.00014 |
| DYNC1H1 | -0.02261 | 0.90898 | 0.13915 | 0.52513 |
| FXN | 0.07769 | 0.77132 | 1.44706 | 0.00000 |
| CHM | -0.11906 | 0.64736 | 0.88912 | 0.00035 |
| VWF | 0.43488 | 0.04803 | 0.12619 | 0.60887 |
| SDC4 | 0.04657 | 0.82061 | -0.78864 | 0.00010 |
| LRCH4 | 0.51761 | 0.20211 | 0.48684 | 0.15157 |
| AHSG | -0.14281 | 0.34524 | -0.08840 | 0.50283 |
| SWAP70 | 0.01271 | 0.95655 | 0.35580 | 0.07878 |

| | | | | |
|---|---|---|---|---|
| MATN2 | -0.08312 | 0.42531 | -0.02933 | 0.80766 |
| TTN | -0.01435 | 0.96340 | 0.17451 | 0.50143 |
| GGA1 | -0.16406 | 0.44207 | -0.04277 | 0.84075 |
| FGF16 | -0.27342 | 0.38025 | -0.14627 | 0.62348 |
| IDUA | 0.01550 | 0.92362 | 0.59261 | 0.00170 |
| GNPDA1 | -0.28130 | 0.07265 | 0.55038 | 0.00044 |
| GTPBP2 | -0.44702 | 0.25957 | -0.35679 | 0.33071 |
| CEACAM18 | -0.18271 | 0.65798 | 0.28646 | 0.47167 |
| IRAG2 | -0.50564 | 0.18364 | -0.17456 | 0.64918 |
| IKZF2 | 0.03853 | 0.89574 | 0.19628 | 0.54032 |
| TSC1 | 0.02641 | 0.92404 | -0.07807 | 0.77557 |
| ACY3 | 0.23301 | 0.40547 | 0.19542 | 0.42139 |
| CD109 | -0.15043 | 0.26129 | -0.34036 | 0.01806 |
| CASP7 | 0.48094 | 0.07956 | 1.55050 | 0.00000 |
| NRP2 | 0.09929 | 0.43490 | 0.01858 | 0.89033 |
| MCEE | 0.09802 | 0.78394 | 1.23656 | 0.00012 |
| GUSB | -0.36029 | 0.03665 | -0.38199 | 0.04604 |
| AZI2 | -0.11002 | 0.69189 | 0.24297 | 0.38673 |
| SMPDL3B | -0.28343 | 0.37775 | -0.57007 | 0.04345 |
| CLSTN2 | 0.07903 | 0.66053 | 0.00156 | 0.99453 |
| TMED8 | 0.37881 | 0.28157 | 1.77601 | 0.00000 |
| STC1 | 0.15079 | 0.58665 | 0.56740 | 0.02535 |
| MLLT1 | 0.24322 | 0.49048 | 0.11033 | 0.76054 |
| PDLIM5 | 0.00224 | 0.99359 | 1.03994 | 0.00053 |
| GLOD4 | 0.01197 | 0.93138 | 1.04713 | 0.00000 |
| DHODH | 0.37046 | 0.19589 | 0.42384 | 0.12294 |
| EPHB6 | 0.12263 | 0.52831 | -0.15537 | 0.46502 |
| MICALL2 | 0.42896 | 0.15273 | 0.77715 | 0.00116 |
| TNFSF12 | 0.09539 | 0.47354 | -0.53445 | 0.03133 |
| SDK2 | -0.20962 | 0.12098 | -0.54529 | 0.00016 |
| CES1 | -0.20398 | 0.50412 | 0.12473 | 0.64379 |
| VAV3 | -0.45191 | 0.25410 | -0.66630 | 0.06240 |
| VWA1 | 0.16221 | 0.50666 | 0.27868 | 0.32533 |
| ABO | -0.32031 | 0.51421 | -0.69432 | 0.13827 |
| TERF1 | -0.11999 | 0.60840 | 0.27403 | 0.25306 |
| LYPD3 | -0.22412 | 0.18759 | -0.19703 | 0.25604 |
| IL7 | -0.01108 | 0.96455 | -0.82511 | 0.00039 |
| PPM1A | -0.26520 | 0.02699 | -0.08868 | 0.40165 |
| ERI1 | 0.28142 | 0.35367 | 0.42866 | 0.10739 |
| VAMP5 | 0.47186 | 0.17955 | 0.32195 | 0.28485 |
| HLA-DRA | -0.15315 | 0.42702 | -0.90057 | 0.00001 |
| AKT1S1 | 0.39971 | 0.09627 | 1.86690 | 0.00000 |
| AP1G2 | -0.40367 | 0.24425 | -0.59587 | 0.07030 |
| SCG2 | 0.33098 | 0.03364 | 0.04074 | 0.84582 |
| CHMP1A | 0.12559 | 0.59529 | 1.41667 | 0.00000 |
| EFCAB14 | 0.13582 | 0.32108 | -0.30722 | 0.01304 |
| IGHMBP2 | -0.31327 | 0.21421 | 0.38985 | 0.10105 |
| TGFB2 | 0.35066 | 0.06497 | -0.23513 | 0.17591 |

| | | | | |
|---|---|---|---|---|
| FES | -0.67701 | 0.01278 | -0.50651 | 0.09288 |
| NGFR | 0.16746 | 0.36155 | 0.06974 | 0.69387 |
| LRP11 | 0.22195 | 0.10189 | 0.16539 | 0.32344 |
| SPRR3 | 0.48848 | 0.09256 | 0.52101 | 0.03022 |
| RBM19 | 0.82343 | 0.00554 | 0.43383 | 0.12610 |
| SOD1 | 0.11298 | 0.54367 | 1.20984 | 0.00000 |
| TNFSF14 | 0.22332 | 0.48978 | -0.65529 | 0.04448 |
| DPP10 | 0.18481 | 0.49539 | -0.06423 | 0.79160 |
| PRAME | 0.25957 | 0.02420 | -0.26345 | 0.03570 |
| GATD3 | -0.05318 | 0.90063 | 1.33471 | 0.00027 |
| CALB2 | 0.12207 | 0.72147 | 0.55002 | 0.10395 |
| PBLD | 0.02671 | 0.93991 | 0.96589 | 0.00622 |
| PVR | 0.17939 | 0.35481 | 0.16652 | 0.40529 |
| PSMD9 | -0.25362 | 0.25120 | 1.13401 | 0.00001 |
